## Supplement 1 for "Challenges of COVID-19 Case Forecasting in the US, 2020-2021"

**Supporting Information 1**: Team submissions, methods, and data

**S1 Figure 1.1**. Forecasts submitted over time at the national, state-territory-DC level in panel A and at the country scale in Panel B. The number of forecasted locations submitted each week nationally or at the state, territory and DC level is included, while the country level forecast submissions shows the percent of counties per quantile that were submitted each week. Sets of team forecasts meeting the inclusion criteria for this main analysis are labeled with an asterisk (*).

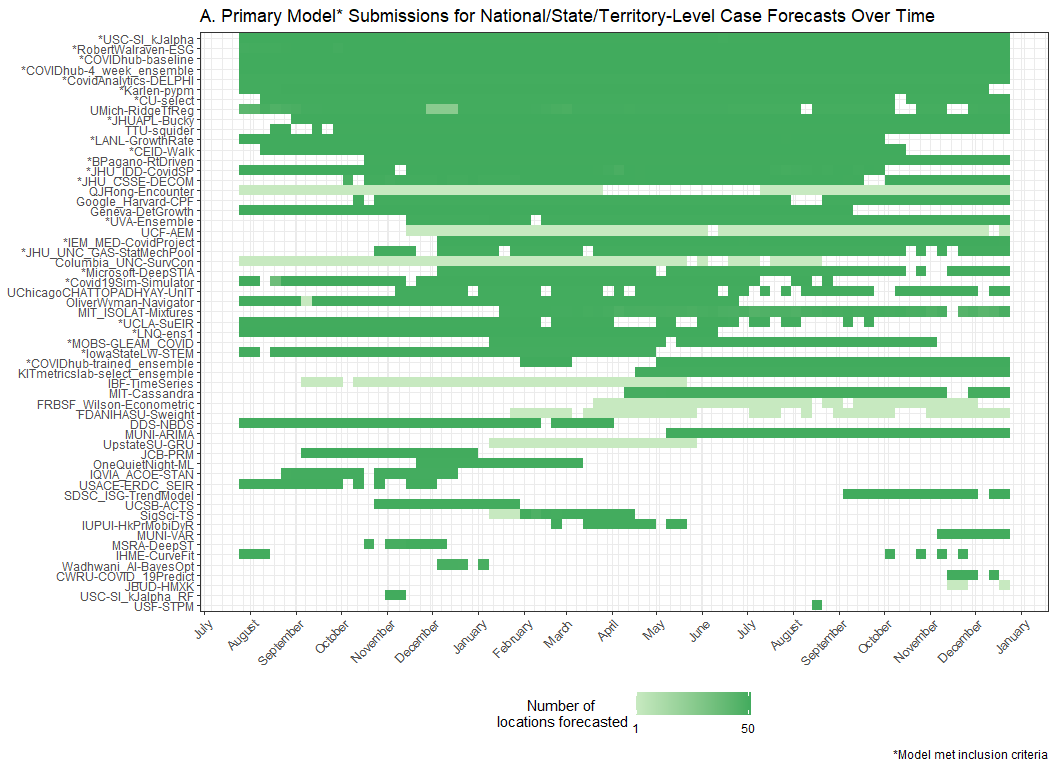

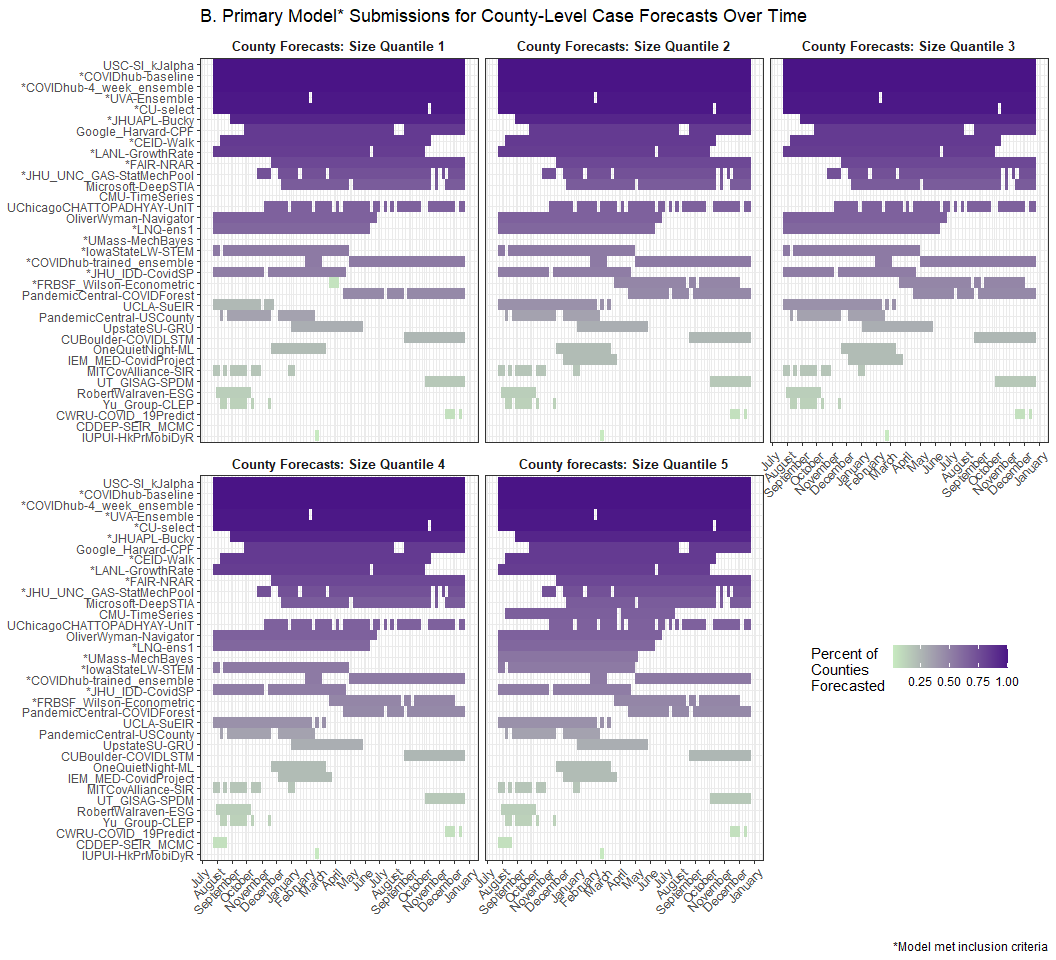

**S1 Table 1.1**. List of models evaluated, including sources for case, hospitalization, death, demographic, and mobility data when used as inputs for the given model. We evaluated 26 models contributed by 24 teams. The COVIDhub team submitted three models including the baseline model and the ensemble model. A brief description is included for each model, with a reference where available. The last column indicates whether the model made assumptions about how and whether social distancing measures were assumed to change during the period for which forecasts were made.

| **Team-Model** | **Data Sources Included** | | | | | **Model Information** | |
| --- | --- | --- | --- | --- | --- | --- | --- |
|  | ***Cases*** | ***Hospitalizations*** | ***Deaths*** | ***Demography*** | ***Mobility*** | ***Description (Reference)*** | ***Assumes future changes to social distancing*** |
| BPagano-RtDriven | J |  |  | J |  | Death-based SIR model that uses the change history of the Covid-19 effective transmission rate to forecast deaths and cases. | No |
| CEID-Walk | J |  |  |  |  | Random walk model starting from the most recent observation with a dispersion based on the spread of the last 5 observations | No |
| COVIDAnalytics-DELPHI | J |  |  | J |  | SEIR model augmented with underdetection and interventions. | Yes |
| COVIDhub-baseline |  |  | J |  |  | Median prediction at all future horizons is equal to the most recent observed incidence | No |
| COVIDhub-4_week_ensemble |  |  |  |  |  | Unweighted average or median of submitted forecasts to the COVID-19 Forecast Hub | n/a |
| COVIDhub-trained_ensemble |  |  |  |  |  | This is a weighted combination of the component model forecasts. All component models that provide forecasts at all required quantile levels for all forecasts at 1 to 4 week ahead horizons (or 1 to 28 day ahead for hospitalizations) are included. | n/a |
| Covid19Sim-Simulator | J | CTP | J |  |  | SEIR model accounting for undiagnosed infections (55) | No |
| CU-select | J | CTP, HHS | J |  | SG | A metapopulation county-level SEIR model for projecting future COVID-19 incidence cases and deaths (56) | Yes |
| FAIR-NRAR | NYT |  |  |  |  | Combine recurrent neural networks with a vector autoregressive model | No |
| FRBSF-Wilson-Econometric | UF |  |  | Cen | MEI | An SIR-derived econometric county panel data model with transmission rate assumed to be function of weather and mobility | No |
| IEM_MED-CovidProject | J |  |  | J |  | SEIR model projections using MCMC to find best parameters to fit actual data. (57) | No |
| IowaStateLW-STEMb | J,NYT |  | J,NYT | Cen | USDT | Nonparametric space-time disease transmission model (58) | No |
| JHUAPL-Bucky | J | HHS | J | Cen | SG, PIQ | Spatial compartment model using public mobility data. Local parameters. | No |
| JHU_CSSE-DECOM | J |  | Cen | J | SG | State-level, empirical machine learning model driven by epidemiological, mobility, demographic, and behavioral data. | No |
| JHU_IDD-CovidSP | J,UF |  | J,UF | Cen | Cen | Metapopulation model with commuting, nonpharmaceutical interventions, and stochastic SEIR disease dynamics (59) | No |
| Karlen-pypm | CTP, HHS | J | J |  |  | Finite time difference equations implemented as a general-purpose population modeling framework (60) | No |
| LNQ-ens1 |  |  |  | J |  | County-level ensemble of boosted tree and neural net models | No |
| LANL-GrowthRate | J |  | J |  |  | Statistical dynamical growth model accounting for population susceptibility (61) | No |
| Microsoft-DeepSTIA | J |  | J | J |  | A deep spatial-temporal network that leverages a hierarchical administration graph and adaptively identified interconnections within the same administrative level (62) | No |
| MOBS-GLEAM_COVID | J | HHS | J | Cen | G | Metapopulation, age-structured SLIR model with mobility and nonpharmaceutical interventions (63) | No |
| RobertWalraven-ESG | J |  | J |  |  | Multiple skewed gaussian mathematical fit | No |
| UCLA-SuEIR | J | CTP | J |  |  | SEIR model variant considering both untested and unreported cases | Yes |
| USC-SIkJalpha | J | HHS | J |  |  | Models temporally varying infection, death, and hospitalization rates. Learning is performed by reducing the problem to multiple simple linear regression problems. (64) | No |
| UMass-MechBayes | J |  | J |  |  | Bayesian compartmental model with observations on incident case counts and incident deaths | No |
| UVA-Ensemble | J |  |  |  |  | An ensemble of multiple methods such as auto-regressive (AR) models with exogenous variables, Long short-term memory (lSTM) models, ensemble Kalman filter (EnKF) and PatchSim (an SEIR model). (65) | No |
| Cen = US Cen (https://www.census.gov/), CTP = COVID Tracking Project (https://covidtracking.com/), G = Google mobility (https://www.google.com/covid19/mobility/), J = JHU CSSE (https://github.com/CSSEGISandData/COVID-19), MEI=Dallas Fed Mobility & Engagement Index (MEI), NYT = New York Times (https://github.com/nytimes/covid-19-data), PIQ = Place IQ (https://github.com/COVIDExposureIndices/COVIDExposureIndices), SEIR = Susceptible-Exposed-Infectious-Recovered compartmental model, SG = SafeGraph mobility (https://www.safegraph.com/), SIR = Susceptible-Infectious-Recovered compartmental model, UF = USA Facts (https://usafacts.org/visualizations/coronavirus-covid-19-spread-map/), USDT = U.S. Department of Transportation Bureau of Transportation Statistics (https://www.transportation.gov/connect/available-datasets); n/a = Not Applicable | | | | | | | |
