## Supplement 2 for "Challenges of COVID-19 Case Forecasting in the US, 2020-2021"

**Supporting Information 2**: Revision and outlier sensitivity analyses

**S2 Figure 2.2.** After identifying weeks with revised case counts, we then excluded them from the dataset and reran the GEE models and estimated the marginal mean Weighted Interval Score (WIS). Panel A shows the estimated marginal mean WIS and 95% confidence intervals for mean cases from team-specific GEE models for all 48 jurisdictions from this sensitivity analysis. The 95% confidence intervals for the COVIDhub-baseline model are shown in dashed red vertical lines. Panel B presents each team’s estimated marginal mean WIS per phase, scaled to the COVIDhub-baseline model’s estimated marginal mean WIS for all epidemic phases, using the dataset with excluded week. Teams with higher estimated marginal mean WIS values (i.e., greater than 1.0) are presented in shades of orange while teams with lower estimated marginal mean WIS (i.e., less than 1.0) are shown in shades of green***.*** Team forecasts are denoted with an asterisk (*) if the 80% confidence interval of the expected WIS outcome (normalized and on the log scale) was estimated by a model to be lower than the expected WIS of the COVIDhub-baseline model for all phases.

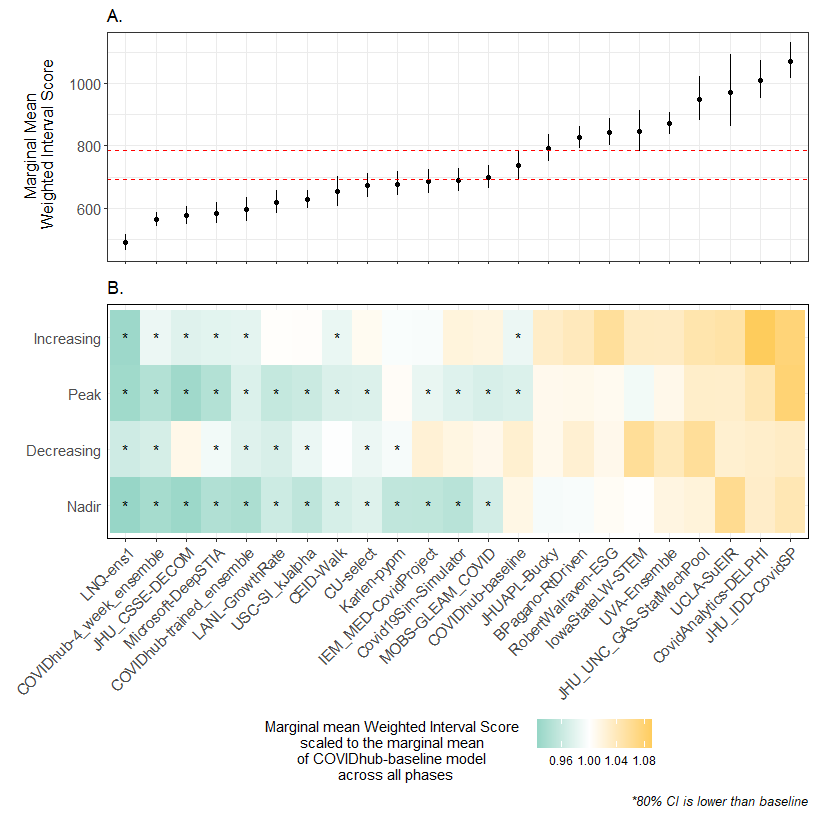

**S2 Figure 2.3**. Outliers were defined as non-revised reported case counts that were outside of the expected range by at least two of the three algorithms: a rolling median, a seasonal trend decomposition, and a seasonal trend decomposition without a seasonality term. Each method used a 21-day window. Approximately three percent of weeks (686 of 27,489 total weeks in the analysis period) had at least one day of reported cases identified as an outlier.

After identifying outlier counts, we then excluded weeks with outliers and the week following an outlier from the dataset and reran the GEE models and estimated the marginal mean Weighted Interval Score (WIS). Panel A shows the estimated marginal mean WIS and 95% confidence intervals for mean cases from team-specific GEE models for all 51 jurisdictions from this sensitivity analysis. The 95% confidence intervals for the COVIDhub-baseline model are shown in dashed red vertical lines. Panel B presents each team’s estimated marginal mean WIS per phase, scaled to the COVIDhub-baseline model’s estimated marginal mean WIS for all epidemic phases, using the dataset with excluded week. Teams with higher estimated marginal mean WIS values (i.e., greater than 1.0) are presented in shades of orange while teams with lower estimated marginal mean WIS (i.e., less than 1.0) are shown in shades of green***.*** WIS values are denoted with an asterisk (*) if the 80% confidence interval of the expected WIS outcome (normalized and on the log scale) was estimated by a model to be lower than the expected WIS of the COVIDhub-baseline model for all phases.

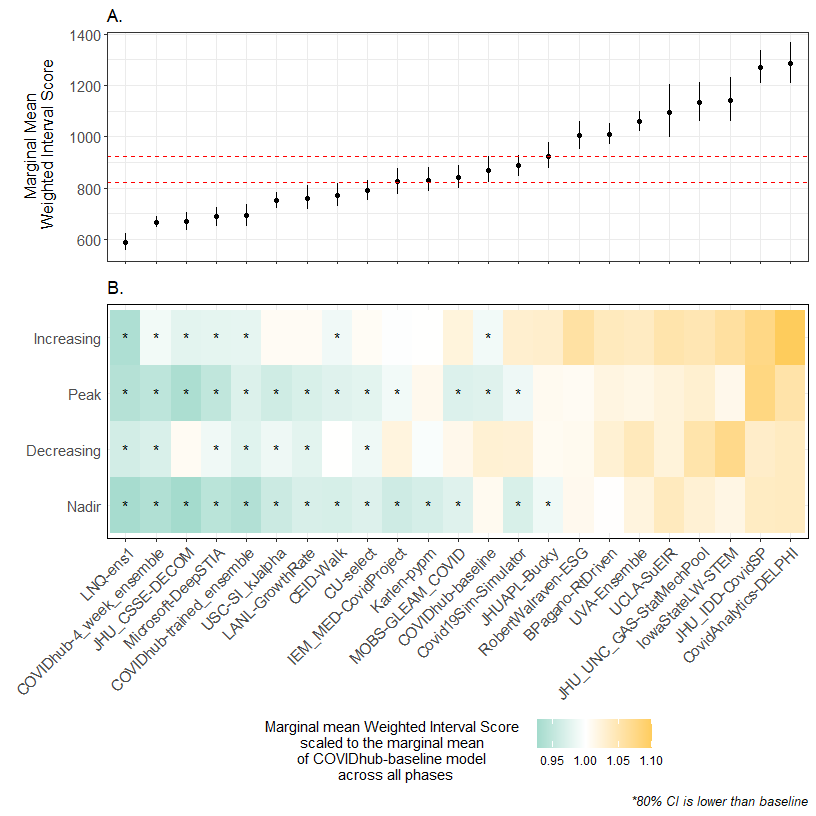
