## Supplemental Figure 2.1 for "Challenges of COVID-19 Case Forecasting in the US, 2020-2021"

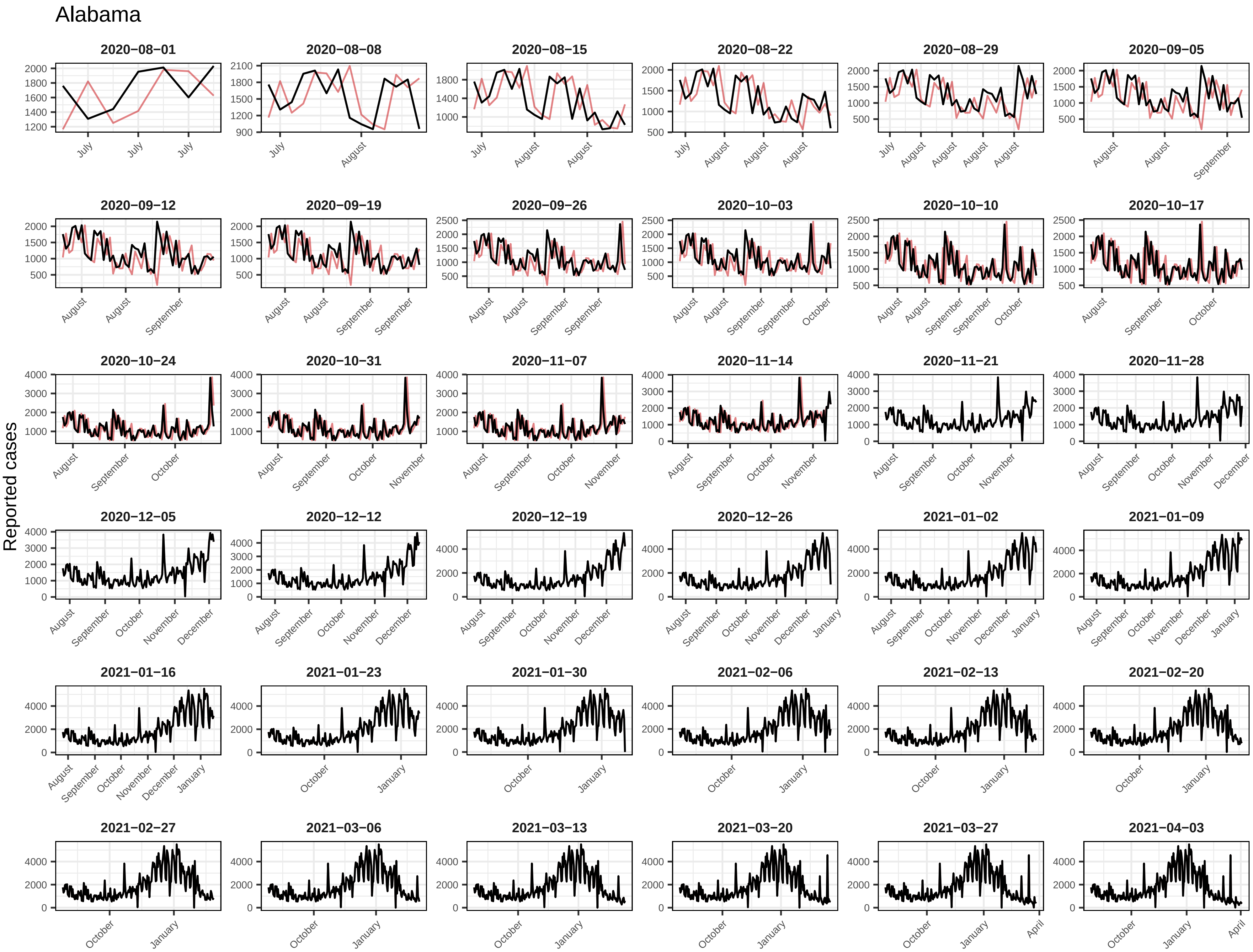
Alabama
Reported cases
2020−08−01
2020−08−08
2020−08−15
2020−08−22
2020−08−29
2020−09−05
2020−09−12
2020−09−19
2020−09−26
2020−10−03
2020−10−10
2020−10−17
2020−10−24
2020−10−31
2020−11−07
2020−11−14
2020−11−21
2020−11−28
2020−12−05
2020−12−12
2020−12−19
2020−12−26
2021−01−02
2021−01−09
2021−01−16
2021−01−23
2021−01−30
2021−02−06
2021−02−13
2021−02−20
2021−02−27
2021−03−06
2021−03−13
2021−03−20
2021−03−27
2021−04−03

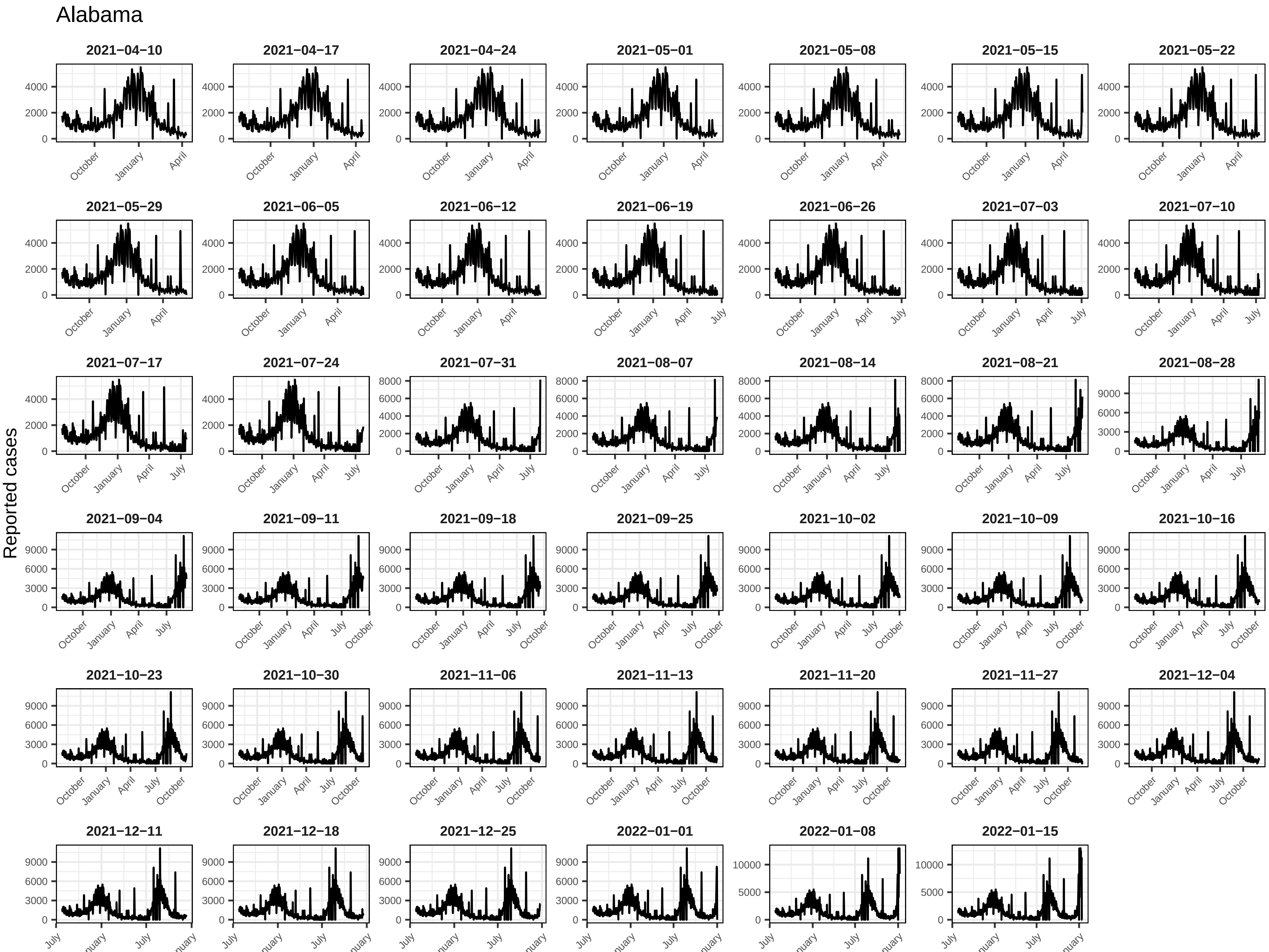
Alabama
Reported cases
2021−04−10
2021−04−17
2021−04−24
2021−05−01
2021−05−08
2021−05−15
2021−05−22
2021−05−29
2021−06−05
2021−06−12
2021−06−19
2021−06−26
2021−07−03
2021−07−10
2021−07−17
2021−07−24
2021−07−31
2021−08−07
2021−08−14
2021−08−21
2021−08−28
2021−09−04
2021−09−11
2021−09−18
2021−09−25
2021−10−02
2021−10−09
2021−10−16
2021−10−23
2021−10−30
2021−11−06
2021−11−13
2021−11−20
2021−11−27
2021−12−04
2021−12−11
2021−12−18
2021−12−25
2022−01−01
2022−01−08
2022−01−15
0
2000
4000
6000
8000
3000
9000
5000
10000
October
January
April
July

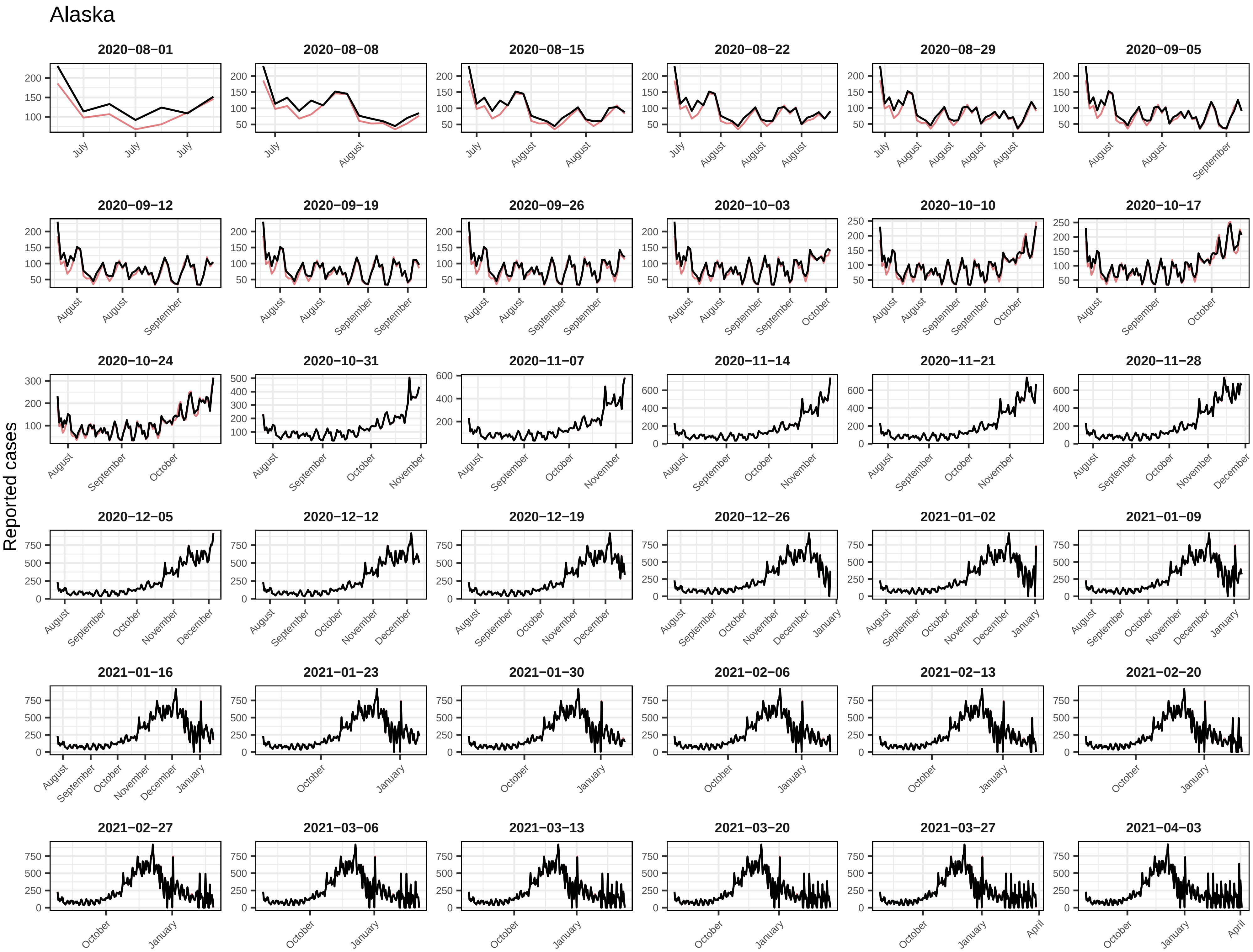
Alaska
Reported cases
2020−08−01
2020−08−08
2020−08−15
2020−08−22
2020−08−29
2020−09−05
2020−09−12
2020−09−19
2020−09−26
2020−10−03
2020−10−10
2020−10−17
2020−10−24
2020−10−31
2020−11−07
2020−11−14
2020−11−21
2020−11−28
2020−12−05
2020−12−12
2020−12−19
2020−12−26
2021−01−02
2021−01−09
2021−01−16
2021−01−23
2021−01−30
2021−02−06
2021−02−13
2021−02−20
2021−02−27
2021−03−06
2021−03−13
2021−03−20
2021−03−27
2021−04−03

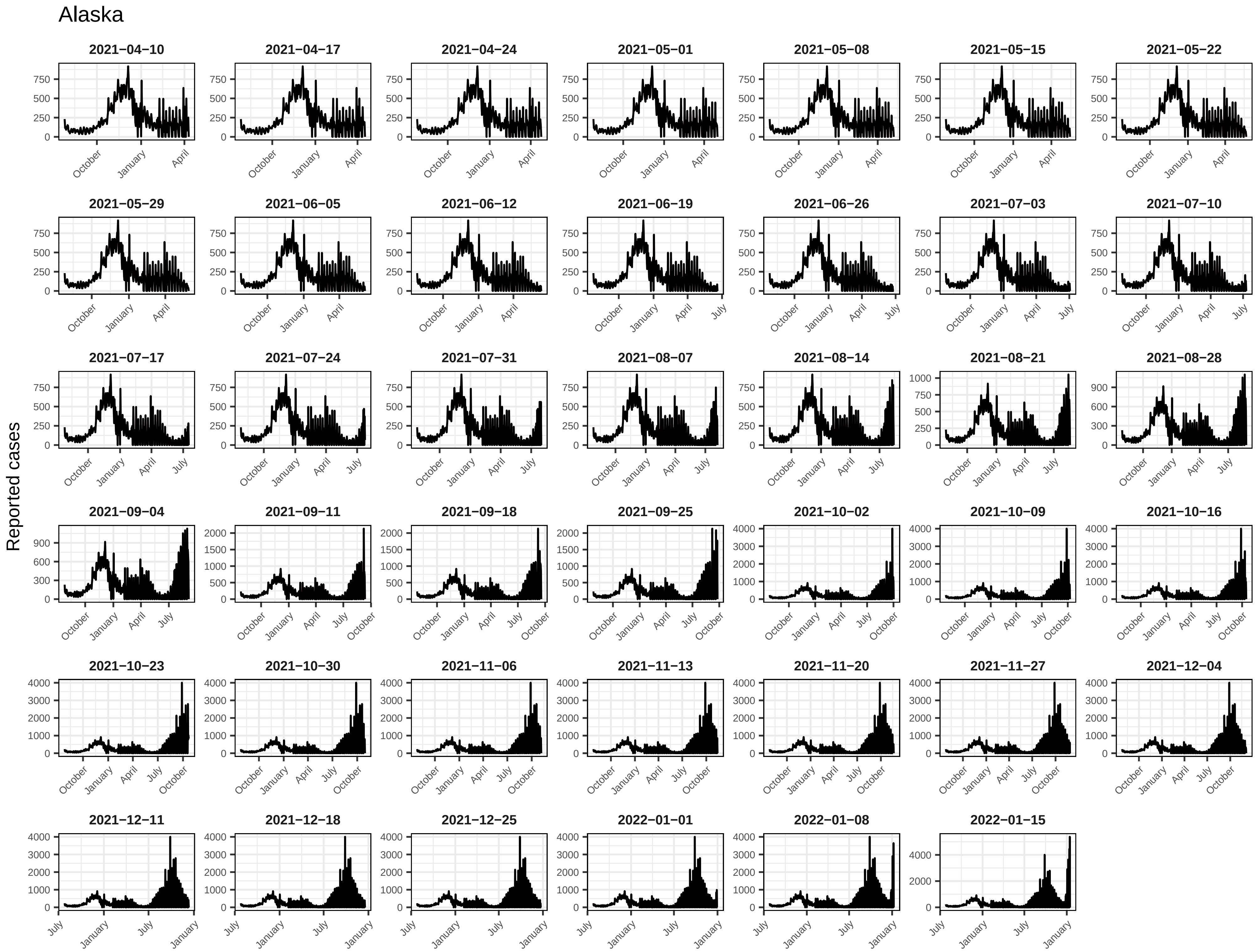
Alaska
Reported cases
2021−04−10
2021−04−17
2021−04−24
2021−05−01
2021−05−08
2021−05−15
2021−05−22
2021−05−29
2021−06−05
2021−06−12
2021−06−19
2021−06−26
2021−07−03
2021−07−10
2021−07−17
2021−07−24
2021−07−31
2021−08−07
2021−08−14
2021−08−21
2021−08−28
2021−09−04
2021−09−11
2021−09−18
2021−09−25
2021−10−02
2021−10−09
2021−10−16
2021−10−23
2021−10−30
2021−11−06
2021−11−13
2021−11−20
2021−11−27
2021−12−04
2021−12−11
2021−12−18
2021−12−25
2022−01−01
2022−01−08
2022−01−15
October
January
April
July

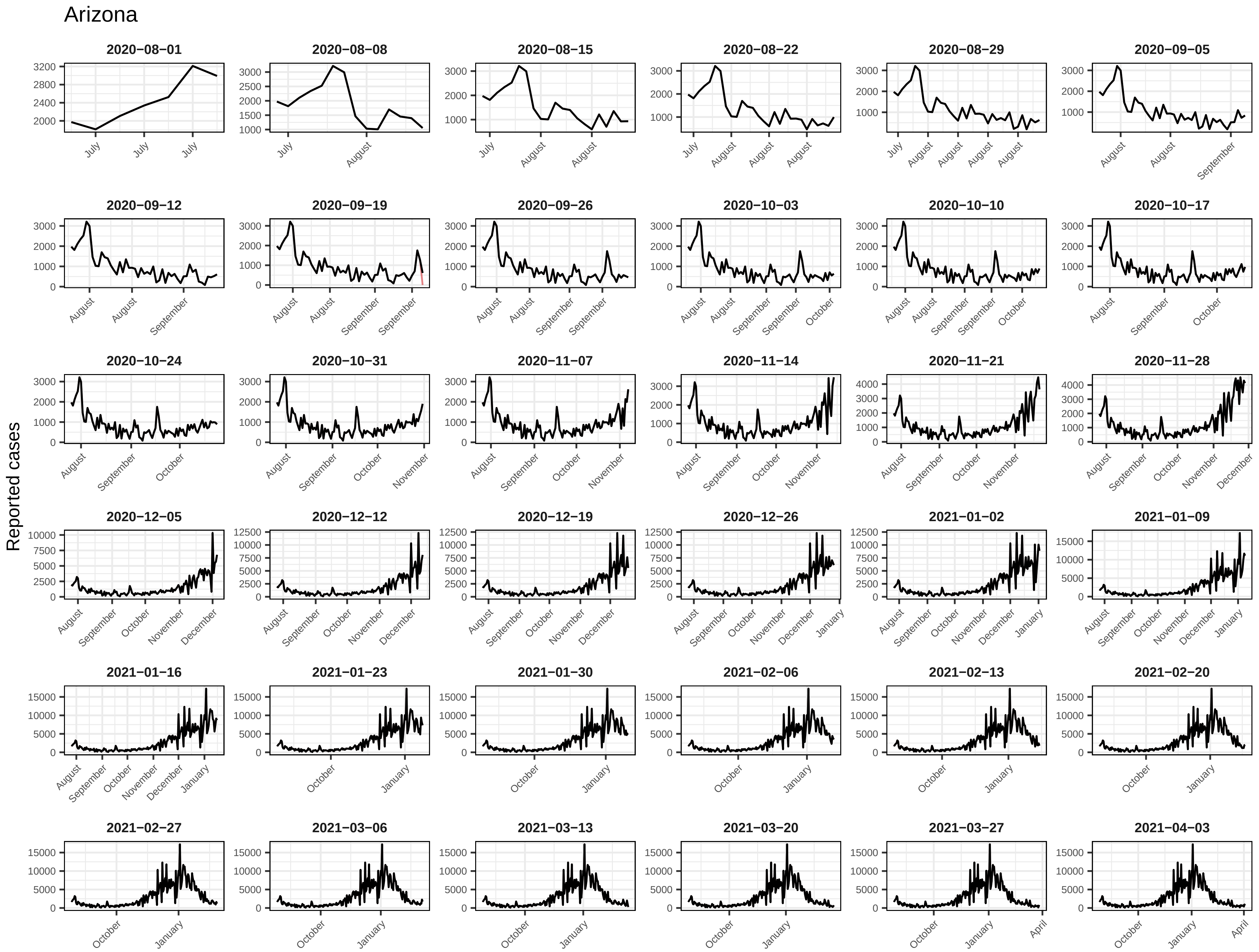
Arizona
Reported cases
2020−08−01
2020−08−08
2020−08−15
2020−08−22
2020−08−29
2020−09−05
2020−09−12
2020−09−19
2020−09−26
2020−10−03
2020−10−10
2020−10−17
2020−10−24
2020−10−31
2020−11−07
2020−11−14
2020−11−21
2020−11−28
2020−12−05
2020−12−12
2020−12−19
2020−12−26
2021−01−02
2021−01−09
2021−01−16
2021−01−23
2021−01−30
2021−02−06
2021−02−13
2021−02−20
2021−02−27
2021−03−06
2021−03−13
2021−03−20
2021−03−27
2021−04−03

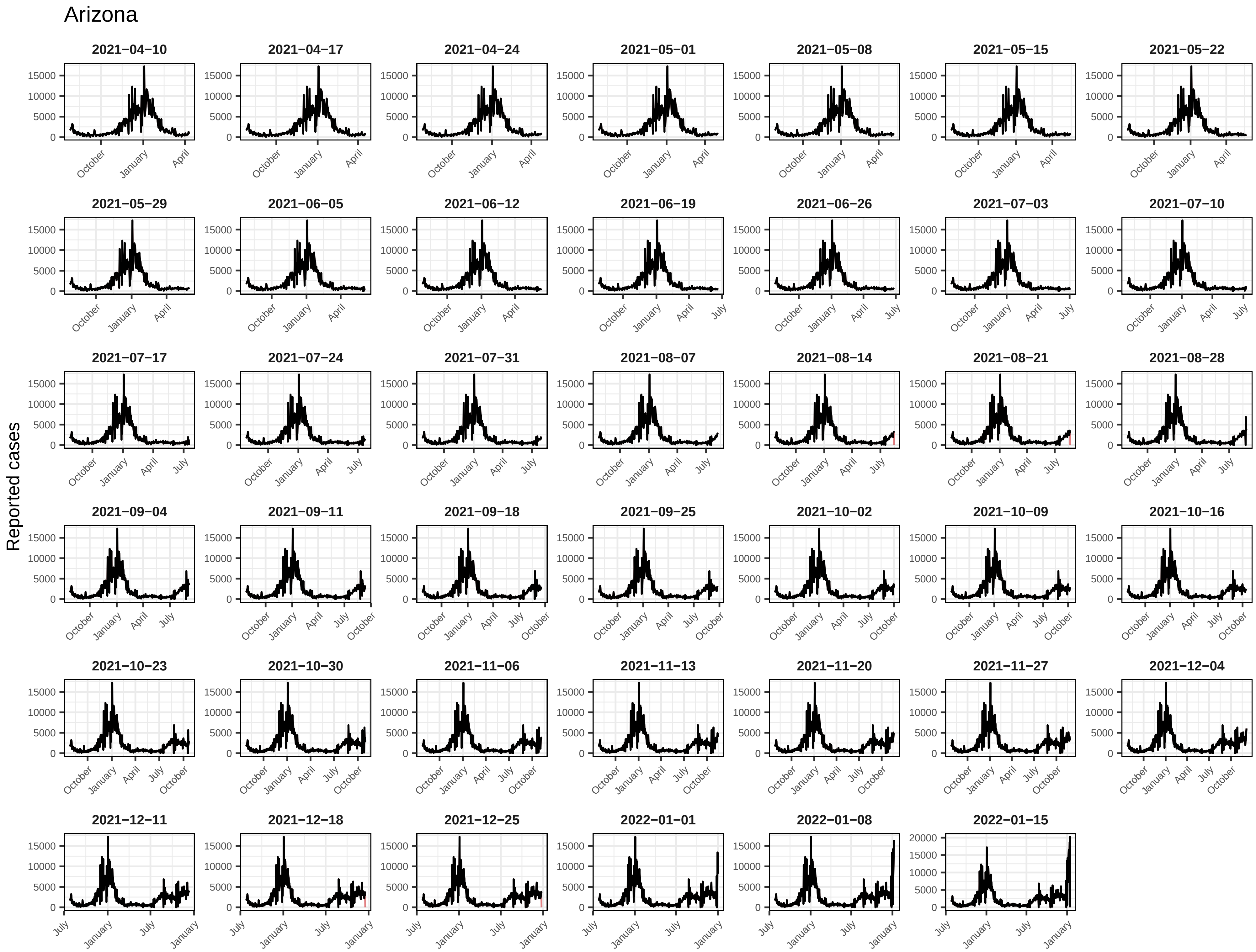
Arizona
Reported cases
2021−04−10
2021−04−17
2021−04−24
2021−05−01
2021−05−08
2021−05−15
2021−05−22
2021−05−29
2021−06−05
2021−06−12
2021−06−19
2021−06−26
2021−07−03
2021−07−10
2021−07−17
2021−07−24
2021−07−31
2021−08−07
2021−08−14
2021−08−21
2021−08−28
2021−09−04
2021−09−11
2021−09−18
2021−09−25
2021−10−02
2021−10−09
2021−10−16
2021−10−23
2021−10−30
2021−11−06
2021−11−13
2021−11−20
2021−11−27
2021−12−04
2021−12−11
2021−12−18
2021−12−25
2022−01−01
2022−01−08
2022−01−15
0
5000
10000
15000
20000
October
January
April
July

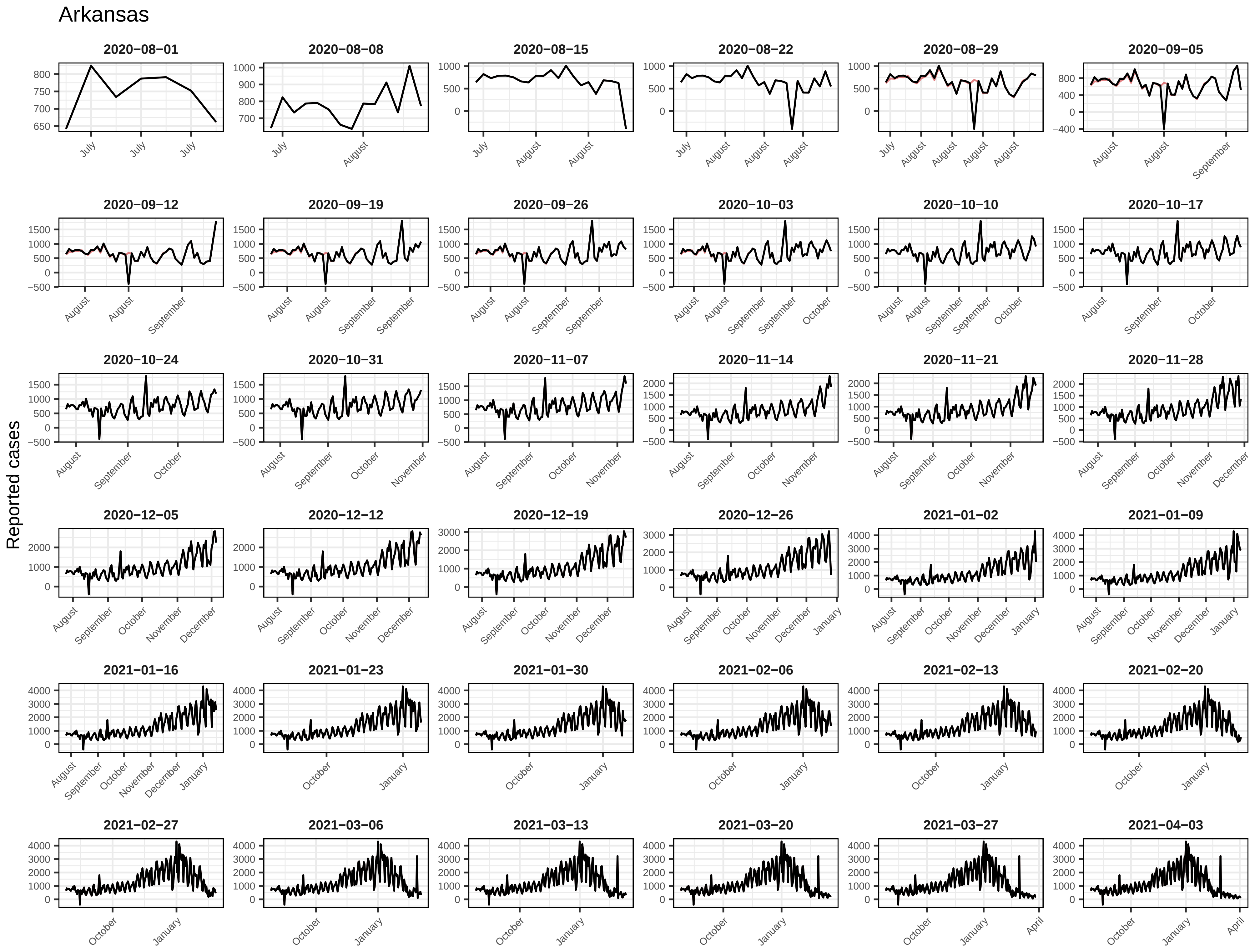
Arkansas
Reported cases
2020−08−01
2020−08−08
2020−08−15
2020−08−22
2020−08−29
2020−09−05
2020−09−12
2020−09−19
2020−09−26
2020−10−03
2020−10−10
2020−10−17
2020−10−24
2020−10−31
2020−11−07
2020−11−14
2020−11−21
2020−11−28
2020−12−05
2020−12−12
2020−12−19
2020−12−26
2021−01−02
2021−01−09
2021−01−16
2021−01−23
2021−01−30
2021−02−06
2021−02−13
2021−02−20
2021−02−27
2021−03−06
2021−03−13
2021−03−20
2021−03−27
2021−04−03

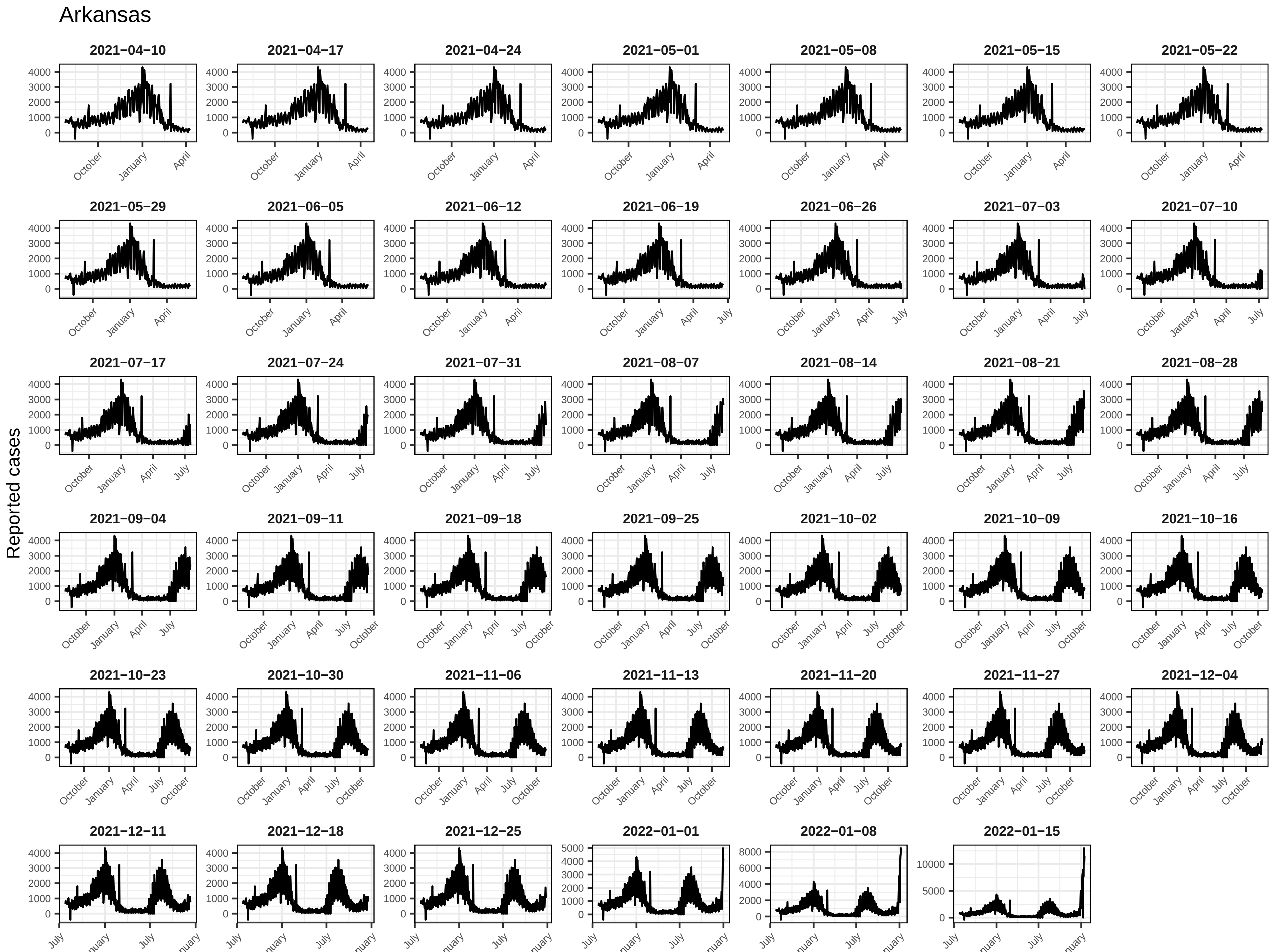
Arkansas
Reported cases
2021−04−10
2021−04−17
2021−04−24
2021−05−01
2021−05−08
2021−05−15
2021−05−22
2021−05−29
2021−06−05
2021−06−12
2021−06−19
2021−06−26
2021−07−03
2021−07−10
2021−07−17
2021−07−24
2021−07−31
2021−08−07
2021−08−14
2021−08−21
2021−08−28
2021−09−04
2021−09−11
2021−09−18
2021−09−25
2021−10−02
2021−10−09
2021−10−16
2021−10−23
2021−10−30
2021−11−06
2021−11−13
2021−11−20
2021−11−27
2021−12−04
2021−12−11
2021−12−18
2021−12−25
2022−01−01
2022−01−08
2022−01−15
October
January
April
July

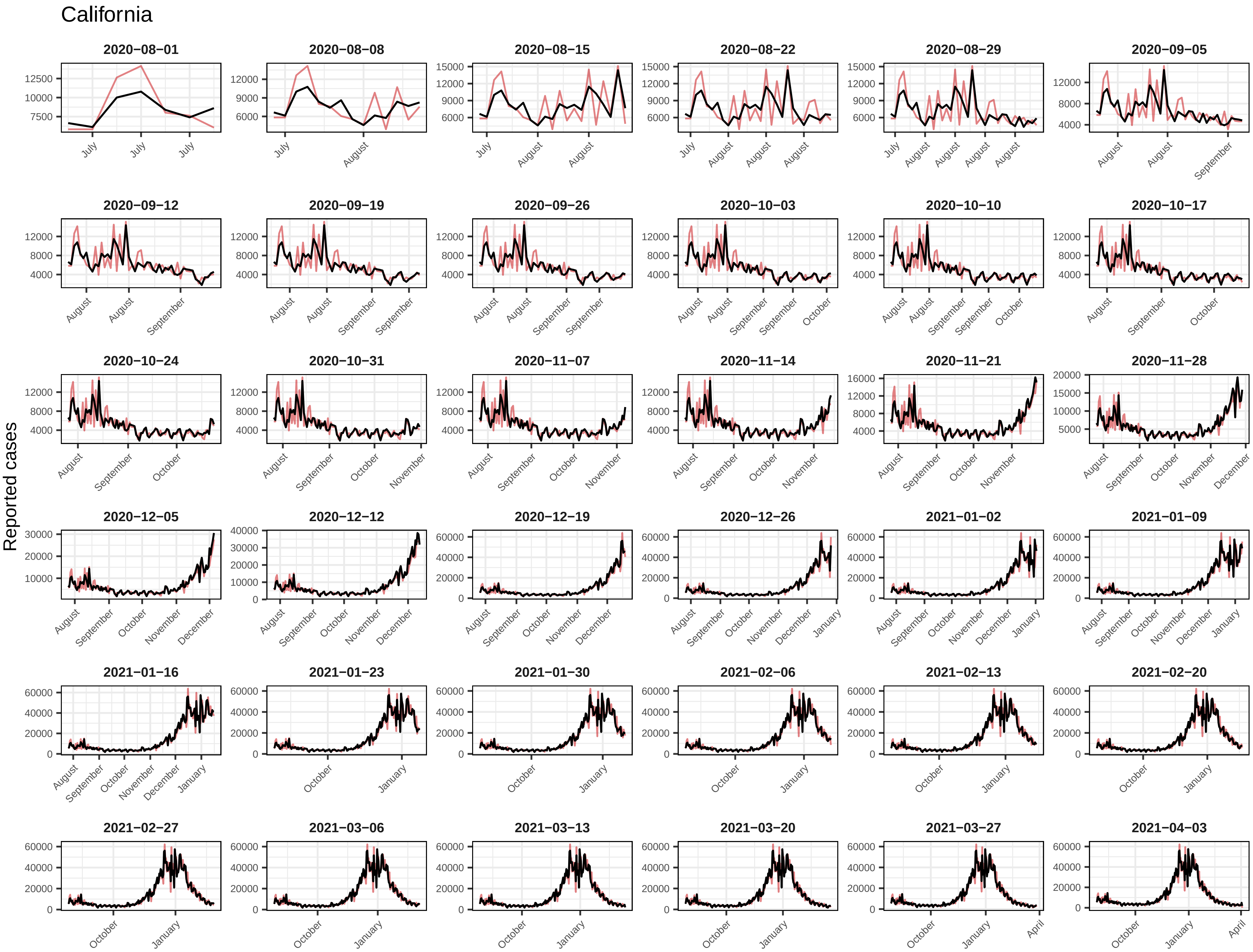
California
Reported cases
2020−08−01
2020−08−08
2020−08−15
2020−08−22
2020−08−29
2020−09−05
2020−09−12
2020−09−19
2020−09−26
2020−10−03
2020−10−10
2020−10−17
2020−10−24
2020−10−31
2020−11−07
2020−11−14
2020−11−21
2020−11−28
2020−12−05
2020−12−12
2020−12−19
2020−12−26
2021−01−02
2021−01−09
2021−01−16
2021−01−23
2021−01−30
2021−02−06
2021−02−13
2021−02−20
2021−02−27
2021−03−06
2021−03−13
2021−03−20
2021−03−27
2021−04−03

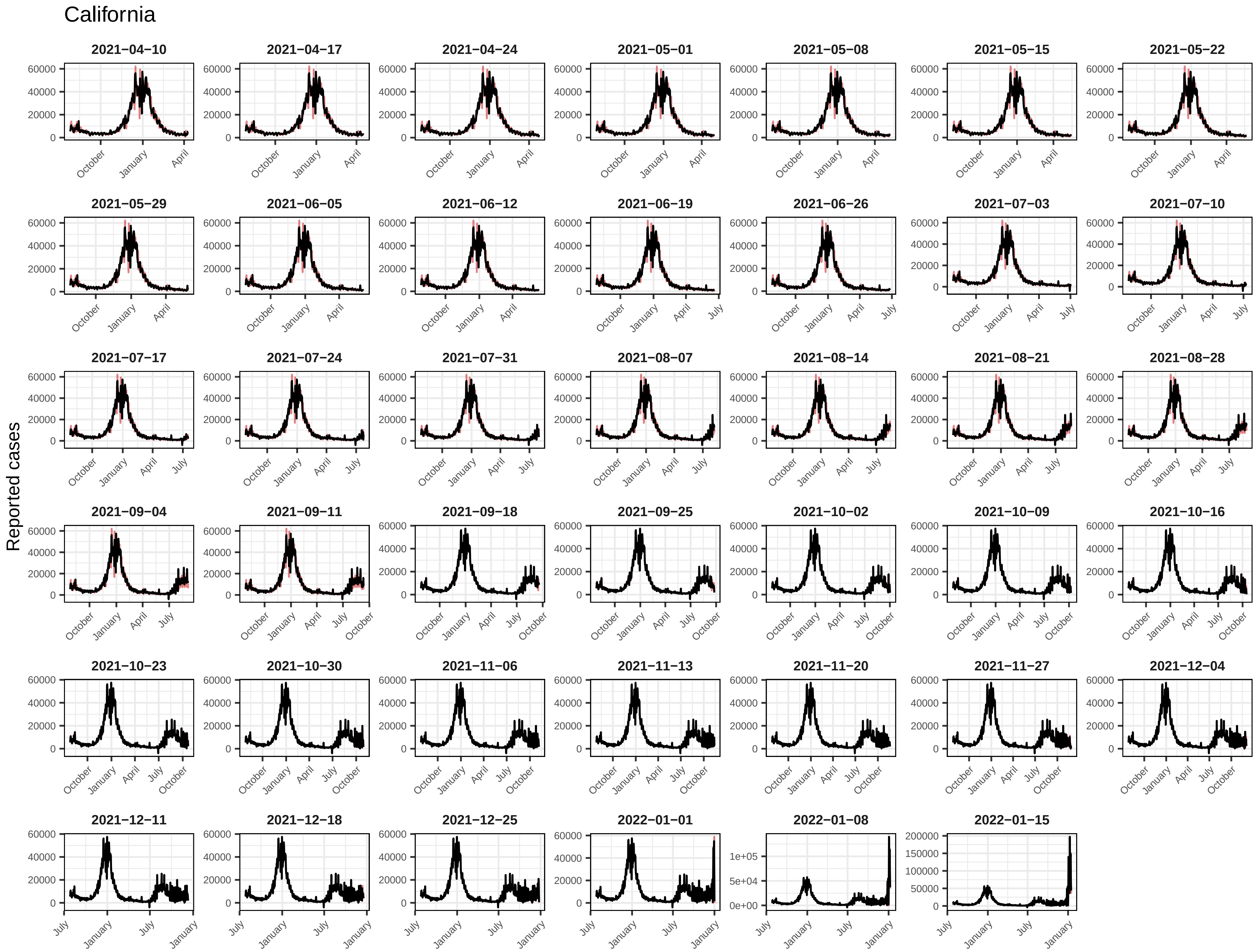
California
Reported cases
2021−04−10
2021−04−17
2021−04−24
2021−05−01
2021−05−08
2021−05−15
2021−05−22
2021−05−29
2021−06−05
2021−06−12
2021−06−19
2021−06−26
2021−07−03
2021−07−10
2021−07−17
2021−07−24
2021−07−31
2021−08−07
2021−08−14
2021−08−21
2021−08−28
2021−09−04
2021−09−11
2021−09−18
2021−09−25
2021−10−02
2021−10−09
2021−10−16
2021−10−23
2021−10−30
2021−11−06
2021−11−13
2021−11−20
2021−11−27
2021−12−04
2021−12−11
2021−12−18
2021−12−25
2022−01−01
2022−01−08
2022−01−15

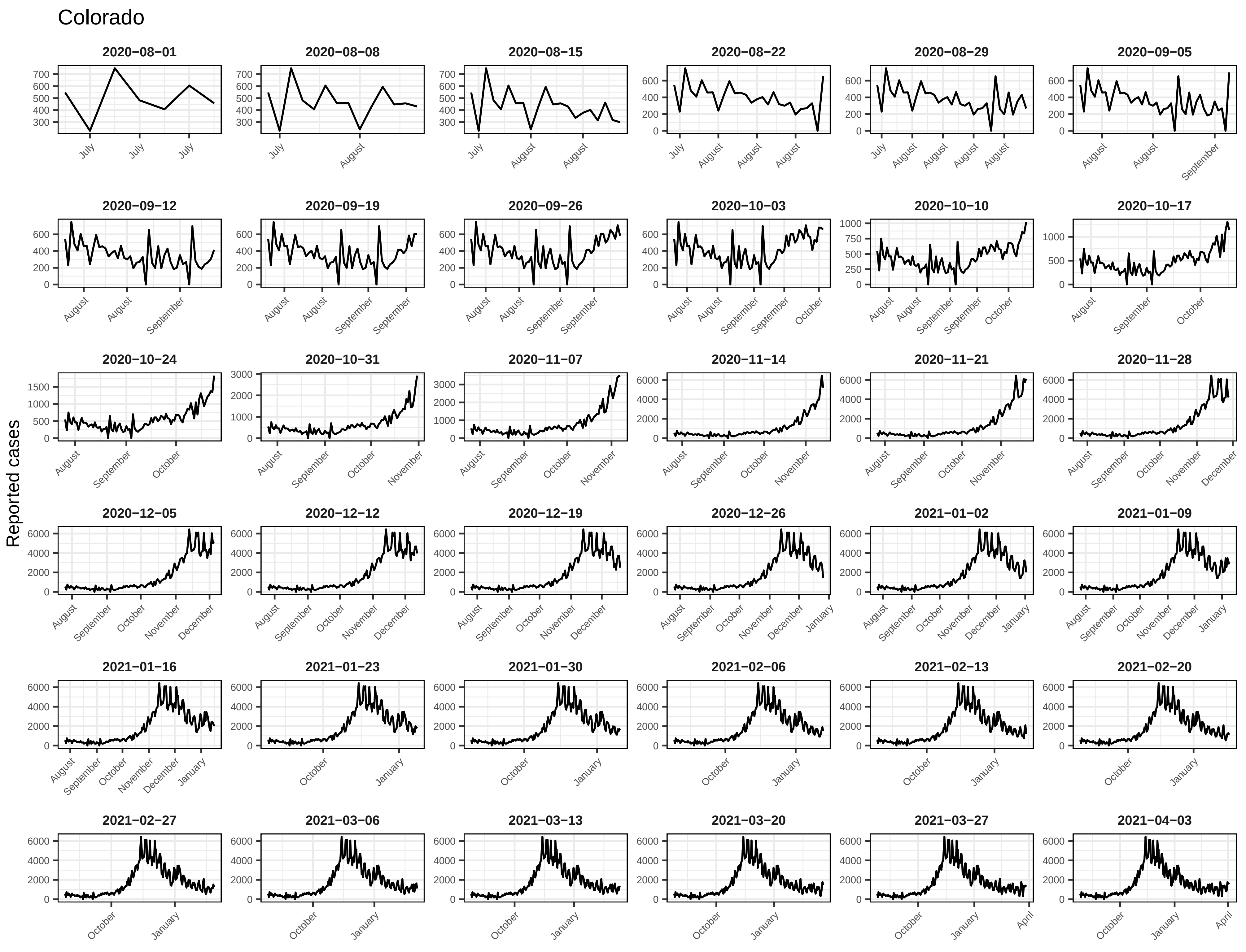
Colorado
Reported cases
2020−08−01
2020−08−08
2020−08−15
2020−08−22
2020−08−29
2020−09−05
2020−09−12
2020−09−19
2020−09−26
2020−10−03
2020−10−10
2020−10−17
2020−10−24
2020−10−31
2020−11−07
2020−11−14
2020−11−21
2020−11−28
2020−12−05
2020−12−12
2020−12−19
2020−12−26
2021−01−02
2021−01−09
2021−01−16
2021−01−23
2021−01−30
2021−02−06
2021−02−13
2021−02−20
2021−02−27
2021−03−06
2021−03−13
2021−03−20
2021−03−27
2021−04−03

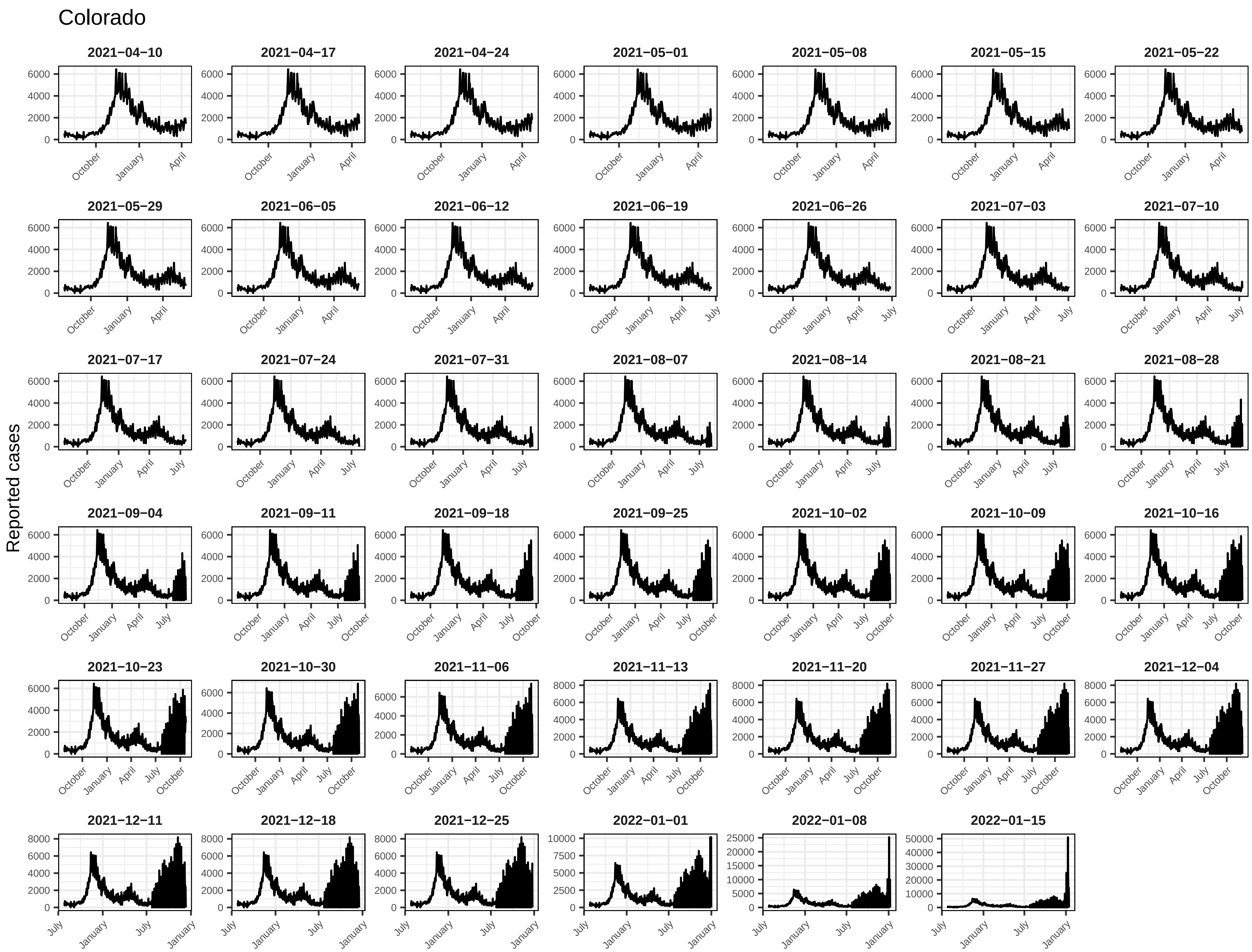
Colorado
Reported cases
2021−04−10
2021−04−17
2021−04−24
2021−05−01
2021−05−08
2021−05−15
2021−05−22
2021−05−29
2021−06−05
2021−06−12
2021−06−19
2021−06−26
2021−07−03
2021−07−10
2021−07−17
2021−07−24
2021−07−31
2021−08−07
2021−08−14
2021−08−21
2021−08−28
2021−09−04
2021−09−11
2021−09−18
2021−09−25
2021−10−02
2021−10−09
2021−10−16
2021−10−23
2021−10−30
2021−11−06
2021−11−13
2021−11−20
2021−11−27
2021−12−04
2021−12−11
2021−12−18
2021−12−25
2022−01−01
2022−01−08
2022−01−15
October
January
April
July

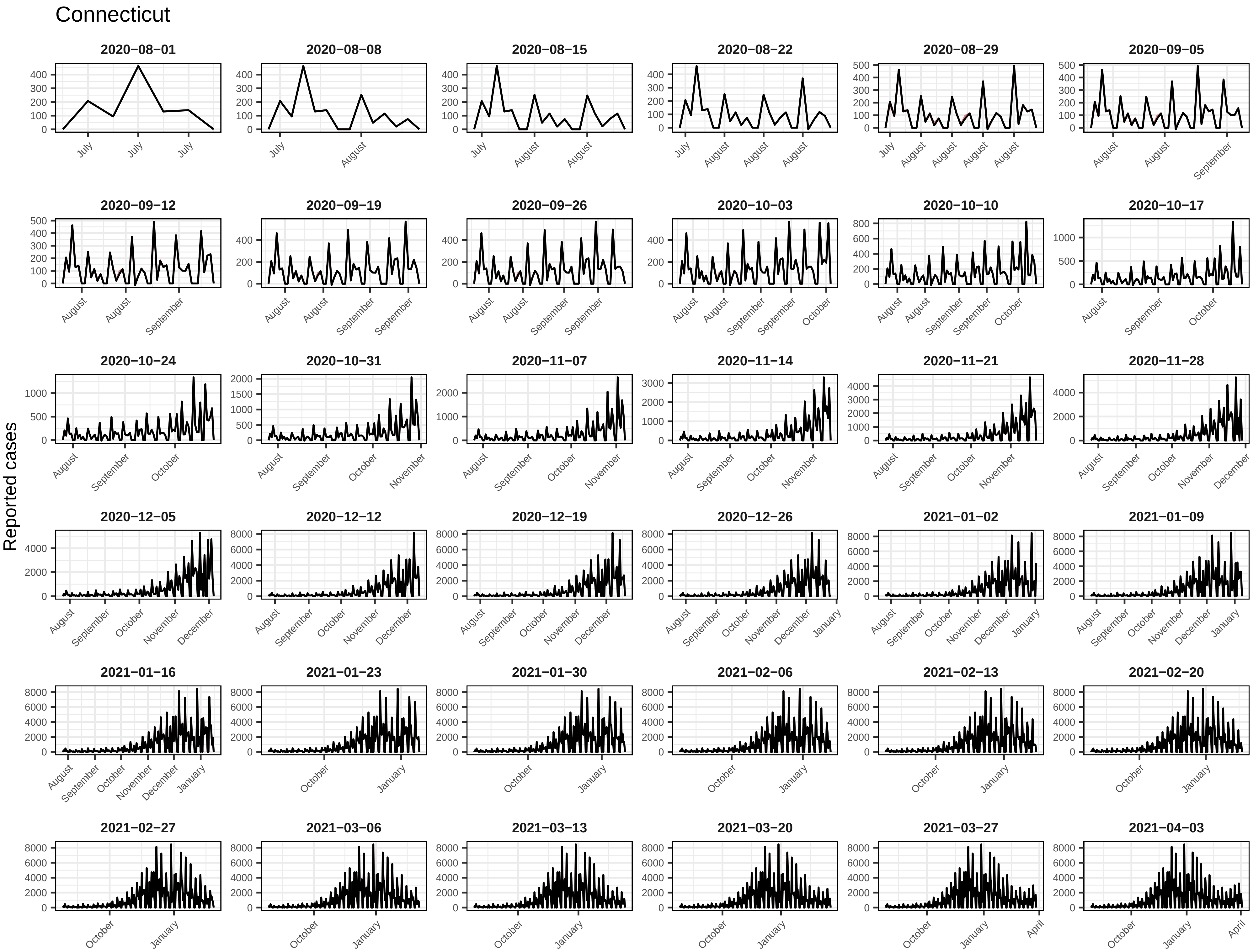
Connecticut
Reported cases
2020−08−01
2020−08−08
2020−08−15
2020−08−22
2020−08−29
2020−09−05
2020−09−12
2020−09−19
2020−09−26
2020−10−03
2020−10−10
2020−10−17
2020−10−24
2020−10−31
2020−11−07
2020−11−14
2020−11−21
2020−11−28
2020−12−05
2020−12−12
2020−12−19
2020−12−26
2021−01−02
2021−01−09
2021−01−16
2021−01−23
2021−01−30
2021−02−06
2021−02−13
2021−02−20
2021−02−27
2021−03−06
2021−03−13
2021−03−20
2021−03−27
2021−04−03

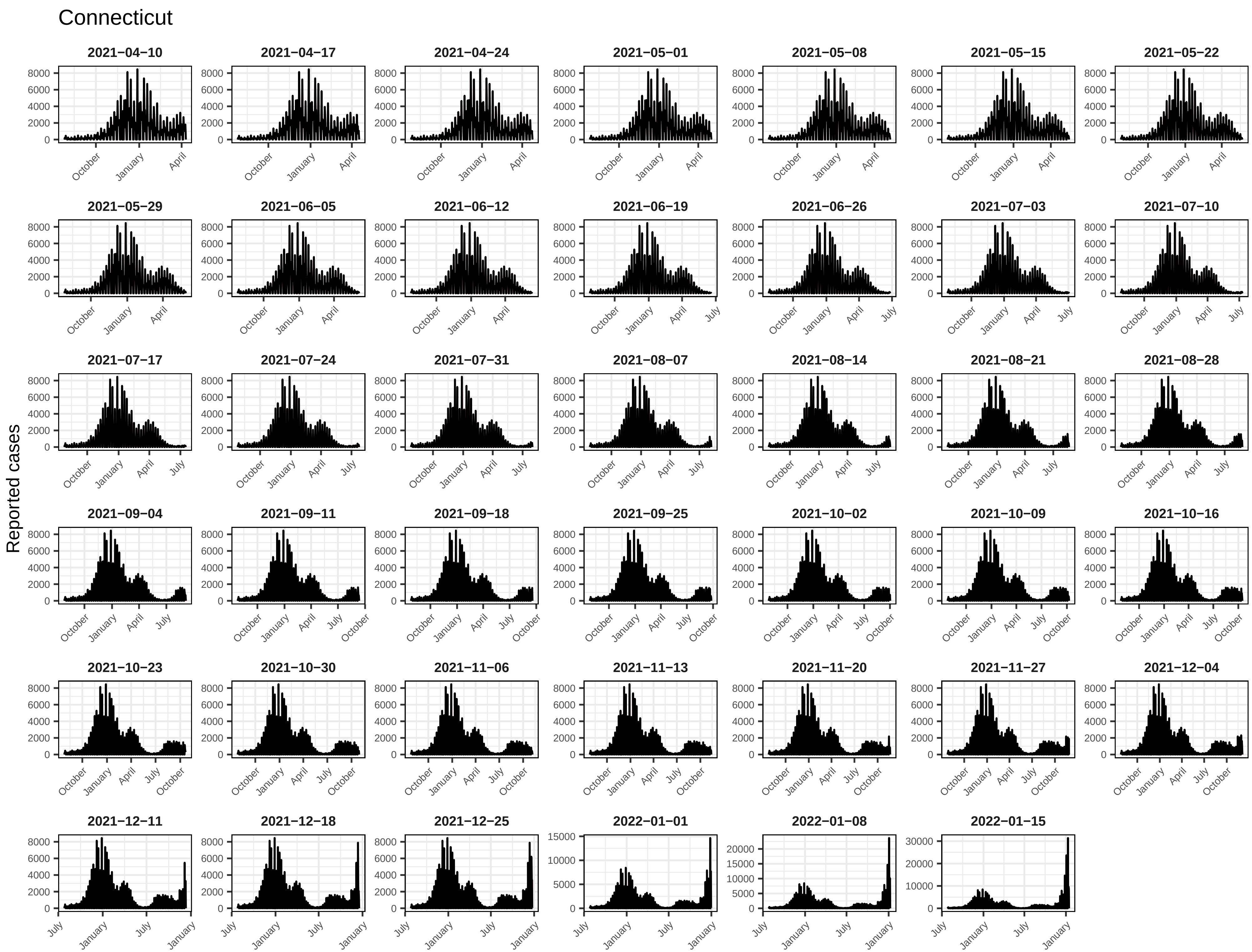
Connecticut
Reported cases
2021−04−10
2021−04−17
2021−04−24
2021−05−01
2021−05−08
2021−05−15
2021−05−22
2021−05−29
2021−06−05
2021−06−12
2021−06−19
2021−06−26
2021−07−03
2021−07−10
2021−07−17
2021−07−24
2021−07−31
2021−08−07
2021−08−14
2021−08−21
2021−08−28
2021−09−04
2021−09−11
2021−09−18
2021−09−25
2021−10−02
2021−10−09
2021−10−16
2021−10−23
2021−10−30
2021−11−06
2021−11−13
2021−11−20
2021−11−27
2021−12−04
2021−12−11
2021−12−18
2021−12−25
2022−01−01
2022−01−08
2022−01−15
October
January
April
July

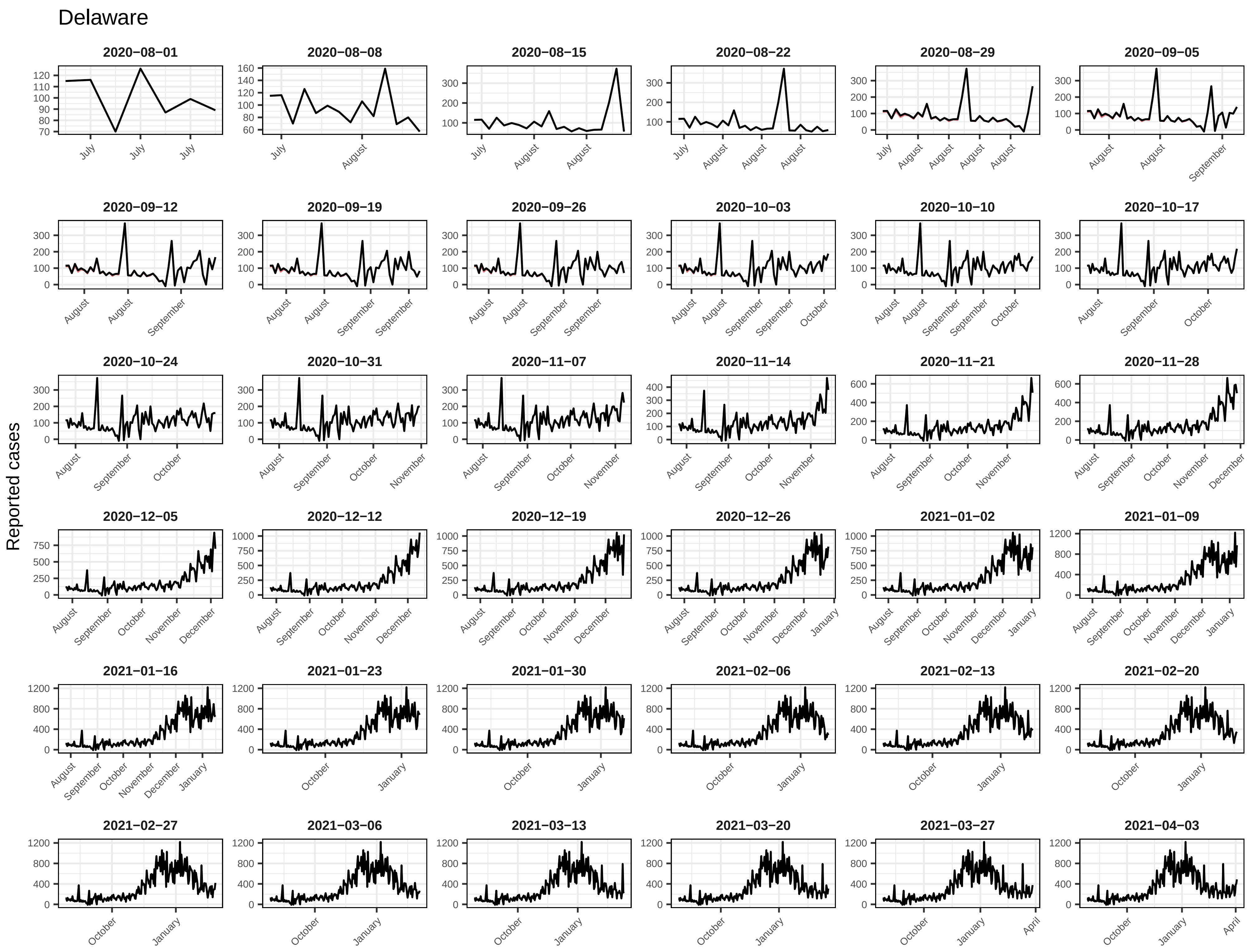
Delaware
Reported cases
2020−08−01
2020−08−08
2020−08−15
2020−08−22
2020−08−29
2020−09−05
2020−09−12
2020−09−19
2020−09−26
2020−10−03
2020−10−10
2020−10−17
2020−10−24
2020−10−31
2020−11−07
2020−11−14
2020−11−21
2020−11−28
2020−12−05
2020−12−12
2020−12−19
2020−12−26
2021−01−02
2021−01−09
2021−01−16
2021−01−23
2021−01−30
2021−02−06
2021−02−13
2021−02−20
2021−02−27
2021−03−06
2021−03−13
2021−03−20
2021−03−27
2021−04−03

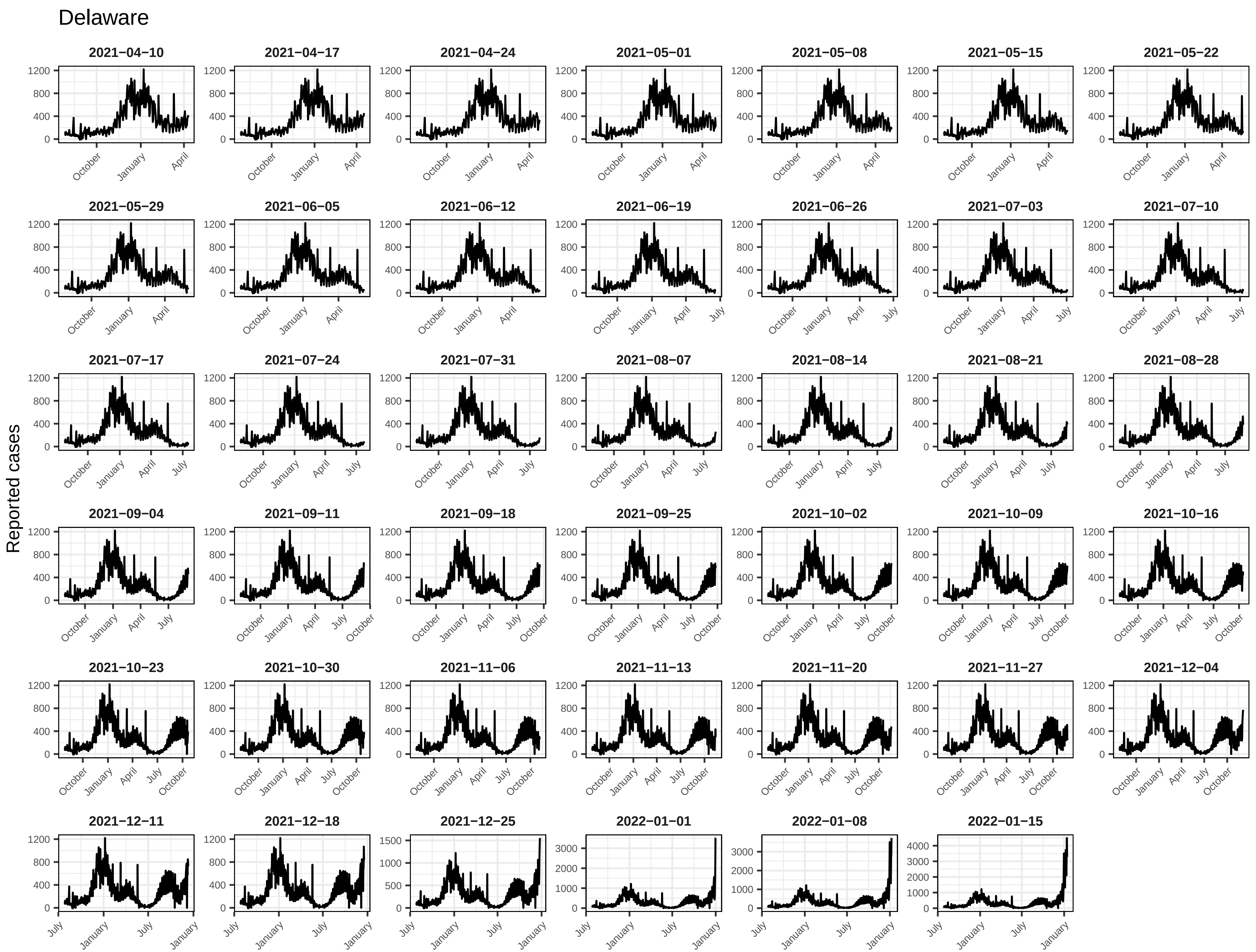
Delaware
Reported cases
2021−04−10
2021−04−17
2021−04−24
2021−05−01
2021−05−08
2021−05−15
2021−05−22
2021−05−29
2021−06−05
2021−06−12
2021−06−19
2021−06−26
2021−07−03
2021−07−10
2021−07−17
2021−07−24
2021−07−31
2021−08−07
2021−08−14
2021−08−21
2021−08−28
2021−09−04
2021−09−11
2021−09−18
2021−09−25
2021−10−02
2021−10−09
2021−10−16
2021−10−23
2021−10−30
2021−11−06
2021−11−13
2021−11−20
2021−11−27
2021−12−04
2021−12−11
2021−12−18
2021−12−25
2022−01−01
2022−01−08
2022−01−15

District of Columbia

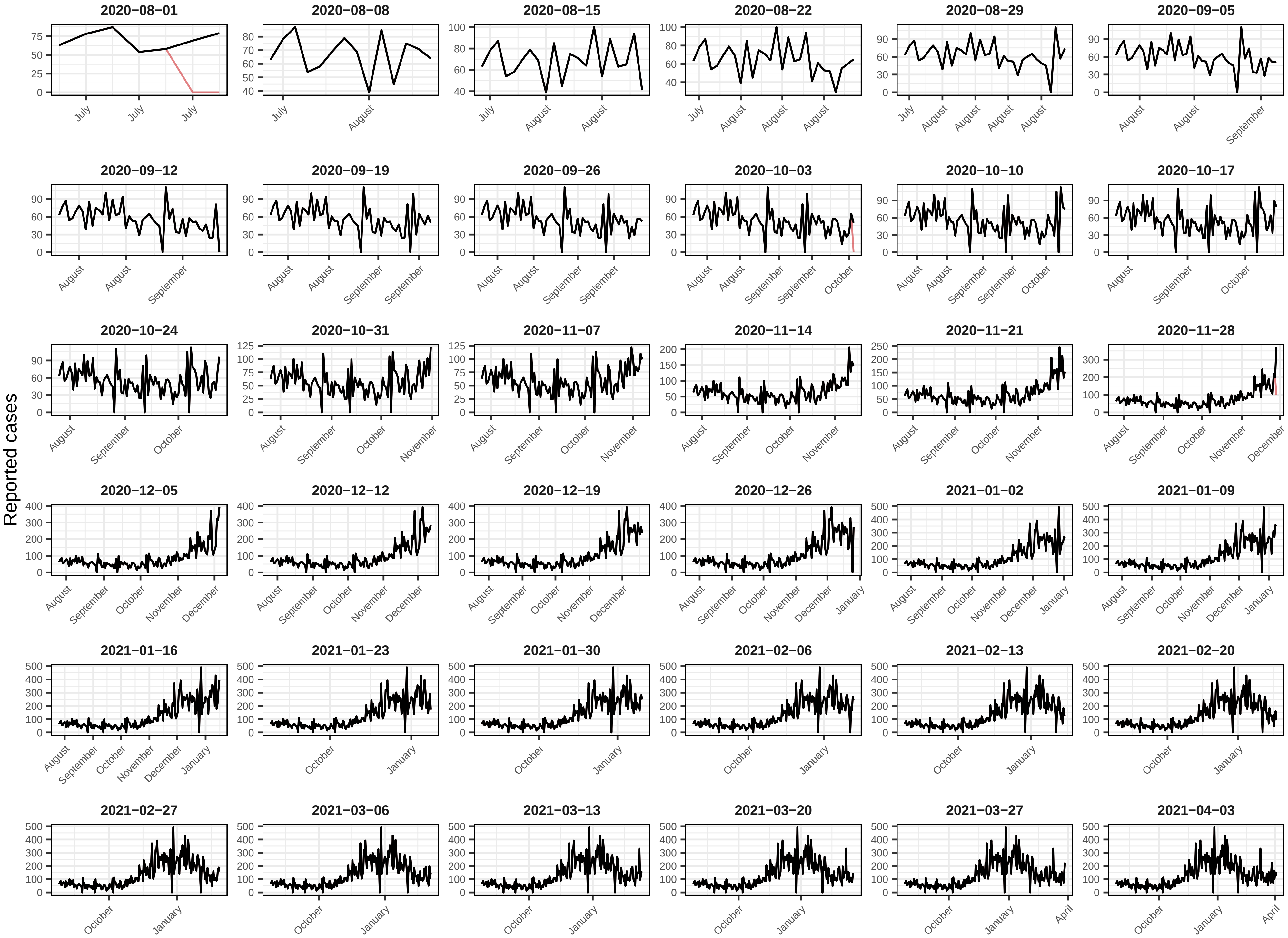

### District of Columbia

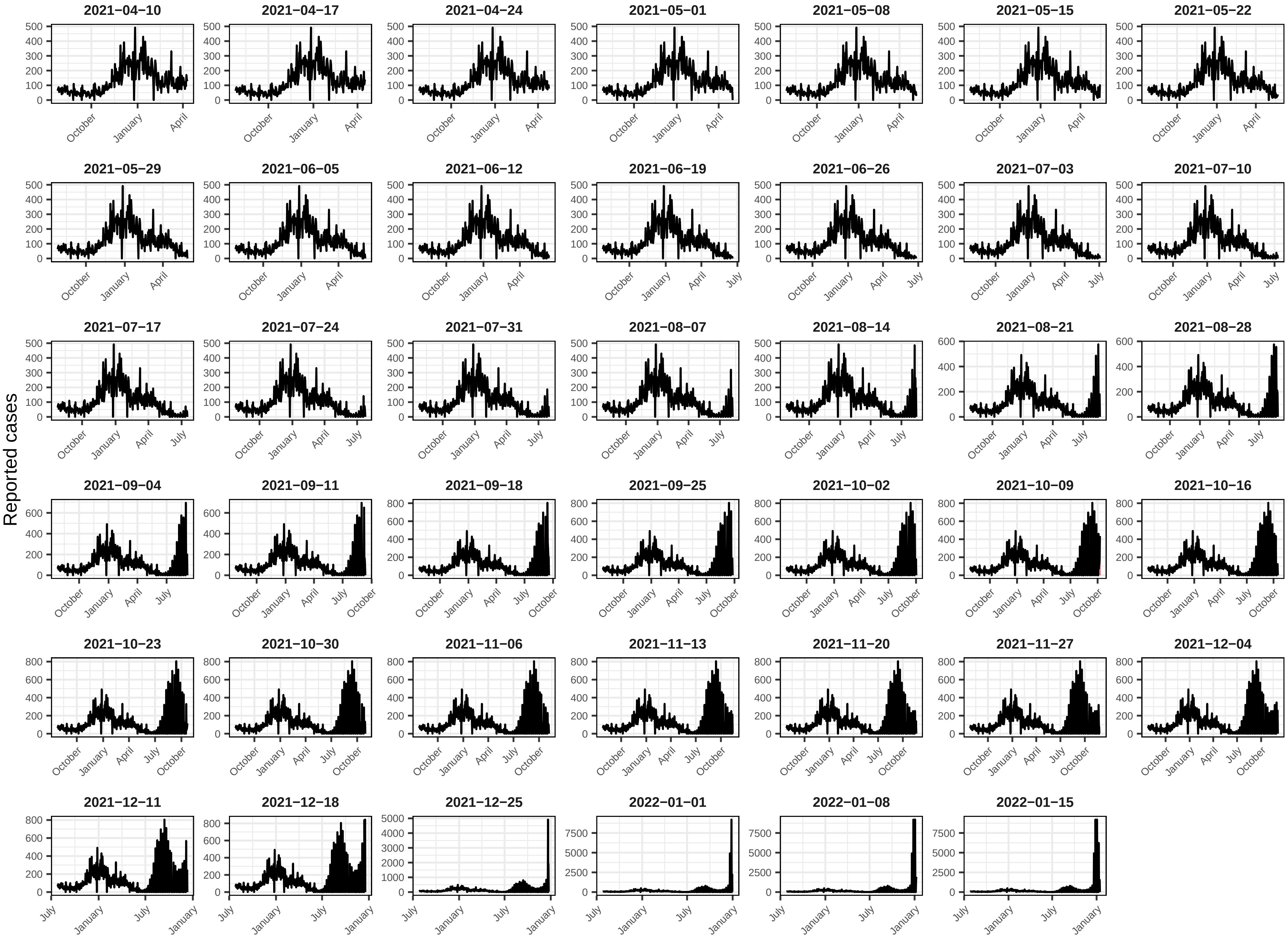

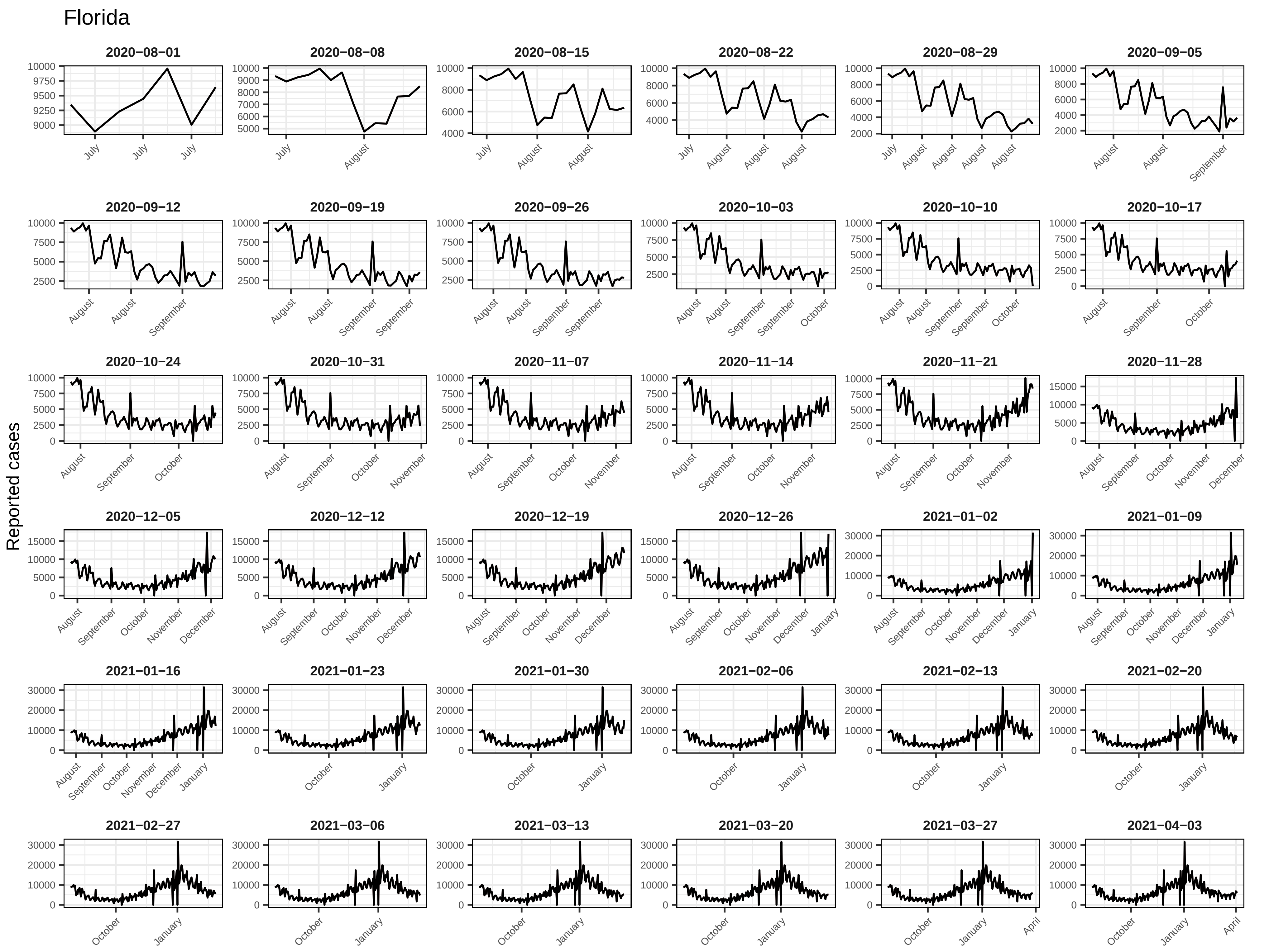
Florida
Reported cases
2020−08−01
2020−08−08
2020−08−15
2020−08−22
2020−08−29
2020−09−05
2020−09−12
2020−09−19
2020−09−26
2020−10−03
2020−10−10
2020−10−17
2020−10−24
2020−10−31
2020−11−07
2020−11−14
2020−11−21
2020−11−28
2020−12−05
2020−12−12
2020−12−19
2020−12−26
2021−01−02
2021−01−09
2021−01−16
2021−01−23
2021−01−30
2021−02−06
2021−02−13
2021−02−20
2021−02−27
2021−03−06
2021−03−13
2021−03−20
2021−03−27
2021−04−03

#### Florida

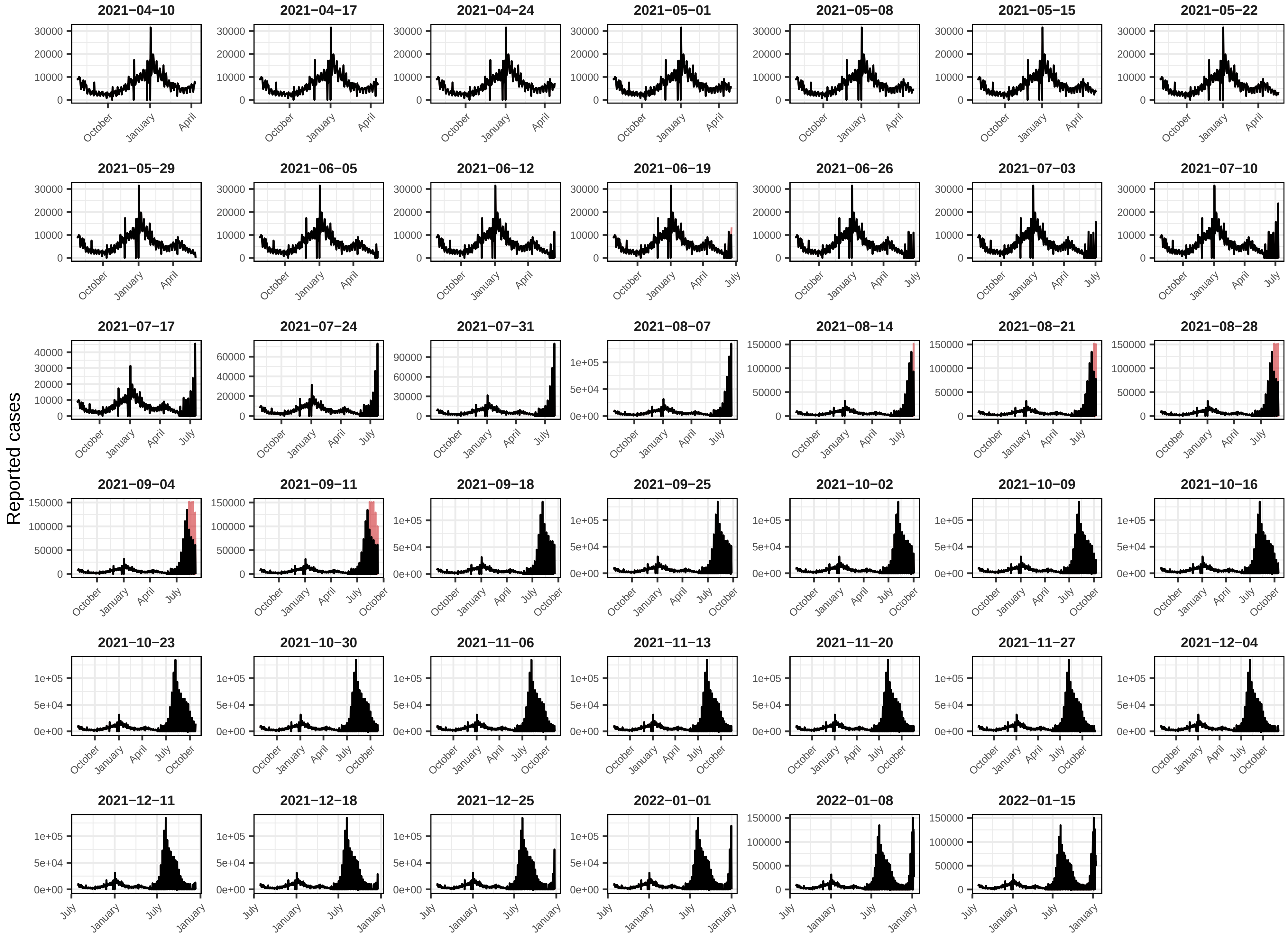

Reported cases
2021−04−10
2021−04−17
2021−04−24
2021−05−01
2021−05−08
2021−05−15
2021−05−22
2021−05−29
2021−06−05
2021−06−12
2021−06−19
2021−06−26
2021−07−03
2021−07−10
2021−07−17
2021−07−24
2021−07−31
2021−08−07
2021−08−14
2021−08−21
2021−08−28
2021−09−04
2021−09−11
2021−09−18
2021−09−25
2021−10−02
2021−10−09
2021−10−16
2021−10−23
2021−10−30
2021−11−06
2021−11−13
2021−11−20
2021−11−27
2021−12−04
2021−12−11
2021−12−18
2021−12−25
2022−01−01
2022−01−08
2022−01−15

#### Georgia

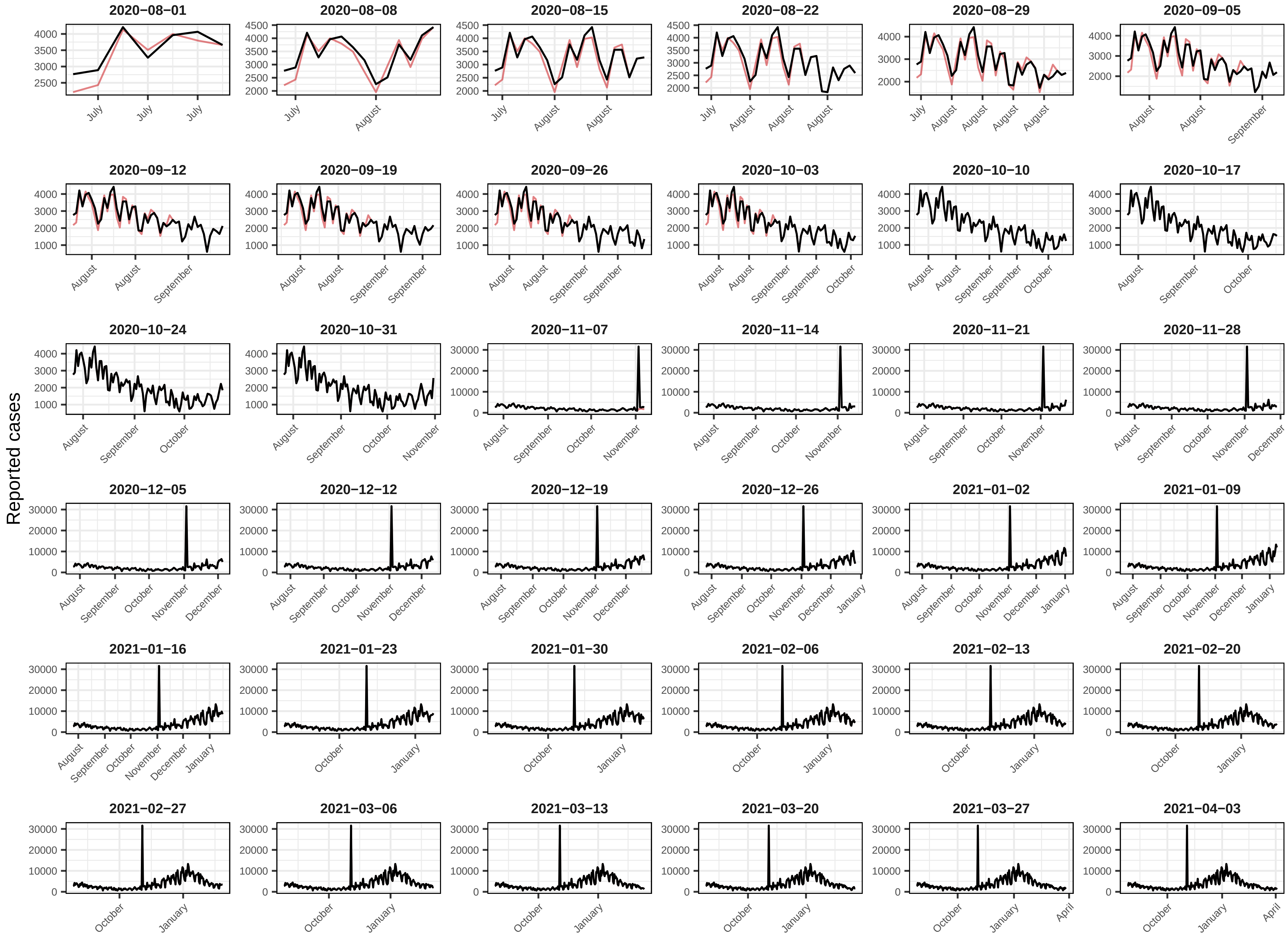
Reported cases
2020−08−01
2020−08−08
2020−08−15
2020−08−22
2020−08−29
2020−09−05
2020−09−12
2020−09−19
2020−09−26
2020−10−03
2020−10−10
2020−10−17
2020−10−24
2020−10−31
2020−11−07
2020−11−14
2020−11−21
2020−11−28
2020−12−05
2020−12−12
2020−12−19
2020−12−26
2021−01−02
2021−01−09
2021−01−16
2021−01−23
2021−01−30
2021−02−06
2021−02−13
2021−02−20
2021−02−27
2021−03−06
2021−03−13
2021−03−20
2021−03−27
2021−04−03

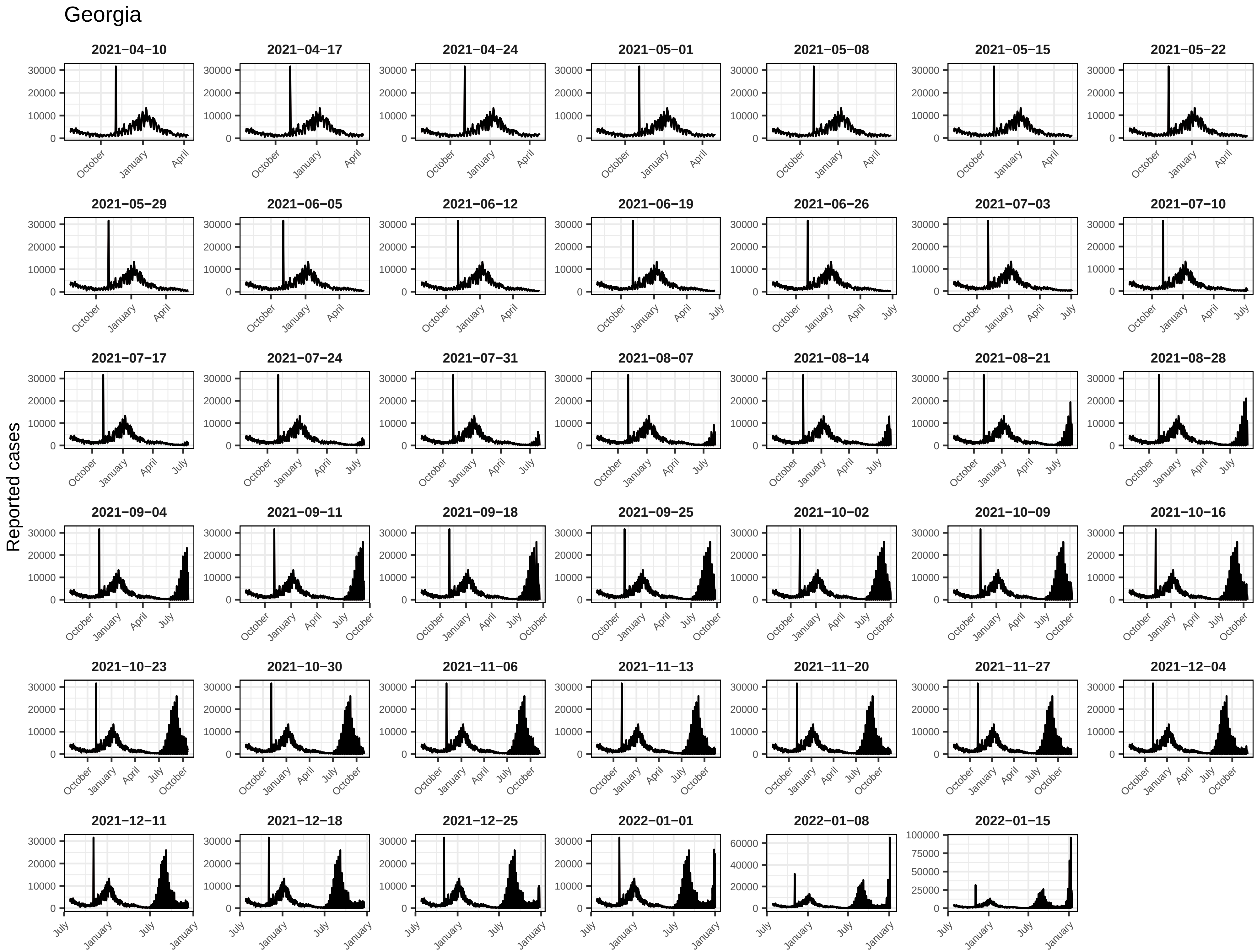
Georgia
Reported cases
2021−04−10
2021−04−17
2021−04−24
2021−05−01
2021−05−08
2021−05−15
2021−05−22
2021−05−29
2021−06−05
2021−06−12
2021−06−19
2021−06−26
2021−07−03
2021−07−10
2021−07−17
2021−07−24
2021−07−31
2021−08−07
2021−08−14
2021−08−21
2021−08−28
2021−09−04
2021−09−11
2021−09−18
2021−09−25
2021−10−02
2021−10−09
2021−10−16
2021−10−23
2021−10−30
2021−11−06
2021−11−13
2021−11−20
2021−11−27
2021−12−04
2021−12−11
2021−12−18
2021−12−25
2022−01−01
2022−01−08
2022−01−15
0
10000
20000
30000
40000
60000
25000
50000
75000
100000
July
October
January
April

Hawaii

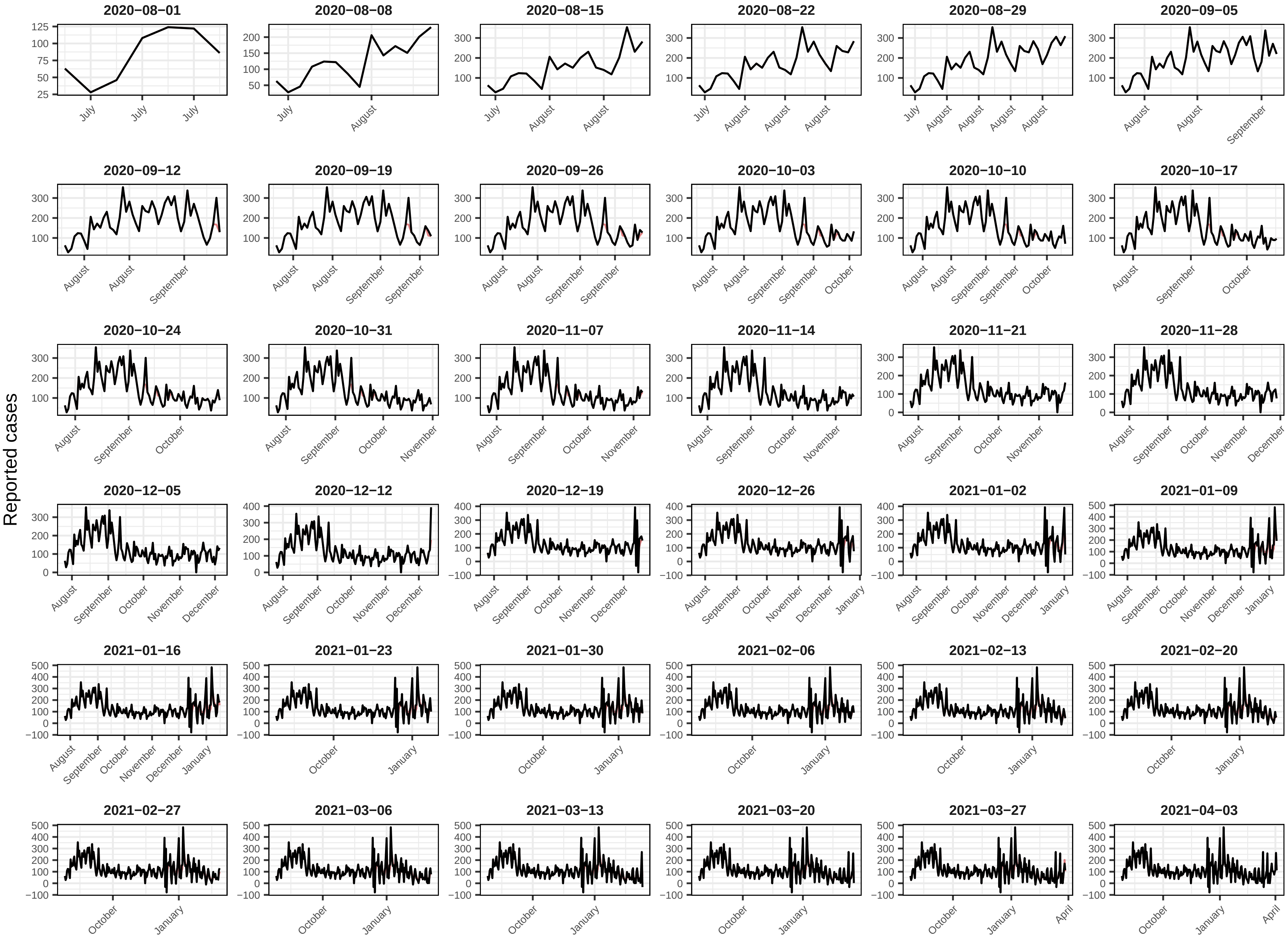
Reported cases
2020−08−01
2020−08−08
2020−08−15
2020−08−22
2020−08−29
2020−09−05
2020−09−12
2020−09−19
2020−09−26
2020−10−03
2020−10−10
2020−10−17
2020−10−24
2020−10−31
2020−11−07
2020−11−14
2020−11−21
2020−11−28
2020−12−05
2020−12−12
2020−12−19
2020−12−26
2021−01−02
2021−01−09
2021−01−16
2021−01−23
2021−01−30
2021−02−06
2021−02−13
2021−02−20
2021−02−27
2021−03−06
2021−03−13
2021−03−20
2021−03−27
2021−04−03

Hawaii

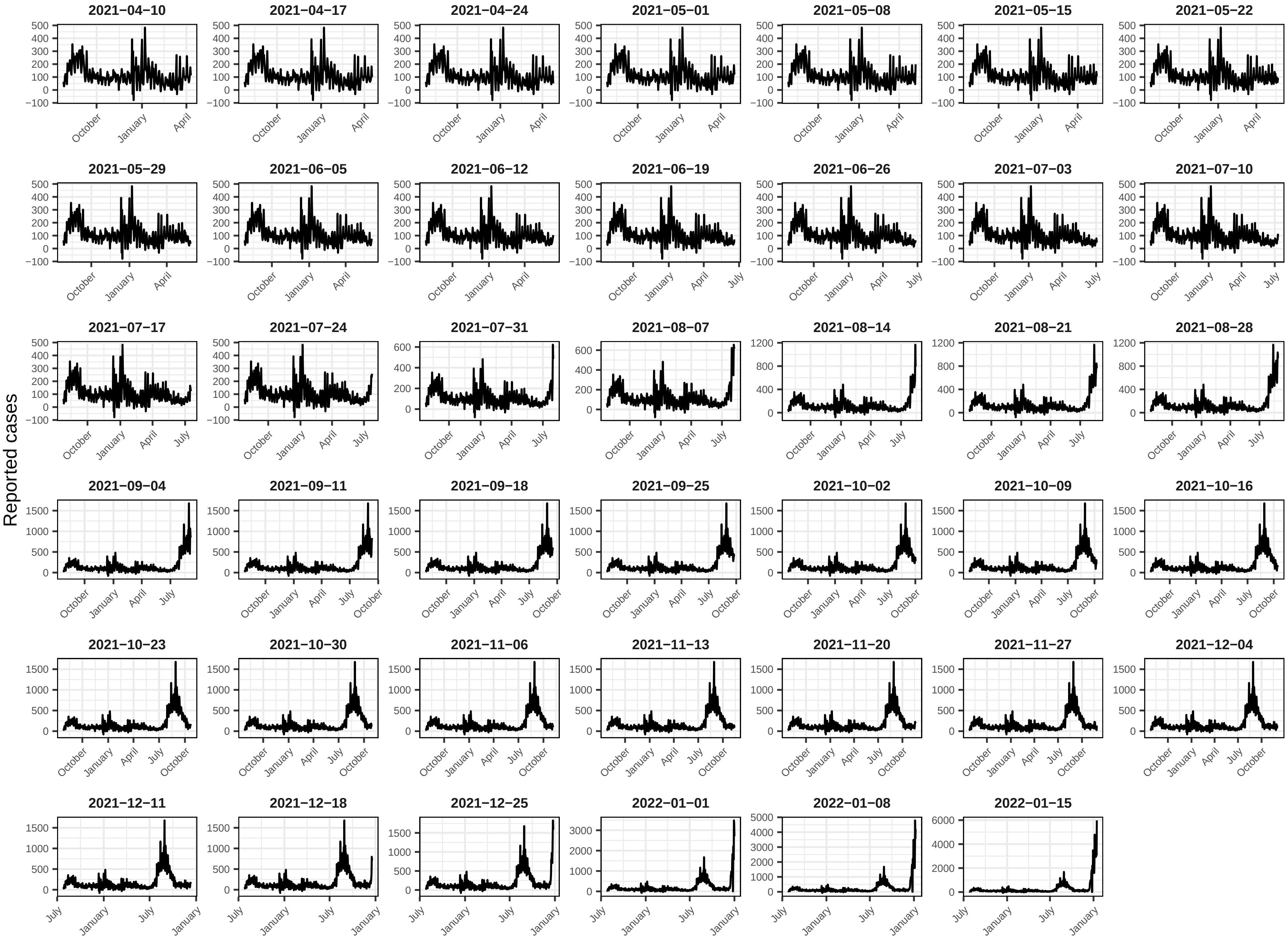
2021−04−10
2021−04−17
2021−04−24
2021−05−01
2021−05−08
2021−05−15
2021−05−22
2021−05−29
2021−06−05
2021−06−12
2021−06−19
2021−06−26
2021−07−03
2021−07−10
2021−07−17
2021−07−24
2021−07−31
2021−08−07
2021−08−14
2021−08−21
2021−08−28
2021−09−04
2021−09−11
2021−09−18
2021−09−25
2021−10−02
2021−10−09
2021−10−16
2021−10−23
2021−10−30
2021−11−06
2021−11−13
2021−11−20
2021−11−27
2021−12−04
2021−12−11
2021−12−18
2021−12−25
2022−01−01
2022−01−08
2022−01−15
Reported cases

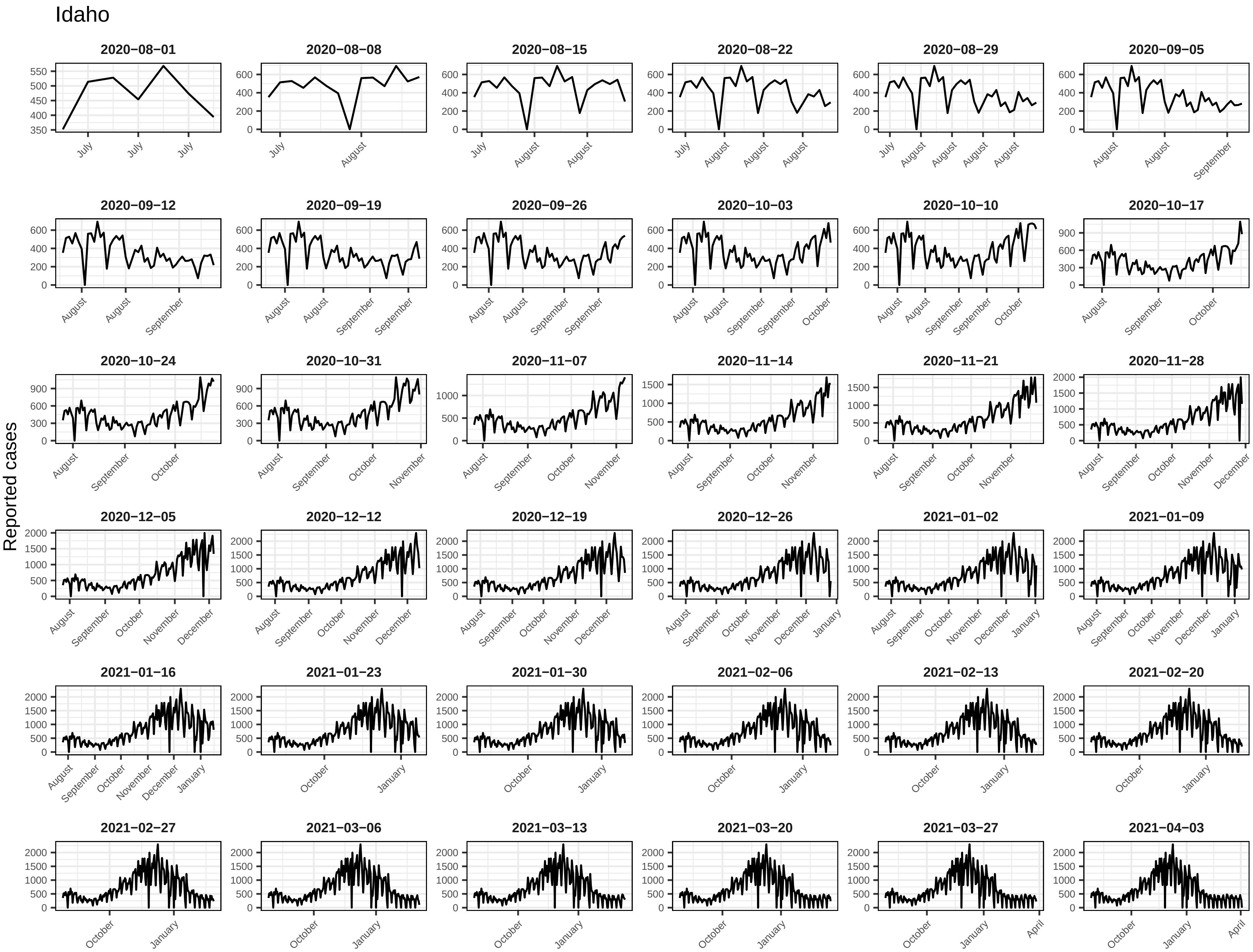
Idaho
Reported cases
2020−08−01
2020−08−08
2020−08−15
2020−08−22
2020−08−29
2020−09−05
2020−09−12
2020−09−19
2020−09−26
2020−10−03
2020−10−10
2020−10−17
2020−10−24
2020−10−31
2020−11−07
2020−11−14
2020−11−21
2020−11−28
2020−12−05
2020−12−12
2020−12−19
2020−12−26
2021−01−02
2021−01−09
2021−01−16
2021−01−23
2021−01−30
2021−02−06
2021−02−13
2021−02−20
2021−02−27
2021−03−06
2021−03−13
2021−03−20
2021−03−27
2021−04−03

Idaho

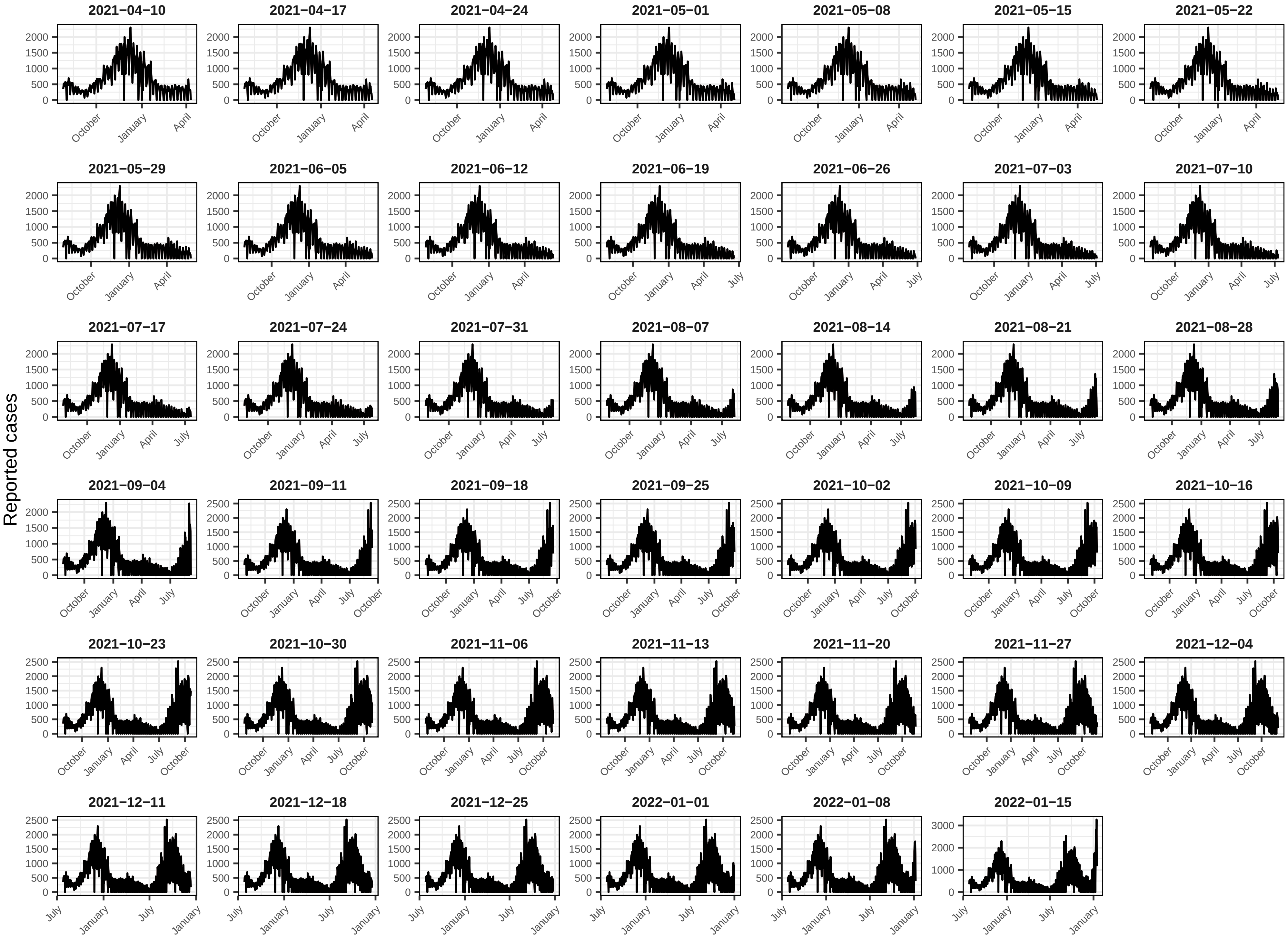
Reported cases
2021−04−10
2021−04−17
2021−04−24
2021−05−01
2021−05−08
2021−05−15
2021−05−22
2021−05−29
2021−06−05
2021−06−12
2021−06−19
2021−06−26
2021−07−03
2021−07−10
2021−07−17
2021−07−24
2021−07−31
2021−08−07
2021−08−14
2021−08−21
2021−08−28
2021−09−04
2021−09−11
2021−09−18
2021−09−25
2021−10−02
2021−10−09
2021−10−16
2021−10−23
2021−10−30
2021−11−06
2021−11−13
2021−11−20
2021−11−27
2021−12−04
2021−12−11
2021−12−18
2021−12−25
2022−01−01
2022−01−08
2022−01−15

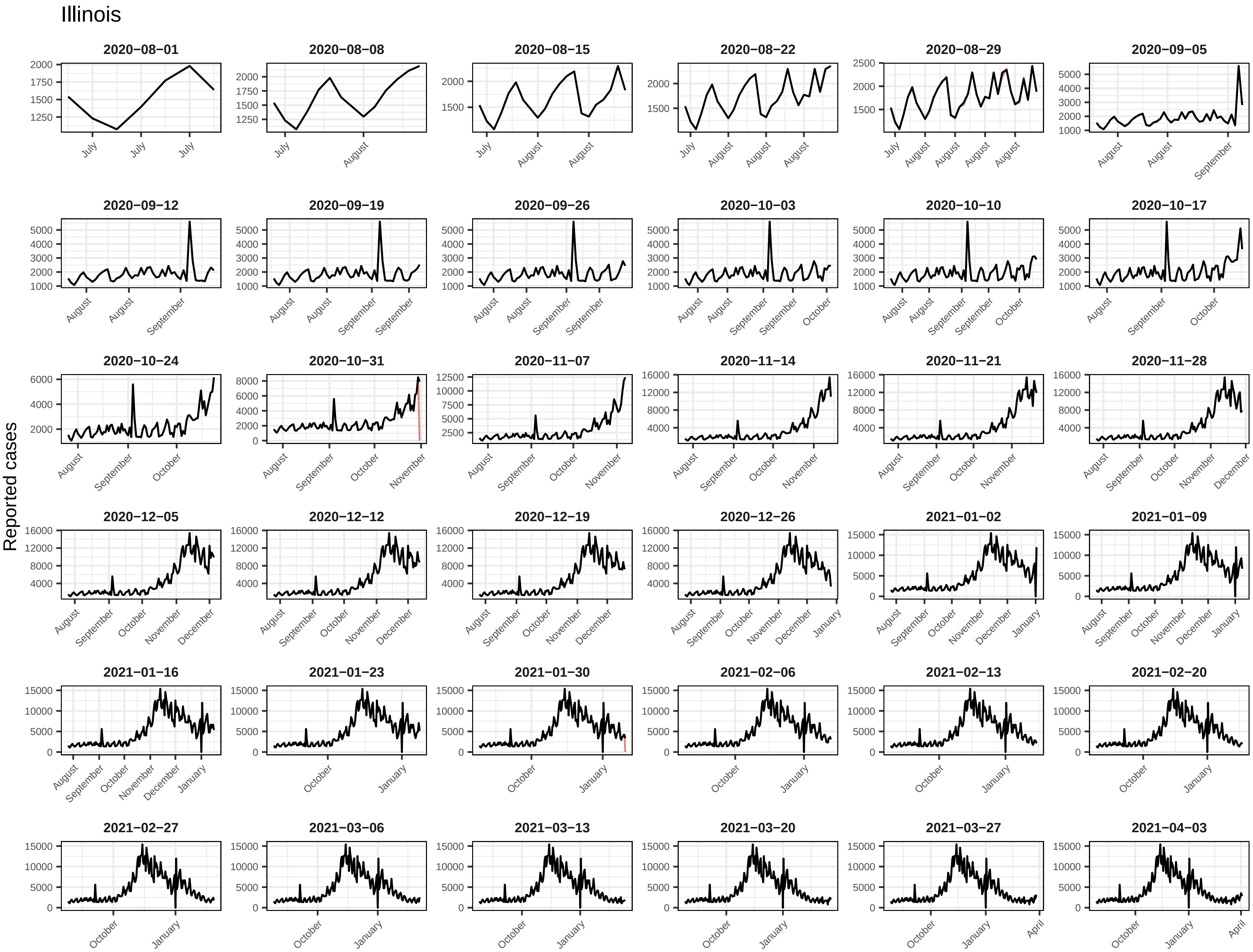
Illinois
Reported cases
2020−08−01
2020−08−08
2020−08−15
2020−08−22
2020−08−29
2020−09−05
2020−09−12
2020−09−19
2020−09−26
2020−10−03
2020−10−10
2020−10−17
2020−10−24
2020−10−31
2020−11−07
2020−11−14
2020−11−21
2020−11−28
2020−12−05
2020−12−12
2020−12−19
2020−12−26
2021−01−02
2021−01−09
2021−01−16
2021−01−23
2021−01−30
2021−02−06
2021−02−13
2021−02−20
2021−02−27
2021−03−06
2021−03−13
2021−03−20
2021−03−27
2021−04−03

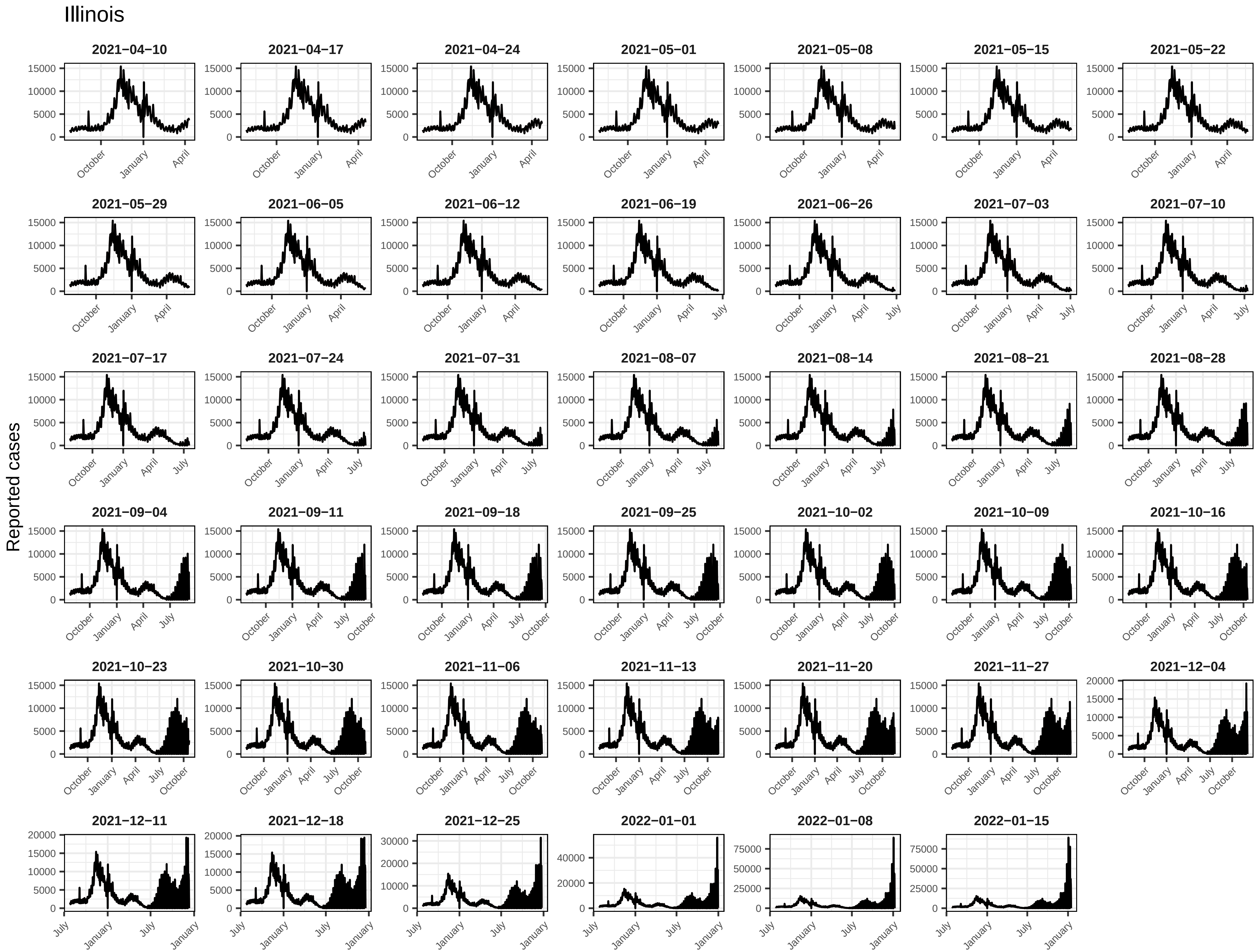
Illinois
Reported cases
2021−04−10
2021−04−17
2021−04−24
2021−05−01
2021−05−08
2021−05−15
2021−05−22
2021−05−29
2021−06−05
2021−06−12
2021−06−19
2021−06−26
2021−07−03
2021−07−10
2021−07−17
2021−07−24
2021−07−31
2021−08−07
2021−08−14
2021−08−21
2021−08−28
2021−09−04
2021−09−11
2021−09−18
2021−09−25
2021−10−02
2021−10−09
2021−10−16
2021−10−23
2021−10−30
2021−11−06
2021−11−13
2021−11−20
2021−11−27
2021−12−04
2021−12−11
2021−12−18
2021−12−25
2022−01−01
2022−01−08
2022−01−15

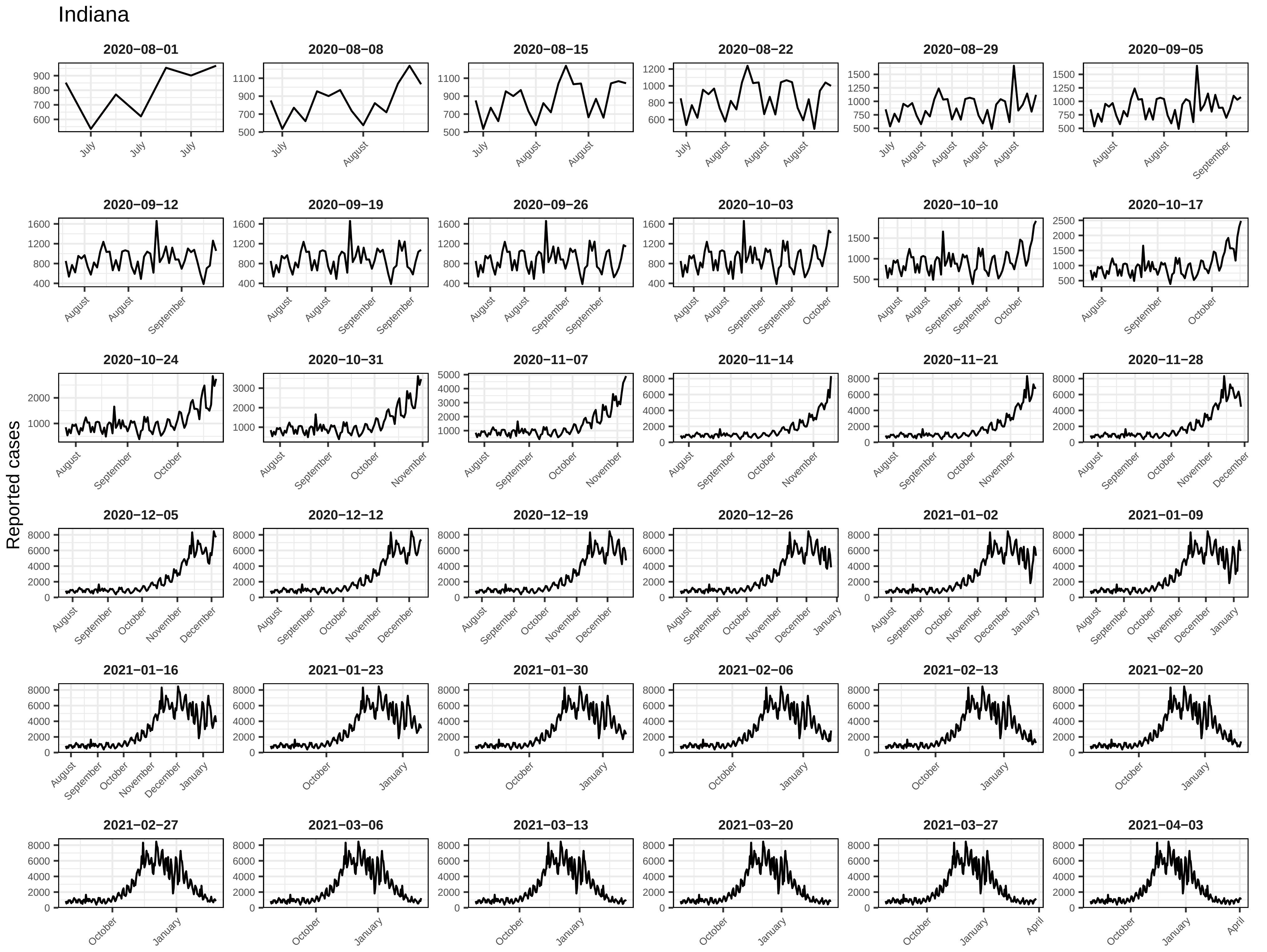
Indiana
Reported cases
2020−08−01
2020−08−08
2020−08−15
2020−08−22
2020−08−29
2020−09−05
2020−09−12
2020−09−19
2020−09−26
2020−10−03
2020−10−10
2020−10−17
2020−10−24
2020−10−31
2020−11−07
2020−11−14
2020−11−21
2020−11−28
2020−12−05
2020−12−12
2020−12−19
2020−12−26
2021−01−02
2021−01−09
2021−01−16
2021−01−23
2021−01−30
2021−02−06
2021−02−13
2021−02−20
2021−02−27
2021−03−06
2021−03−13
2021−03−20
2021−03−27
2021−04−03

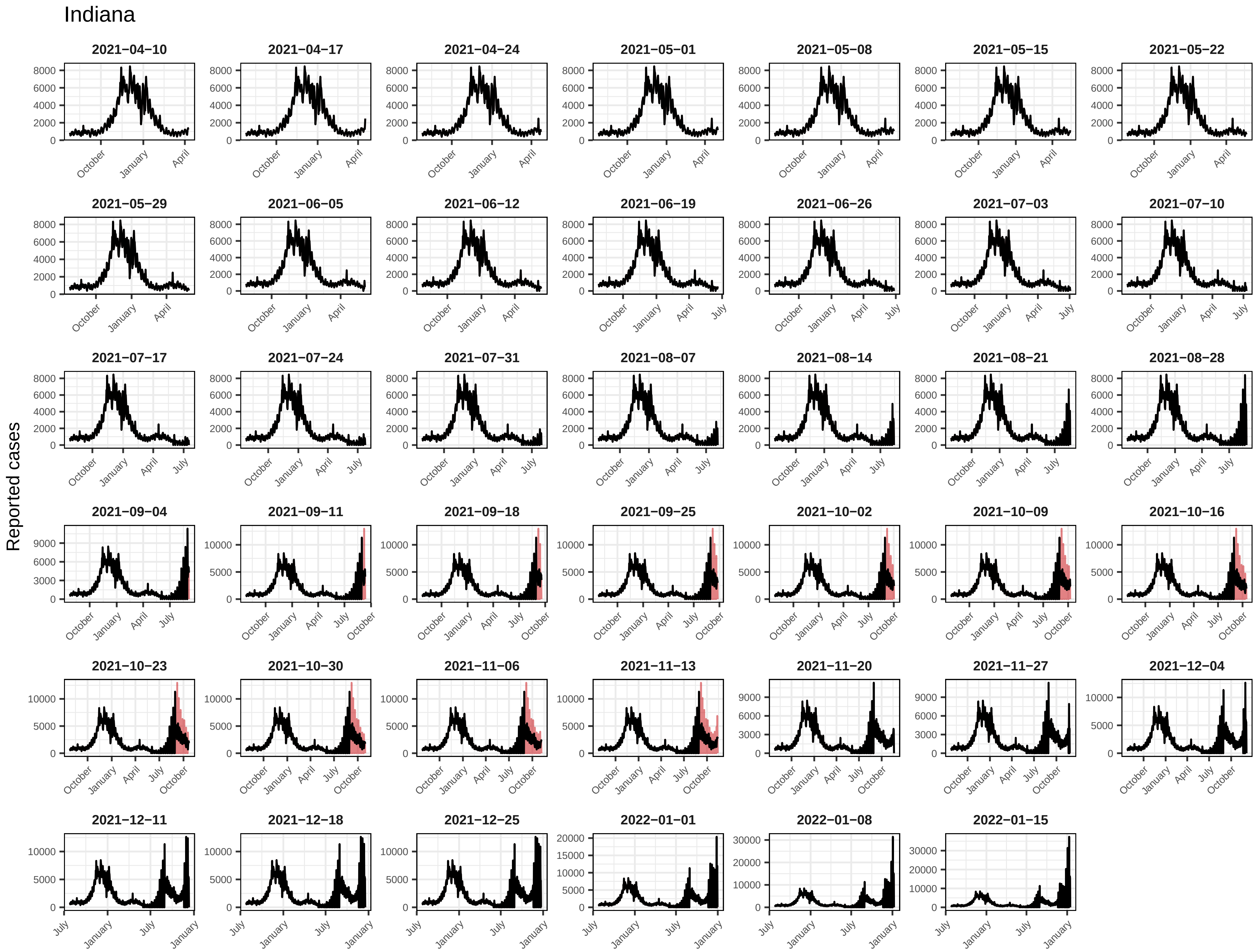
Indiana
Reported cases
2021−04−10
2021−04−17
2021−04−24
2021−05−01
2021−05−08
2021−05−15
2021−05−22
2021−05−29
2021−06−05
2021−06−12
2021−06−19
2021−06−26
2021−07−03
2021−07−10
2021−07−17
2021−07−24
2021−07−31
2021−08−07
2021−08−14
2021−08−21
2021−08−28
2021−09−04
2021−09−11
2021−09−18
2021−09−25
2021−10−02
2021−10−09
2021−10−16
2021−10−23
2021−10−30
2021−11−06
2021−11−13
2021−11−20
2021−11−27
2021−12−04
2021−12−11
2021−12−18
2021−12−25
2022−01−01
2022−01−08
2022−01−15
October
January
April
July

### Iowa

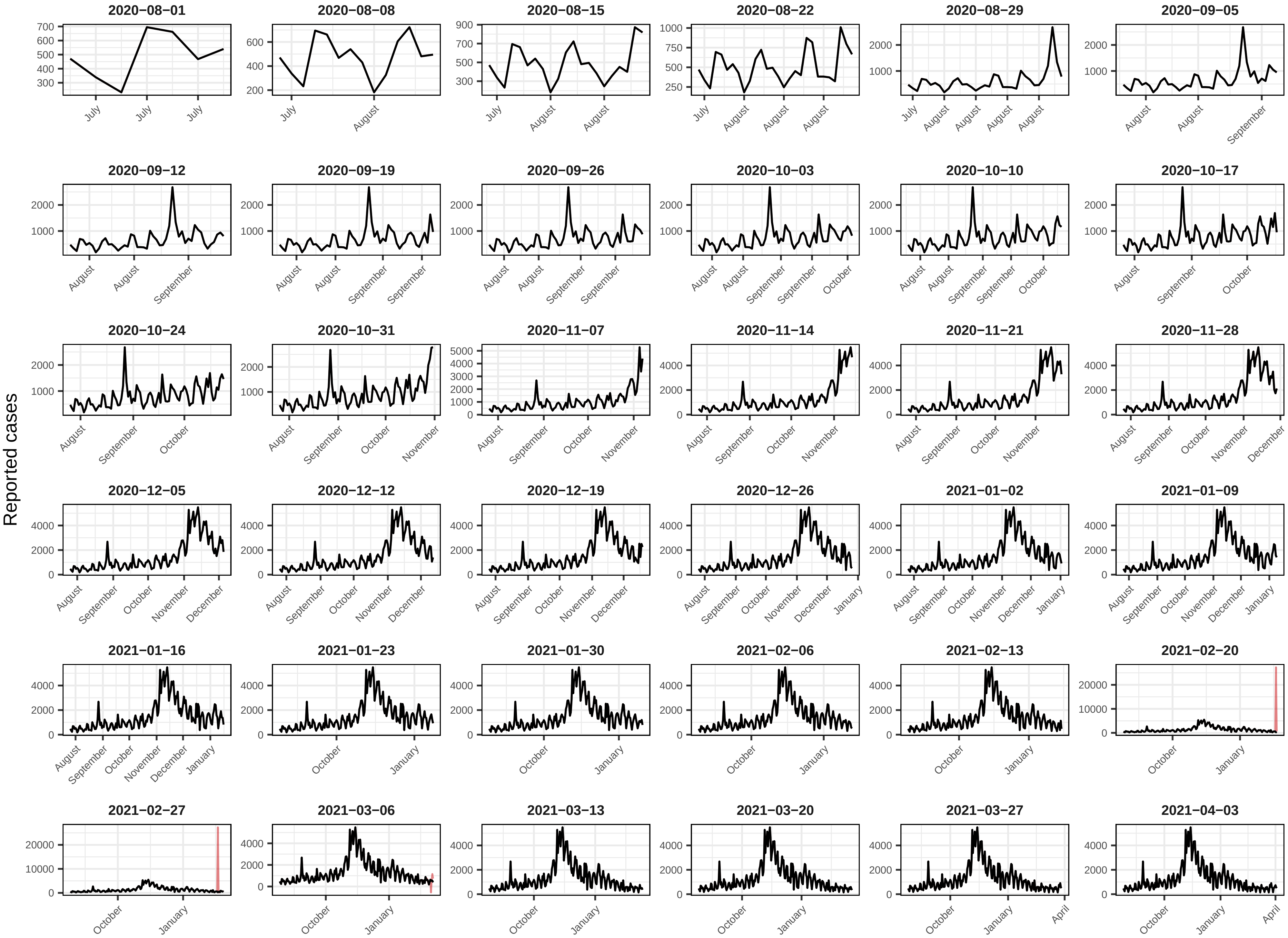

Reported cases
2020−08−01
2020−08−08
2020−08−15
2020−08−22
2020−08−29
2020−09−05
2020−09−12
2020−09−19
2020−09−26
2020−10−03
2020−10−10
2020−10−17
2020−10−24
2020−10−31
2020−11−07
2020−11−14
2020−11−21
2020−11−28
2020−12−05
2020−12−12
2020−12−19
2020−12−26
2021−01−02
2021−01−09
2021−01−16
2021−01−23
2021−01−30
2021−02−06
2021−02−13
2021−02−20
2021−02−27
2021−03−06
2021−03−13
2021−03−20
2021−03−27
2021−04−03

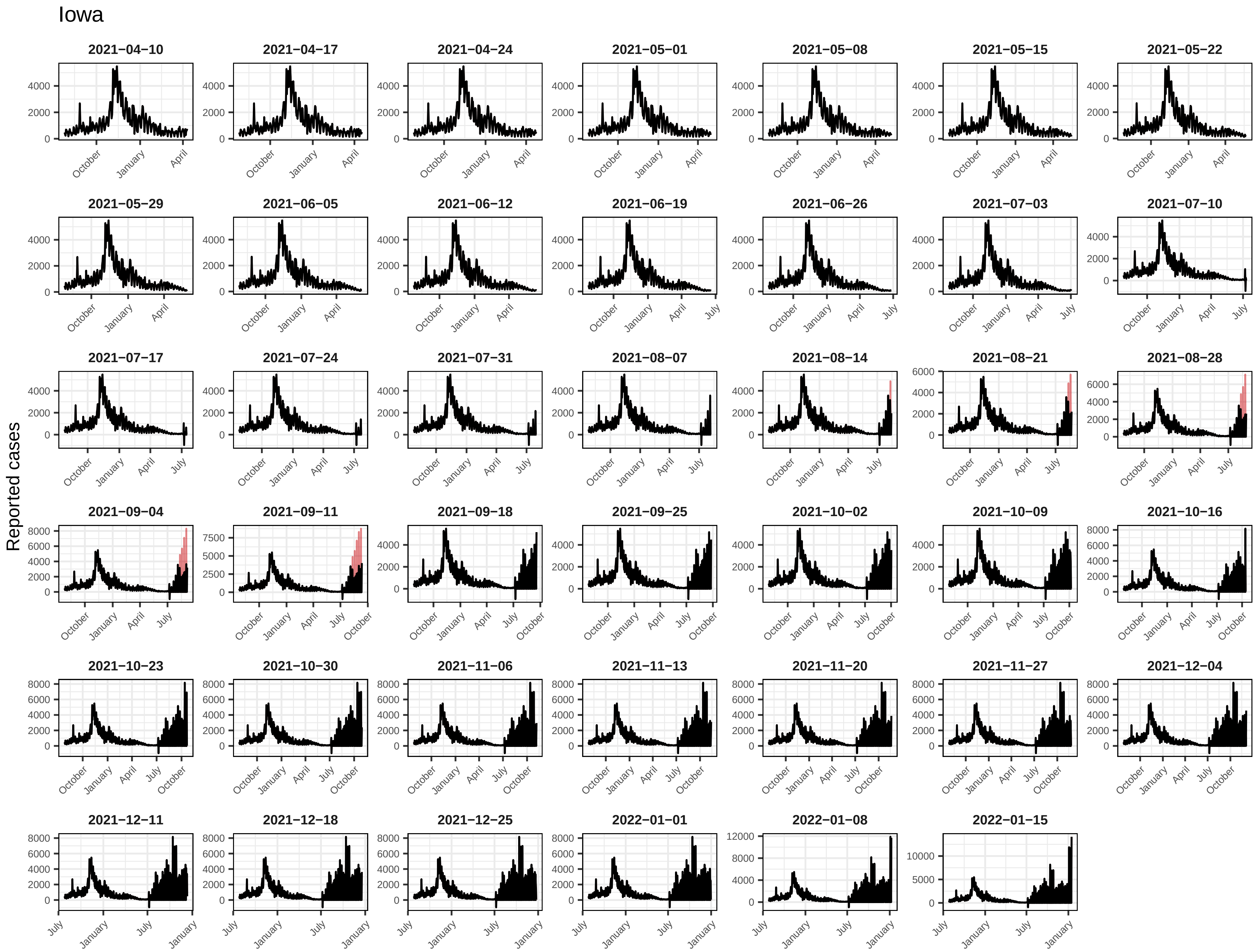
Iowa
Reported cases
2021−04−10
2021−04−17
2021−04−24
2021−05−01
2021−05−08
2021−05−15
2021−05−22
2021−05−29
2021−06−05
2021−06−12
2021−06−19
2021−06−26
2021−07−03
2021−07−10
2021−07−17
2021−07−24
2021−07−31
2021−08−07
2021−08−14
2021−08−21
2021−08−28
2021−09−04
2021−09−11
2021−09−18
2021−09−25
2021−10−02
2021−10−09
2021−10−16
2021−10−23
2021−10−30
2021−11−06
2021−11−13
2021−11−20
2021−11−27
2021−12−04
2021−12−11
2021−12−18
2021−12−25
2022−01−01
2022−01−08
2022−01−15

### Kansas

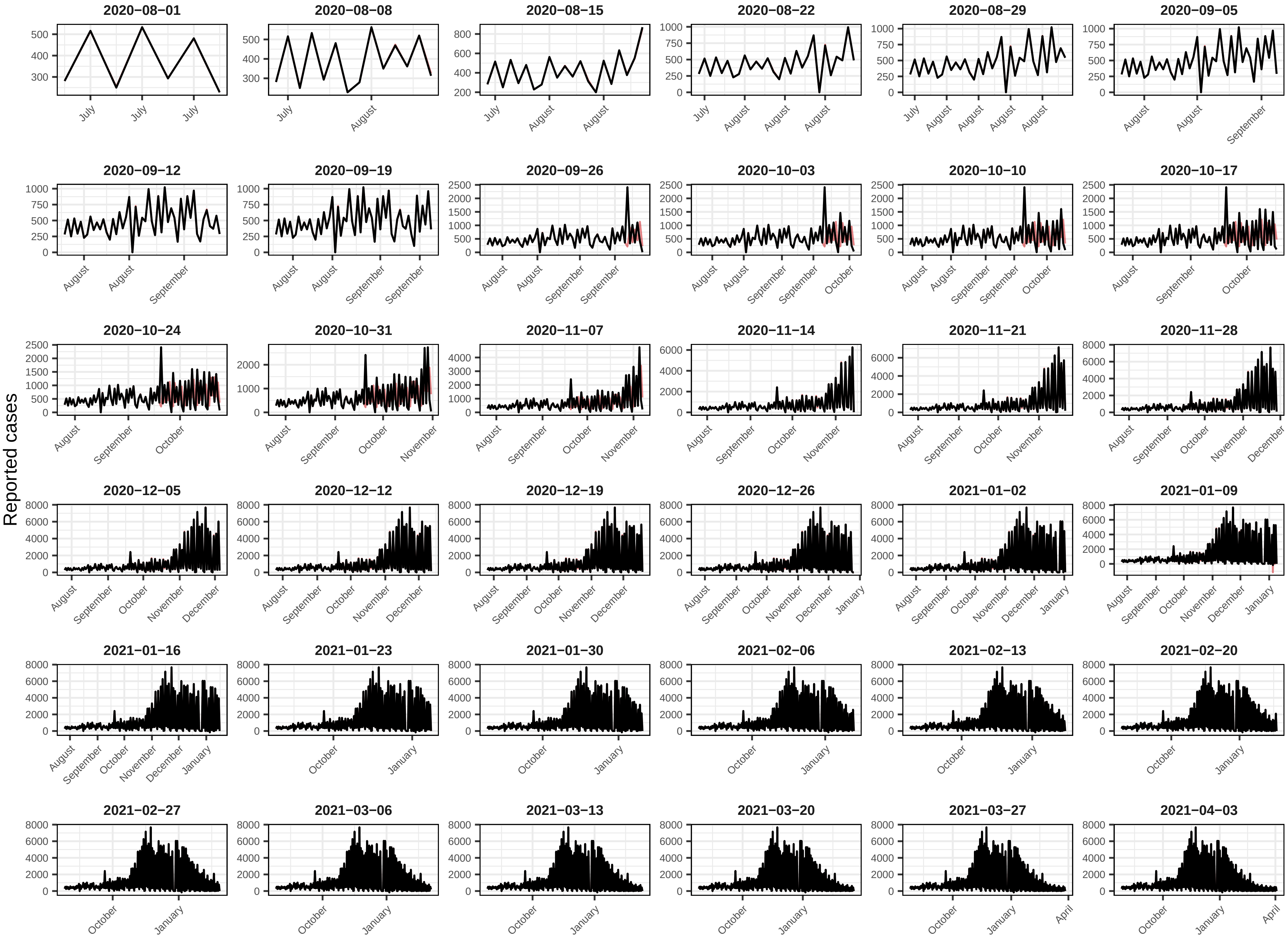

2020−08−01
2020−08−08
2020−08−15
2020−08−22
2020−08−29
2020−09−05
2020−09−12
2020−09−19
2020−09−26
2020−10−03
2020−10−10
2020−10−17
2020−10−24
2020−10−31
2020−11−07
2020−11−14
2020−11−21
2020−11−28
2020−12−05
2020−12−12
2020−12−19
2020−12−26
2021−01−02
2021−01−09
2021−01−16
2021−01−23
2021−01−30
2021−02−06
2021−02−13
2021−02−20
2021−02−27
2021−03−06
2021−03−13
2021−03−20
2021−03−27
2021−04−03
Reported cases

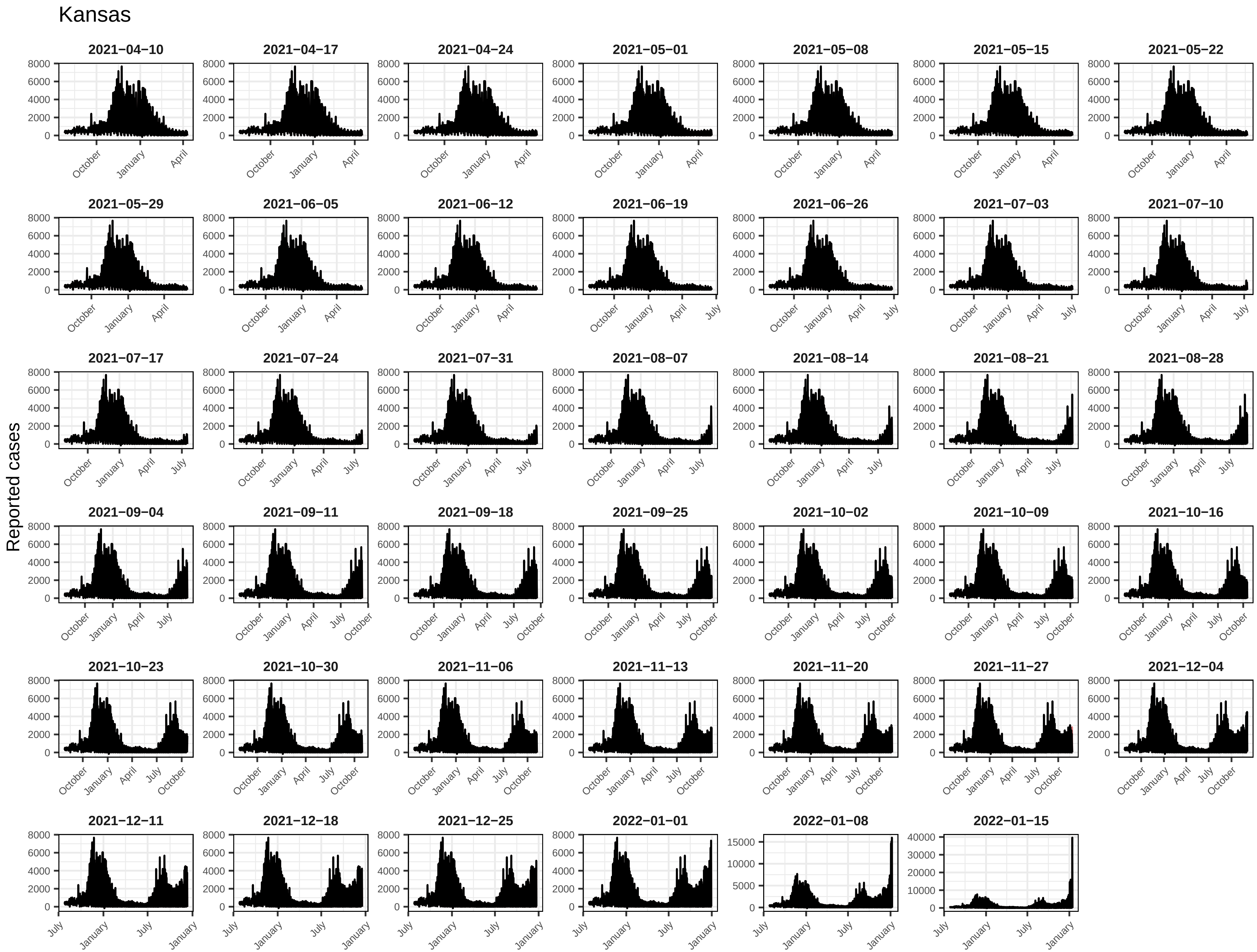
Kansas
Reported cases
2021−04−10
2021−04−17
2021−04−24
2021−05−01
2021−05−08
2021−05−15
2021−05−22
2021−05−29
2021−06−05
2021−06−12
2021−06−19
2021−06−26
2021−07−03
2021−07−10
2021−07−17
2021−07−24
2021−07−31
2021−08−07
2021−08−14
2021−08−21
2021−08−28
2021−09−04
2021−09−11
2021−09−18
2021−09−25
2021−10−02
2021−10−09
2021−10−16
2021−10−23
2021−10−30
2021−11−06
2021−11−13
2021−11−20
2021−11−27
2021−12−04
2021−12−11
2021−12−18
2021−12−25
2022−01−01
2022−01−08
2022−01−15

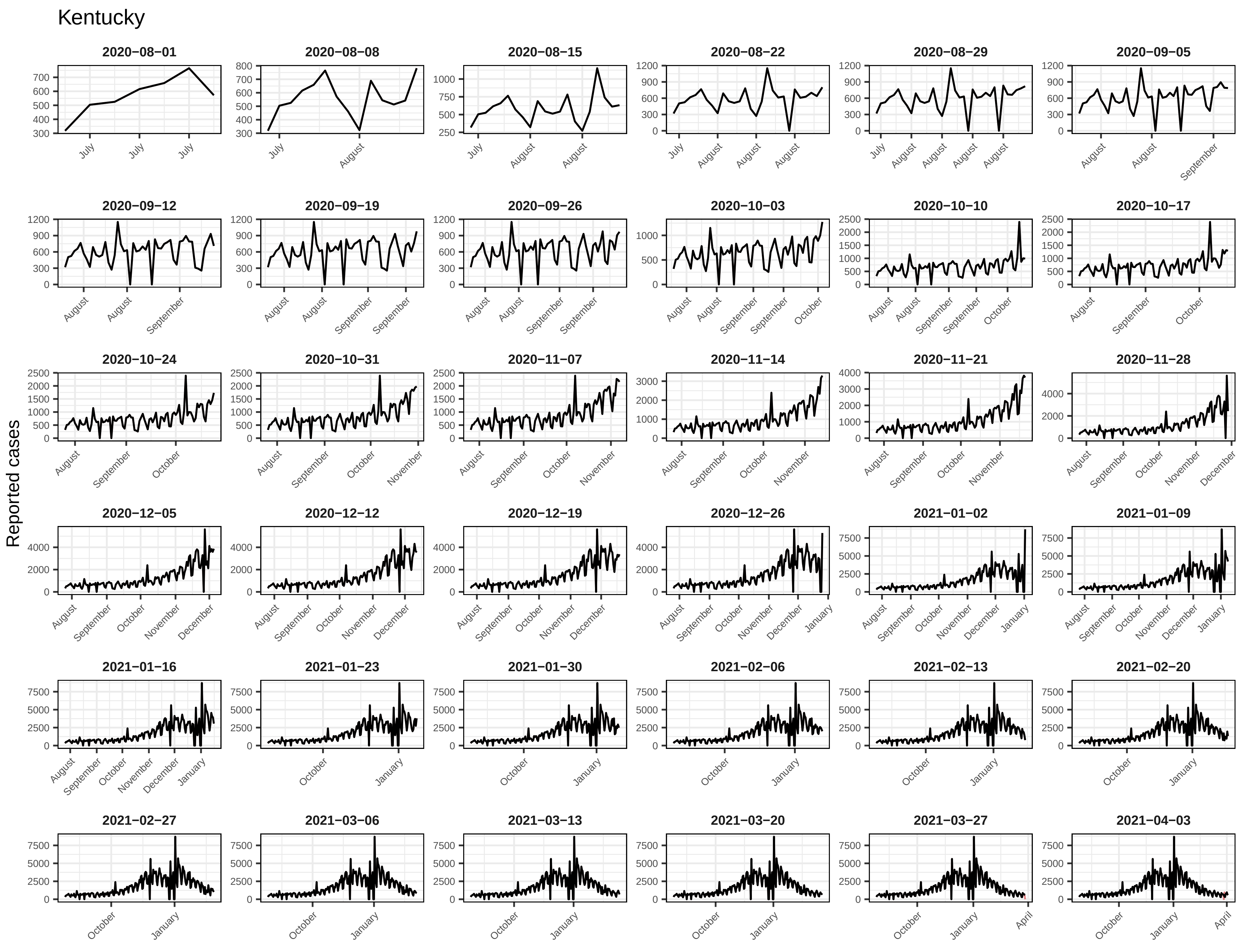
Kentucky
Reported cases
2020−08−01
2020−08−08
2020−08−15
2020−08−22
2020−08−29
2020−09−05
2020−09−12
2020−09−19
2020−09−26
2020−10−03
2020−10−10
2020−10−17
2020−10−24
2020−10−31
2020−11−07
2020−11−14
2020−11−21
2020−11−28
2020−12−05
2020−12−12
2020−12−19
2020−12−26
2021−01−02
2021−01−09
2021−01−16
2021−01−23
2021−01−30
2021−02−06
2021−02−13
2021−02−20
2021−02−27
2021−03−06
2021−03−13
2021−03−20
2021−03−27
2021−04−03

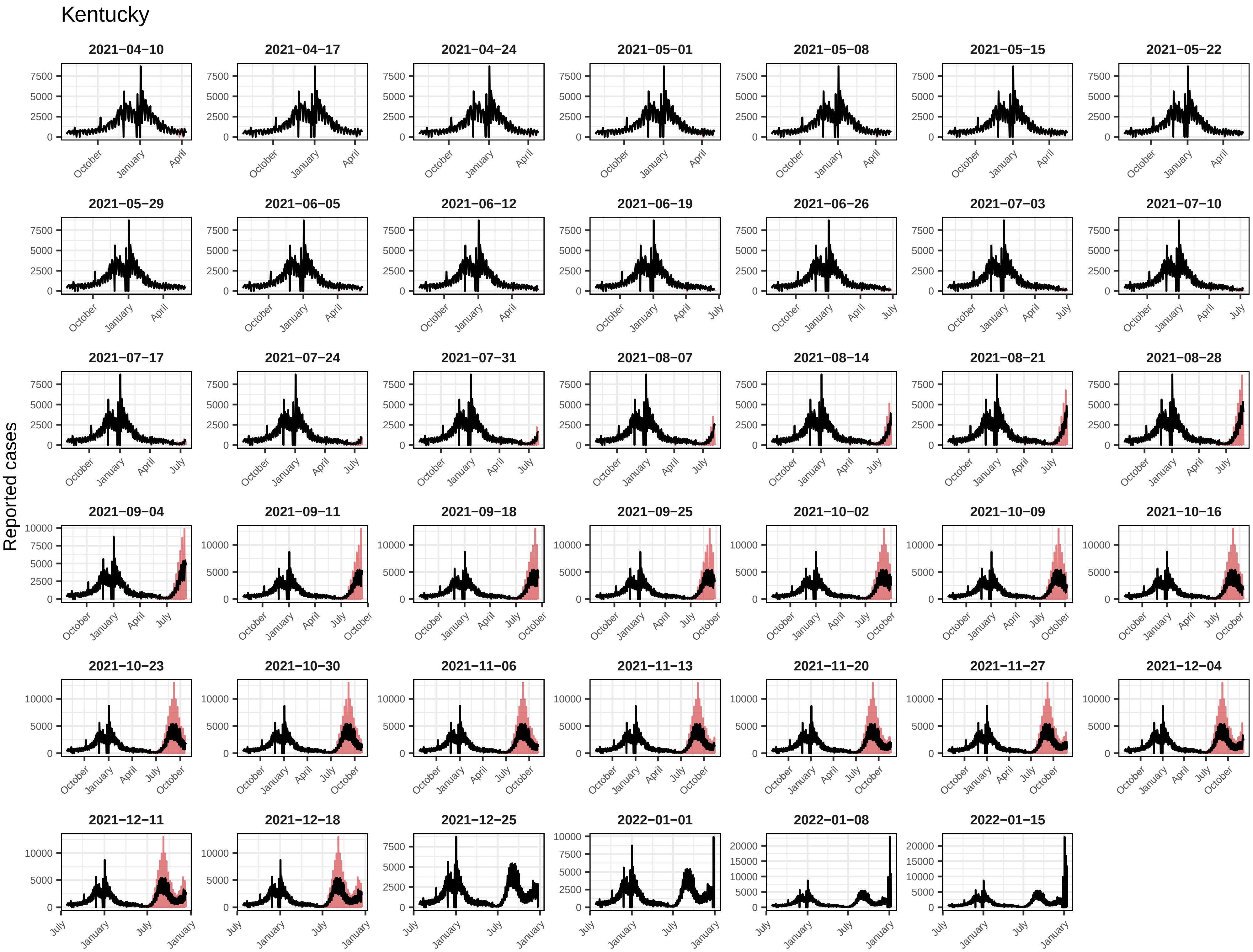
Kentucky
Reported cases
2021−04−10
2021−04−17
2021−04−24
2021−05−01
2021−05−08
2021−05−15
2021−05−22
2021−05−29
2021−06−05
2021−06−12
2021−06−19
2021−06−26
2021−07−03
2021−07−10
2021−07−17
2021−07−24
2021−07−31
2021−08−07
2021−08−14
2021−08−21
2021−08−28
2021−09−04
2021−09−11
2021−09−18
2021−09−25
2021−10−02
2021−10−09
2021−10−16
2021−10−23
2021−10−30
2021−11−06
2021−11−13
2021−11−20
2021−11−27
2021−12−04
2021−12−11
2021−12−18
2021−12−25
2022−01−01
2022−01−08
2022−01−15
0
2500
5000
7500
10000
15000
20000
October
January
April
July

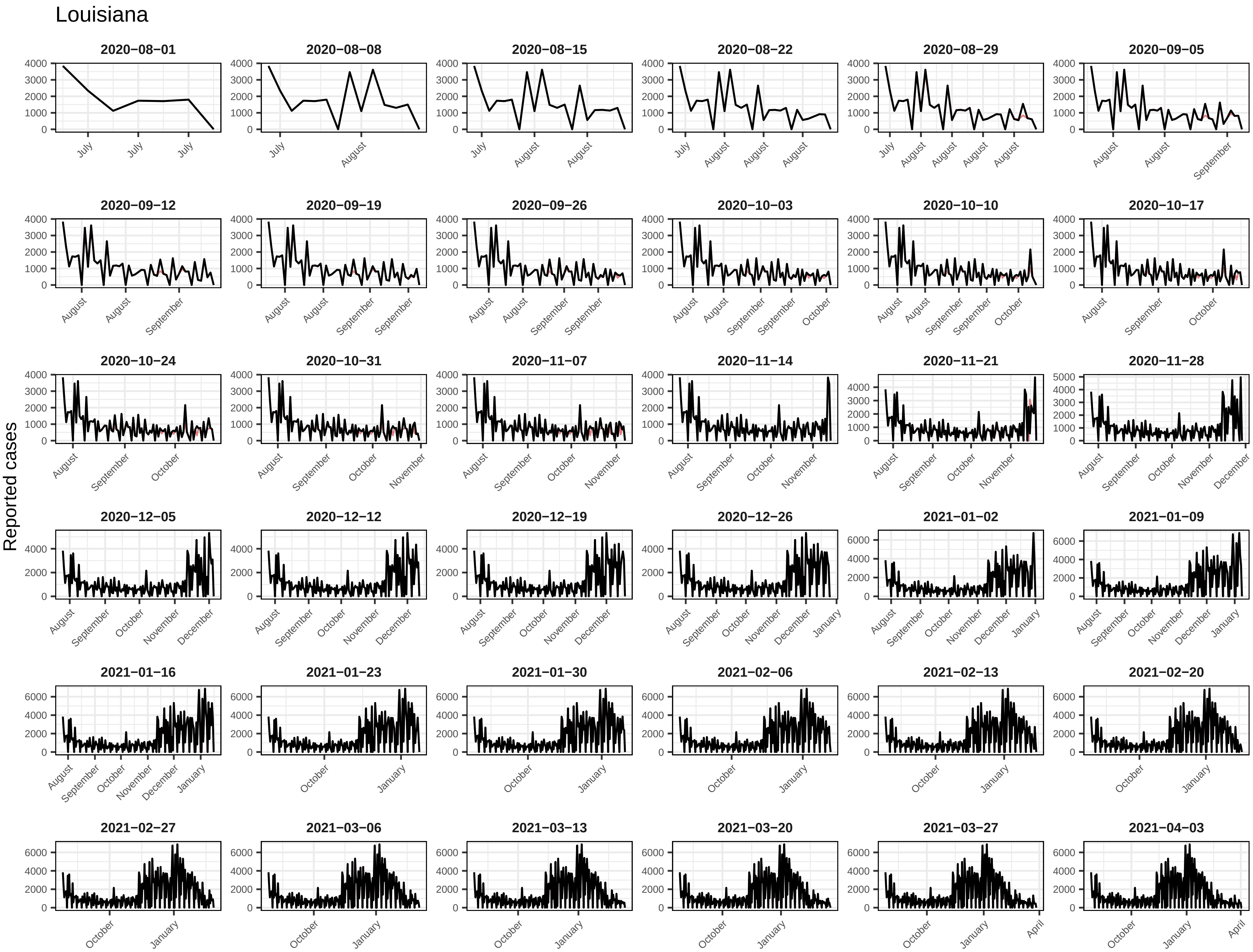
Louisiana
Reported cases
2020−08−01
2020−08−08
2020−08−15
2020−08−22
2020−08−29
2020−09−05
2020−09−12
2020−09−19
2020−09−26
2020−10−03
2020−10−10
2020−10−17
2020−10−24
2020−10−31
2020−11−07
2020−11−14
2020−11−21
2020−11−28
2020−12−05
2020−12−12
2020−12−19
2020−12−26
2021−01−02
2021−01−09
2021−01−16
2021−01−23
2021−01−30
2021−02−06
2021−02−13
2021−02−20
2021−02−27
2021−03−06
2021−03−13
2021−03−20
2021−03−27
2021−04−03

Louisiana

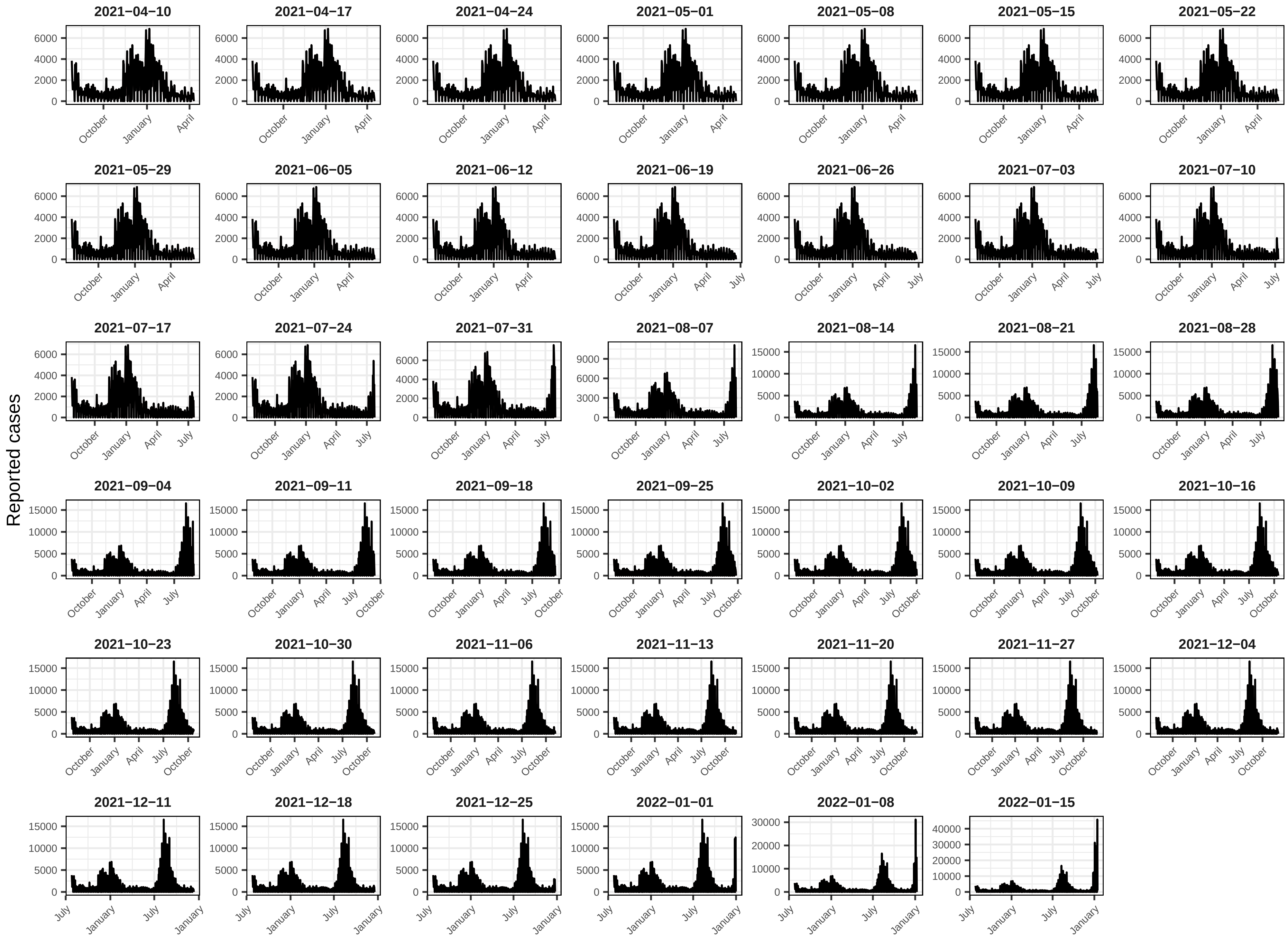
Reported cases
2021−04−10
2021−04−17
2021−04−24
2021−05−01
2021−05−08
2021−05−15
2021−05−22
2021−05−29
2021−06−05
2021−06−12
2021−06−19
2021−06−26
2021−07−03
2021−07−10
2021−07−17
2021−07−24
2021−07−31
2021−08−07
2021−08−14
2021−08−21
2021−08−28
2021−09−04
2021−09−11
2021−09−18
2021−09−25
2021−10−02
2021−10−09
2021−10−16
2021−10−23
2021−10−30
2021−11−06
2021−11−13
2021−11−20
2021−11−27
2021−12−04
2021−12−11
2021−12−18
2021−12−25
2022−01−01
2022−01−08
2022−01−15

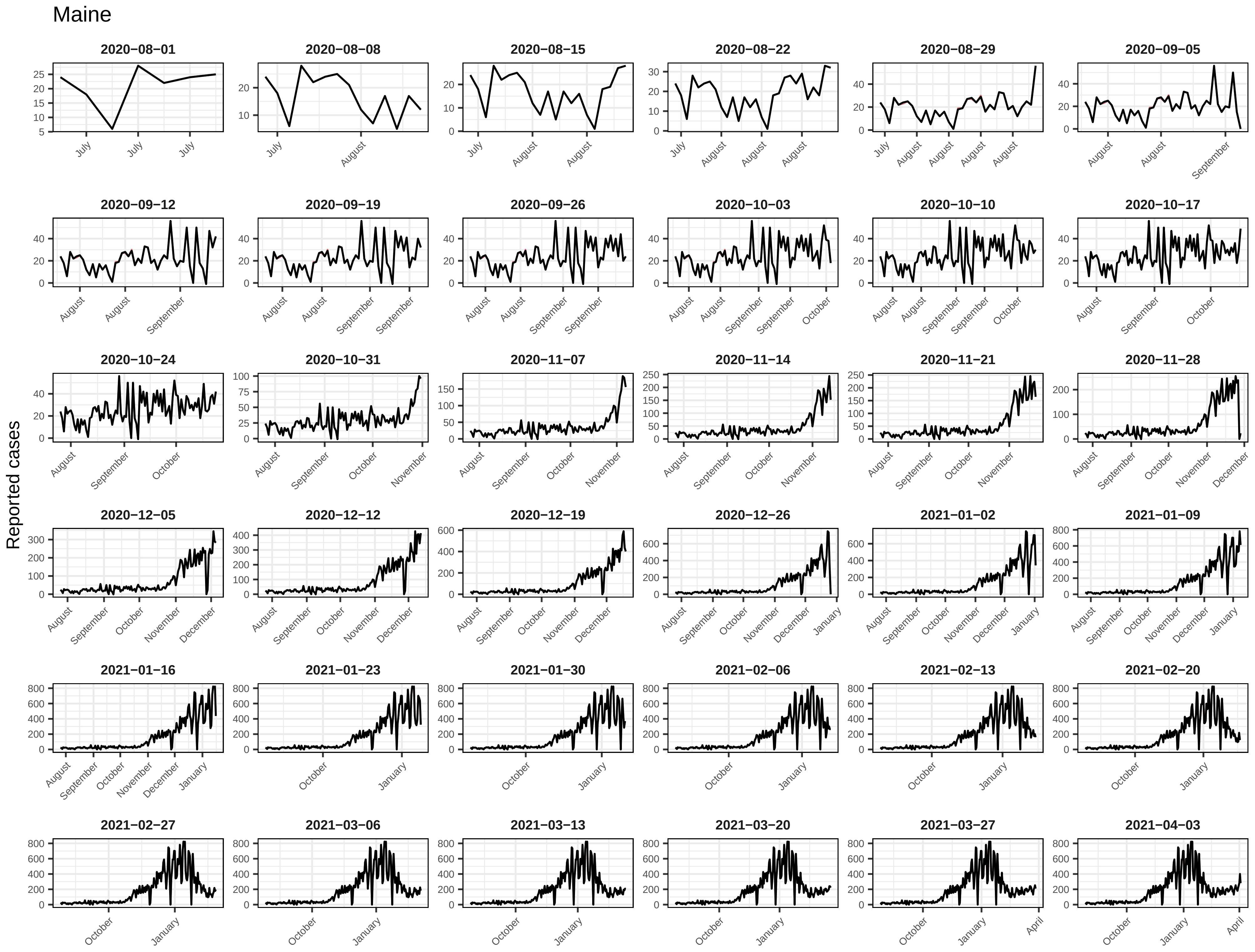

Maine
Reported cases
2020−08−01
2020−08−08
2020−08−15
2020−08−22
2020−08−29
2020−09−05
2020−09−12
2020−09−19
2020−09−26
2020−10−03
2020−10−10
2020−10−17
2020−10−24
2020−10−31
2020−11−07
2020−11−14
2020−11−21
2020−11−28
2020−12−05
2020−12−12
2020−12−19
2020−12−26
2021−01−02
2021−01−09
2021−01−16
2021−01−23
2021−01−30
2021−02−06
2021−02−13
2021−02−20
2021−02−27
2021−03−06
2021−03−13
2021−03−20
2021−03−27
2021−04−03

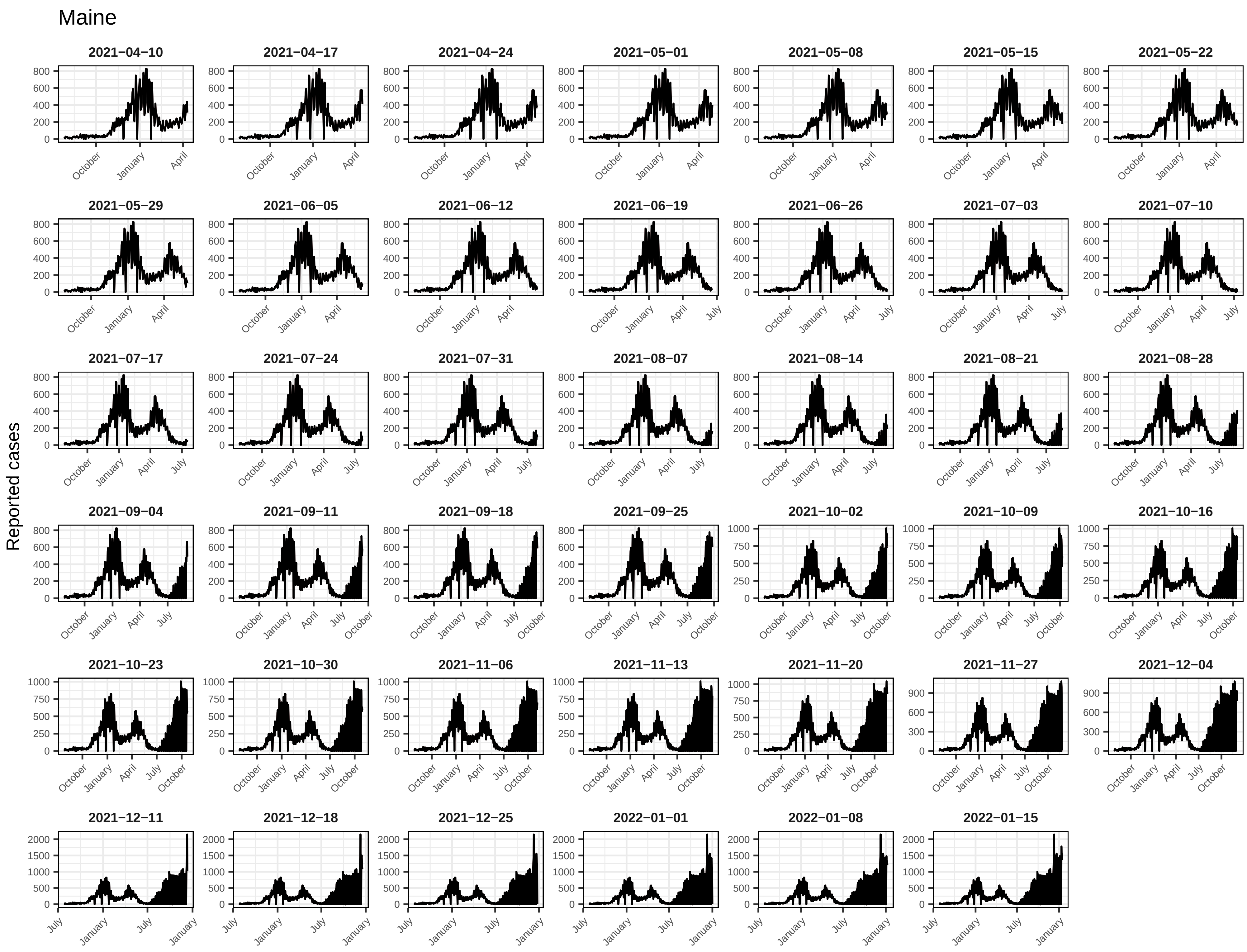
Maine
Reported cases
2021−04−10
2021−04−17
2021−04−24
2021−05−01
2021−05−08
2021−05−15
2021−05−22
2021−05−29
2021−06−05
2021−06−12
2021−06−19
2021−06−26
2021−07−03
2021−07−10
2021−07−17
2021−07−24
2021−07−31
2021−08−07
2021−08−14
2021−08−21
2021−08−28
2021−09−04
2021−09−11
2021−09−18
2021−09−25
2021−10−02
2021−10−09
2021−10−16
2021−10−23
2021−10−30
2021−11−06
2021−11−13
2021−11−20
2021−11−27
2021−12−04
2021−12−11
2021−12−18
2021−12−25
2022−01−01
2022−01−08
2022−01−15

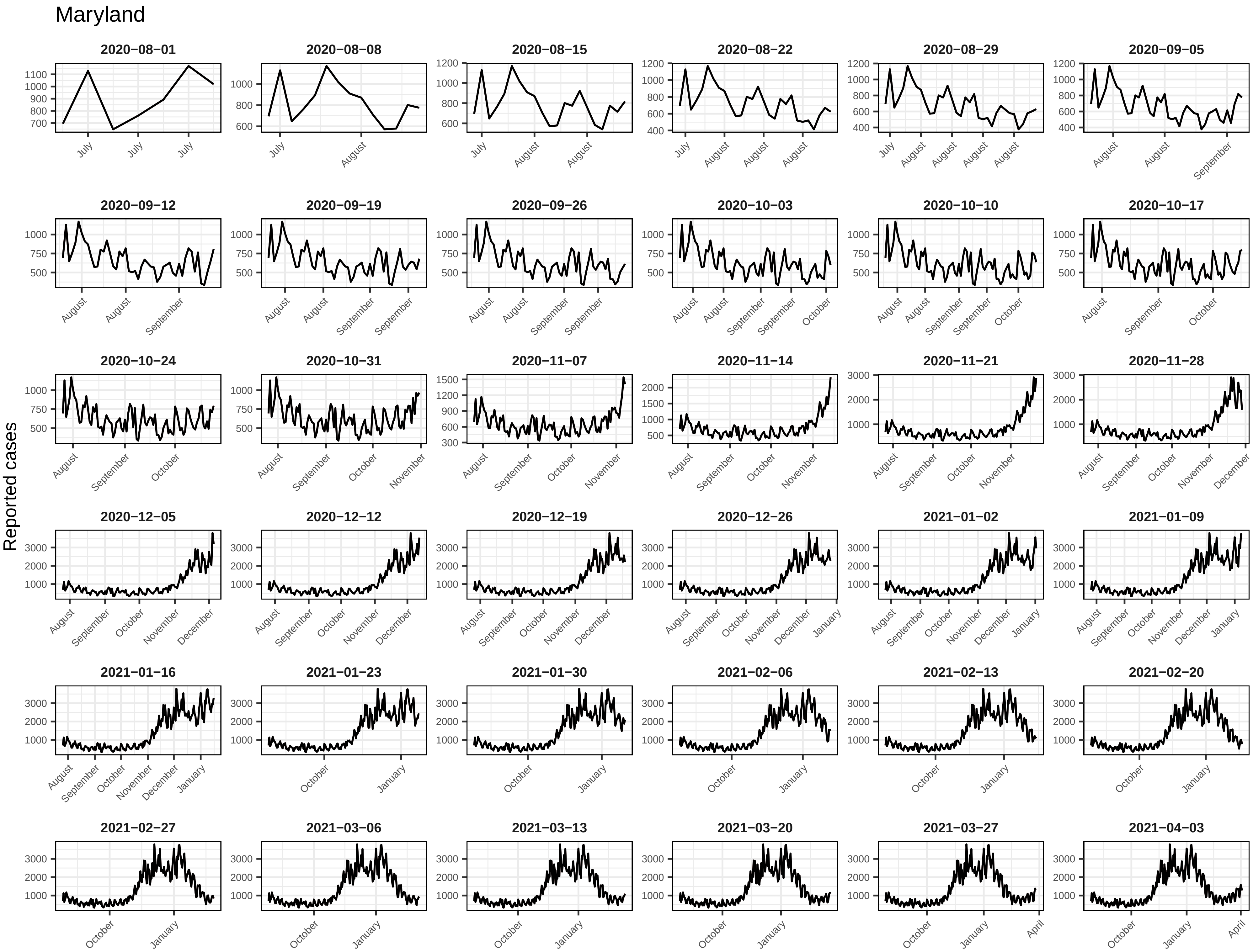
Maryland
Reported cases
2020−08−01
2020−08−08
2020−08−15
2020−08−22
2020−08−29
2020−09−05
2020−09−12
2020−09−19
2020−09−26
2020−10−03
2020−10−10
2020−10−17
2020−10−24
2020−10−31
2020−11−07
2020−11−14
2020−11−21
2020−11−28
2020−12−05
2020−12−12
2020−12−19
2020−12−26
2021−01−02
2021−01−09
2021−01−16
2021−01−23
2021−01−30
2021−02−06
2021−02−13
2021−02−20
2021−02−27
2021−03−06
2021−03−13
2021−03−20
2021−03−27
2021−04−03

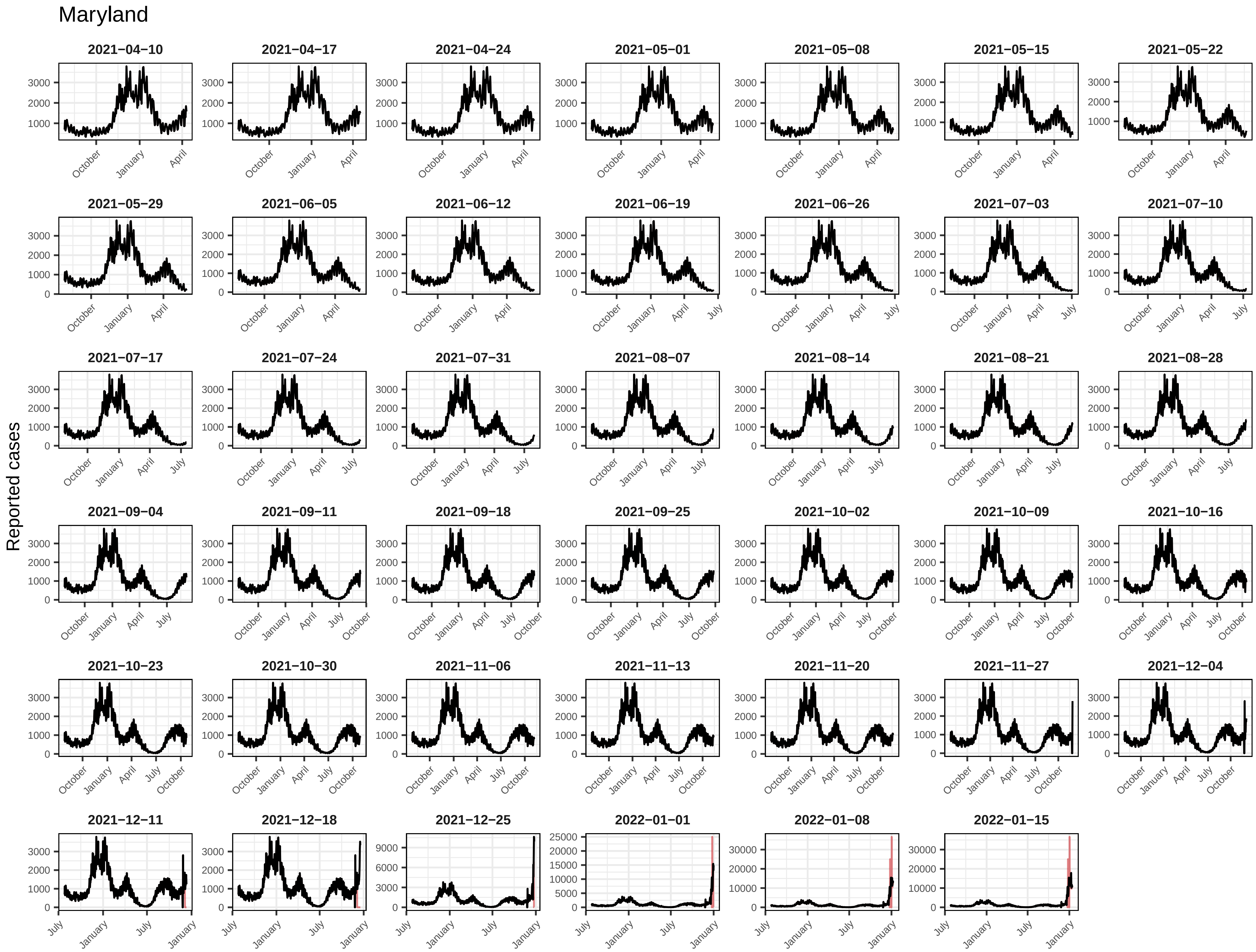
Maryland
Reported cases
2021−04−10
2021−04−17
2021−04−24
2021−05−01
2021−05−08
2021−05−15
2021−05−22
2021−05−29
2021−06−05
2021−06−12
2021−06−19
2021−06−26
2021−07−03
2021−07−10
2021−07−17
2021−07−24
2021−07−31
2021−08−07
2021−08−14
2021−08−21
2021−08−28
2021−09−04
2021−09−11
2021−09−18
2021−09−25
2021−10−02
2021−10−09
2021−10−16
2021−10−23
2021−10−30
2021−11−06
2021−11−13
2021−11−20
2021−11−27
2021−12−04
2021−12−11
2021−12−18
2021−12−25
2022−01−01
2022−01−08
2022−01−15
October
January
April
July

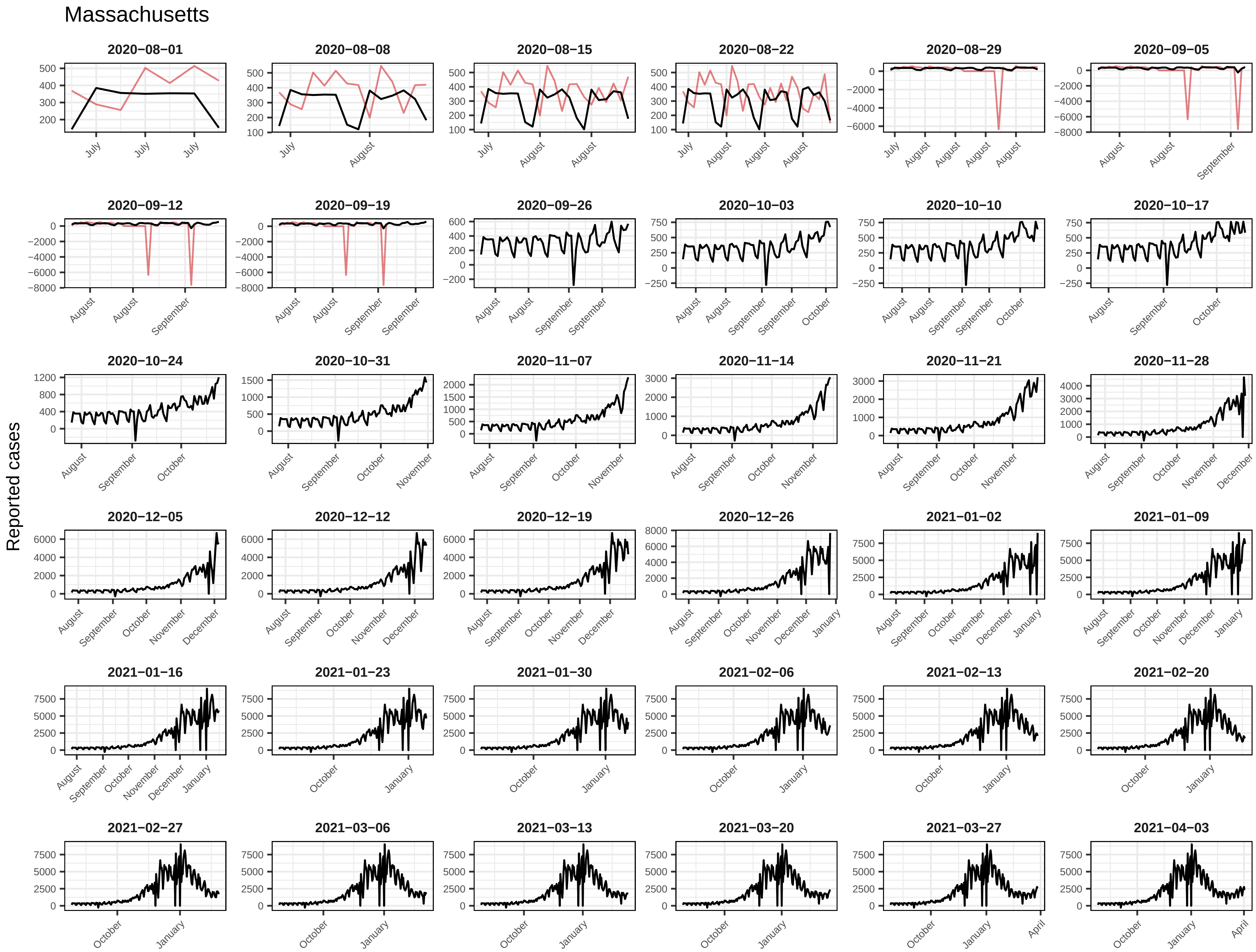
Massachusetts
Reported cases
2020−08−01
2020−08−08
2020−08−15
2020−08−22
2020−08−29
2020−09−05
2020−09−12
2020−09−19
2020−09−26
2020−10−03
2020−10−10
2020−10−17
2020−10−24
2020−10−31
2020−11−07
2020−11−14
2020−11−21
2020−11−28
2020−12−05
2020−12−12
2020−12−19
2020−12−26
2021−01−02
2021−01−09
2021−01−16
2021−01−23
2021−01−30
2021−02−06
2021−02−13
2021−02−20
2021−02−27
2021−03−06
2021−03−13
2021−03−20
2021−03−27
2021−04−03

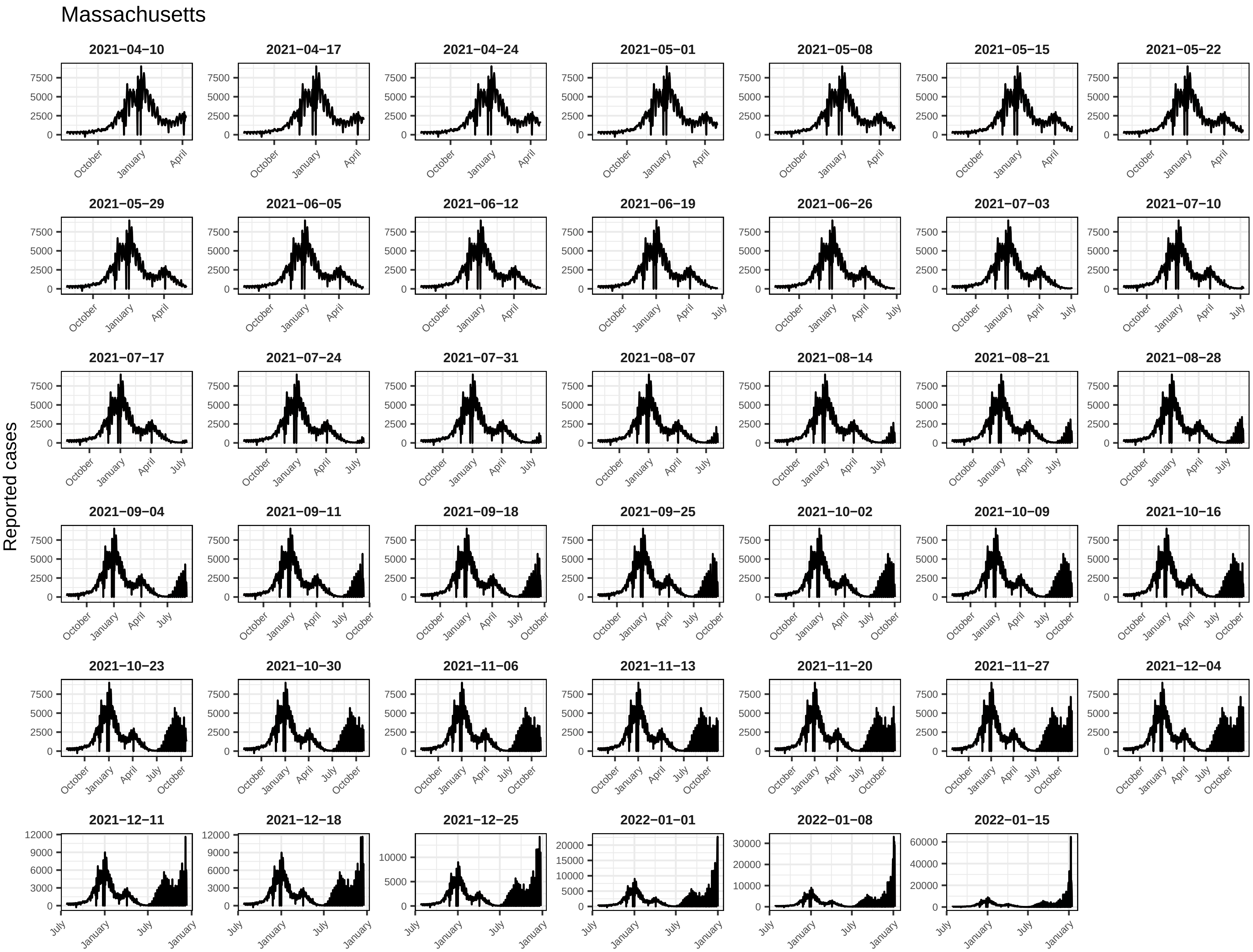
Massachusetts
Reported cases
2021−04−10
2021−04−17
2021−04−24
2021−05−01
2021−05−08
2021−05−15
2021−05−22
2021−05−29
2021−06−05
2021−06−12
2021−06−19
2021−06−26
2021−07−03
2021−07−10
2021−07−17
2021−07−24
2021−07−31
2021−08−07
2021−08−14
2021−08−21
2021−08−28
2021−09−04
2021−09−11
2021−09−18
2021−09−25
2021−10−02
2021−10−09
2021−10−16
2021−10−23
2021−10−30
2021−11−06
2021−11−13
2021−11−20
2021−11−27
2021−12−04
2021−12−11
2021−12−18
2021−12−25
2022−01−01
2022−01−08
2022−01−15
0
2500
5000
7500
3000
6000
9000
12000
10000
15000
20000
30000
40000
60000
October
January
April
July

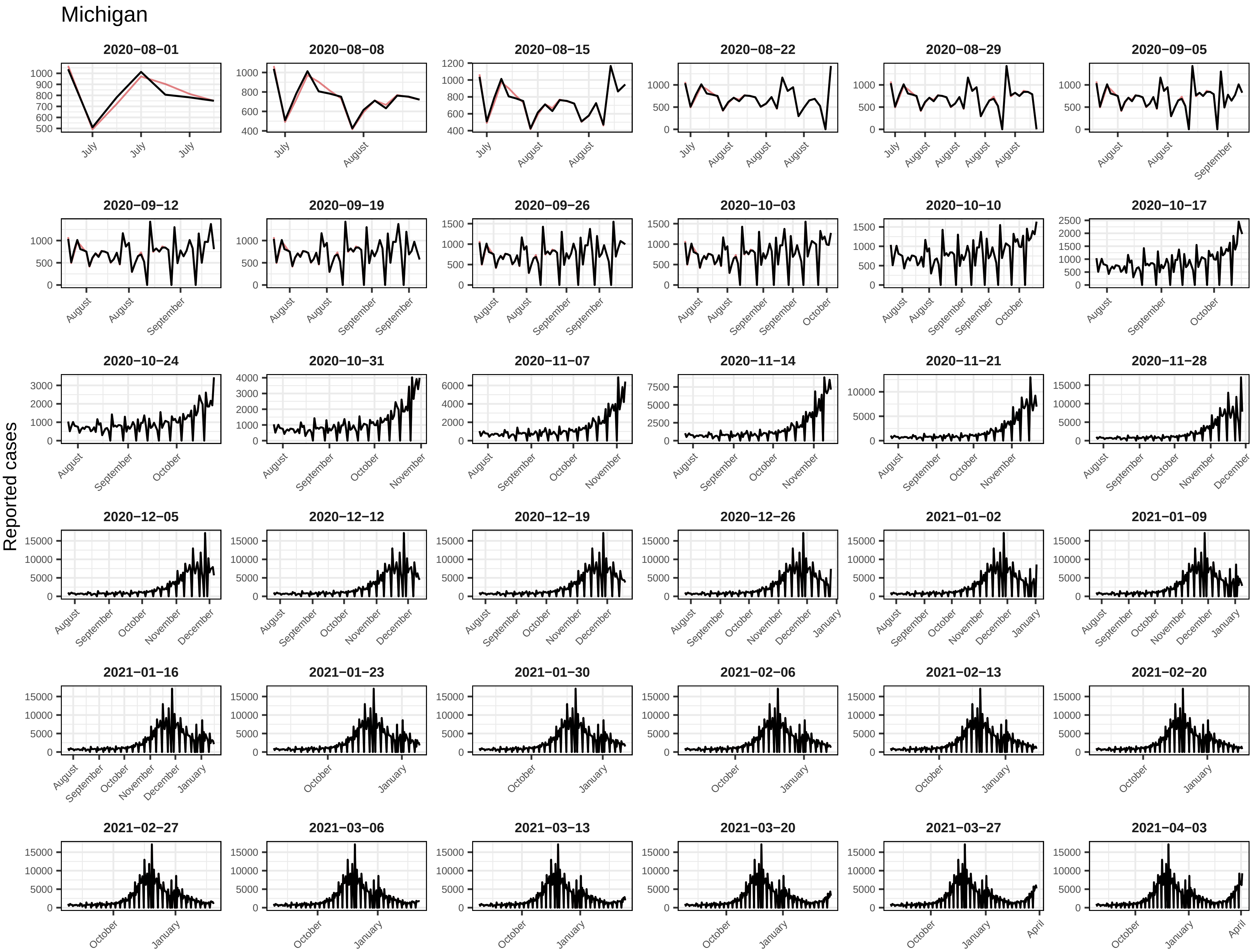
Michigan
Reported cases
2020−08−01
2020−08−08
2020−08−15
2020−08−22
2020−08−29
2020−09−05
2020−09−12
2020−09−19
2020−09−26
2020−10−03
2020−10−10
2020−10−17
2020−10−24
2020−10−31
2020−11−07
2020−11−14
2020−11−21
2020−11−28
2020−12−05
2020−12−12
2020−12−19
2020−12−26
2021−01−02
2021−01−09
2021−01−16
2021−01−23
2021−01−30
2021−02−06
2021−02−13
2021−02−20
2021−02−27
2021−03−06
2021−03−13
2021−03−20
2021−03−27
2021−04−03

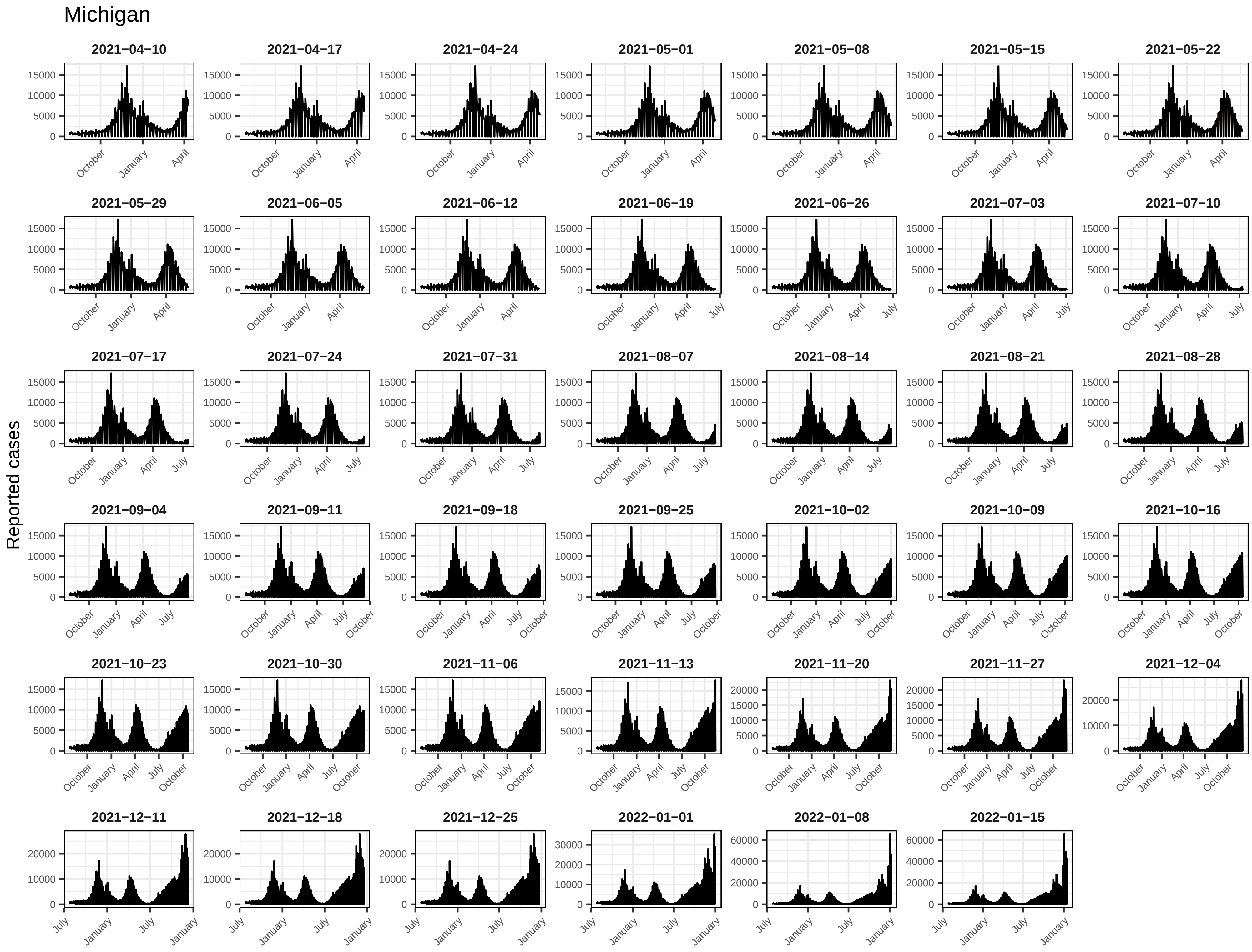
Michigan
Reported cases
2021−04−10
2021−04−17
2021−04−24
2021−05−01
2021−05−08
2021−05−15
2021−05−22
2021−05−29
2021−06−05
2021−06−12
2021−06−19
2021−06−26
2021−07−03
2021−07−10
2021−07−17
2021−07−24
2021−07−31
2021−08−07
2021−08−14
2021−08−21
2021−08−28
2021−09−04
2021−09−11
2021−09−18
2021−09−25
2021−10−02
2021−10−09
2021−10−16
2021−10−23
2021−10−30
2021−11−06
2021−11−13
2021−11−20
2021−11−27
2021−12−04
2021−12−11
2021−12−18
2021−12−25
2022−01−01
2022−01−08
2022−01−15
0
5000
10000
15000
20000
30000
40000
60000
July
October
January
April

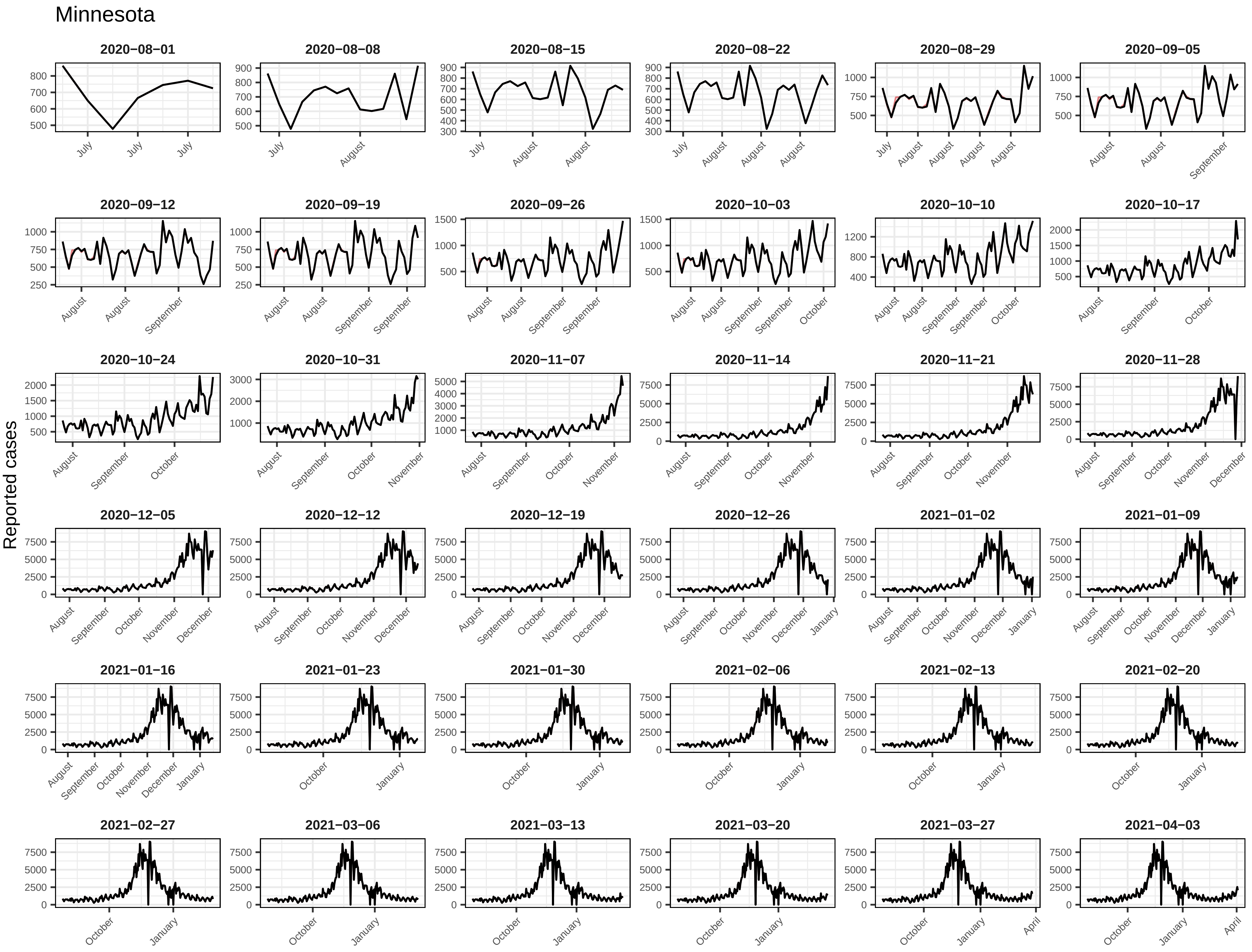
Minnesota
Reported cases
2020−08−01
2020−08−08
2020−08−15
2020−08−22
2020−08−29
2020−09−05
2020−09−12
2020−09−19
2020−09−26
2020−10−03
2020−10−10
2020−10−17
2020−10−24
2020−10−31
2020−11−07
2020−11−14
2020−11−21
2020−11−28
2020−12−05
2020−12−12
2020−12−19
2020−12−26
2021−01−02
2021−01−09
2021−01−16
2021−01−23
2021−01−30
2021−02−06
2021−02−13
2021−02−20
2021−02−27
2021−03−06
2021−03−13
2021−03−20
2021−03−27
2021−04−03

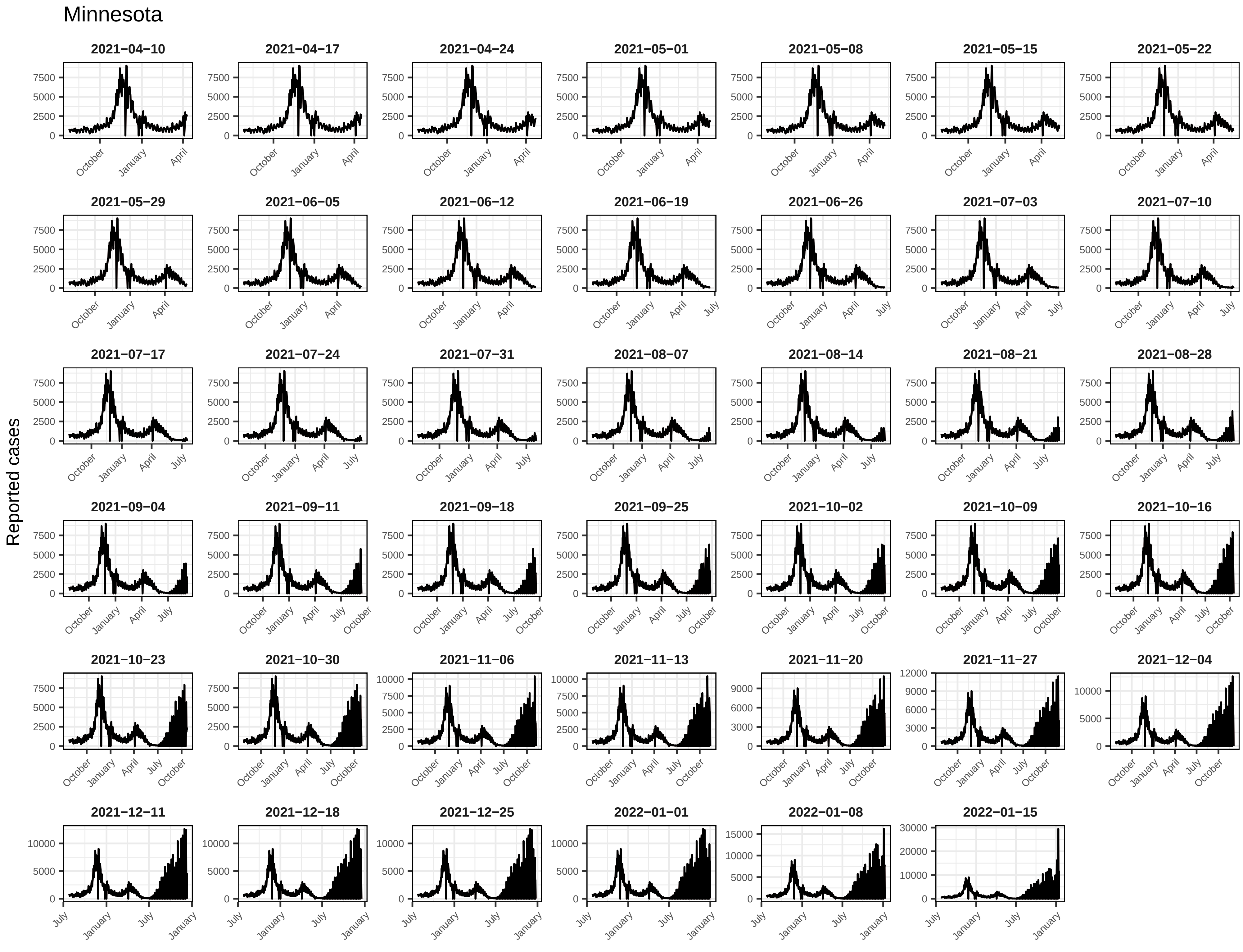

Minnesota
Reported cases
2021−04−10
2021−04−17
2021−04−24
2021−05−01
2021−05−08
2021−05−15
2021−05−22
2021−05−29
2021−06−05
2021−06−12
2021−06−19
2021−06−26
2021−07−03
2021−07−10
2021−07−17
2021−07−24
2021−07−31
2021−08−07
2021−08−14
2021−08−21
2021−08−28
2021−09−04
2021−09−11
2021−09−18
2021−09−25
2021−10−02
2021−10−09
2021−10−16
2021−10−23
2021−10−30
2021−11−06
2021−11−13
2021−11−20
2021−11−27
2021−12−04
2021−12−11
2021−12−18
2021−12−25
2022−01−01
2022−01−08
2022−01−15

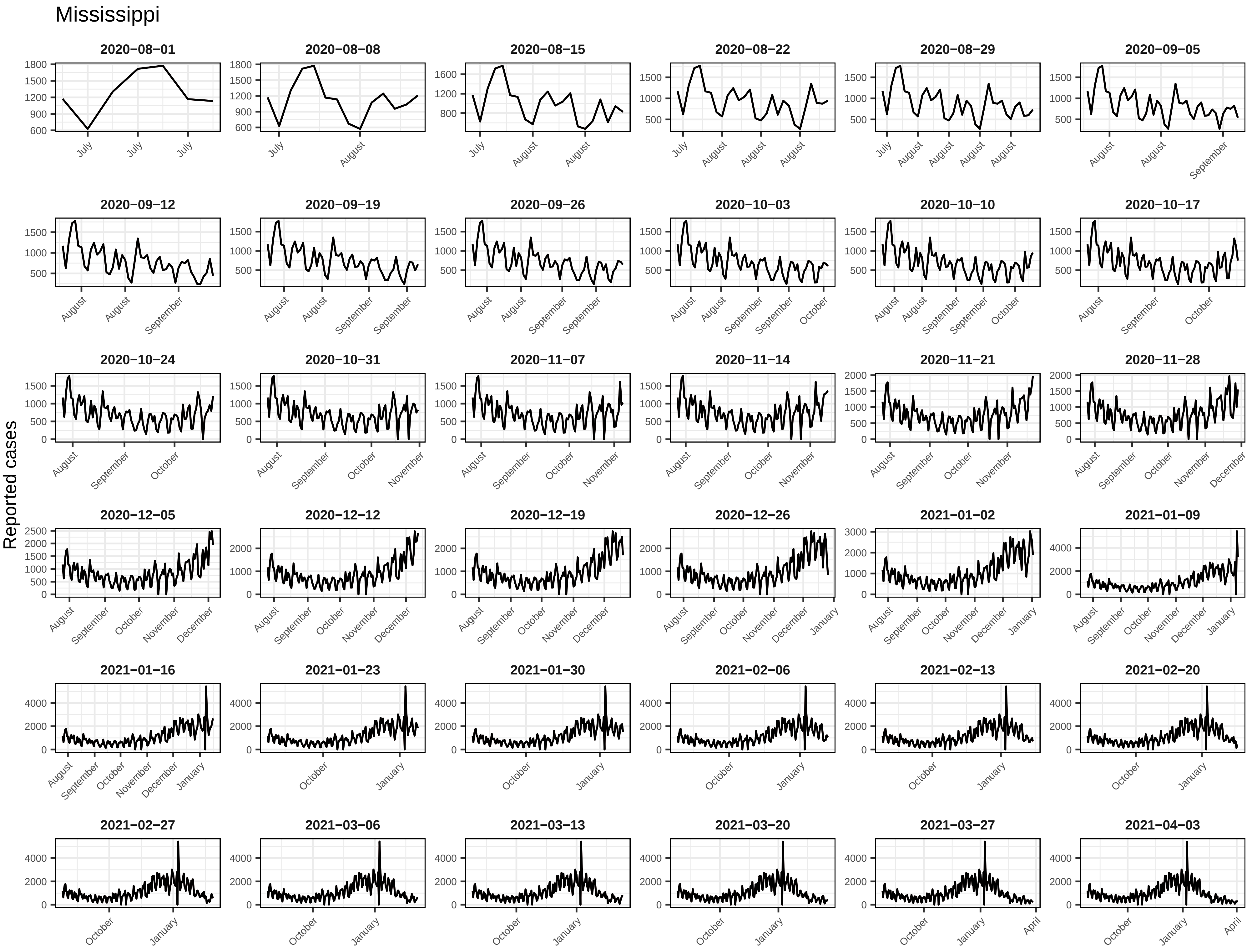
Mississippi
Reported cases
2020−08−01
2020−08−08
2020−08−15
2020−08−22
2020−08−29
2020−09−05
2020−09−12
2020−09−19
2020−09−26
2020−10−03
2020−10−10
2020−10−17
2020−10−24
2020−10−31
2020−11−07
2020−11−14
2020−11−21
2020−11−28
2020−12−05
2020−12−12
2020−12−19
2020−12−26
2021−01−02
2021−01−09
2021−01−16
2021−01−23
2021−01−30
2021−02−06
2021−02−13
2021−02−20
2021−02−27
2021−03−06
2021−03−13
2021−03−20
2021−03−27
2021−04−03

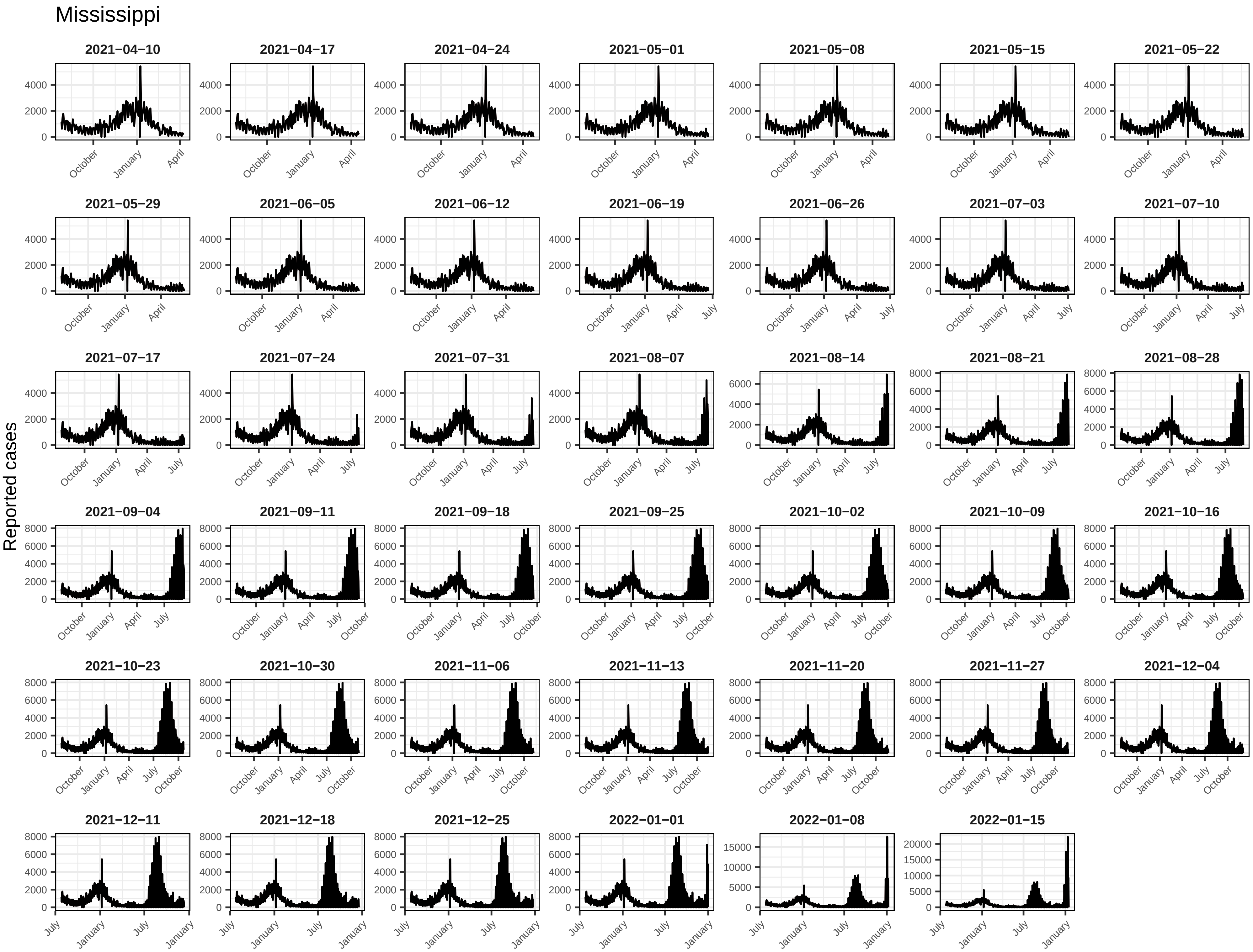
Mississippi
Reported cases
2021−04−10
2021−04−17
2021−04−24
2021−05−01
2021−05−08
2021−05−15
2021−05−22
2021−05−29
2021−06−05
2021−06−12
2021−06−19
2021−06−26
2021−07−03
2021−07−10
2021−07−17
2021−07−24
2021−07−31
2021−08−07
2021−08−14
2021−08−21
2021−08−28
2021−09−04
2021−09−11
2021−09−18
2021−09−25
2021−10−02
2021−10−09
2021−10−16
2021−10−23
2021−10−30
2021−11−06
2021−11−13
2021−11−20
2021−11−27
2021−12−04
2021−12−11
2021−12−18
2021−12−25
2022−01−01
2022−01−08
2022−01−15

Missouri

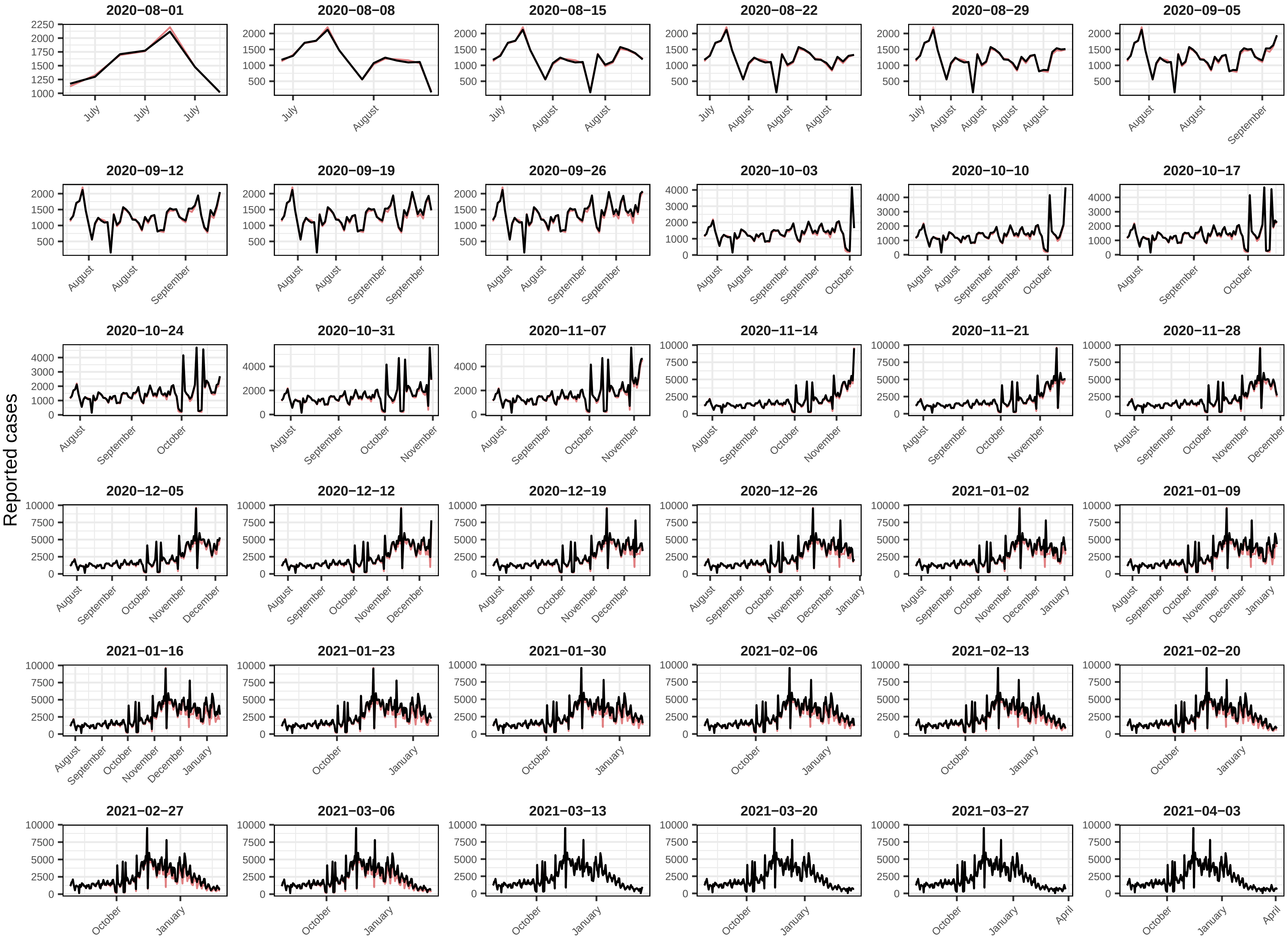

Reported cases
2020−08−01
2020−08−08
2020−08−15
2020−08−22
2020−08−29
2020−09−05
2020−09−12
2020−09−19
2020−09−26
2020−10−03
2020−10−10
2020−10−17
2020−10−24
2020−10−31
2020−11−07
2020−11−14
2020−11−21
2020−11−28
2020−12−05
2020−12−12
2020−12−19
2020−12−26
2021−01−02
2021−01−09
2021−01−16
2021−01−23
2021−01−30
2021−02−06
2021−02−13
2021−02−20
2021−02−27
2021−03−06
2021−03−13
2021−03−20
2021−03−27
2021−04−03

### Missouri

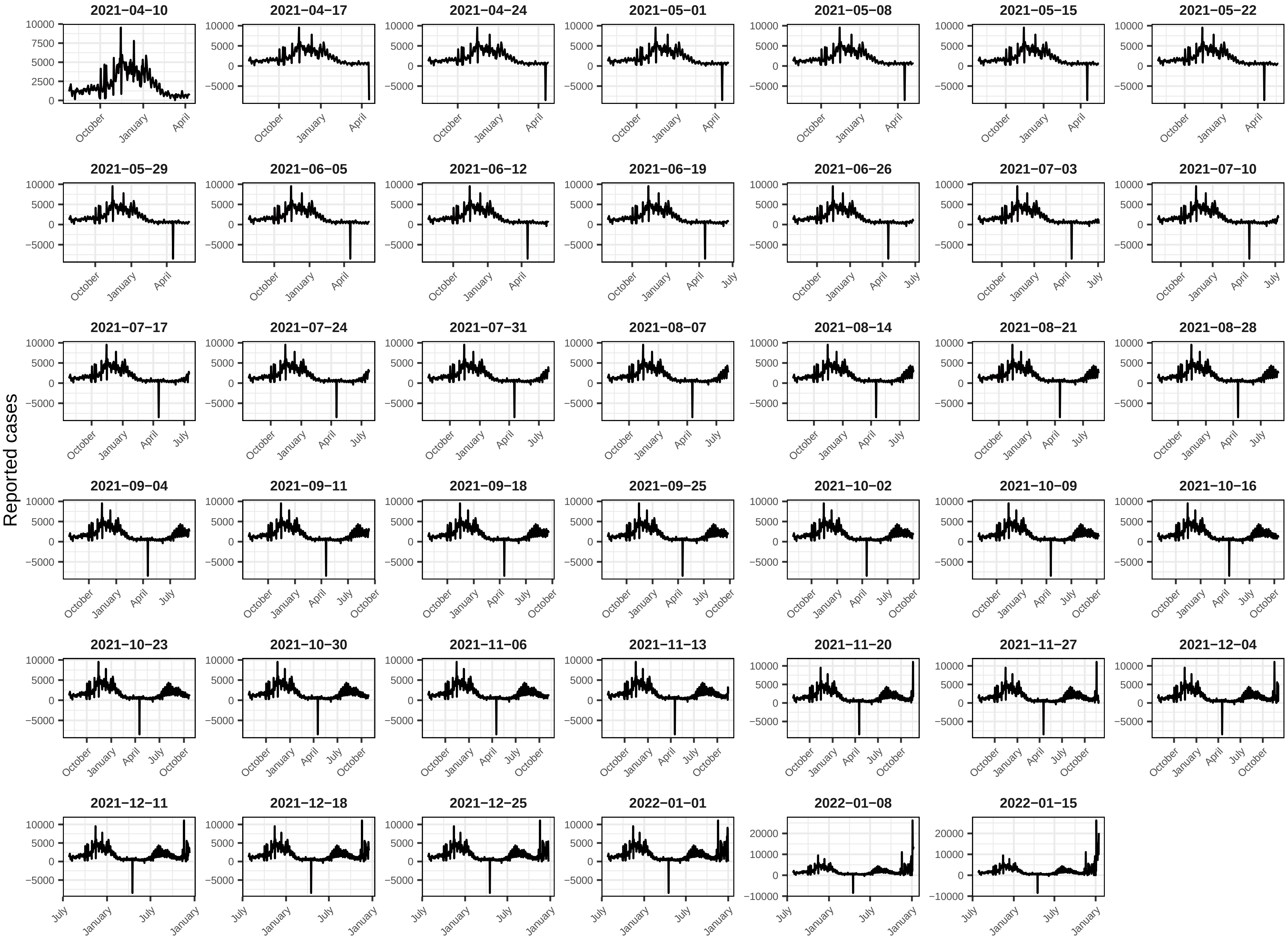
Reported cases
2021−04−10
2021−04−17
2021−04−24
2021−05−01
2021−05−08
2021−05−15
2021−05−22
2021−05−29
2021−06−05
2021−06−12
2021−06−19
2021−06−26
2021−07−03
2021−07−10
2021−07−17
2021−07−24
2021−07−31
2021−08−07
2021−08−14
2021−08−21
2021−08−28
2021−09−04
2021−09−11
2021−09−18
2021−09−25
2021−10−02
2021−10−09
2021−10−16
2021−10−23
2021−10−30
2021−11−06
2021−11−13
2021−11−20
2021−11−27
2021−12−04
2021−12−11
2021−12−18
2021−12−25
2022−01−01
2022−01−08
2022−01−15
10000
7500
5000
2500
0
−5000
20000
−10000
July
October
January
April

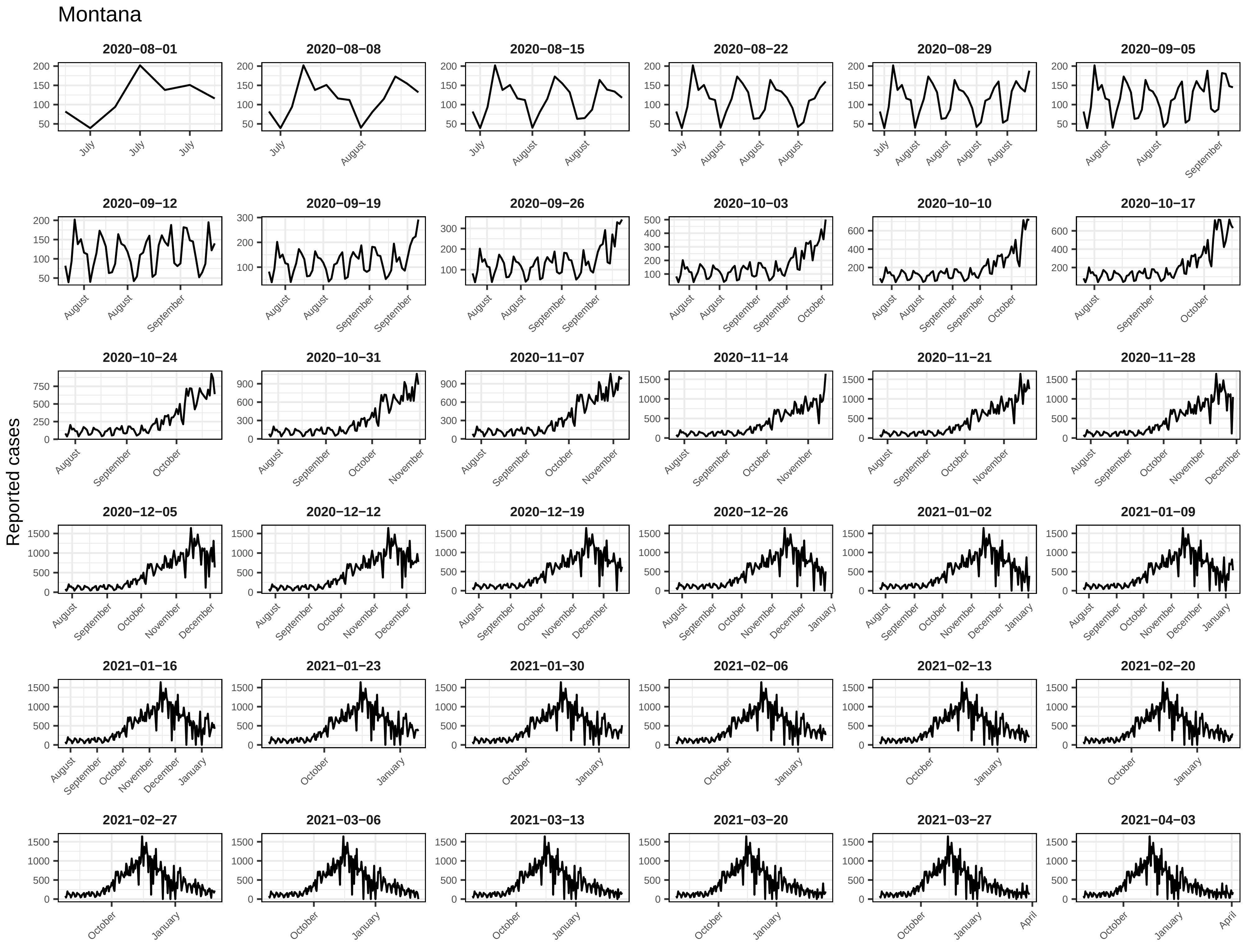
Montana
Reported cases
2020−08−01
2020−08−08
2020−08−15
2020−08−22
2020−08−29
2020−09−05
2020−09−12
2020−09−19
2020−09−26
2020−10−03
2020−10−10
2020−10−17
2020−10−24
2020−10−31
2020−11−07
2020−11−14
2020−11−21
2020−11−28
2020−12−05
2020−12−12
2020−12−19
2020−12−26
2021−01−02
2021−01−09
2021−01−16
2021−01−23
2021−01−30
2021−02−06
2021−02−13
2021−02−20
2021−02−27
2021−03−06
2021−03−13
2021−03−20
2021−03−27
2021−04−03

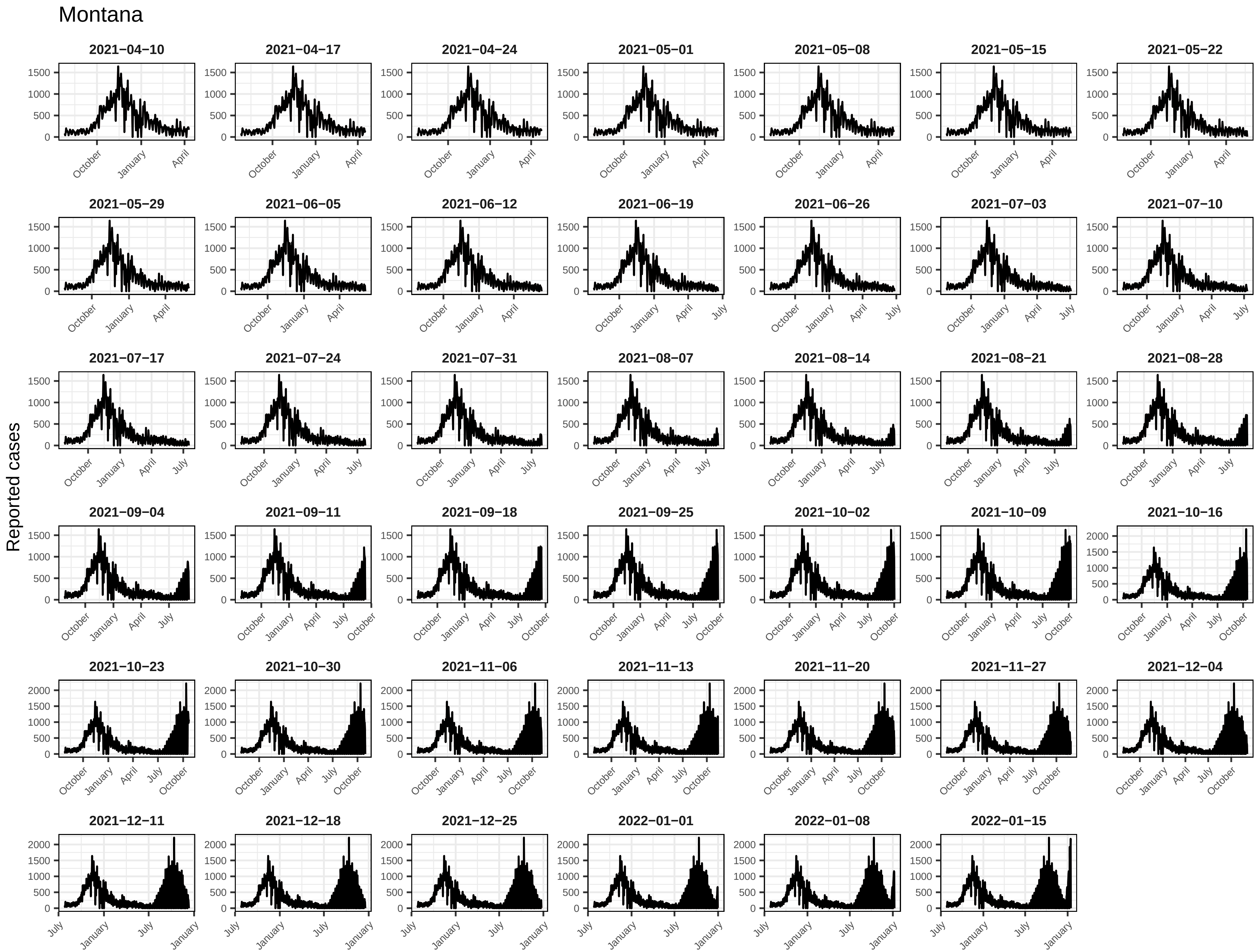
Montana
Reported cases
2021−04−10
2021−04−17
2021−04−24
2021−05−01
2021−05−08
2021−05−15
2021−05−22
2021−05−29
2021−06−05
2021−06−12
2021−06−19
2021−06−26
2021−07−03
2021−07−10
2021−07−17
2021−07−24
2021−07−31
2021−08−07
2021−08−14
2021−08−21
2021−08−28
2021−09−04
2021−09−11
2021−09−18
2021−09−25
2021−10−02
2021−10−09
2021−10−16
2021−10−23
2021−10−30
2021−11−06
2021−11−13
2021−11−20
2021−11−27
2021−12−04
2021−12−11
2021−12−18
2021−12−25
2022−01−01
2022−01−08
2022−01−15
0
500
1000
1500
2000
October
January
April
July

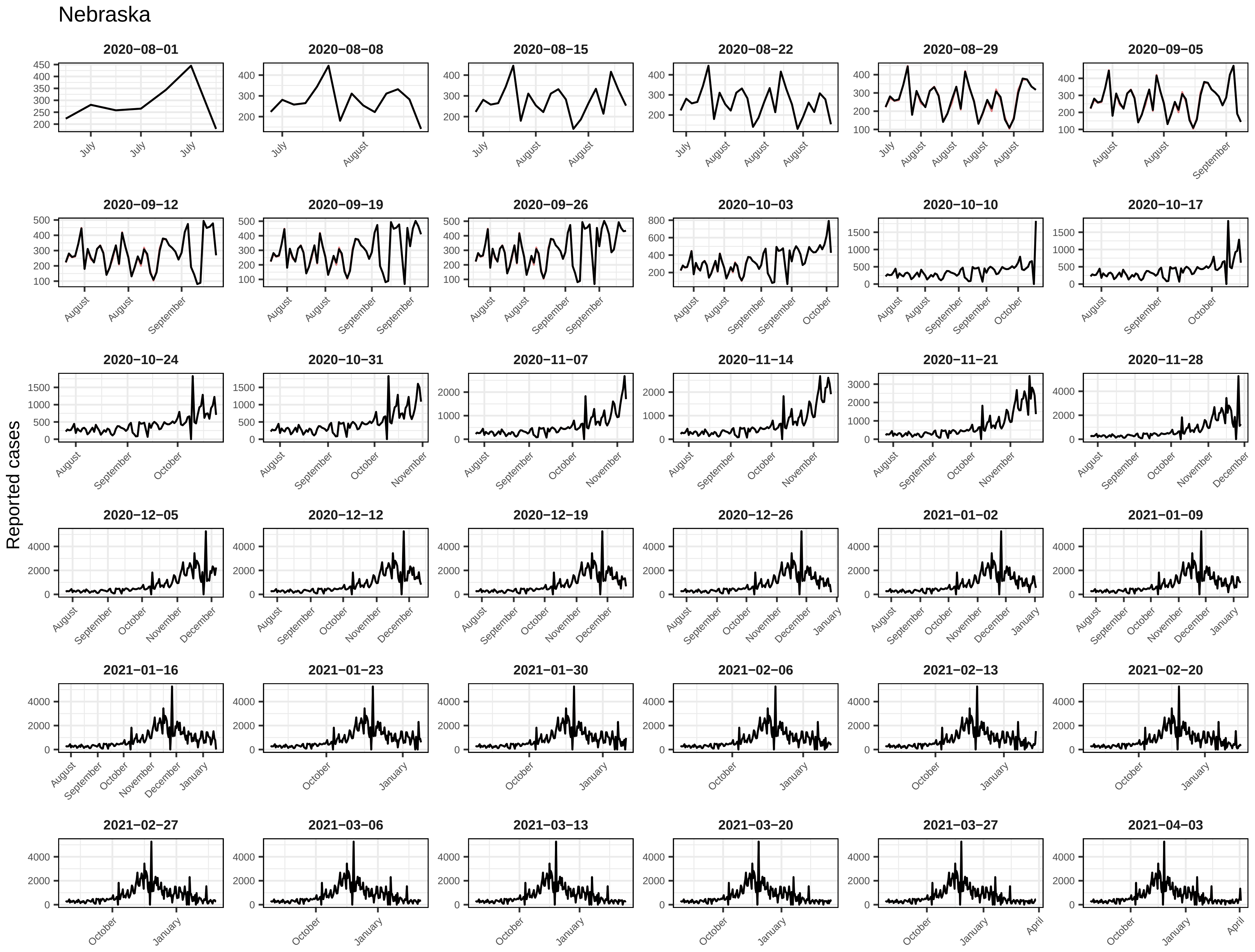
Nebraska
Reported cases
2020−08−01
2020−08−08
2020−08−15
2020−08−22
2020−08−29
2020−09−05
2020−09−12
2020−09−19
2020−09−26
2020−10−03
2020−10−10
2020−10−17
2020−10−24
2020−10−31
2020−11−07
2020−11−14
2020−11−21
2020−11−28
2020−12−05
2020−12−12
2020−12−19
2020−12−26
2021−01−02
2021−01−09
2021−01−16
2021−01−23
2021−01−30
2021−02−06
2021−02−13
2021−02−20
2021−02−27
2021−03−06
2021−03−13
2021−03−20
2021−03−27
2021−04−03

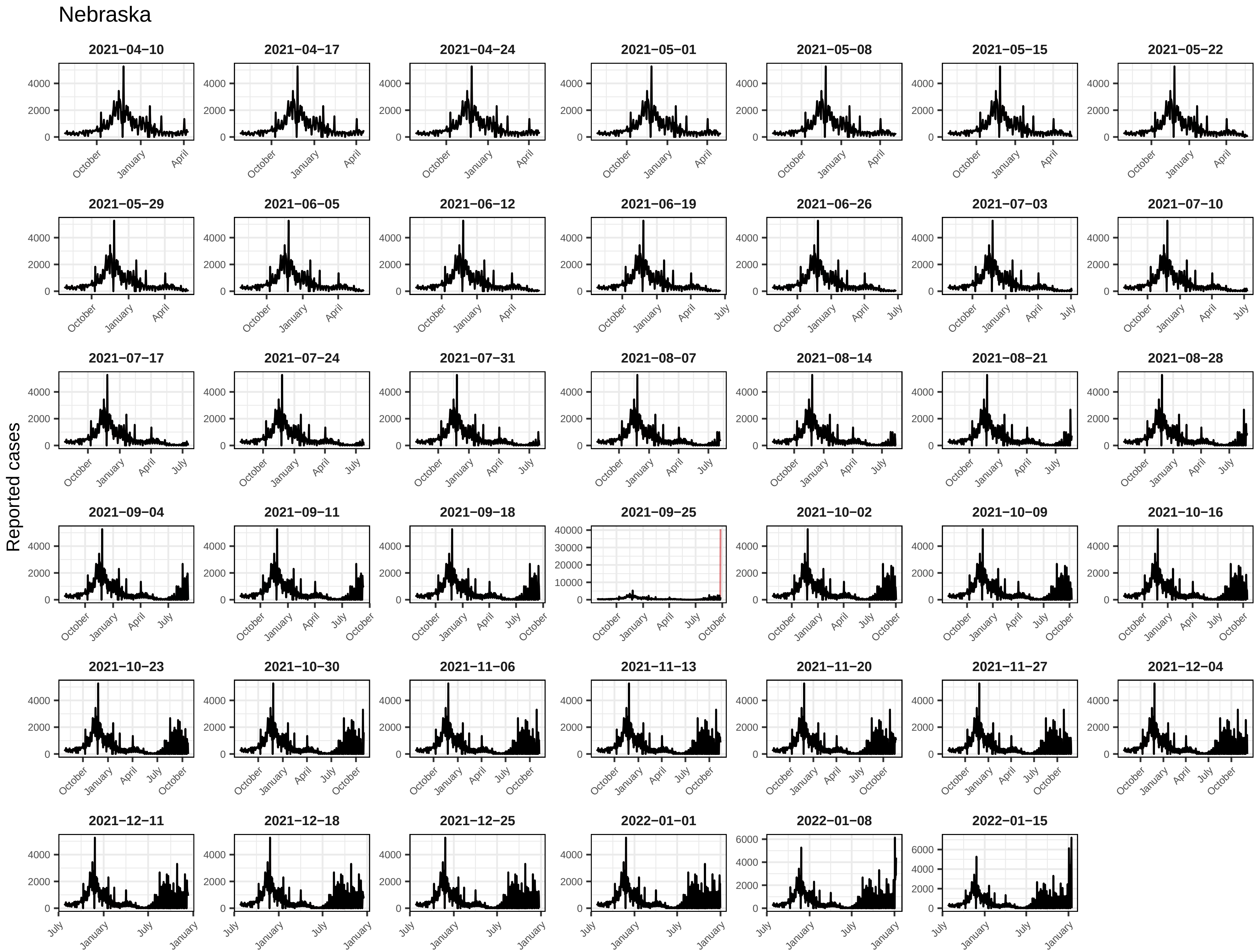

Nebraska
Reported cases
2021−04−10
2021−04−17
2021−04−24
2021−05−01
2021−05−08
2021−05−15
2021−05−22
2021−05−29
2021−06−05
2021−06−12
2021−06−19
2021−06−26
2021−07−03
2021−07−10
2021−07−17
2021−07−24
2021−07−31
2021−08−07
2021−08−14
2021−08−21
2021−08−28
2021−09−04
2021−09−11
2021−09−18
2021−09−25
2021−10−02
2021−10−09
2021−10−16
2021−10−23
2021−10−30
2021−11−06
2021−11−13
2021−11−20
2021−11−27
2021−12−04
2021−12−11
2021−12−18
2021−12−25
2022−01−01
2022−01−08
2022−01−15

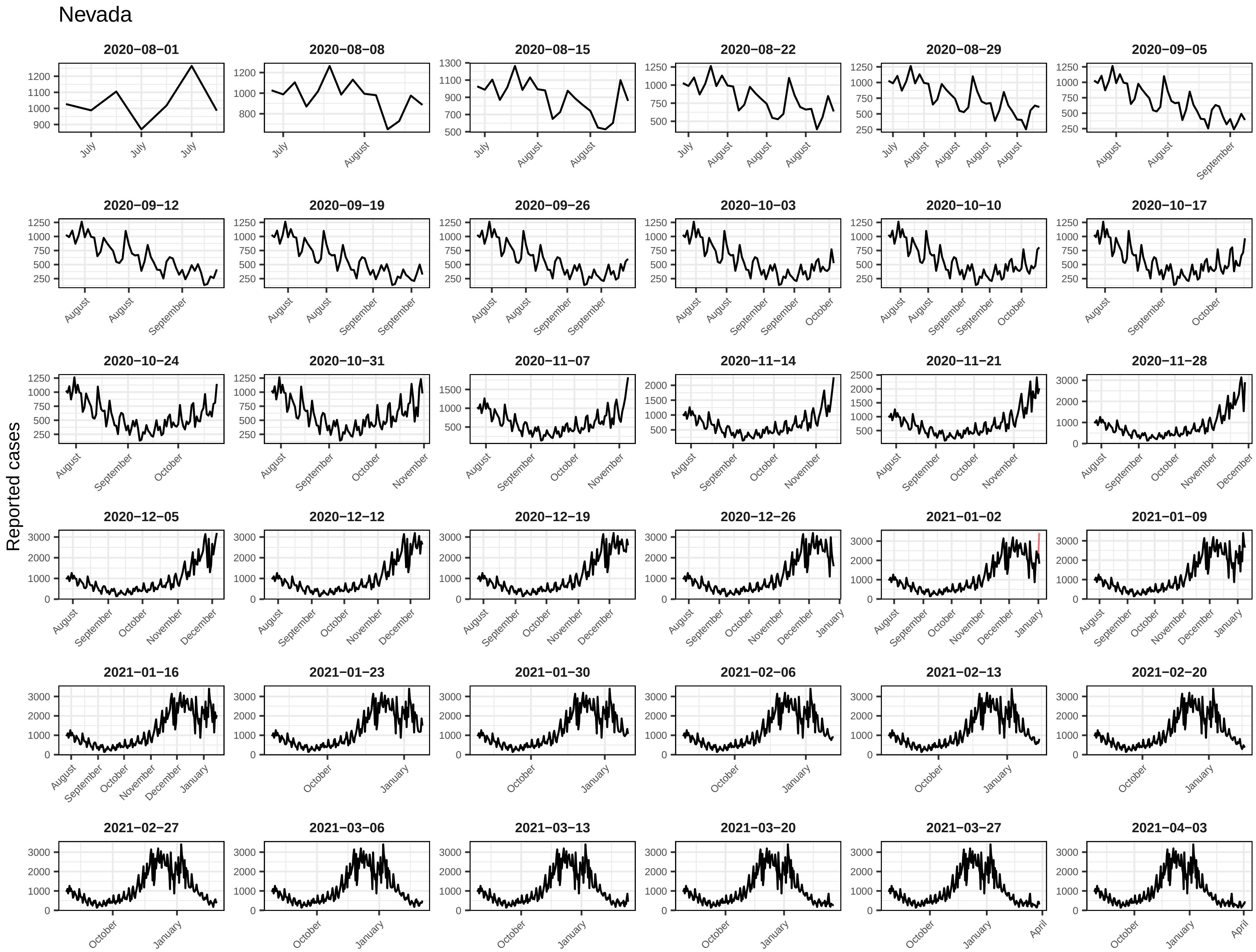
Nevada
Reported cases
2020−08−01
2020−08−08
2020−08−15
2020−08−22
2020−08−29
2020−09−05
2020−09−12
2020−09−19
2020−09−26
2020−10−03
2020−10−10
2020−10−17
2020−10−24
2020−10−31
2020−11−07
2020−11−14
2020−11−21
2020−11−28
2020−12−05
2020−12−12
2020−12−19
2020−12−26
2021−01−02
2021−01−09
2021−01−16
2021−01−23
2021−01−30
2021−02−06
2021−02−13
2021−02−20
2021−02−27
2021−03−06
2021−03−13
2021−03−20
2021−03−27
2021−04−03

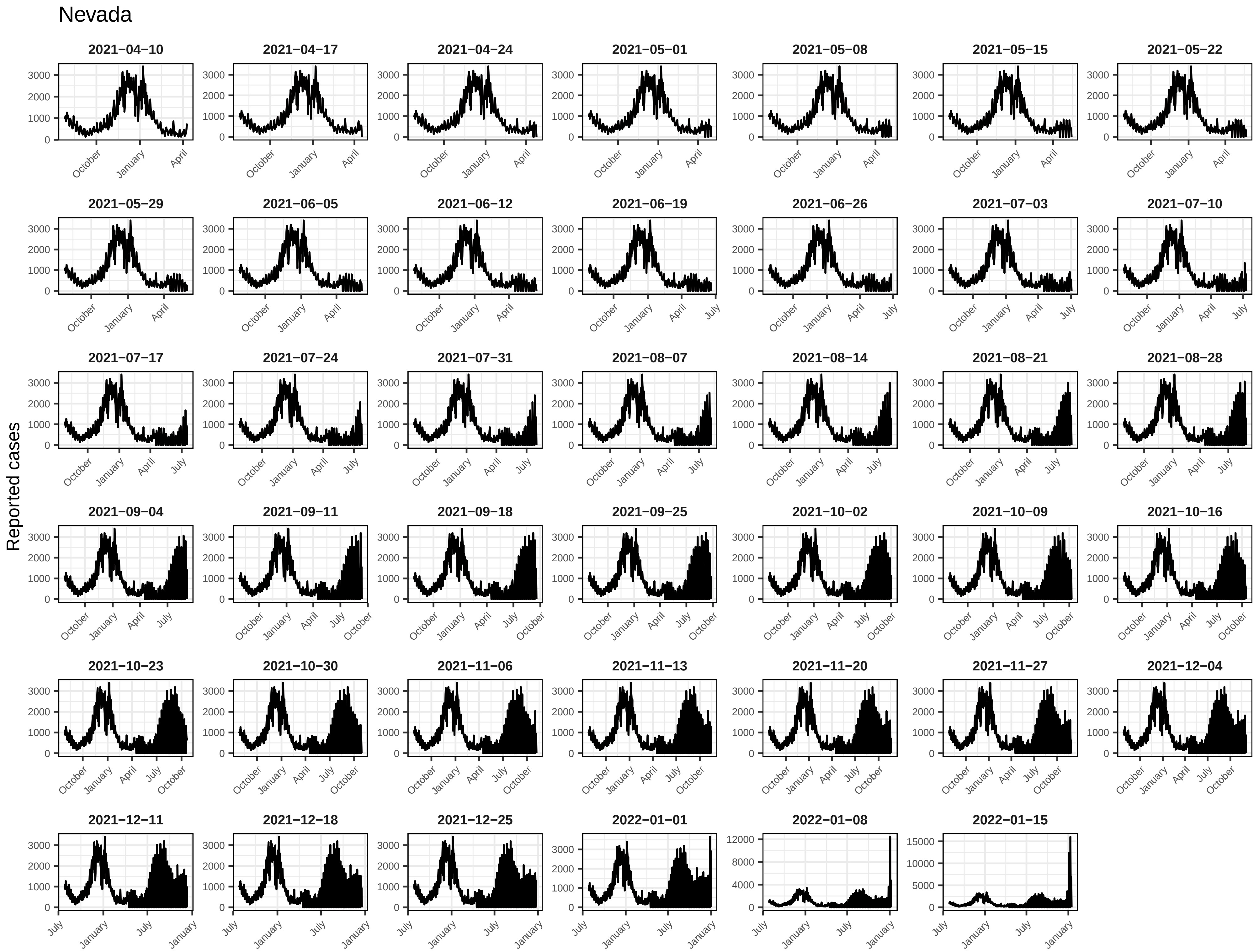
Nevada
Reported cases
2021−04−10
2021−04−17
2021−04−24
2021−05−01
2021−05−08
2021−05−15
2021−05−22
2021−05−29
2021−06−05
2021−06−12
2021−06−19
2021−06−26
2021−07−03
2021−07−10
2021−07−17
2021−07−24
2021−07−31
2021−08−07
2021−08−14
2021−08−21
2021−08−28
2021−09−04
2021−09−11
2021−09−18
2021−09−25
2021−10−02
2021−10−09
2021−10−16
2021−10−23
2021−10−30
2021−11−06
2021−11−13
2021−11−20
2021−11−27
2021−12−04
2021−12−11
2021−12−18
2021−12−25
2022−01−01
2022−01−08
2022−01−15
October
January
April
July

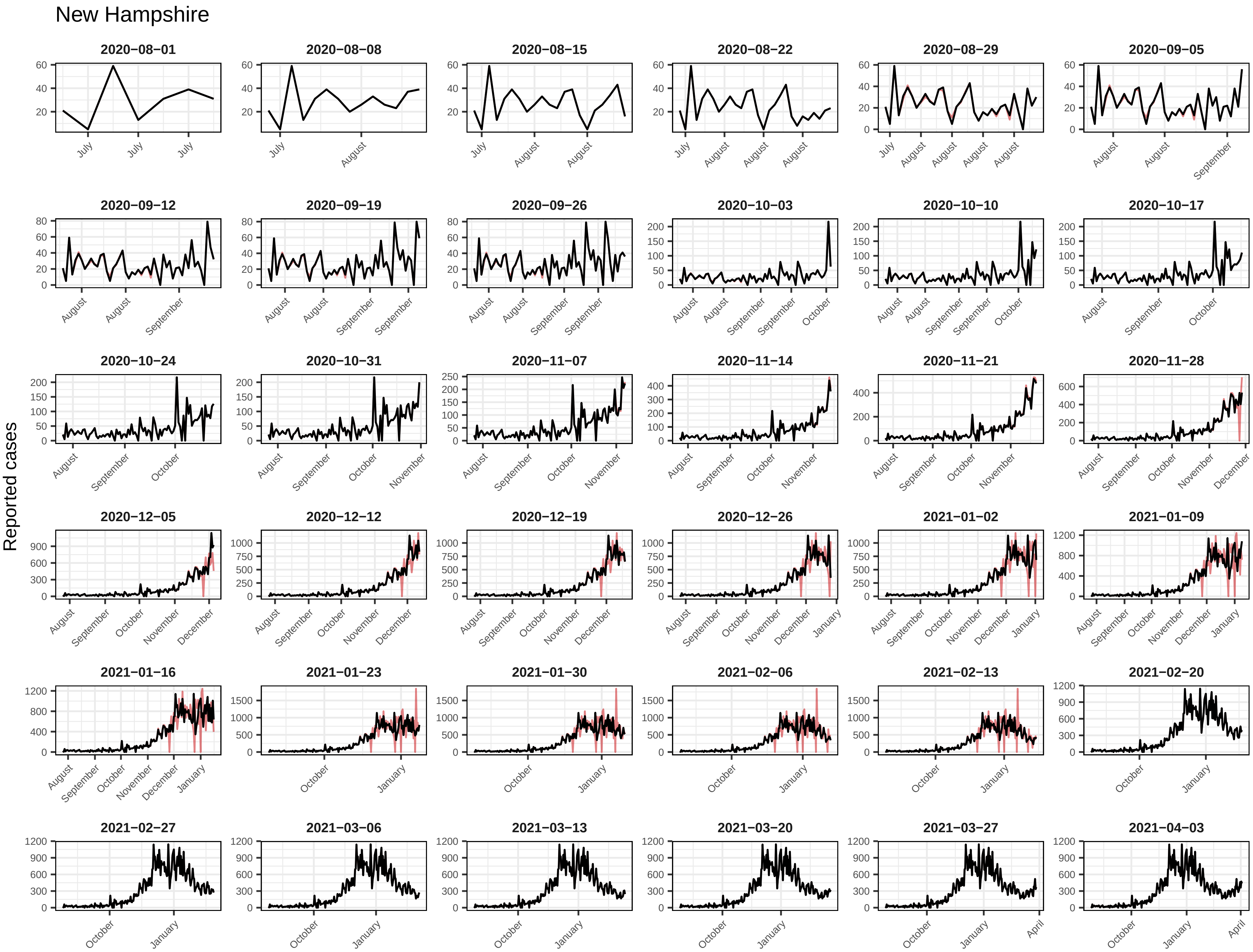
New Hampshire
Reported cases
2020−08−01
2020−08−08
2020−08−15
2020−08−22
2020−08−29
2020−09−05
2020−09−12
2020−09−19
2020−09−26
2020−10−03
2020−10−10
2020−10−17
2020−10−24
2020−10−31
2020−11−07
2020−11−14
2020−11−21
2020−11−28
2020−12−05
2020−12−12
2020−12−19
2020−12−26
2021−01−02
2021−01−09
2021−01−16
2021−01−23
2021−01−30
2021−02−06
2021−02−13
2021−02−20
2021−02−27
2021−03−06
2021−03−13
2021−03−20
2021−03−27
2021−04−03
July
August
September
October
November
December
January
April

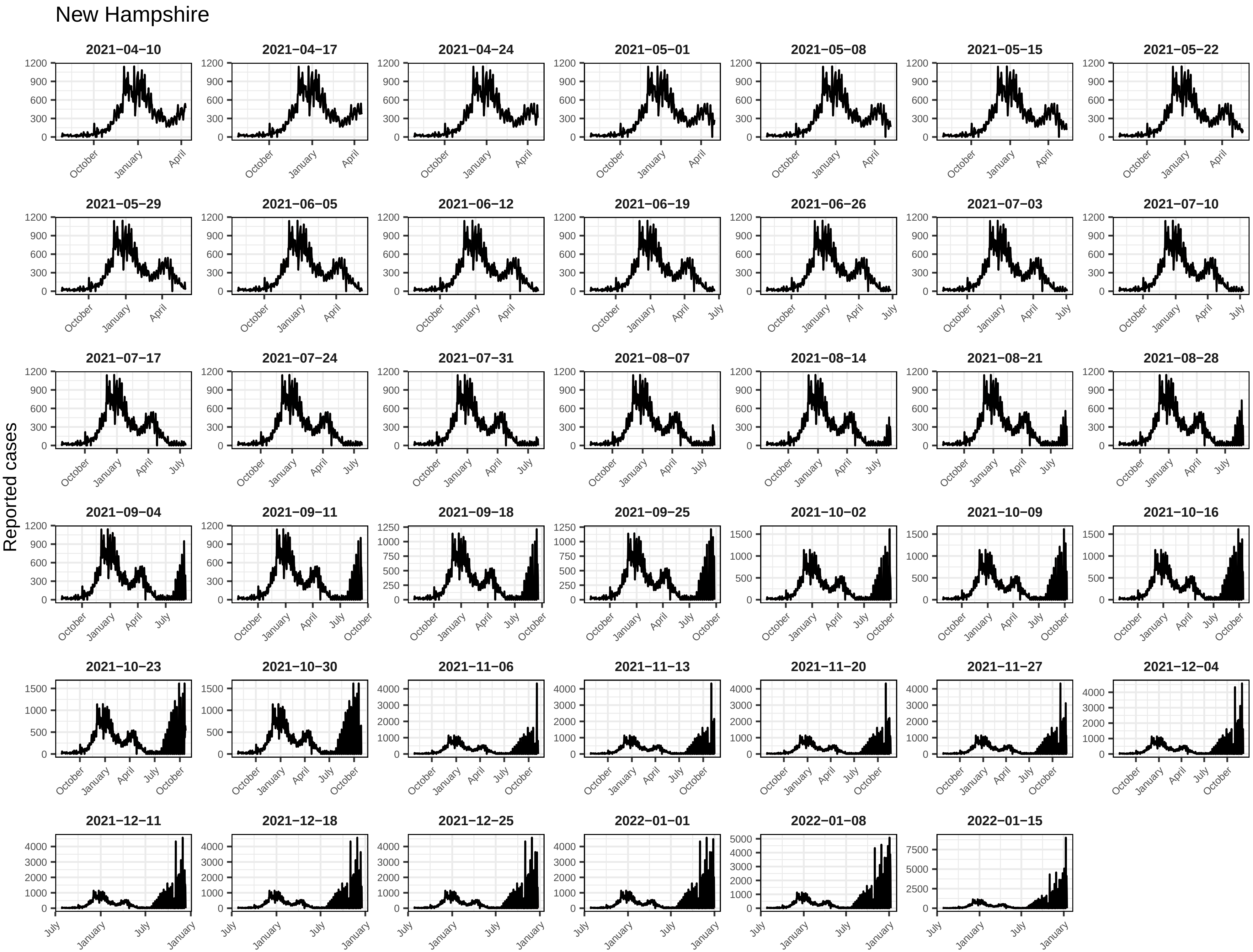
New Hampshire
Reported cases
2021−04−10
2021−04−17
2021−04−24
2021−05−01
2021−05−08
2021−05−15
2021−05−22
2021−05−29
2021−06−05
2021−06−12
2021−06−19
2021−06−26
2021−07−03
2021−07−10
2021−07−17
2021−07−24
2021−07−31
2021−08−07
2021−08−14
2021−08−21
2021−08−28
2021−09−04
2021−09−11
2021−09−18
2021−09−25
2021−10−02
2021−10−09
2021−10−16
2021−10−23
2021−10−30
2021−11−06
2021−11−13
2021−11−20
2021−11−27
2021−12−04
2021−12−11
2021−12−18
2021−12−25
2022−01−01
2022−01−08
2022−01−15
July
October
January
April

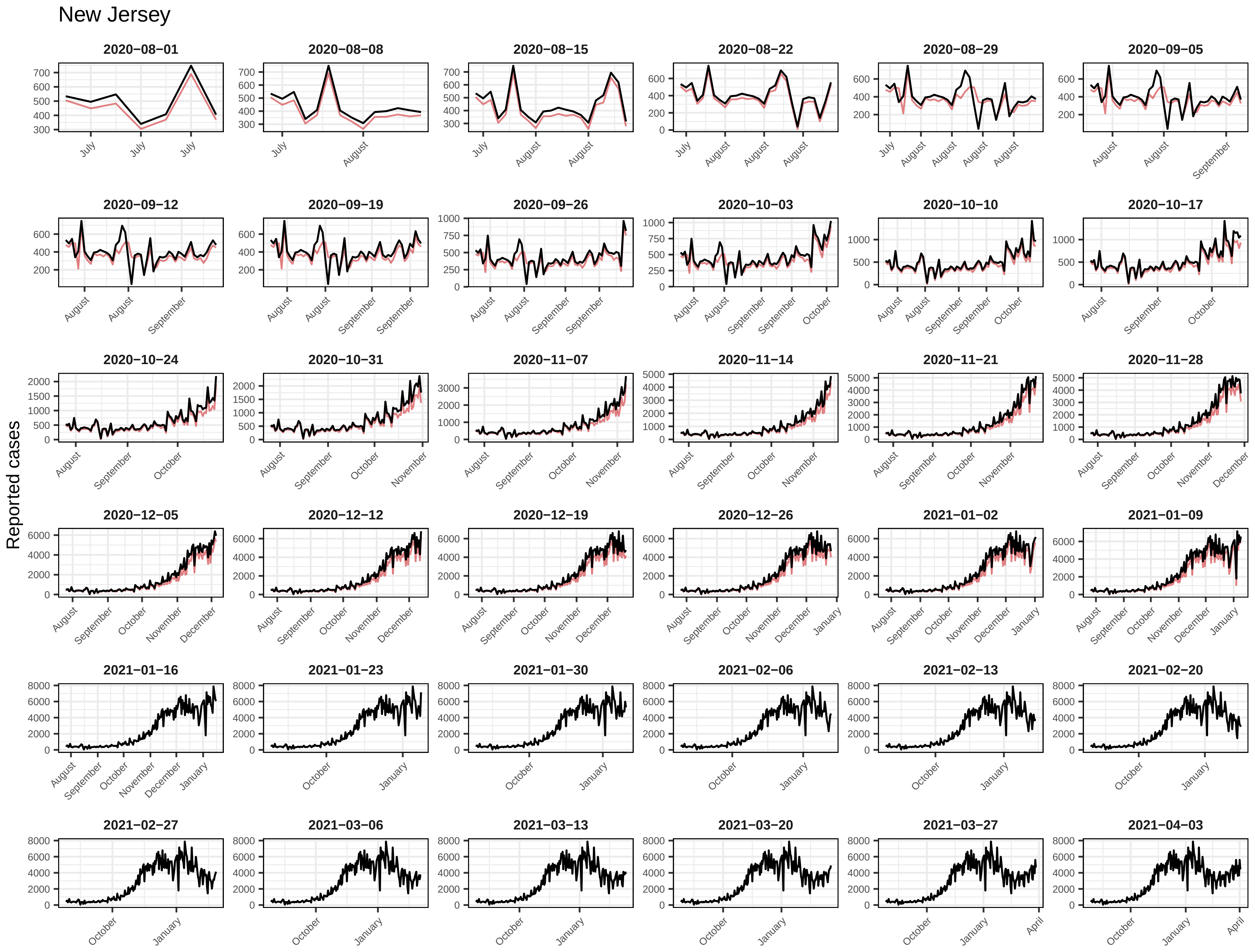
New Jersey
Reported cases
2020−08−01
2020−08−08
2020−08−15
2020−08−22
2020−08−29
2020−09−05
2020−09−12
2020−09−19
2020−09−26
2020−10−03
2020−10−10
2020−10−17
2020−10−24
2020−10−31
2020−11−07
2020−11−14
2020−11−21
2020−11−28
2020−12−05
2020−12−12
2020−12−19
2020−12−26
2021−01−02
2021−01−09
2021−01−16
2021−01−23
2021−01−30
2021−02−06
2021−02−13
2021−02−20
2021−02−27
2021−03−06
2021−03−13
2021−03−20
2021−03−27
2021−04−03

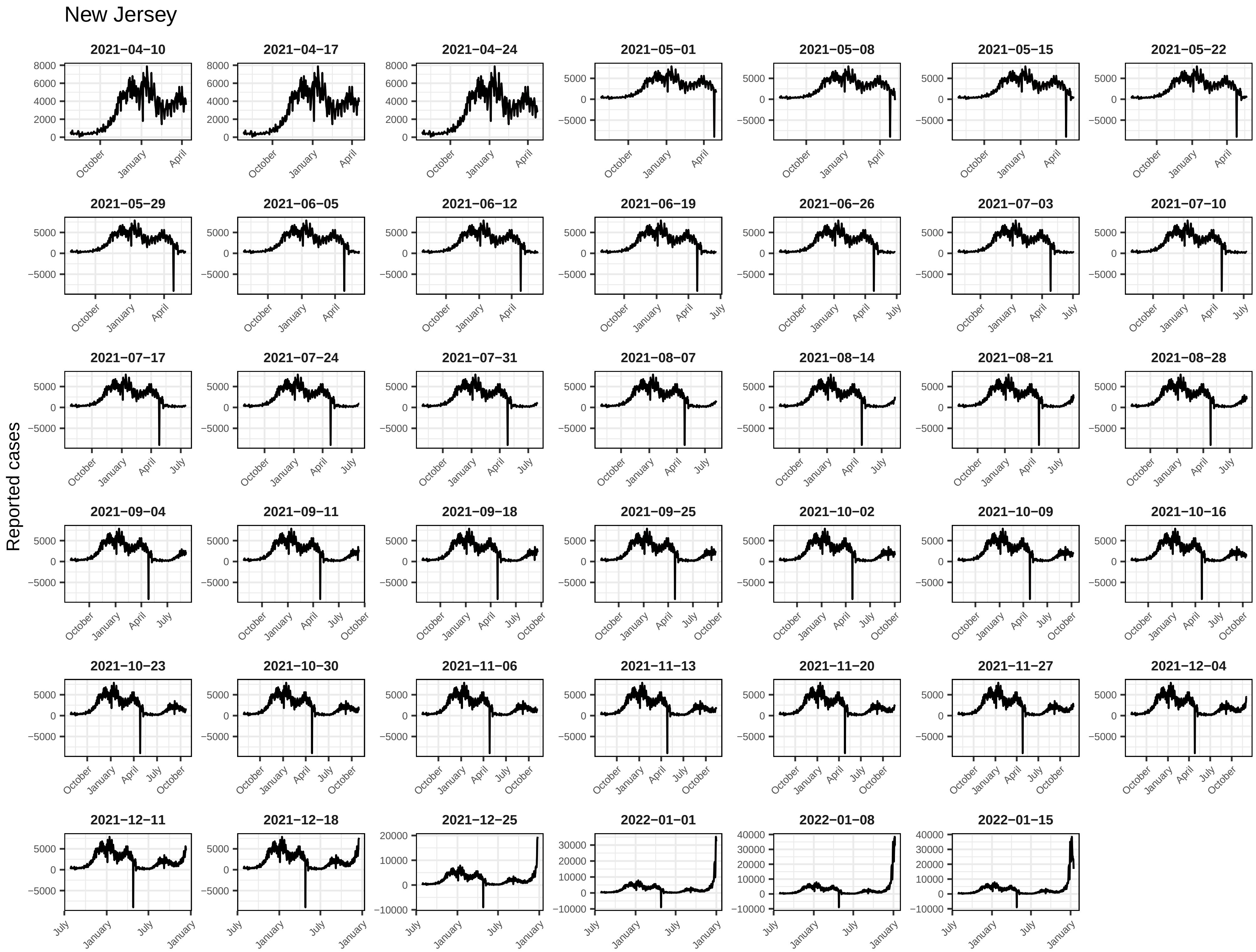
New Jersey
Reported cases
2021−04−10
2021−04−17
2021−04−24
2021−05−01
2021−05−08
2021−05−15
2021−05−22
2021−05−29
2021−06−05
2021−06−12
2021−06−19
2021−06−26
2021−07−03
2021−07−10
2021−07−17
2021−07−24
2021−07−31
2021−08−07
2021−08−14
2021−08−21
2021−08−28
2021−09−04
2021−09−11
2021−09−18
2021−09−25
2021−10−02
2021−10−09
2021−10−16
2021−10−23
2021−10−30
2021−11−06
2021−11−13
2021−11−20
2021−11−27
2021−12−04
2021−12−11
2021−12−18
2021−12−25
2022−01−01
2022−01−08
2022−01−15

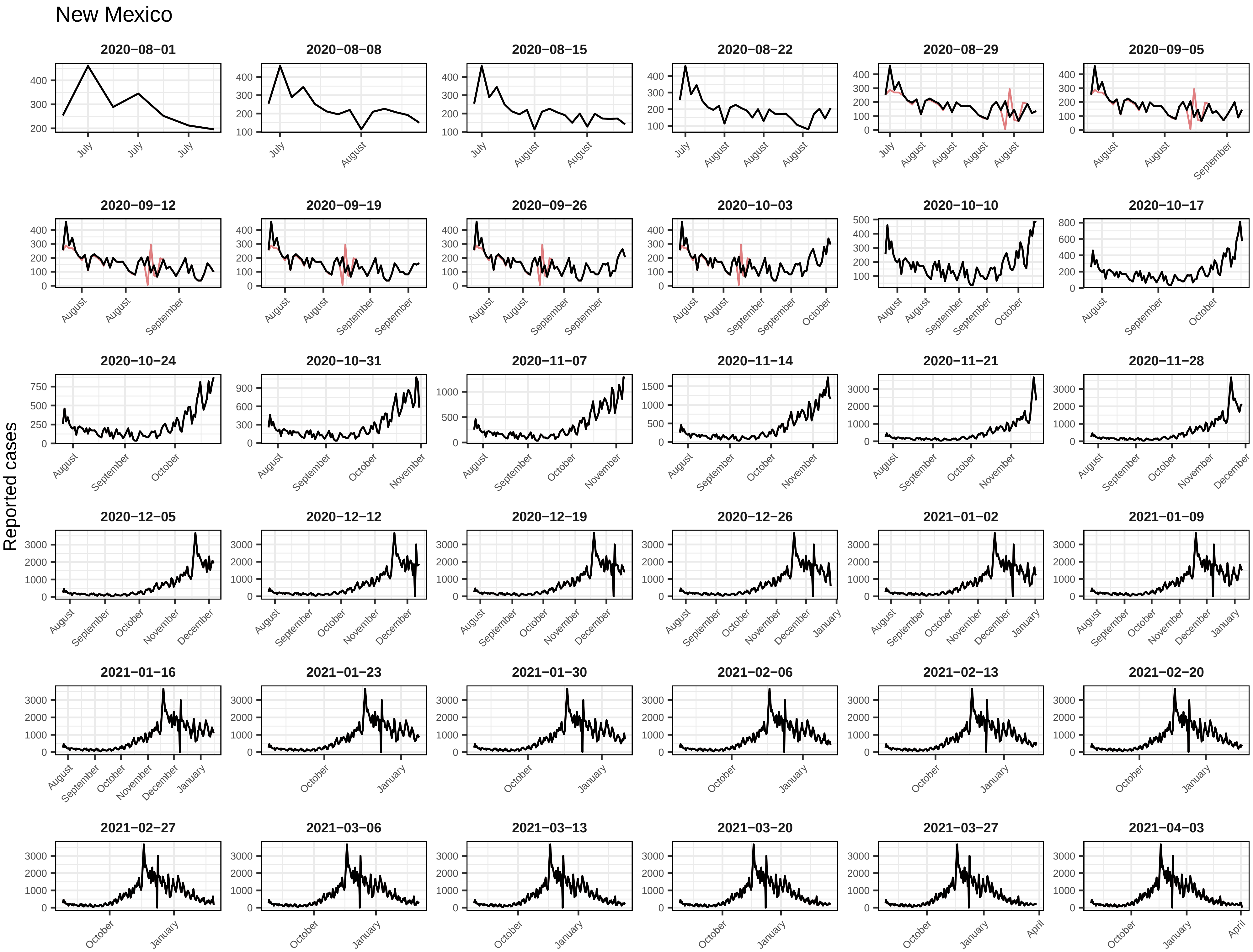
New Mexico
Reported cases
2020−08−01
2020−08−08
2020−08−15
2020−08−22
2020−08−29
2020−09−05
2020−09−12
2020−09−19
2020−09−26
2020−10−03
2020−10−10
2020−10−17
2020−10−24
2020−10−31
2020−11−07
2020−11−14
2020−11−21
2020−11−28
2020−12−05
2020−12−12
2020−12−19
2020−12−26
2021−01−02
2021−01−09
2021−01−16
2021−01−23
2021−01−30
2021−02−06
2021−02−13
2021−02−20
2021−02−27
2021−03−06
2021−03−13
2021−03−20
2021−03−27
2021−04−03

New Mexico

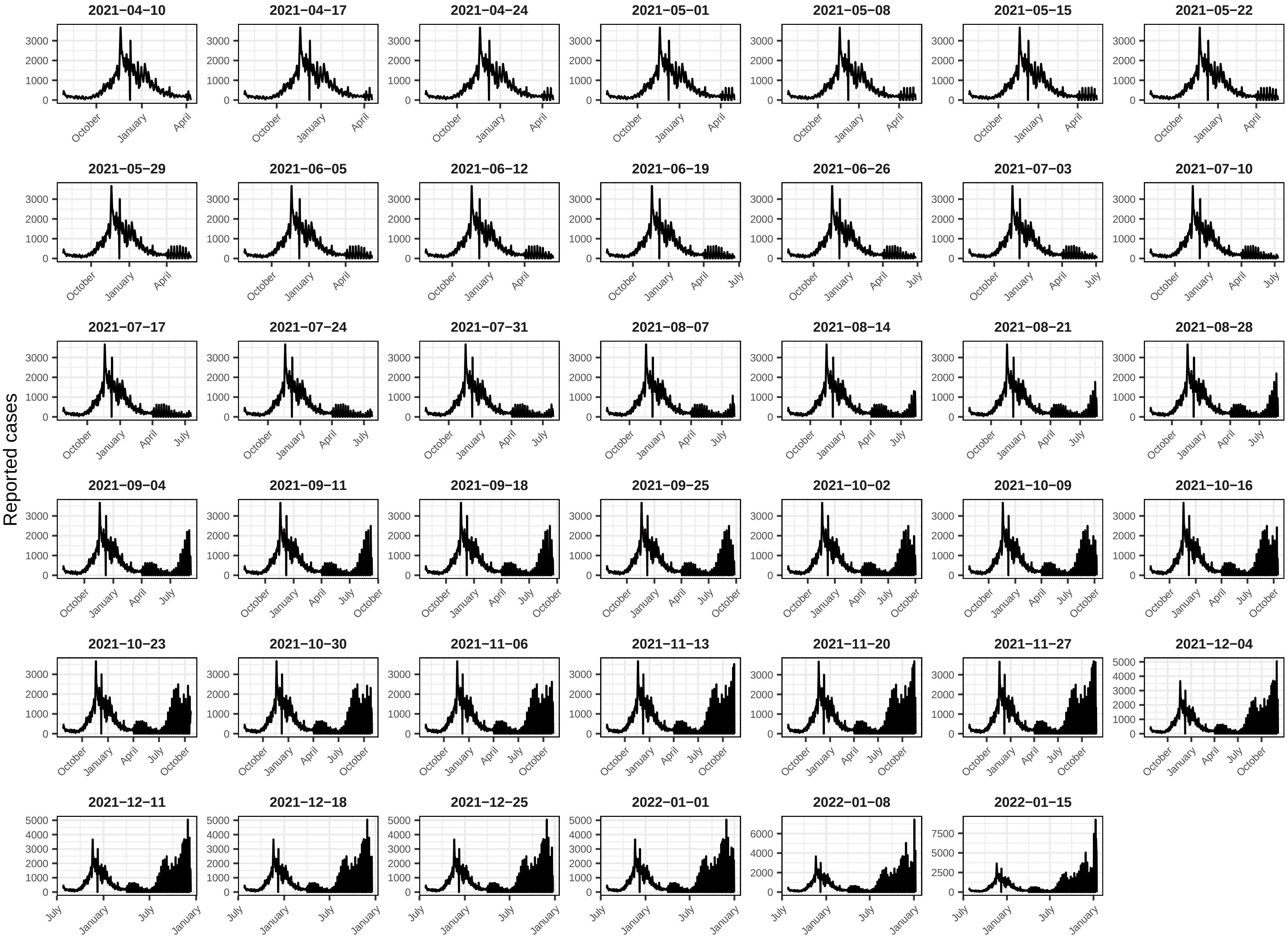

Reported cases
2021−04−10
2021−04−17
2021−04−24
2021−05−01
2021−05−08
2021−05−15
2021−05−22
2021−05−29
2021−06−05
2021−06−12
2021−06−19
2021−06−26
2021−07−03
2021−07−10
2021−07−17
2021−07−24
2021−07−31
2021−08−07
2021−08−14
2021−08−21
2021−08−28
2021−09−04
2021−09−11
2021−09−18
2021−09−25
2021−10−02
2021−10−09
2021−10−16
2021−10−23
2021−10−30
2021−11−06
2021−11−13
2021−11−20
2021−11−27
2021−12−04
2021−12−11
2021−12−18
2021−12−25
2022−01−01
2022−01−08
2022−01−15
October
January
April
July

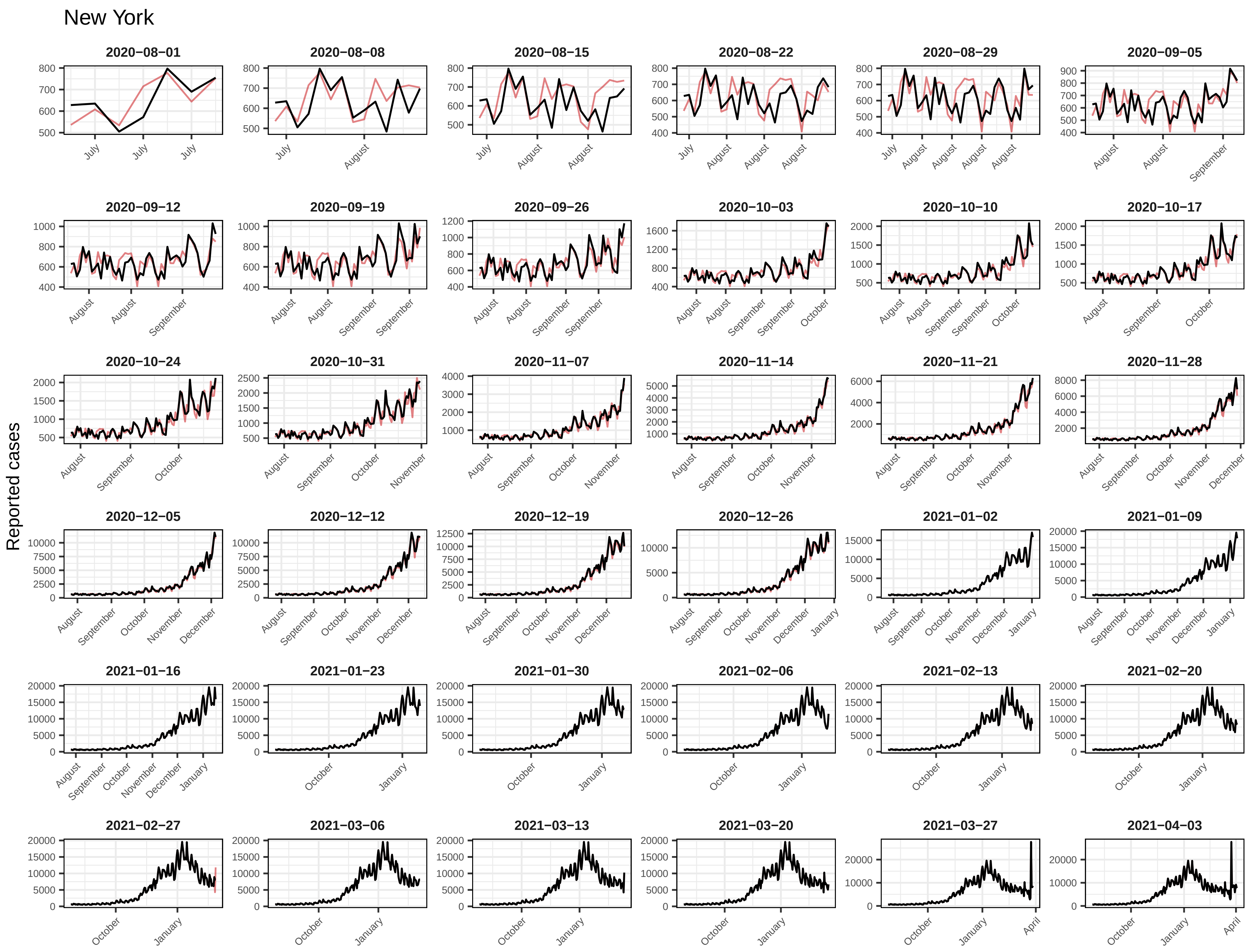
New York
Reported cases
2020−08−01
2020−08−08
2020−08−15
2020−08−22
2020−08−29
2020−09−05
2020−09−12
2020−09−19
2020−09−26
2020−10−03
2020−10−10
2020−10−17
2020−10−24
2020−10−31
2020−11−07
2020−11−14
2020−11−21
2020−11−28
2020−12−05
2020−12−12
2020−12−19
2020−12−26
2021−01−02
2021−01−09
2021−01−16
2021−01−23
2021−01−30
2021−02−06
2021−02−13
2021−02−20
2021−02−27
2021−03−06
2021−03−13
2021−03−20
2021−03−27
2021−04−03

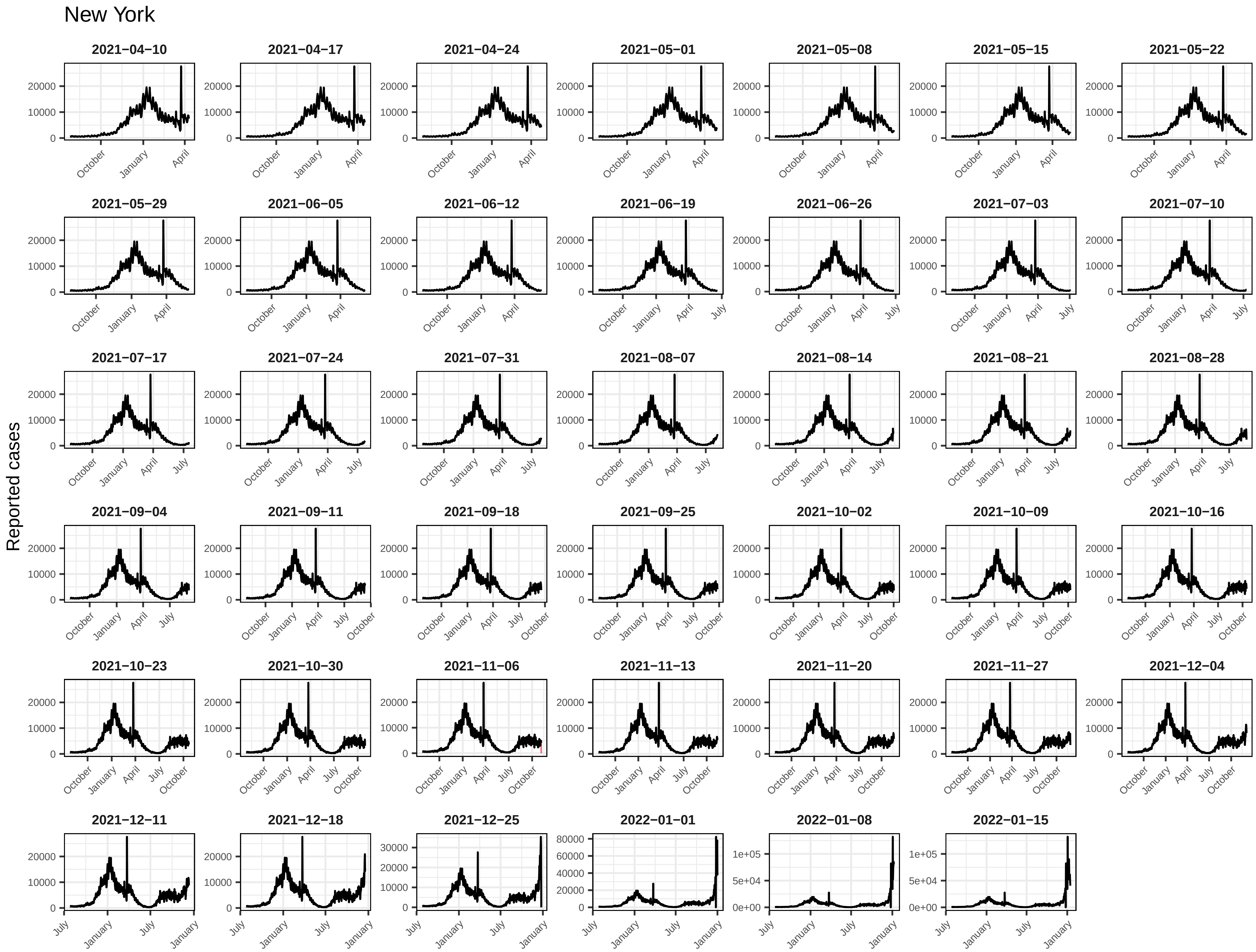
New York
Reported cases
2021−04−10
2021−04−17
2021−04−24
2021−05−01
2021−05−08
2021−05−15
2021−05−22
2021−05−29
2021−06−05
2021−06−12
2021−06−19
2021−06−26
2021−07−03
2021−07−10
2021−07−17
2021−07−24
2021−07−31
2021−08−07
2021−08−14
2021−08−21
2021−08−28
2021−09−04
2021−09−11
2021−09−18
2021−09−25
2021−10−02
2021−10−09
2021−10−16
2021−10−23
2021−10−30
2021−11−06
2021−11−13
2021−11−20
2021−11−27
2021−12−04
2021−12−11
2021−12−18
2021−12−25
2022−01−01
2022−01−08
2022−01−15
0
10000
20000
30000
40000
60000
80000
0e+00
5e+04
1e+05
July
October
January
April

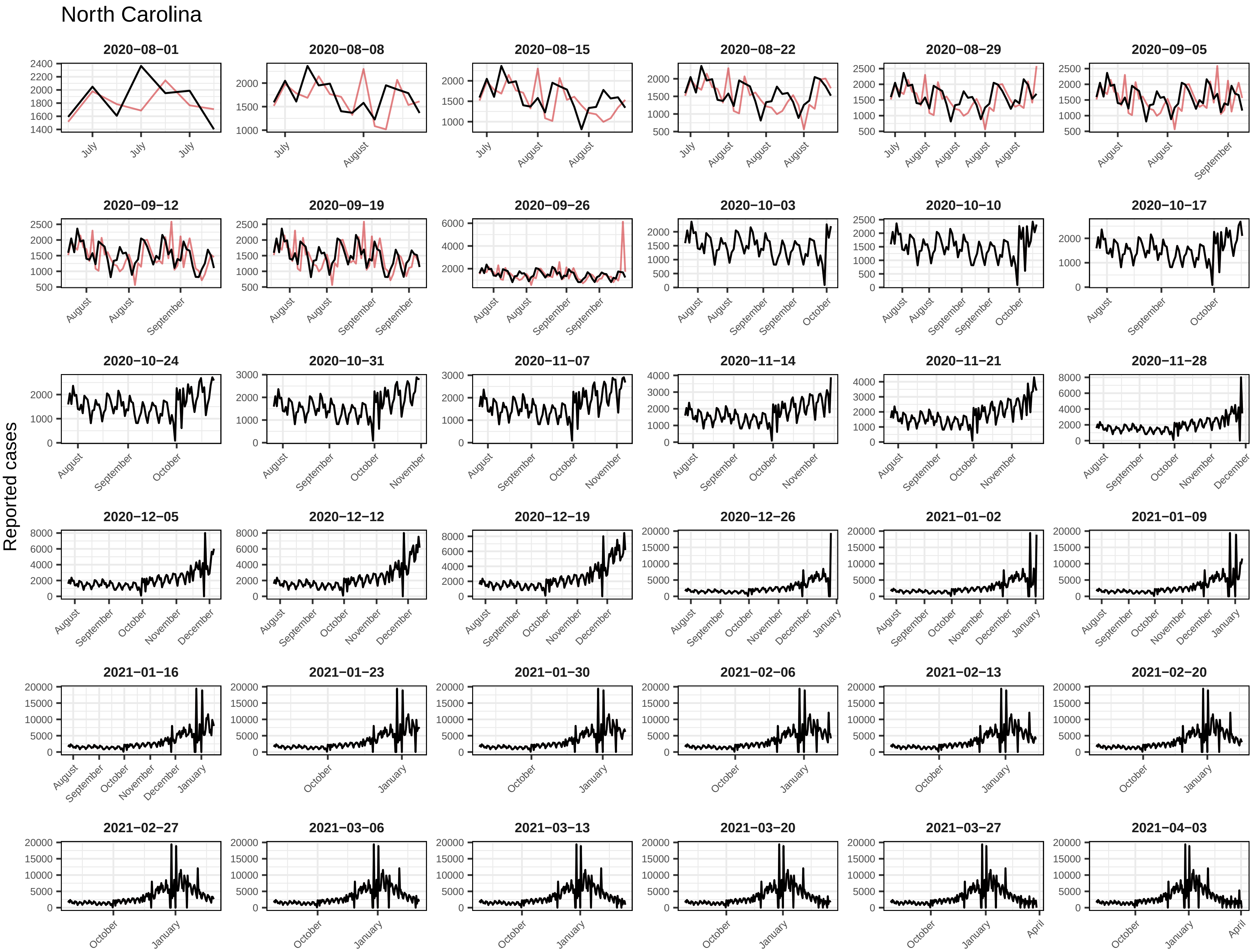
North Carolina
Reported cases
2020−08−01
2020−08−08
2020−08−15
2020−08−22
2020−08−29
2020−09−05
2020−09−12
2020−09−19
2020−09−26
2020−10−03
2020−10−10
2020−10−17
2020−10−24
2020−10−31
2020−11−07
2020−11−14
2020−11−21
2020−11−28
2020−12−05
2020−12−12
2020−12−19
2020−12−26
2021−01−02
2021−01−09
2021−01−16
2021−01−23
2021−01−30
2021−02−06
2021−02−13
2021−02−20
2021−02−27
2021−03−06
2021−03−13
2021−03−20
2021−03−27
2021−04−03

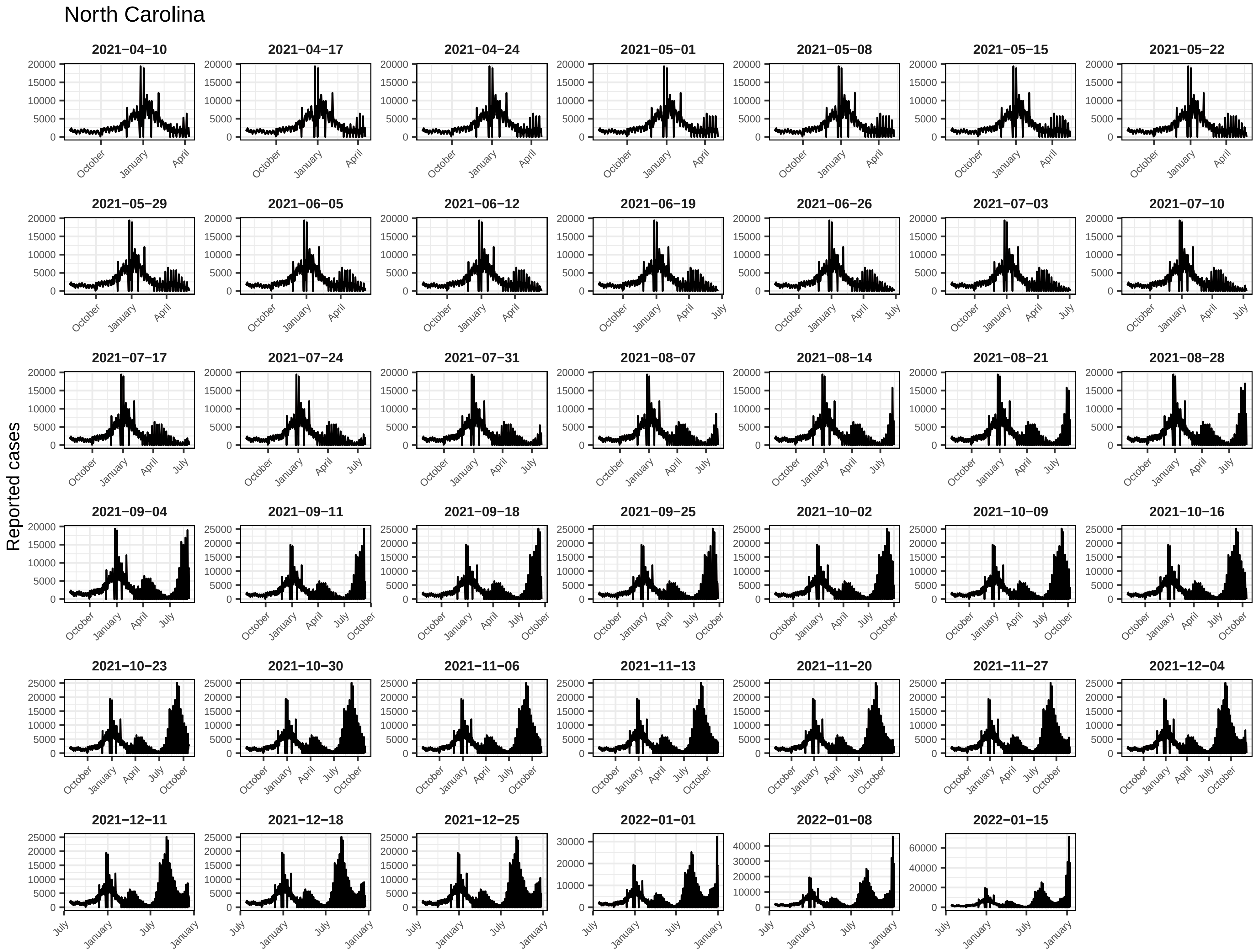
North Carolina
Reported cases
2021−04−10
2021−04−17
2021−04−24
2021−05−01
2021−05−08
2021−05−15
2021−05−22
2021−05−29
2021−06−05
2021−06−12
2021−06−19
2021−06−26
2021−07−03
2021−07−10
2021−07−17
2021−07−24
2021−07−31
2021−08−07
2021−08−14
2021−08−21
2021−08−28
2021−09−04
2021−09−11
2021−09−18
2021−09−25
2021−10−02
2021−10−09
2021−10−16
2021−10−23
2021−10−30
2021−11−06
2021−11−13
2021−11−20
2021−11−27
2021−12−04
2021−12−11
2021−12−18
2021−12−25
2022−01−01
2022−01−08
2022−01−15
October
January
April
July

### North Dakota

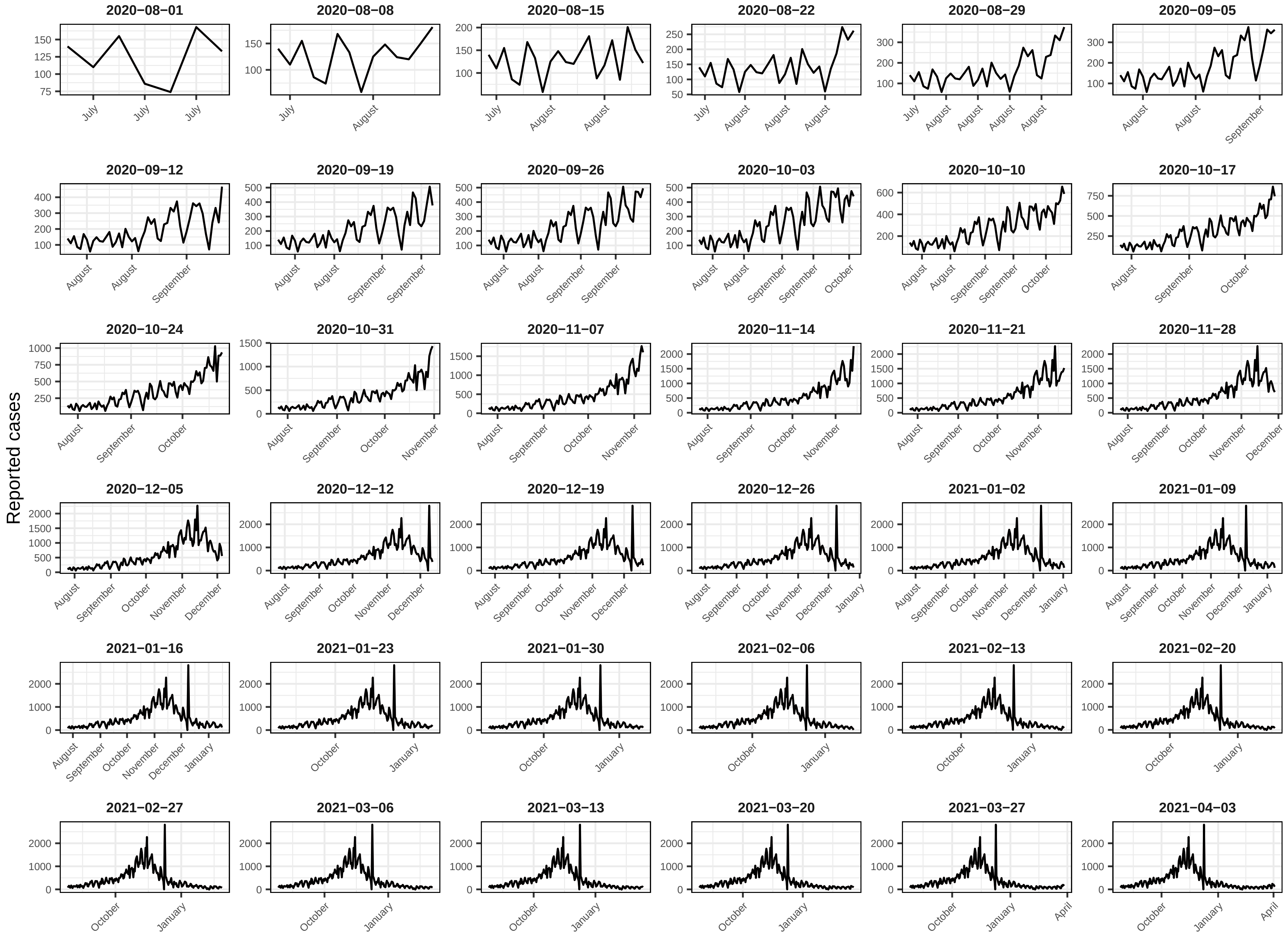

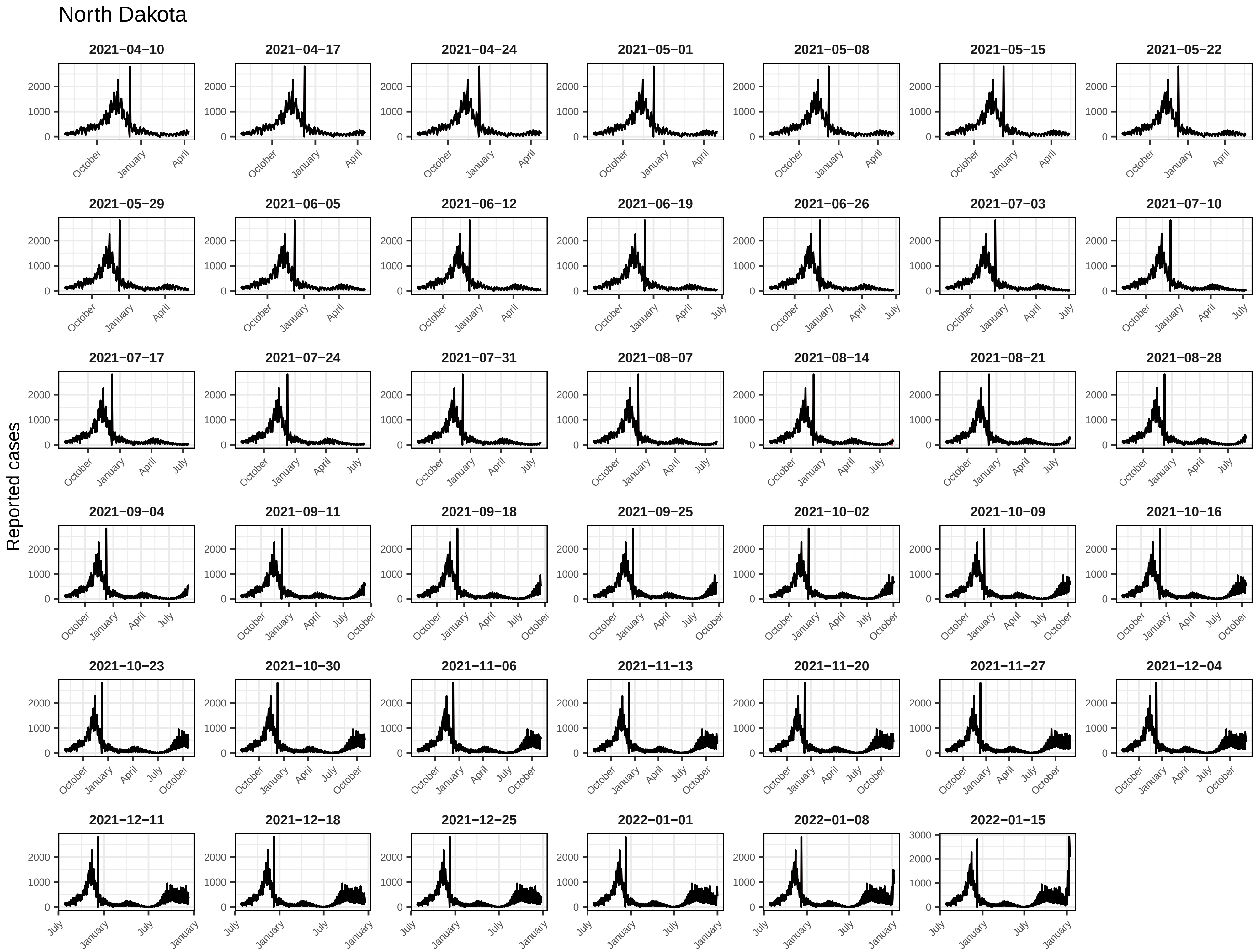
North Dakota
Reported cases
2021−04−10
2021−04−17
2021−04−24
2021−05−01
2021−05−08
2021−05−15
2021−05−22
2021−05−29
2021−06−05
2021−06−12
2021−06−19
2021−06−26
2021−07−03
2021−07−10
2021−07−17
2021−07−24
2021−07−31
2021−08−07
2021−08−14
2021−08−21
2021−08−28
2021−09−04
2021−09−11
2021−09−18
2021−09−25
2021−10−02
2021−10−09
2021−10−16
2021−10−23
2021−10−30
2021−11−06
2021−11−13
2021−11−20
2021−11−27
2021−12−04
2021−12−11
2021−12−18
2021−12−25
2022−01−01
2022−01−08
2022−01−15

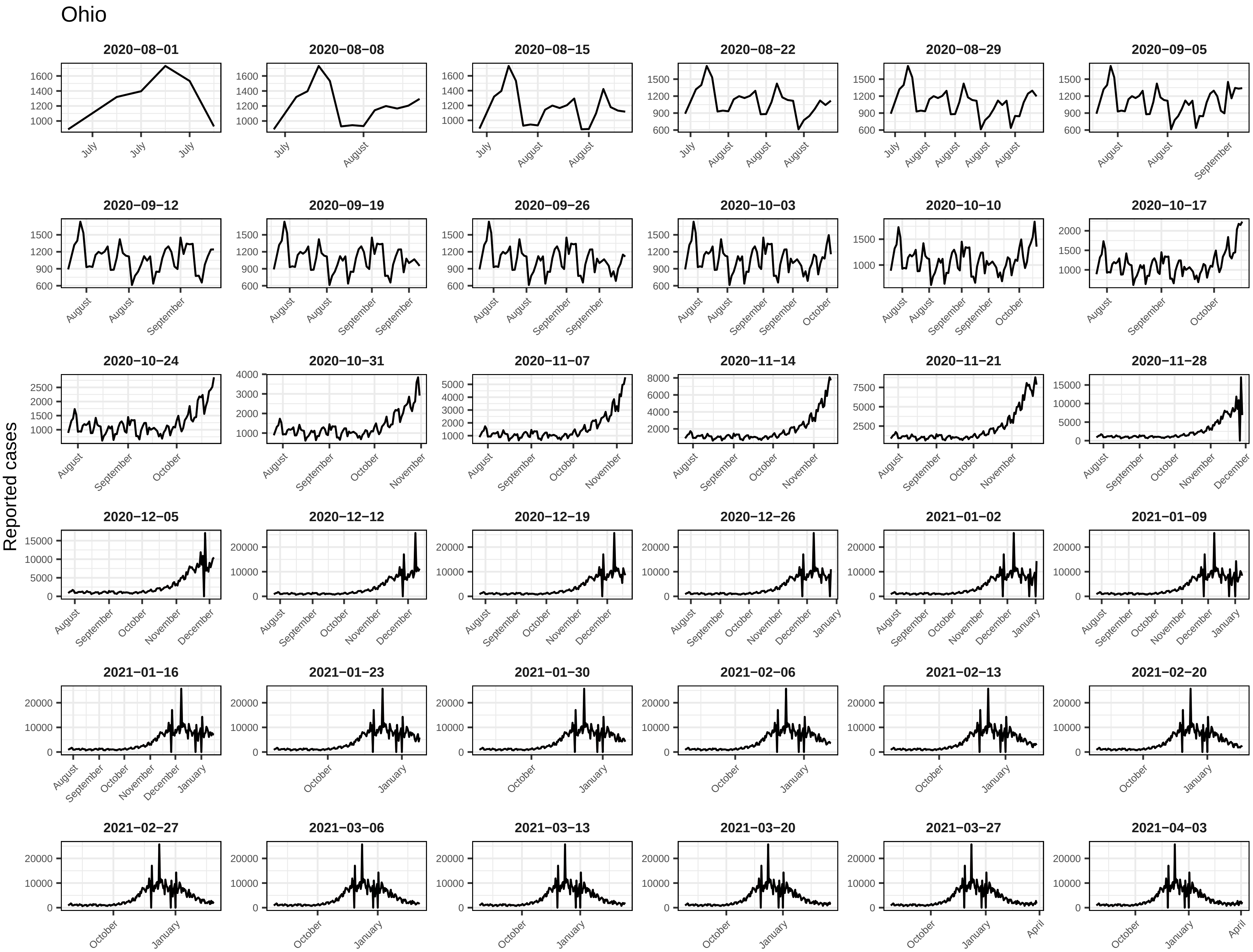

Ohio
Reported cases
2020−08−01
2020−08−08
2020−08−15
2020−08−22
2020−08−29
2020−09−05
2020−09−12
2020−09−19
2020−09−26
2020−10−03
2020−10−10
2020−10−17
2020−10−24
2020−10−31
2020−11−07
2020−11−14
2020−11−21
2020−11−28
2020−12−05
2020−12−12
2020−12−19
2020−12−26
2021−01−02
2021−01−09
2021−01−16
2021−01−23
2021−01−30
2021−02−06
2021−02−13
2021−02−20
2021−02−27
2021−03−06
2021−03−13
2021−03−20
2021−03−27
2021−04−03

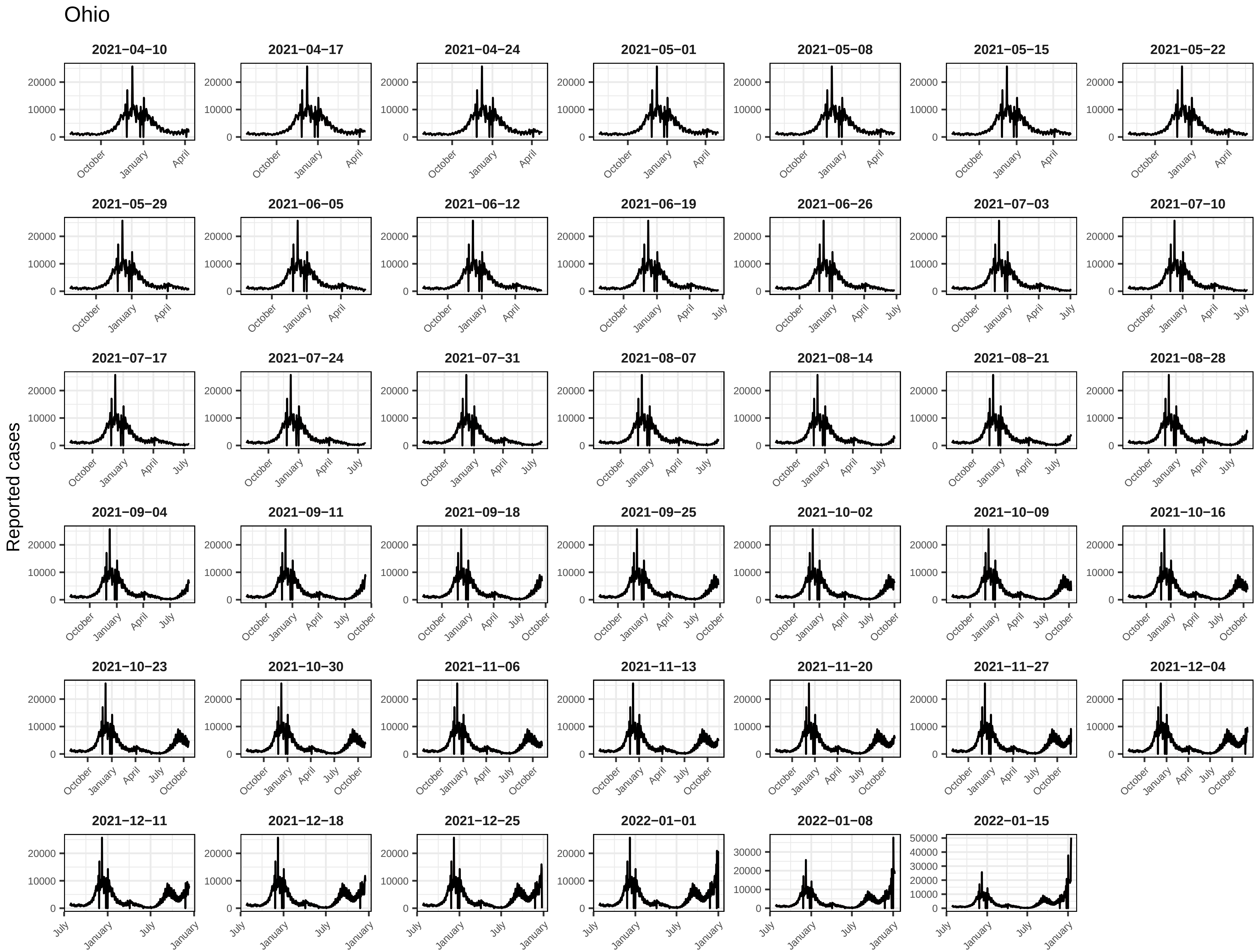
Ohio
2021−04−10
2021−04−17
2021−04−24
2021−05−01
2021−05−08
2021−05−15
2021−05−22
2021−05−29
2021−06−05
2021−06−12
2021−06−19
2021−06−26
2021−07−03
2021−07−10
2021−07−17
2021−07−24
2021−07−31
2021−08−07
2021−08−14
2021−08−21
2021−08−28
2021−09−04
2021−09−11
2021−09−18
2021−09−25
2021−10−02
2021−10−09
2021−10−16
2021−10−23
2021−10−30
2021−11−06
2021−11−13
2021−11−20
2021−11−27
2021−12−04
2021−12−11
2021−12−18
2021−12−25
2022−01−01
2022−01−08
2022−01−15
Reported cases

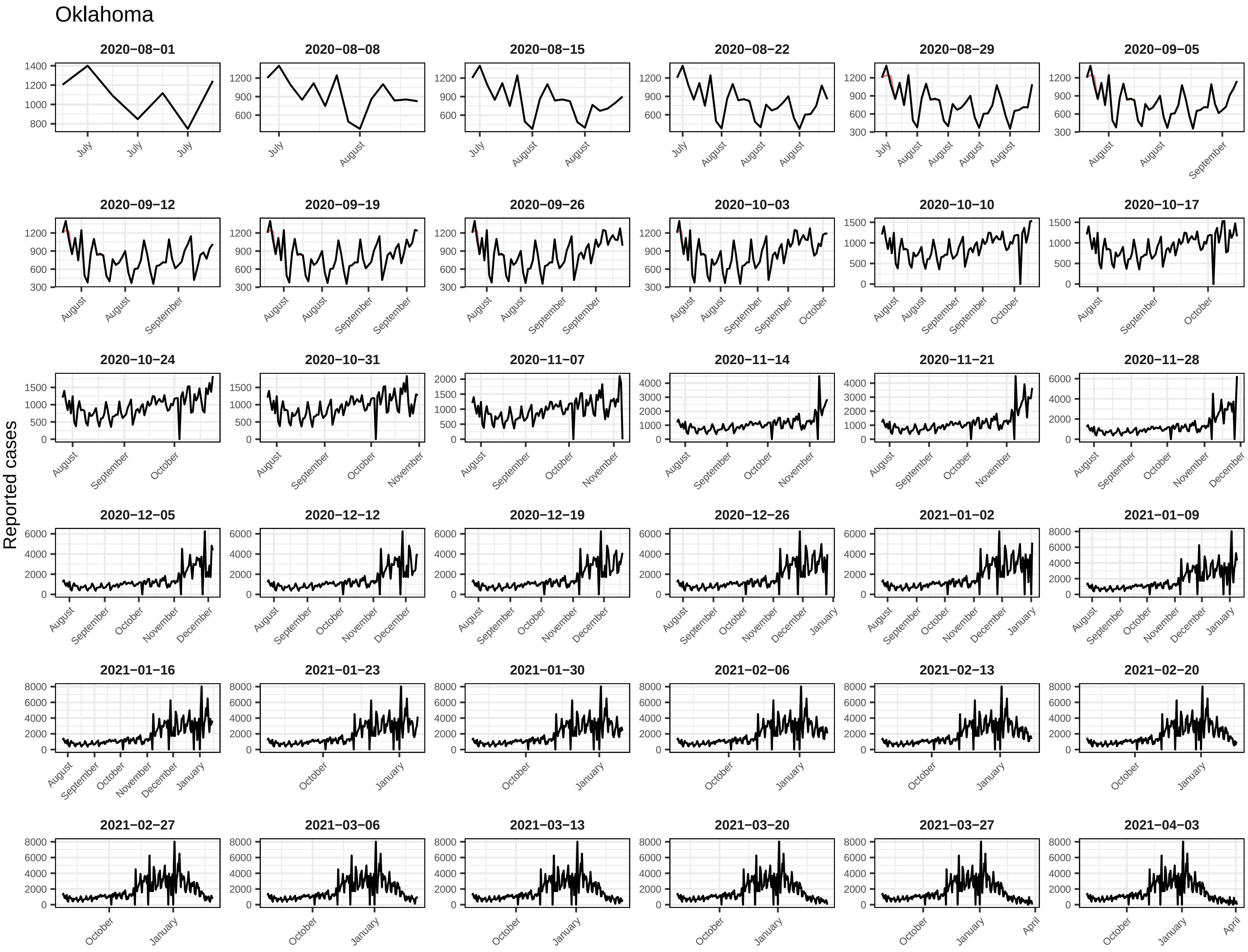
Oklahoma
Reported cases
2020−08−01
2020−08−08
2020−08−15
2020−08−22
2020−08−29
2020−09−05
2020−09−12
2020−09−19
2020−09−26
2020−10−03
2020−10−10
2020−10−17
2020−10−24
2020−10−31
2020−11−07
2020−11−14
2020−11−21
2020−11−28
2020−12−05
2020−12−12
2020−12−19
2020−12−26
2021−01−02
2021−01−09
2021−01−16
2021−01−23
2021−01−30
2021−02−06
2021−02−13
2021−02−20
2021−02−27
2021−03−06
2021−03−13
2021−03−20
2021−03−27
2021−04−03

### Oklahoma

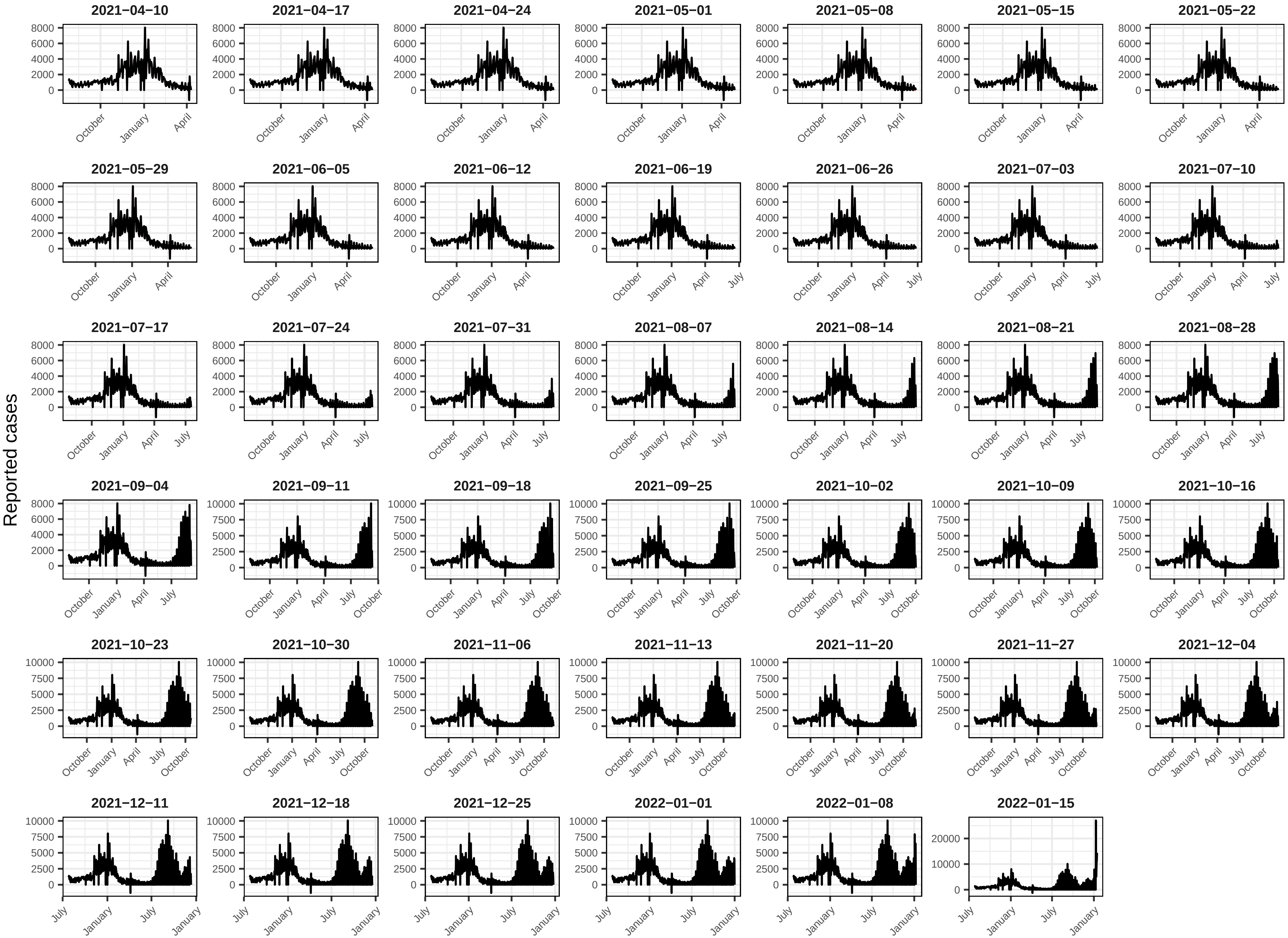
Reported cases
2021−04−10
2021−04−17
2021−04−24
2021−05−01
2021−05−08
2021−05−15
2021−05−22
2021−05−29
2021−06−05
2021−06−12
2021−06−19
2021−06−26
2021−07−03
2021−07−10
2021−07−17
2021−07−24
2021−07−31
2021−08−07
2021−08−14
2021−08−21
2021−08−28
2021−09−04
2021−09−11
2021−09−18
2021−09−25
2021−10−02
2021−10−09
2021−10−16
2021−10−23
2021−10−30
2021−11−06
2021−11−13
2021−11−20
2021−11−27
2021−12−04
2021−12−11
2021−12−18
2021−12−25
2022−01−01
2022−01−08
2022−01−15
0
2000
4000
6000
8000
2500
5000
7500
10000
20000
July
October
January
April

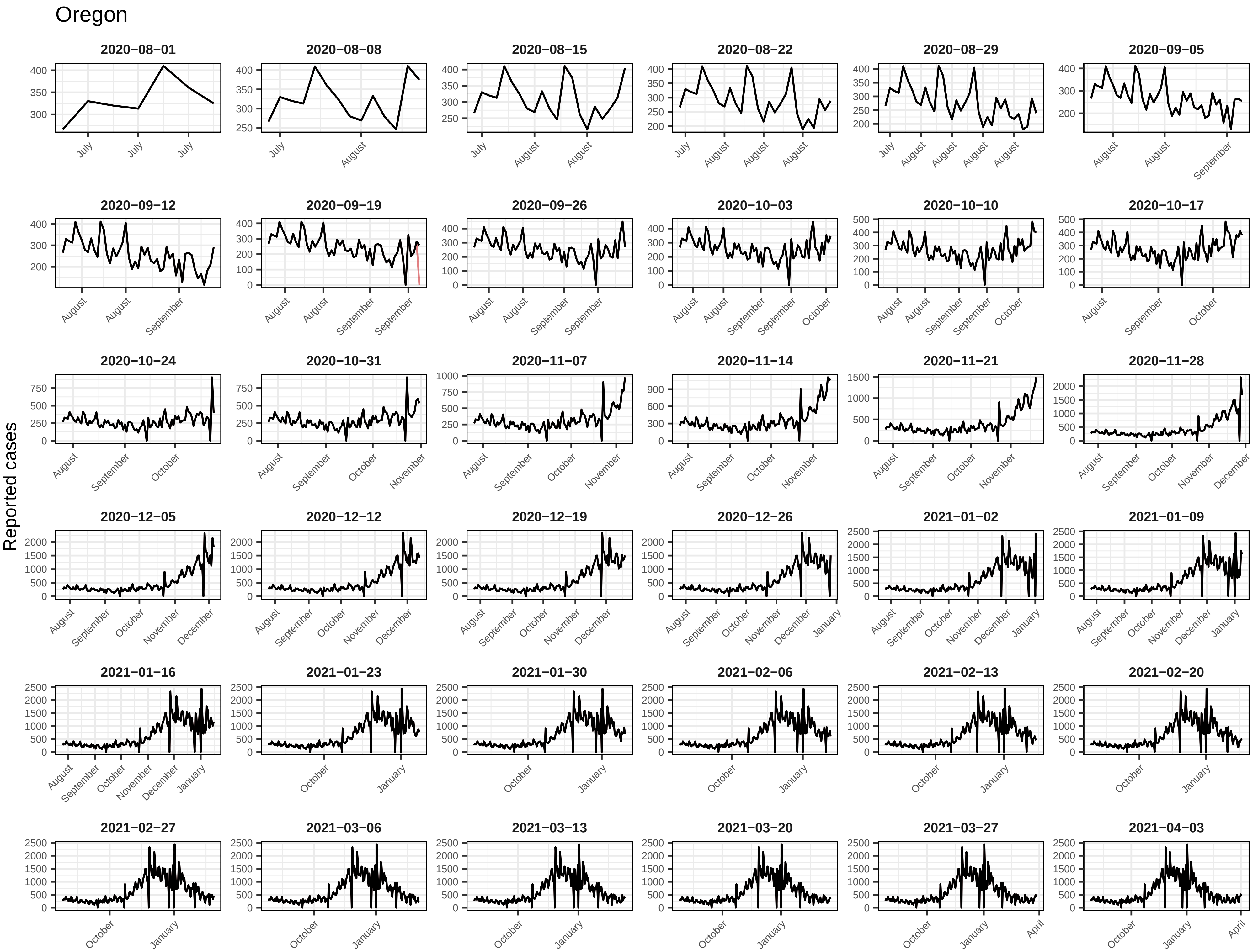
Oregon
Reported cases
2020−08−01
2020−08−08
2020−08−15
2020−08−22
2020−08−29
2020−09−05
2020−09−12
2020−09−19
2020−09−26
2020−10−03
2020−10−10
2020−10−17
2020−10−24
2020−10−31
2020−11−07
2020−11−14
2020−11−21
2020−11−28
2020−12−05
2020−12−12
2020−12−19
2020−12−26
2021−01−02
2021−01−09
2021−01−16
2021−01−23
2021−01−30
2021−02−06
2021−02−13
2021−02−20
2021−02−27
2021−03−06
2021−03−13
2021−03−20
2021−03−27
2021−04−03

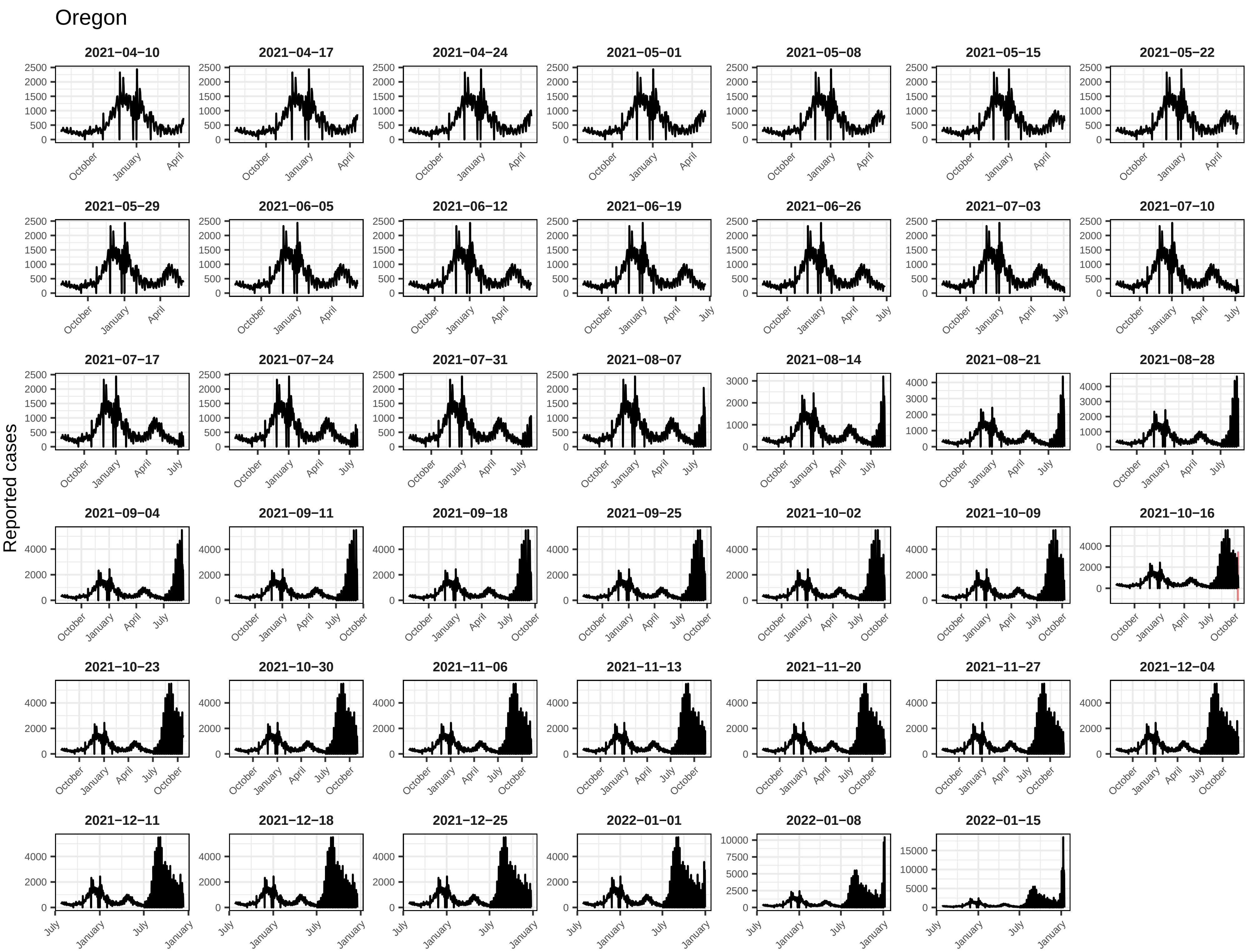
Oregon
Reported cases
2021−04−10
2021−04−17
2021−04−24
2021−05−01
2021−05−08
2021−05−15
2021−05−22
2021−05−29
2021−06−05
2021−06−12
2021−06−19
2021−06−26
2021−07−03
2021−07−10
2021−07−17
2021−07−24
2021−07−31
2021−08−07
2021−08−14
2021−08−21
2021−08−28
2021−09−04
2021−09−11
2021−09−18
2021−09−25
2021−10−02
2021−10−09
2021−10−16
2021−10−23
2021−10−30
2021−11−06
2021−11−13
2021−11−20
2021−11−27
2021−12−04
2021−12−11
2021−12−18
2021−12−25
2022−01−01
2022−01−08
2022−01−15

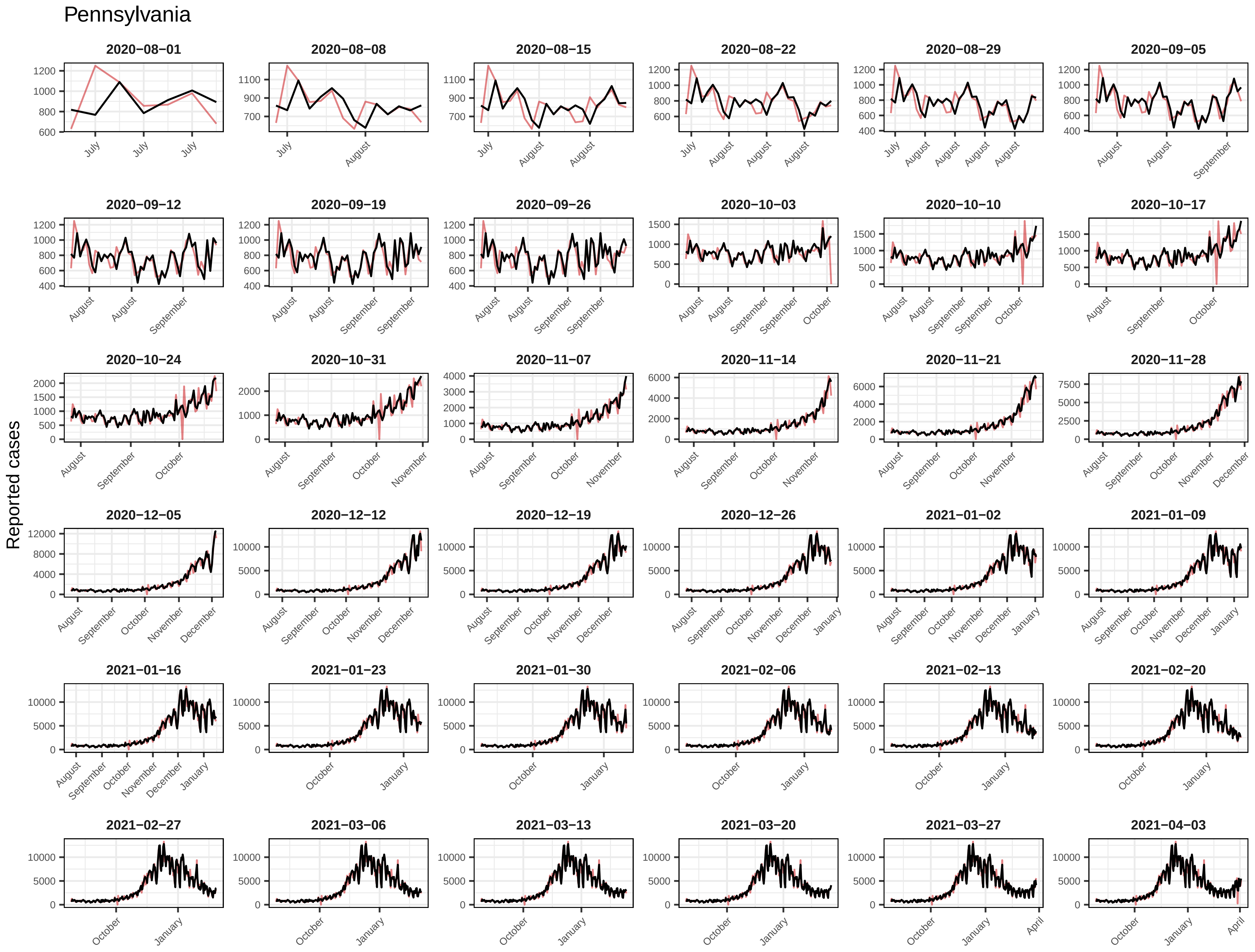

Pennsylvania
Reported cases
2020−08−01
2020−08−08
2020−08−15
2020−08−22
2020−08−29
2020−09−05
2020−09−12
2020−09−19
2020−09−26
2020−10−03
2020−10−10
2020−10−17
2020−10−24
2020−10−31
2020−11−07
2020−11−14
2020−11−21
2020−11−28
2020−12−05
2020−12−12
2020−12−19
2020−12−26
2021−01−02
2021−01−09
2021−01−16
2021−01−23
2021−01−30
2021−02−06
2021−02−13
2021−02−20
2021−02−27
2021−03−06
2021−03−13
2021−03−20
2021−03−27
2021−04−03

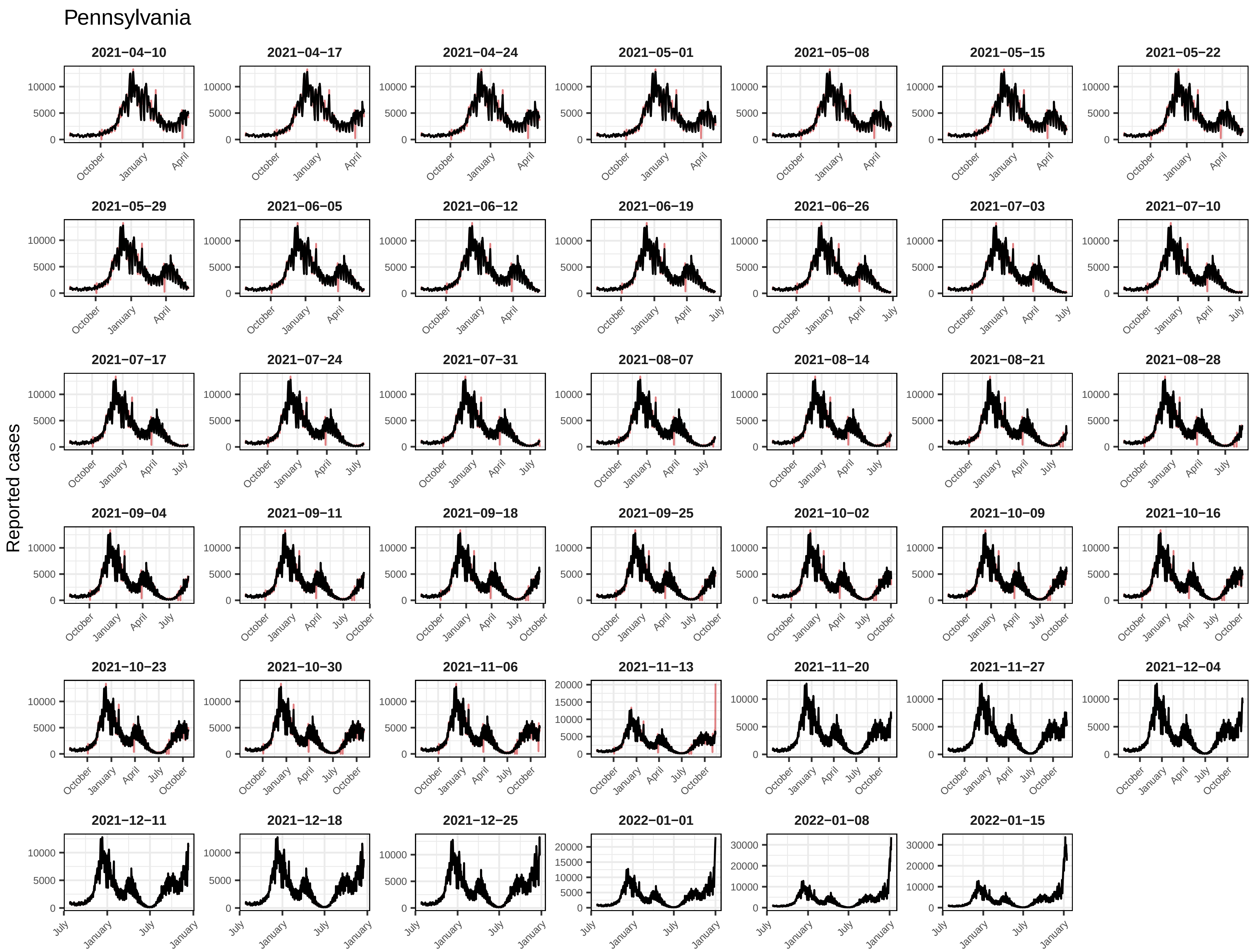
Pennsylvania
Reported cases
2021−04−10
2021−04−17
2021−04−24
2021−05−01
2021−05−08
2021−05−15
2021−05−22
2021−05−29
2021−06−05
2021−06−12
2021−06−19
2021−06−26
2021−07−03
2021−07−10
2021−07−17
2021−07−24
2021−07−31
2021−08−07
2021−08−14
2021−08−21
2021−08−28
2021−09−04
2021−09−11
2021−09−18
2021−09−25
2021−10−02
2021−10−09
2021−10−16
2021−10−23
2021−10−30
2021−11−06
2021−11−13
2021−11−20
2021−11−27
2021−12−04
2021−12−11
2021−12−18
2021−12−25
2022−01−01
2022−01−08
2022−01−15
October
January
April
July

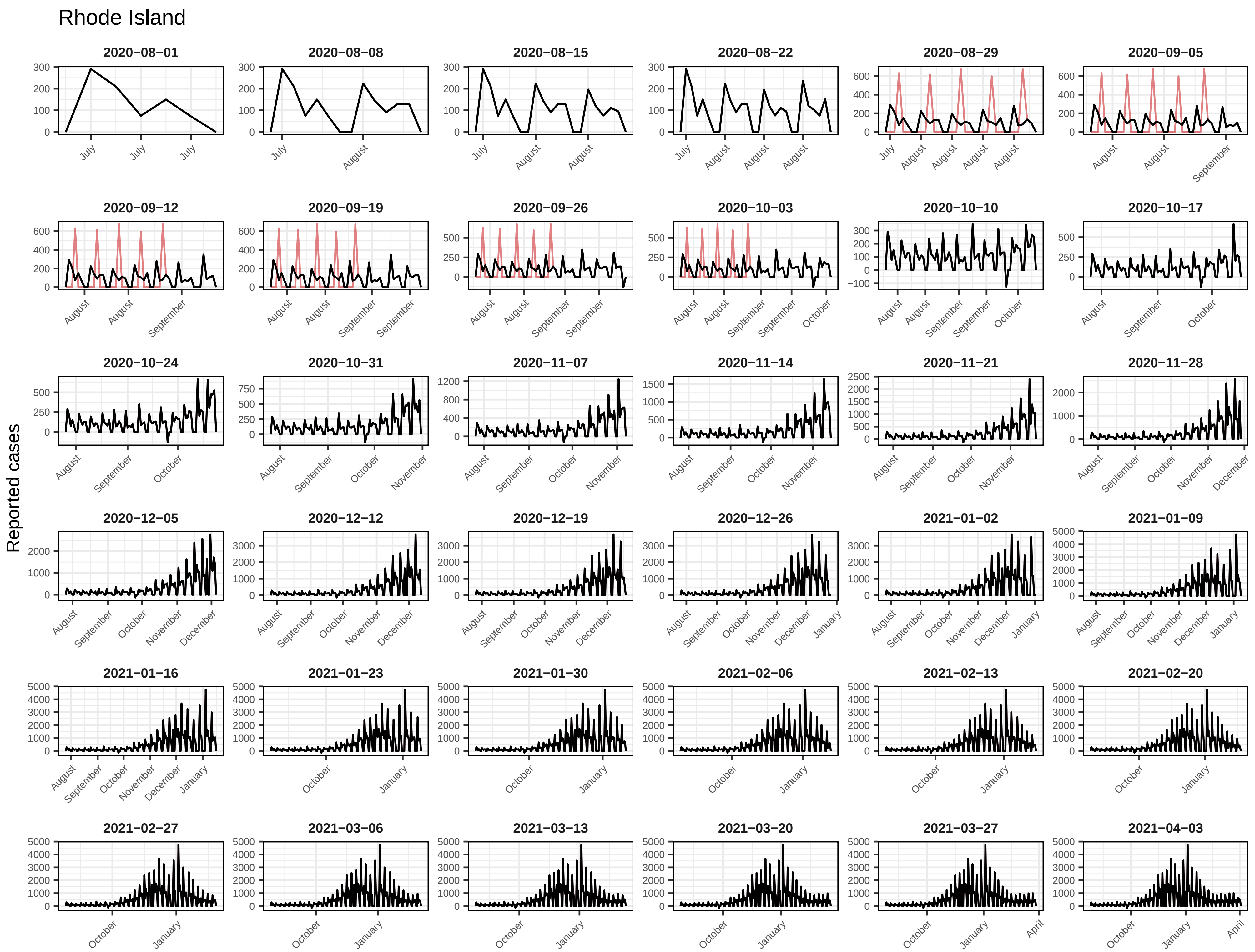

Rhode Island
Reported cases
2020−08−01
2020−08−08
2020−08−15
2020−08−22
2020−08−29
2020−09−05
2020−09−12
2020−09−19
2020−09−26
2020−10−03
2020−10−10
2020−10−17
2020−10−24
2020−10−31
2020−11−07
2020−11−14
2020−11−21
2020−11−28
2020−12−05
2020−12−12
2020−12−19
2020−12−26
2021−01−02
2021−01−09
2021−01−16
2021−01−23
2021−01−30
2021−02−06
2021−02−13
2021−02−20
2021−02−27
2021−03−06
2021−03−13
2021−03−20
2021−03−27
2021−04−03

#### Rhode Island

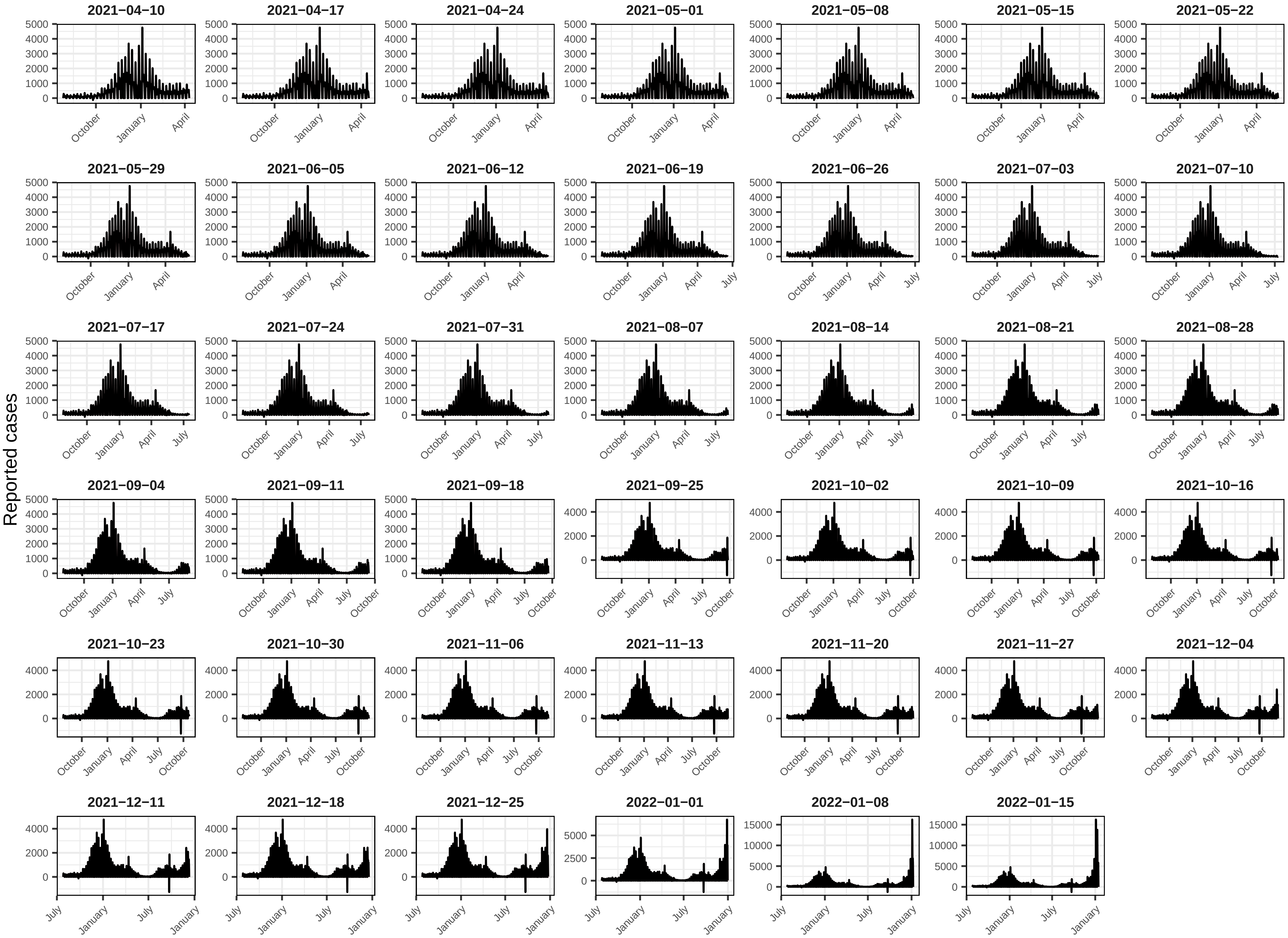

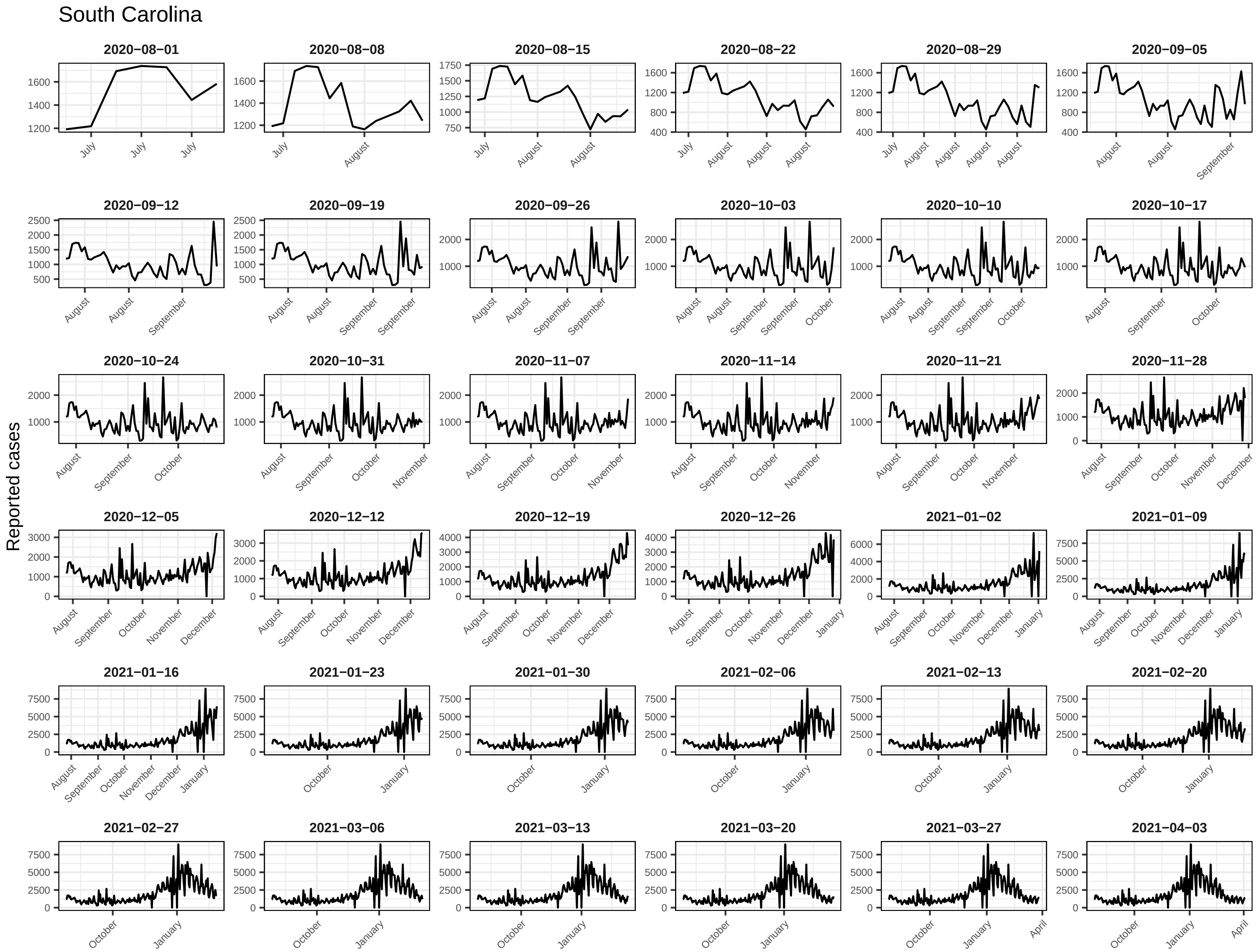
South Carolina
Reported cases
2020−08−01
2020−08−08
2020−08−15
2020−08−22
2020−08−29
2020−09−05
2020−09−12
2020−09−19
2020−09−26
2020−10−03
2020−10−10
2020−10−17
2020−10−24
2020−10−31
2020−11−07
2020−11−14
2020−11−21
2020−11−28
2020−12−05
2020−12−12
2020−12−19
2020−12−26
2021−01−02
2021−01−09
2021−01−16
2021−01−23
2021−01−30
2021−02−06
2021−02−13
2021−02−20
2021−02−27
2021−03−06
2021−03−13
2021−03−20
2021−03−27
2021−04−03

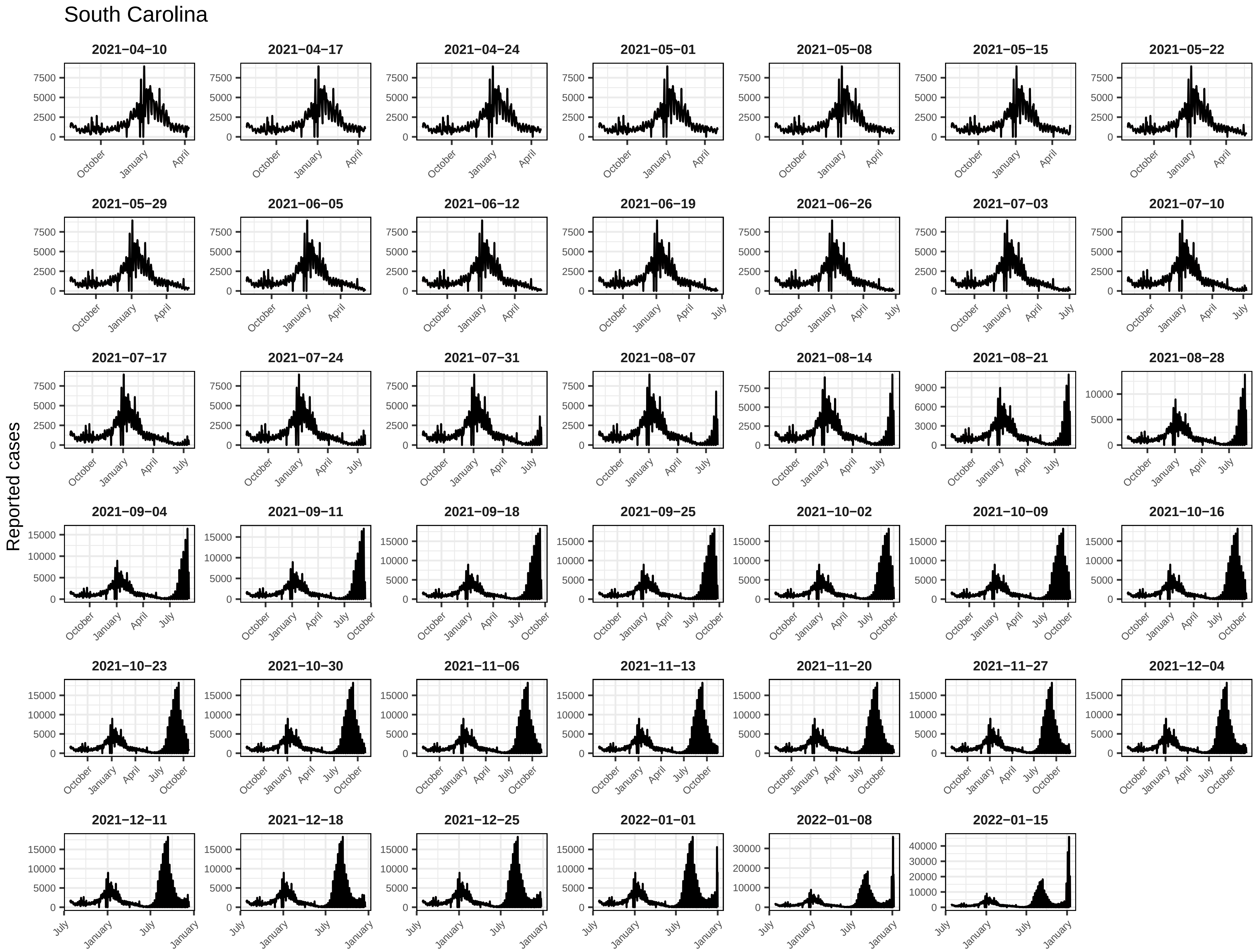
South Carolina
Reported cases
2021−04−10
2021−04−17
2021−04−24
2021−05−01
2021−05−08
2021−05−15
2021−05−22
2021−05−29
2021−06−05
2021−06−12
2021−06−19
2021−06−26
2021−07−03
2021−07−10
2021−07−17
2021−07−24
2021−07−31
2021−08−07
2021−08−14
2021−08−21
2021−08−28
2021−09−04
2021−09−11
2021−09−18
2021−09−25
2021−10−02
2021−10−09
2021−10−16
2021−10−23
2021−10−30
2021−11−06
2021−11−13
2021−11−20
2021−11−27
2021−12−04
2021−12−11
2021−12−18
2021−12−25
2022−01−01
2022−01−08
2022−01−15
October
January
April
July

### South Dakota

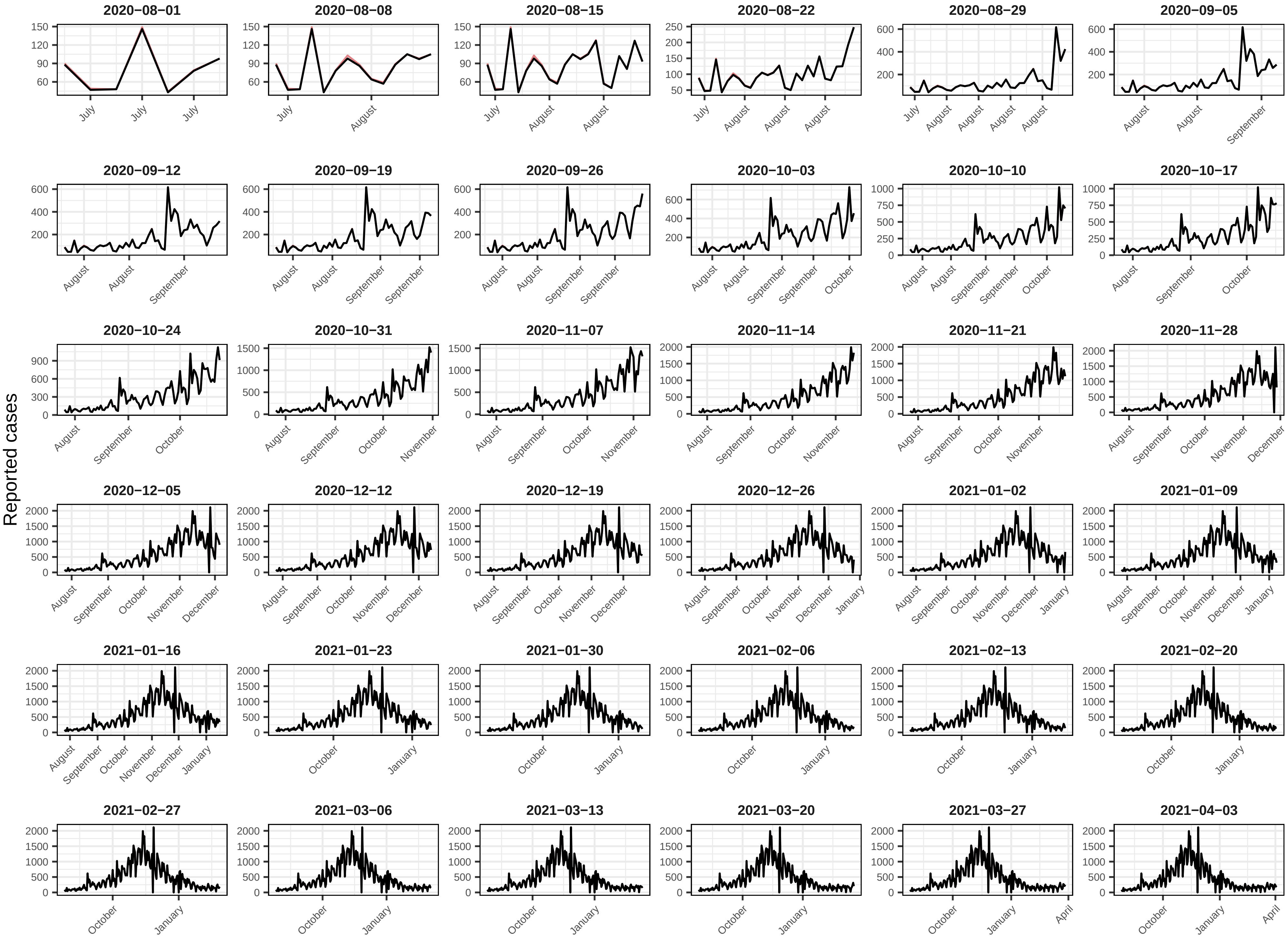

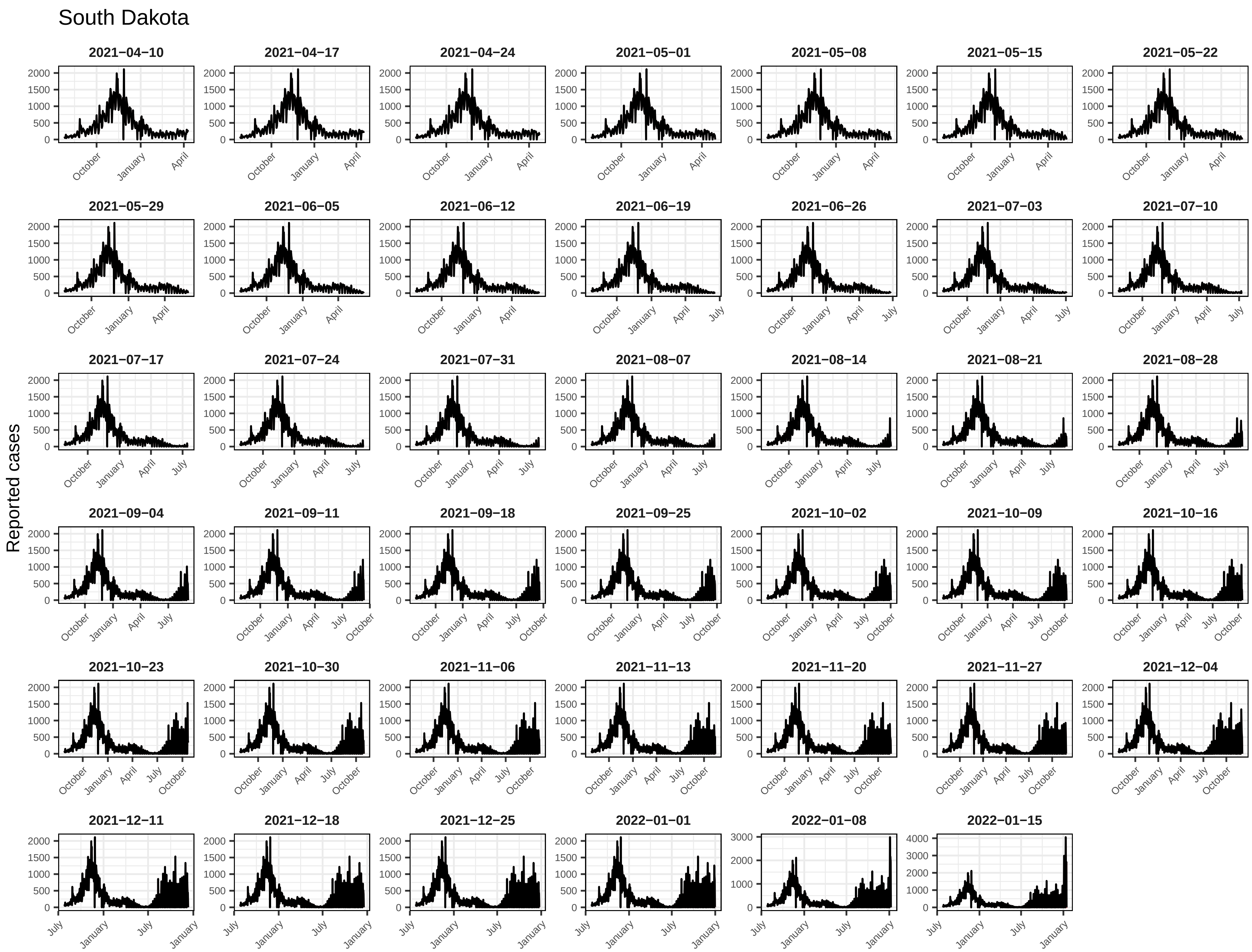
South Dakota
Reported cases
2021−04−10
2021−04−17
2021−04−24
2021−05−01
2021−05−08
2021−05−15
2021−05−22
2021−05−29
2021−06−05
2021−06−12
2021−06−19
2021−06−26
2021−07−03
2021−07−10
2021−07−17
2021−07−24
2021−07−31
2021−08−07
2021−08−14
2021−08−21
2021−08−28
2021−09−04
2021−09−11
2021−09−18
2021−09−25
2021−10−02
2021−10−09
2021−10−16
2021−10−23
2021−10−30
2021−11−06
2021−11−13
2021−11−20
2021−11−27
2021−12−04
2021−12−11
2021−12−18
2021−12−25
2022−01−01
2022−01−08
2022−01−15

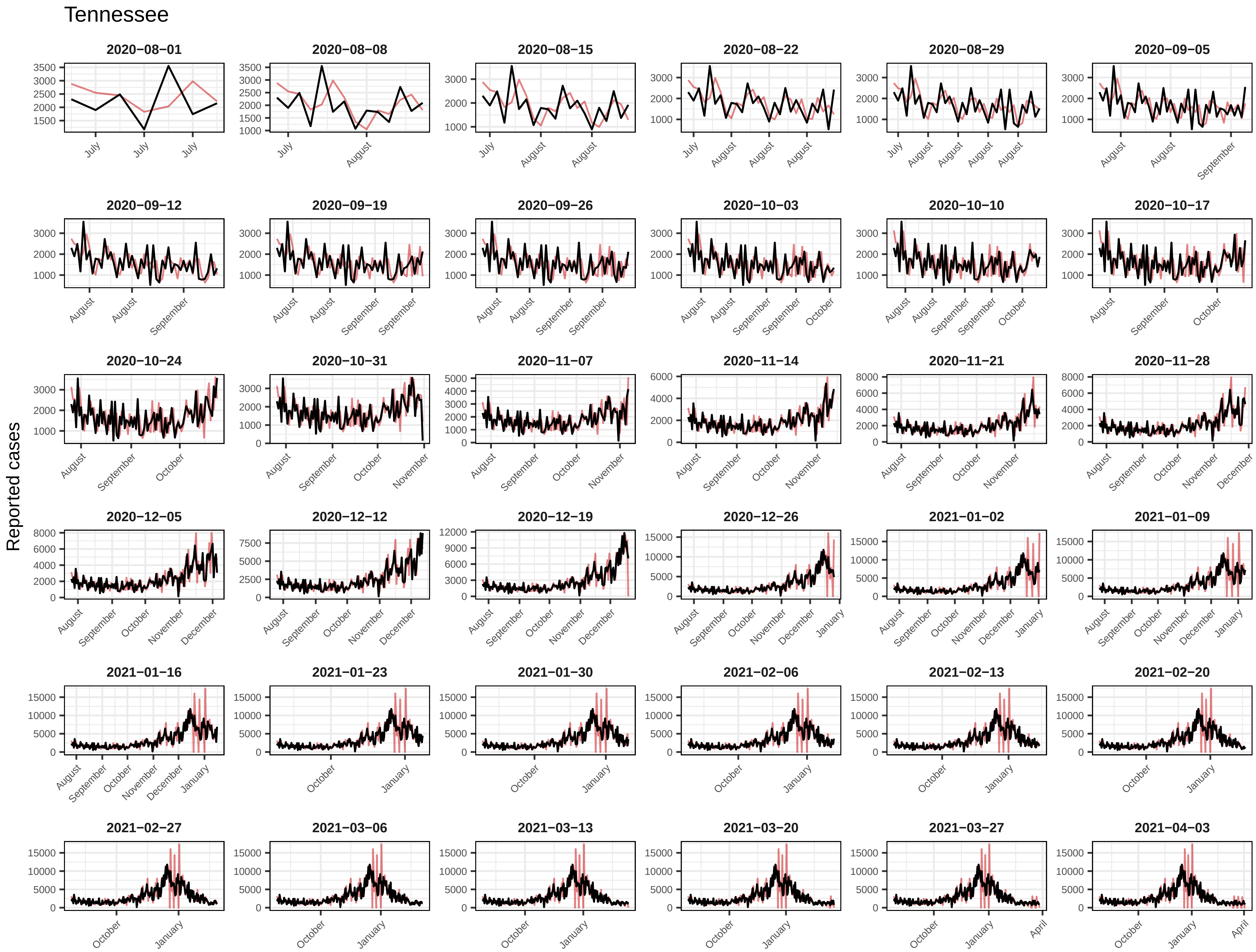
Tennessee
Reported cases
2020−08−01
2020−08−08
2020−08−15
2020−08−22
2020−08−29
2020−09−05
2020−09−12
2020−09−19
2020−09−26
2020−10−03
2020−10−10
2020−10−17
2020−10−24
2020−10−31
2020−11−07
2020−11−14
2020−11−21
2020−11−28
2020−12−05
2020−12−12
2020−12−19
2020−12−26
2021−01−02
2021−01−09
2021−01−16
2021−01−23
2021−01−30
2021−02−06
2021−02−13
2021−02−20
2021−02−27
2021−03−06
2021−03−13
2021−03−20
2021−03−27
2021−04−03

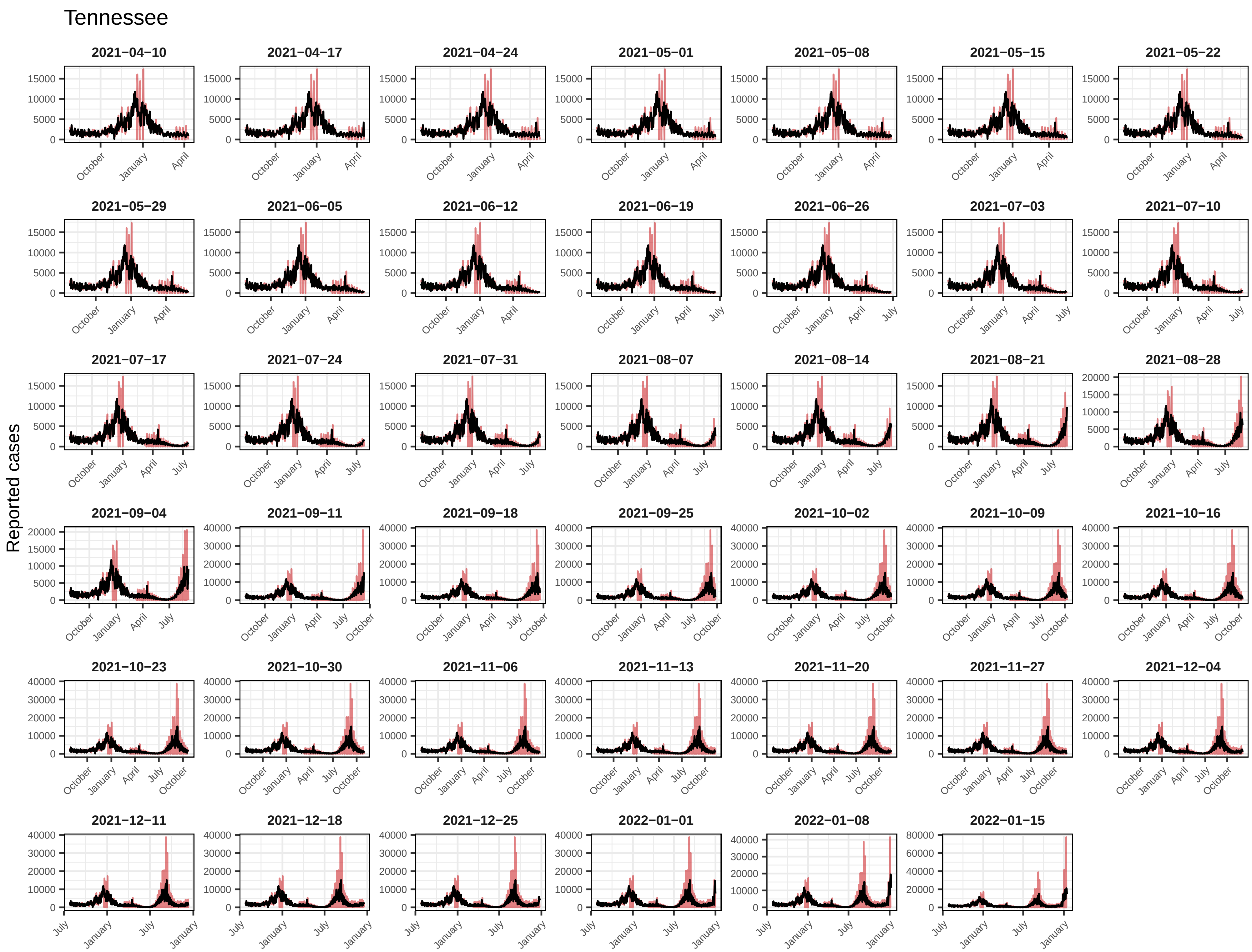
Tennessee
Reported cases
2021−04−10
2021−04−17
2021−04−24
2021−05−01
2021−05−08
2021−05−15
2021−05−22
2021−05−29
2021−06−05
2021−06−12
2021−06−19
2021−06−26
2021−07−03
2021−07−10
2021−07−17
2021−07−24
2021−07−31
2021−08−07
2021−08−14
2021−08−21
2021−08−28
2021−09−04
2021−09−11
2021−09−18
2021−09−25
2021−10−02
2021−10−09
2021−10−16
2021−10−23
2021−10−30
2021−11−06
2021−11−13
2021−11−20
2021−11−27
2021−12−04
2021−12−11
2021−12−18
2021−12−25
2022−01−01
2022−01−08
2022−01−15

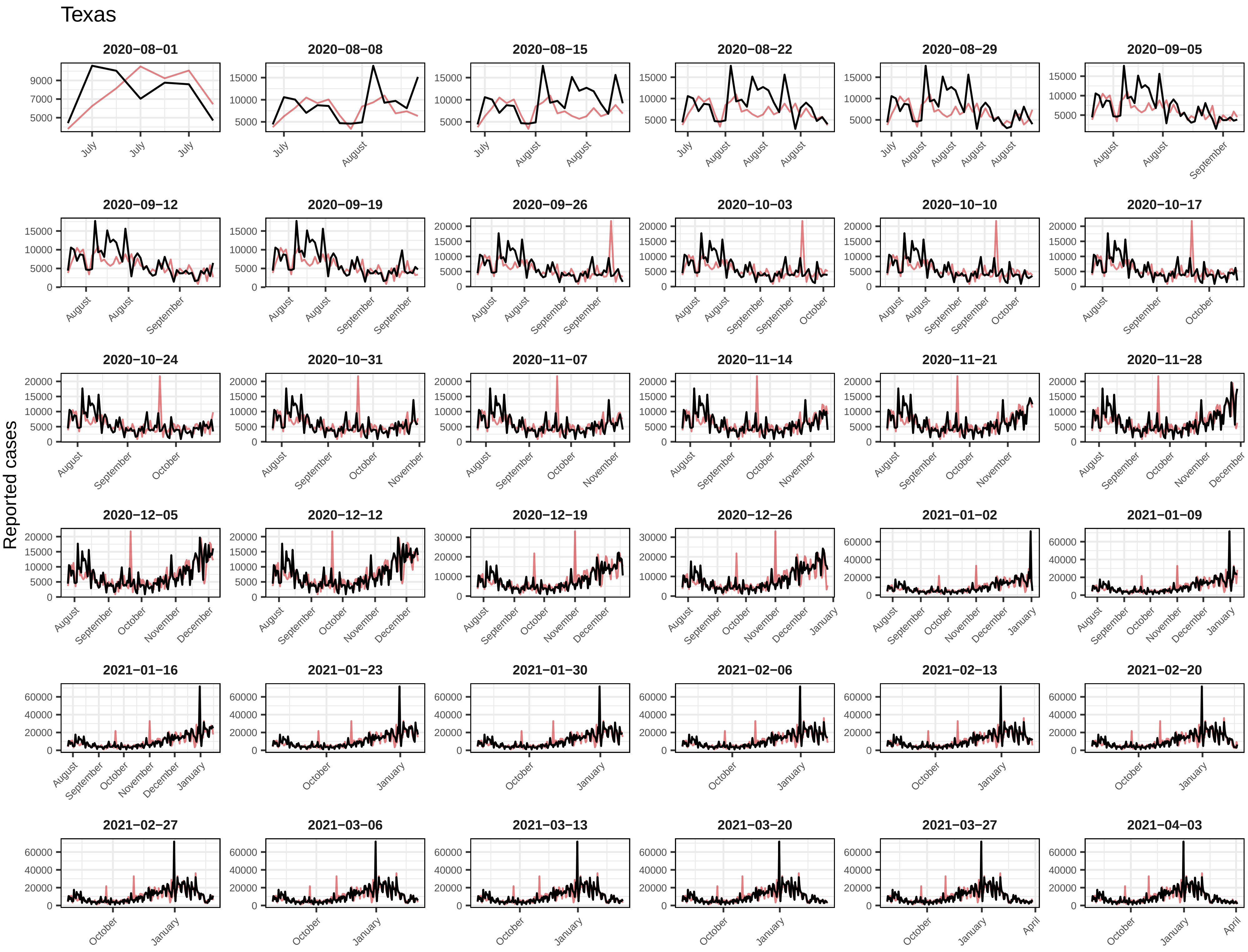
Texas
Reported cases
2020−08−01
2020−08−08
2020−08−15
2020−08−22
2020−08−29
2020−09−05
2020−09−12
2020−09−19
2020−09−26
2020−10−03
2020−10−10
2020−10−17
2020−10−24
2020−10−31
2020−11−07
2020−11−14
2020−11−21
2020−11−28
2020−12−05
2020−12−12
2020−12−19
2020−12−26
2021−01−02
2021−01−09
2021−01−16
2021−01−23
2021−01−30
2021−02−06
2021−02−13
2021−02−20
2021−02−27
2021−03−06
2021−03−13
2021−03−20
2021−03−27
2021−04−03

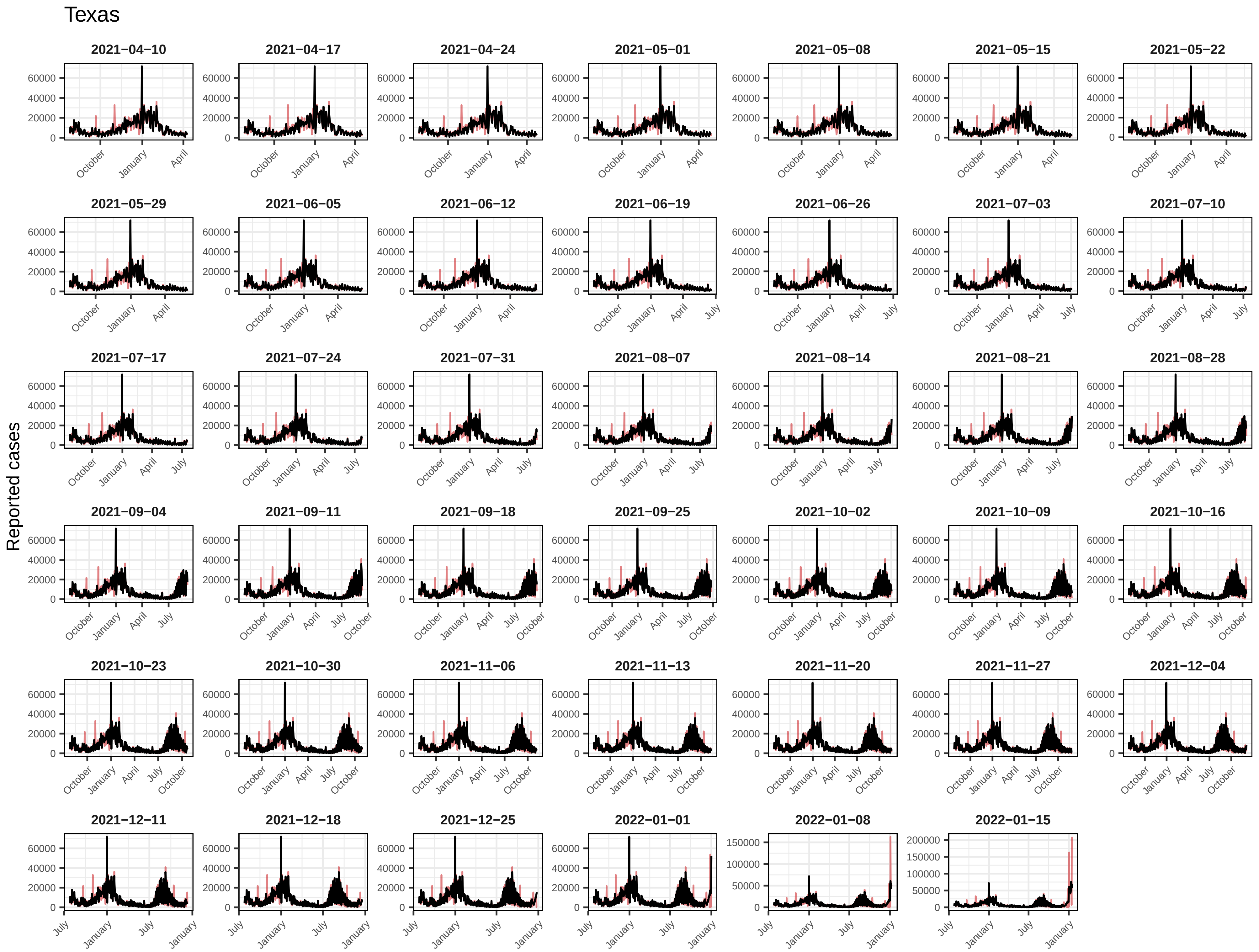

Texas
Reported cases
2021−04−10
2021−04−17
2021−04−24
2021−05−01
2021−05−08
2021−05−15
2021−05−22
2021−05−29
2021−06−05
2021−06−12
2021−06−19
2021−06−26
2021−07−03
2021−07−10
2021−07−17
2021−07−24
2021−07−31
2021−08−07
2021−08−14
2021−08−21
2021−08−28
2021−09−04
2021−09−11
2021−09−18
2021−09−25
2021−10−02
2021−10−09
2021−10−16
2021−10−23
2021−10−30
2021−11−06
2021−11−13
2021−11−20
2021−11−27
2021−12−04
2021−12−11
2021−12−18
2021−12−25
2022−01−01
2022−01−08
2022−01−15
0
20000
40000
60000
50000
100000
150000
200000
October
January
April
July

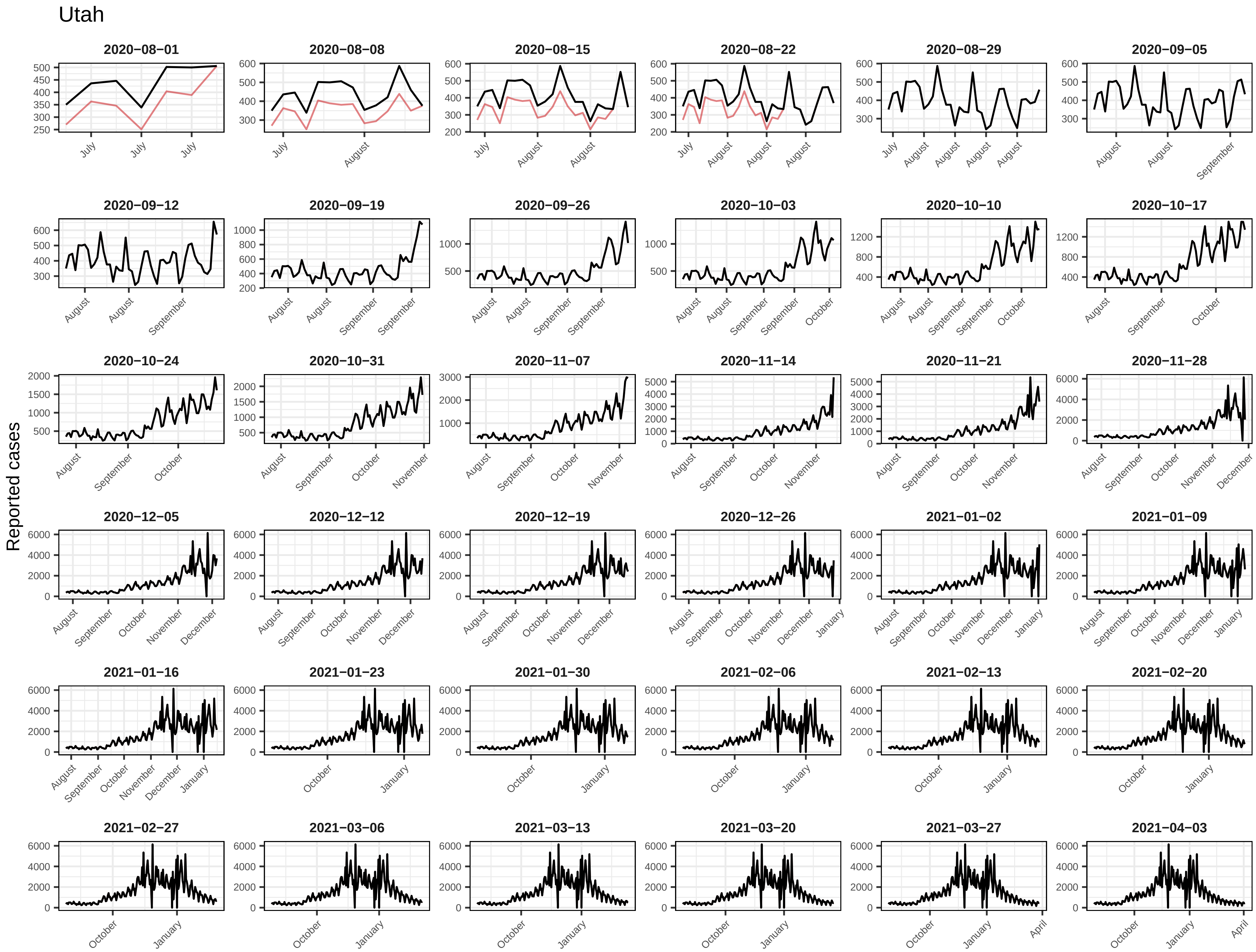
Utah
Reported cases
2020−08−01
2020−08−08
2020−08−15
2020−08−22
2020−08−29
2020−09−05
2020−09−12
2020−09−19
2020−09−26
2020−10−03
2020−10−10
2020−10−17
2020−10−24
2020−10−31
2020−11−07
2020−11−14
2020−11−21
2020−11−28
2020−12−05
2020−12−12
2020−12−19
2020−12−26
2021−01−02
2021−01−09
2021−01−16
2021−01−23
2021−01−30
2021−02−06
2021−02−13
2021−02−20
2021−02−27
2021−03−06
2021−03−13
2021−03−20
2021−03−27
2021−04−03

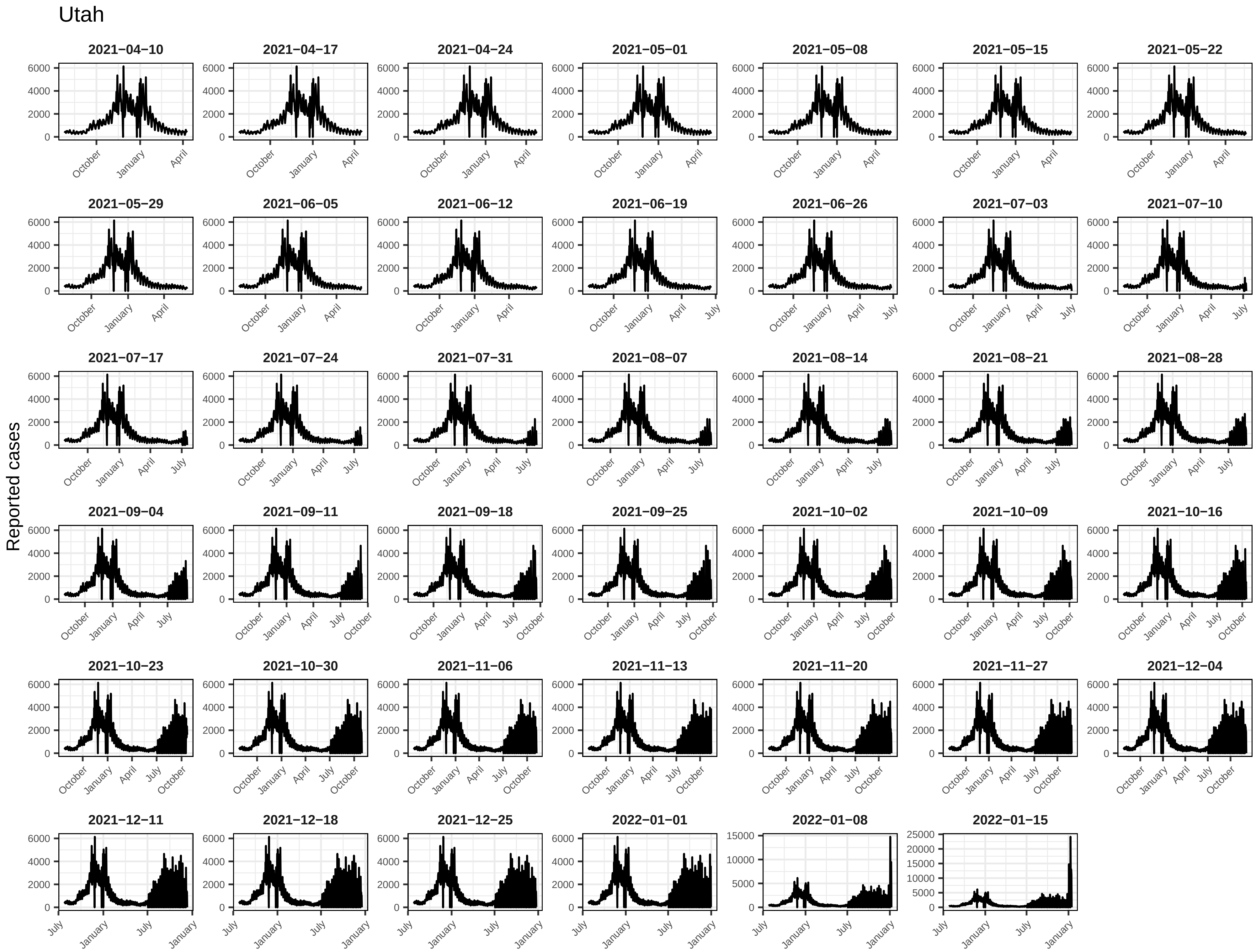
Utah
Reported cases
2021−04−10
2021−04−17
2021−04−24
2021−05−01
2021−05−08
2021−05−15
2021−05−22
2021−05−29
2021−06−05
2021−06−12
2021−06−19
2021−06−26
2021−07−03
2021−07−10
2021−07−17
2021−07−24
2021−07−31
2021−08−07
2021−08−14
2021−08−21
2021−08−28
2021−09−04
2021−09−11
2021−09−18
2021−09−25
2021−10−02
2021−10−09
2021−10−16
2021−10−23
2021−10−30
2021−11−06
2021−11−13
2021−11−20
2021−11−27
2021−12−04
2021−12−11
2021−12−18
2021−12−25
2022−01−01
2022−01−08
2022−01−15
0
2000
4000
6000
5000
10000
15000
20000
25000
October
January
April
July

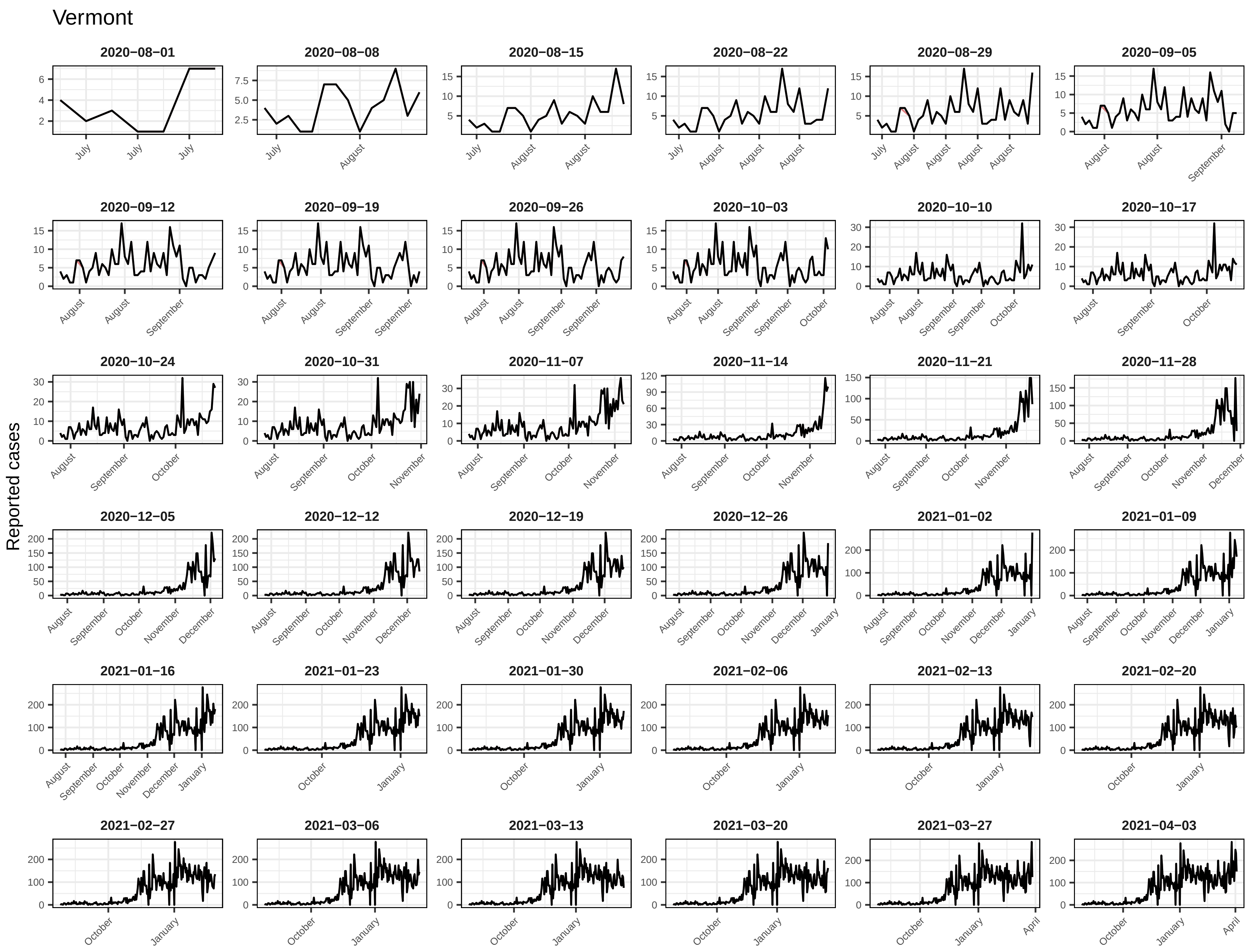
Vermont
Reported cases
2020−08−01
2020−08−08
2020−08−15
2020−08−22
2020−08−29
2020−09−05
2020−09−12
2020−09−19
2020−09−26
2020−10−03
2020−10−10
2020−10−17
2020−10−24
2020−10−31
2020−11−07
2020−11−14
2020−11−21
2020−11−28
2020−12−05
2020−12−12
2020−12−19
2020−12−26
2021−01−02
2021−01−09
2021−01−16
2021−01−23
2021−01−30
2021−02−06
2021−02−13
2021−02−20
2021−02−27
2021−03−06
2021−03−13
2021−03−20
2021−03−27
2021−04−03

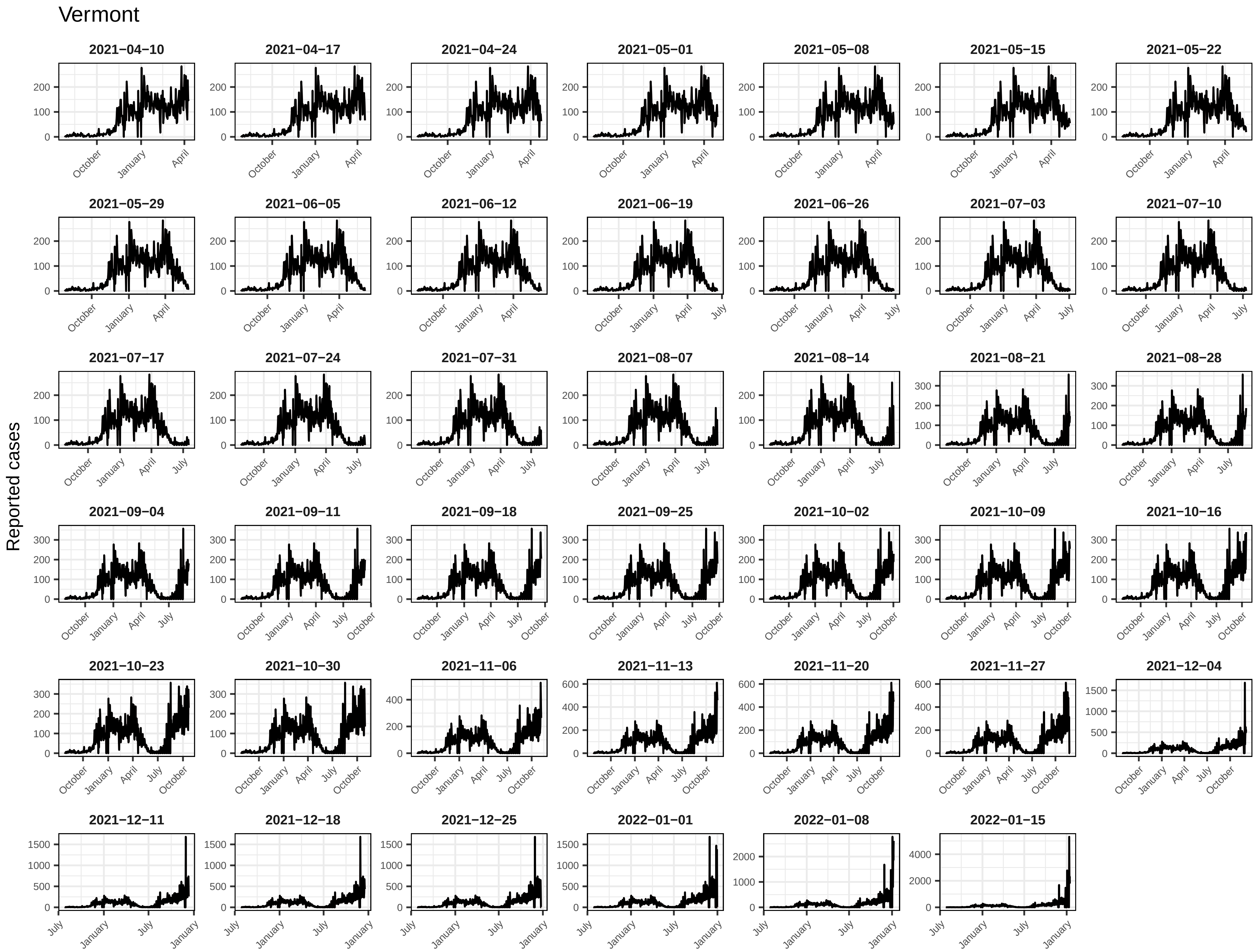
Vermont
Reported cases
2021−04−10
2021−04−17
2021−04−24
2021−05−01
2021−05−08
2021−05−15
2021−05−22
2021−05−29
2021−06−05
2021−06−12
2021−06−19
2021−06−26
2021−07−03
2021−07−10
2021−07−17
2021−07−24
2021−07−31
2021−08−07
2021−08−14
2021−08−21
2021−08−28
2021−09−04
2021−09−11
2021−09−18
2021−09−25
2021−10−02
2021−10−09
2021−10−16
2021−10−23
2021−10−30
2021−11−06
2021−11−13
2021−11−20
2021−11−27
2021−12−04
2021−12−11
2021−12−18
2021−12−25
2022−01−01
2022−01−08
2022−01−15

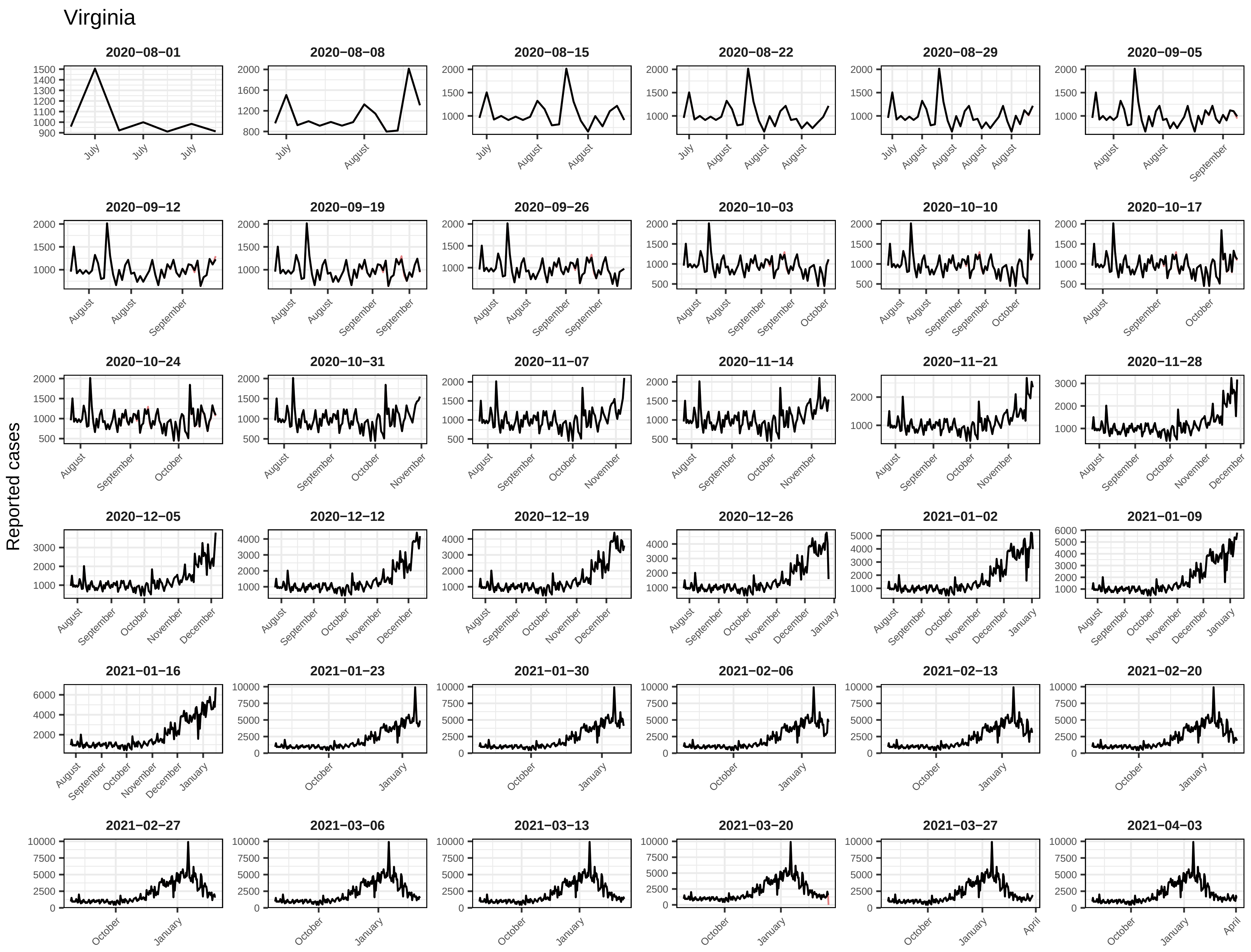
Virginia
Reported cases
2020−08−01
2020−08−08
2020−08−15
2020−08−22
2020−08−29
2020−09−05
2020−09−12
2020−09−19
2020−09−26
2020−10−03
2020−10−10
2020−10−17
2020−10−24
2020−10−31
2020−11−07
2020−11−14
2020−11−21
2020−11−28
2020−12−05
2020−12−12
2020−12−19
2020−12−26
2021−01−02
2021−01−09
2021−01−16
2021−01−23
2021−01−30
2021−02−06
2021−02−13
2021−02−20
2021−02−27
2021−03−06
2021−03−13
2021−03−20
2021−03−27
2021−04−03

#### Virginia

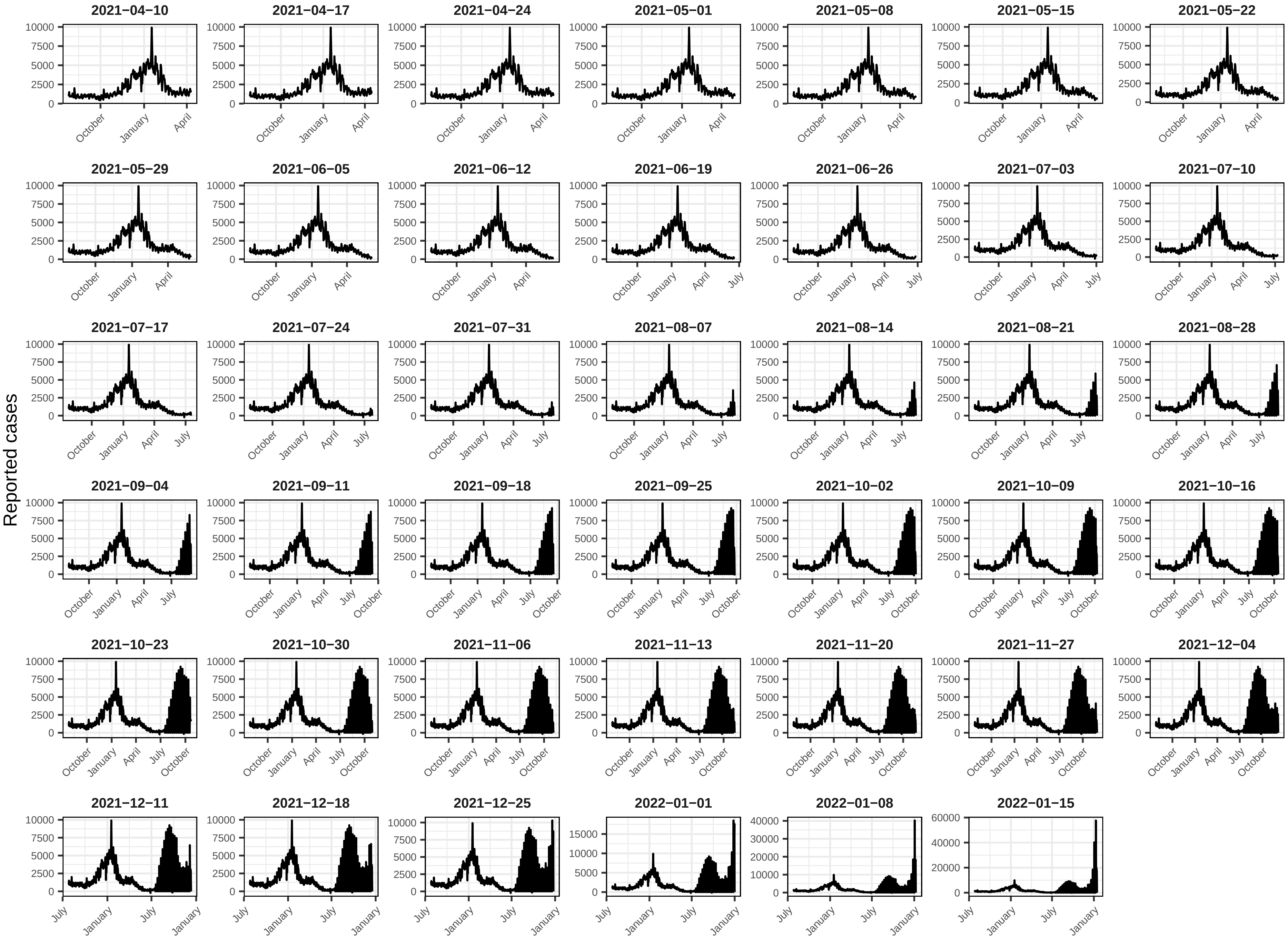

Reported cases
2021−04−10
2021−04−17
2021−04−24
2021−05−01
2021−05−08
2021−05−15
2021−05−22
2021−05−29
2021−06−05
2021−06−12
2021−06−19
2021−06−26
2021−07−03
2021−07−10
2021−07−17
2021−07−24
2021−07−31
2021−08−07
2021−08−14
2021−08−21
2021−08−28
2021−09−04
2021−09−11
2021−09−18
2021−09−25
2021−10−02
2021−10−09
2021−10−16
2021−10−23
2021−10−30
2021−11−06
2021−11−13
2021−11−20
2021−11−27
2021−12−04
2021−12−11
2021−12−18
2021−12−25
2022−01−01
2022−01−08
2022−01−15

Washington

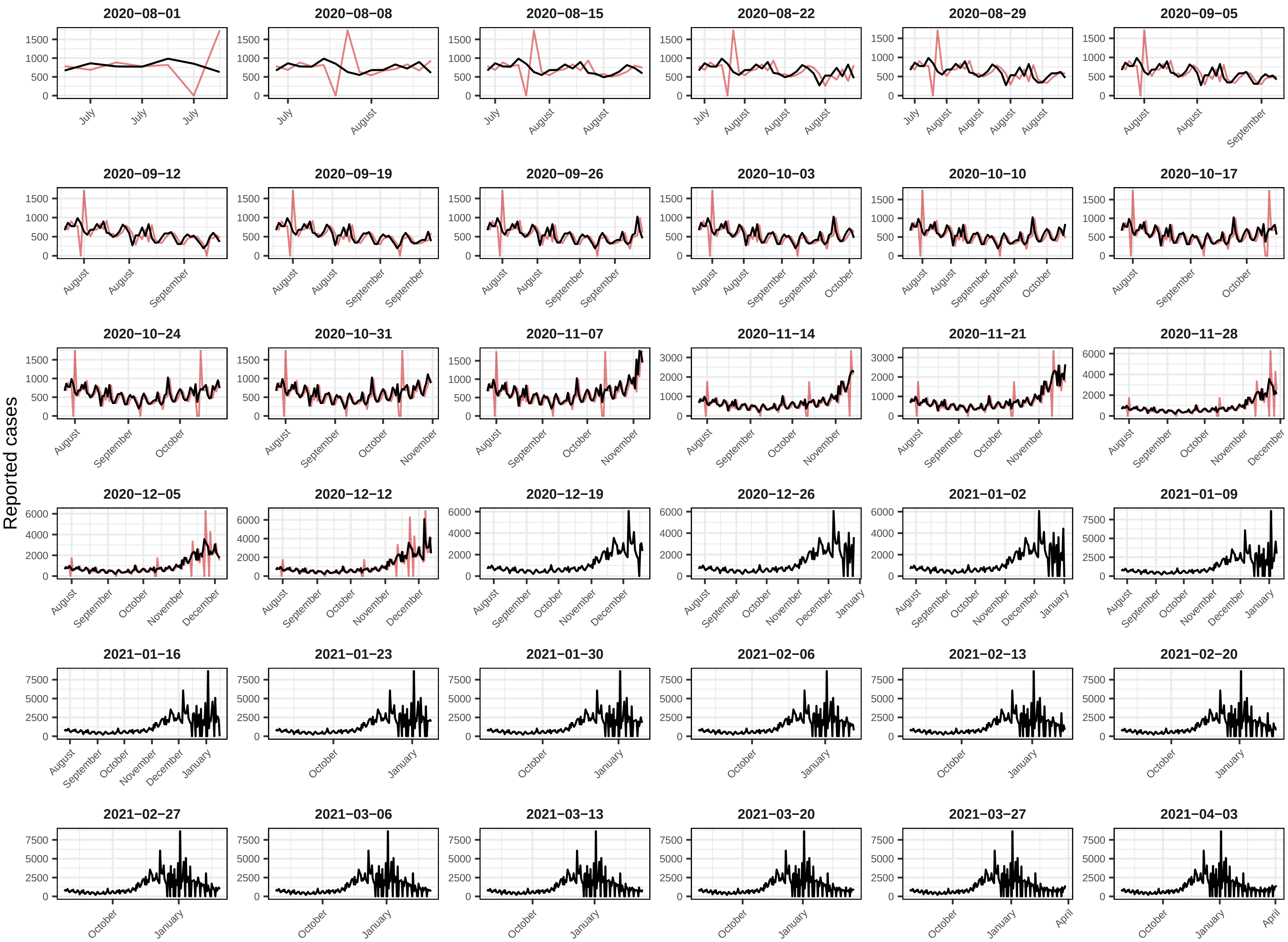
Reported cases
2020−08−01
2020−08−08
2020−08−15
2020−08−22
2020−08−29
2020−09−05
2020−09−12
2020−09−19
2020−09−26
2020−10−03
2020−10−10
2020−10−17
2020−10−24
2020−10−31
2020−11−07
2020−11−14
2020−11−21
2020−11−28
2020−12−05
2020−12−12
2020−12−19
2020−12−26
2021−01−02
2021−01−09
2021−01−16
2021−01−23
2021−01−30
2021−02−06
2021−02−13
2021−02−20
2021−02−27
2021−03−06
2021−03−13
2021−03−20
2021−03−27
2021−04−03

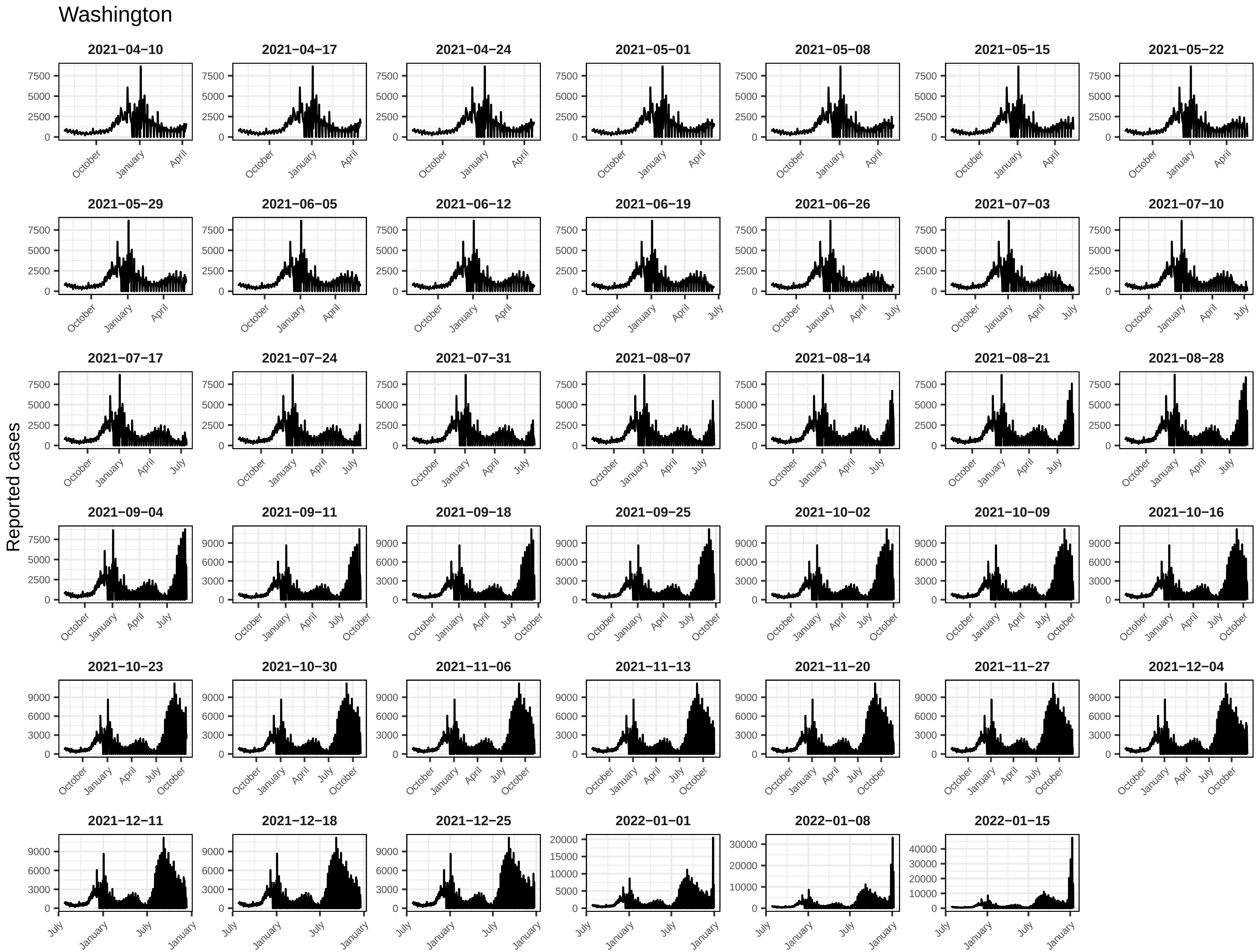
Washington
Reported cases
2021−04−10
2021−04−17
2021−04−24
2021−05−01
2021−05−08
2021−05−15
2021−05−22
2021−05−29
2021−06−05
2021−06−12
2021−06−19
2021−06−26
2021−07−03
2021−07−10
2021−07−17
2021−07−24
2021−07−31
2021−08−07
2021−08−14
2021−08−21
2021−08−28
2021−09−04
2021−09−11
2021−09−18
2021−09−25
2021−10−02
2021−10−09
2021−10−16
2021−10−23
2021−10−30
2021−11−06
2021−11−13
2021−11−20
2021−11−27
2021−12−04
2021−12−11
2021−12−18
2021−12−25
2022−01−01
2022−01−08
2022−01−15
0
2500
5000
7500
3000
6000
9000
10000
15000
20000
30000
40000
October
January
April
July

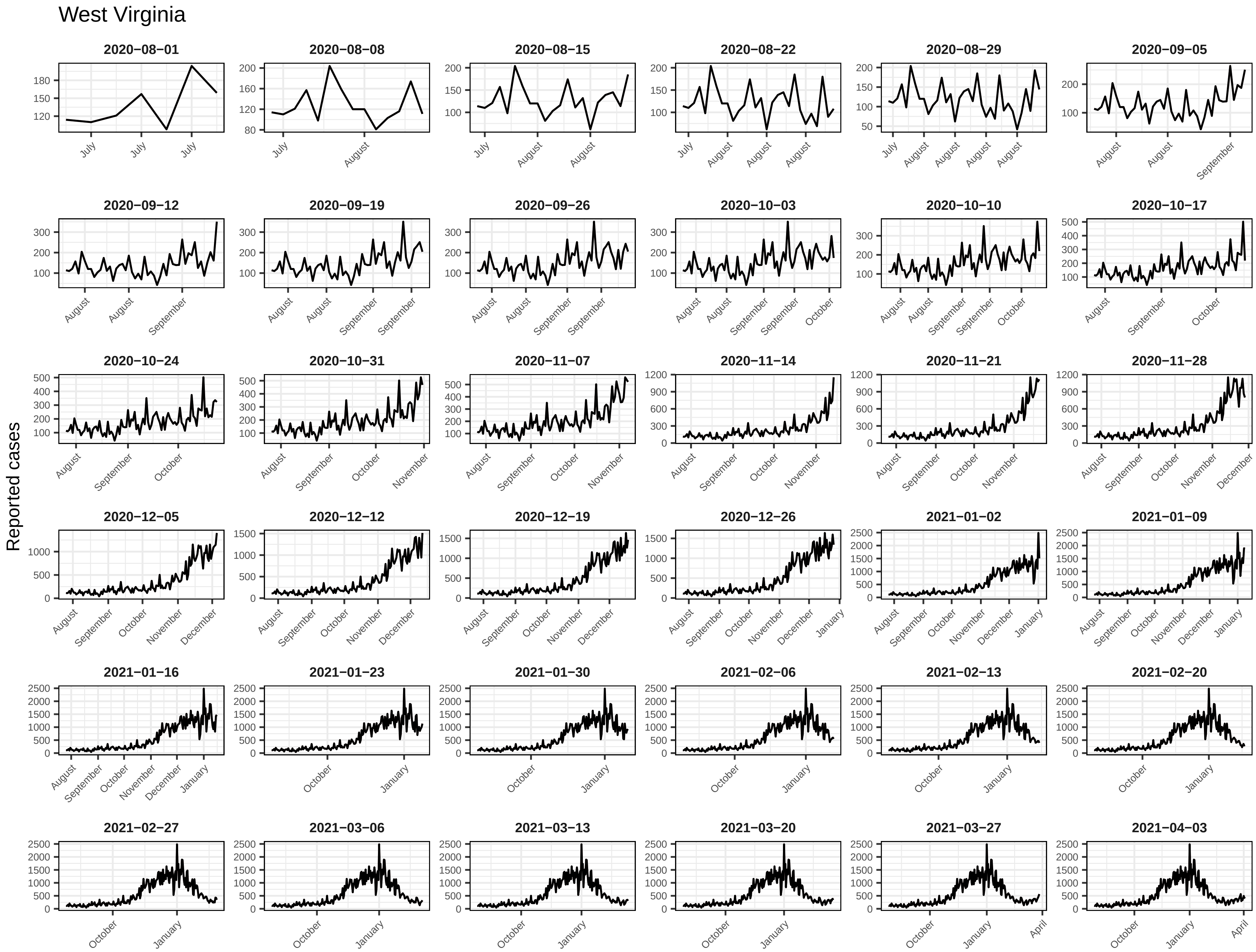
West Virginia
Reported cases
2020−08−01
2020−08−08
2020−08−15
2020−08−22
2020−08−29
2020−09−05
2020−09−12
2020−09−19
2020−09−26
2020−10−03
2020−10−10
2020−10−17
2020−10−24
2020−10−31
2020−11−07
2020−11−14
2020−11−21
2020−11−28
2020−12−05
2020−12−12
2020−12−19
2020−12−26
2021−01−02
2021−01−09
2021−01−16
2021−01−23
2021−01−30
2021−02−06
2021−02−13
2021−02−20
2021−02−27
2021−03−06
2021−03−13
2021−03−20
2021−03−27
2021−04−03

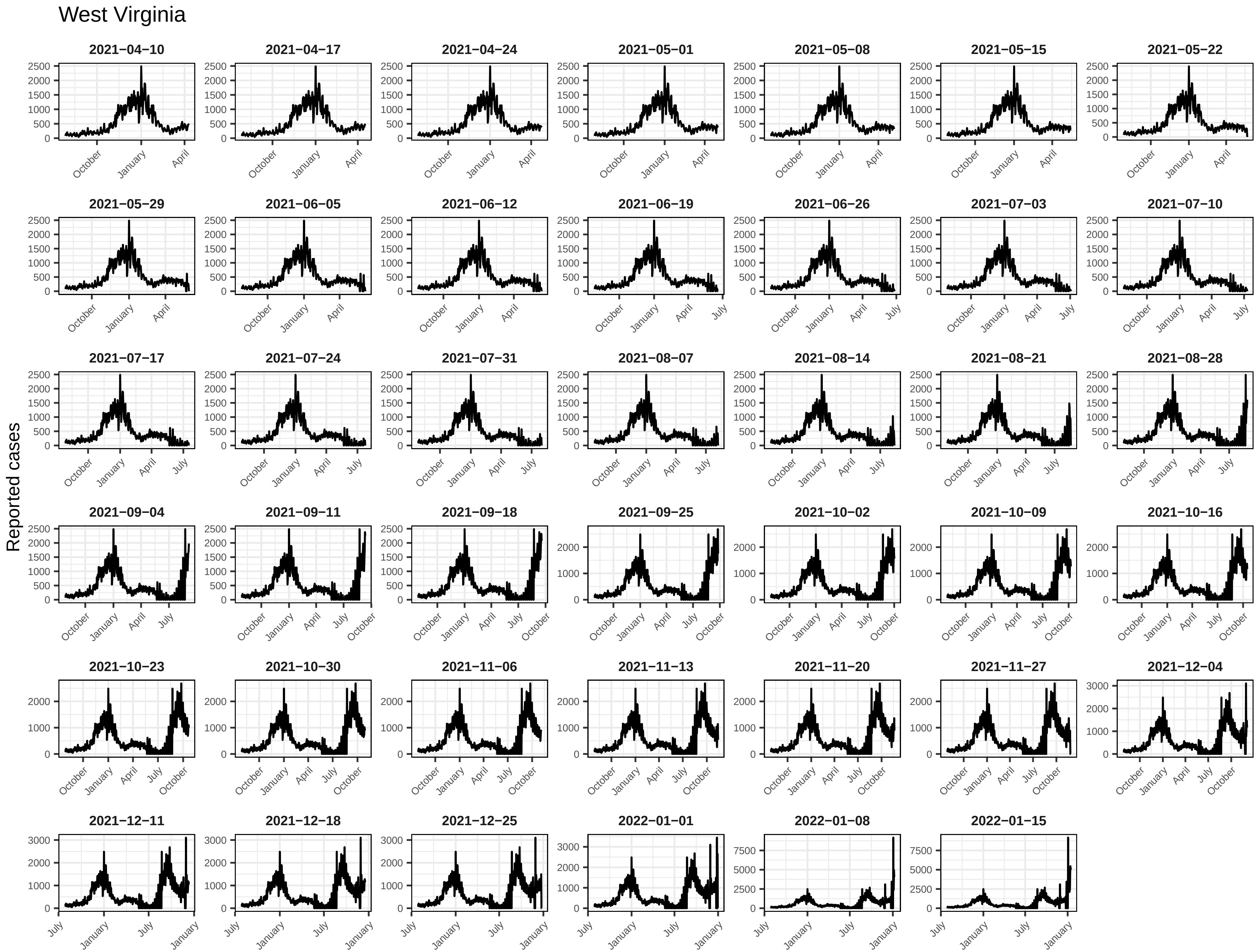
West Virginia
Reported cases
2021−04−10
2021−04−17
2021−04−24
2021−05−01
2021−05−08
2021−05−15
2021−05−22
2021−05−29
2021−06−05
2021−06−12
2021−06−19
2021−06−26
2021−07−03
2021−07−10
2021−07−17
2021−07−24
2021−07−31
2021−08−07
2021−08−14
2021−08−21
2021−08−28
2021−09−04
2021−09−11
2021−09−18
2021−09−25
2021−10−02
2021−10−09
2021−10−16
2021−10−23
2021−10−30
2021−11−06
2021−11−13
2021−11−20
2021−11−27
2021−12−04
2021−12−11
2021−12−18
2021−12−25
2022−01−01
2022−01−08
2022−01−15
October
January
April
July

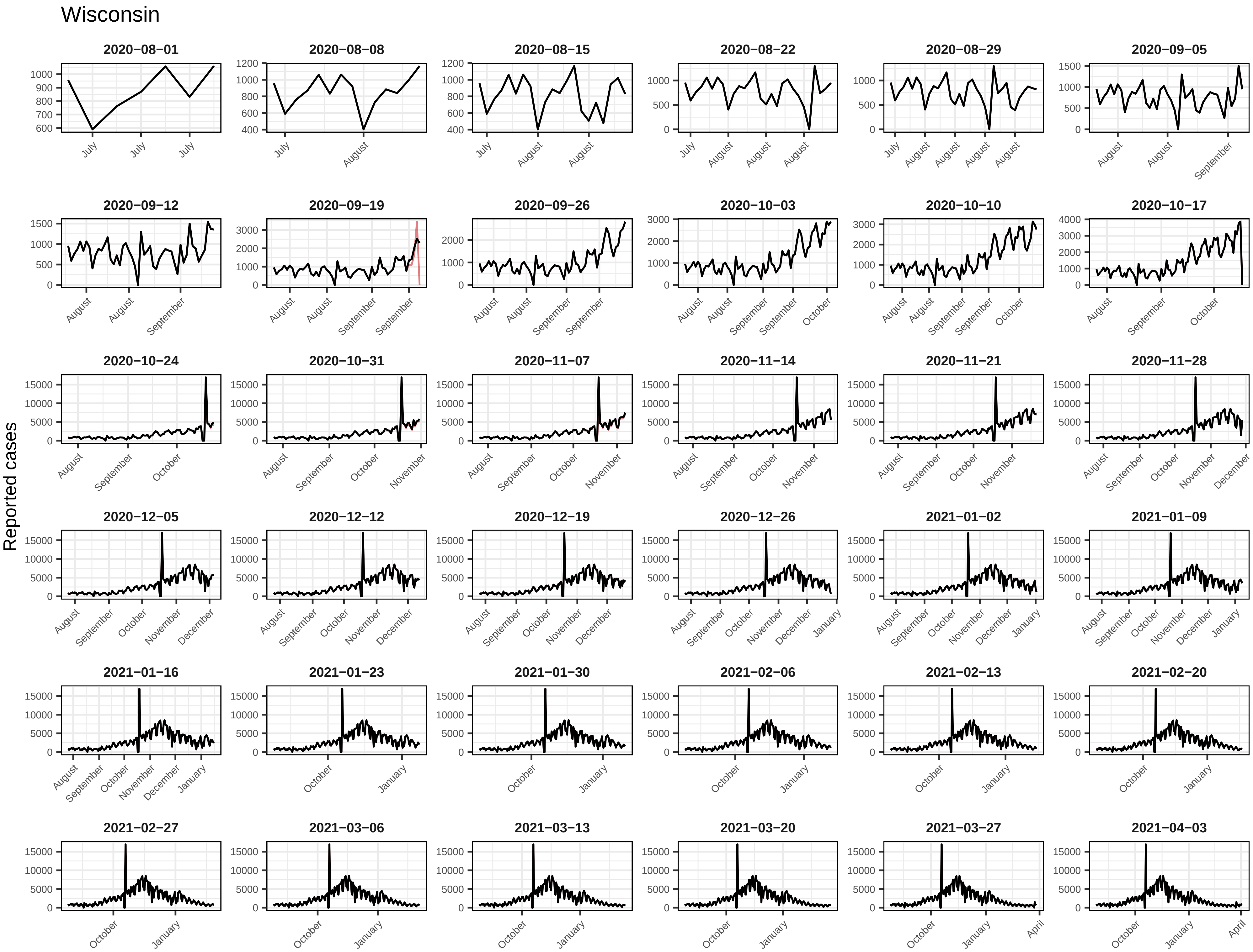
Wisconsin
Reported cases
2020−08−01
2020−08−08
2020−08−15
2020−08−22
2020−08−29
2020−09−05
2020−09−12
2020−09−19
2020−09−26
2020−10−03
2020−10−10
2020−10−17
2020−10−24
2020−10−31
2020−11−07
2020−11−14
2020−11−21
2020−11−28
2020−12−05
2020−12−12
2020−12−19
2020−12−26
2021−01−02
2021−01−09
2021−01−16
2021−01−23
2021−01−30
2021−02−06
2021−02−13
2021−02−20
2021−02−27
2021−03−06
2021−03−13
2021−03−20
2021−03−27
2021−04−03

Wisconsin

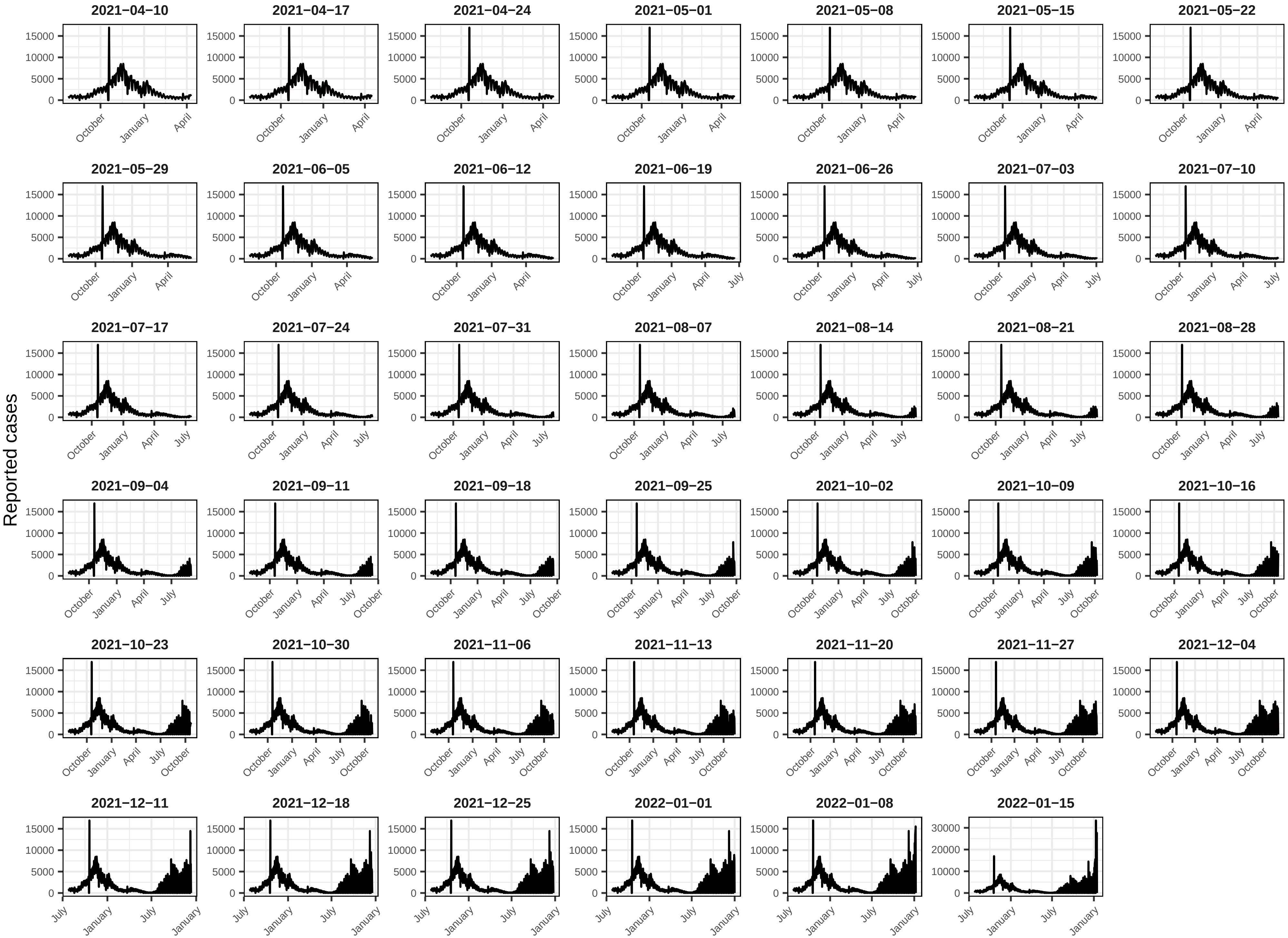
Reported cases
2021−04−10
2021−04−17
2021−04−24
2021−05−01
2021−05−08
2021−05−15
2021−05−22
2021−05−29
2021−06−05
2021−06−12
2021−06−19
2021−06−26
2021−07−03
2021−07−10
2021−07−17
2021−07−24
2021−07−31
2021−08−07
2021−08−14
2021−08−21
2021−08−28
2021−09−04
2021−09−11
2021−09−18
2021−09−25
2021−10−02
2021−10−09
2021−10−16
2021−10−23
2021−10−30
2021−11−06
2021−11−13
2021−11−20
2021−11−27
2021−12−04
2021−12−11
2021−12−18
2021−12−25
2022−01−01
2022−01−08
2022−01−15

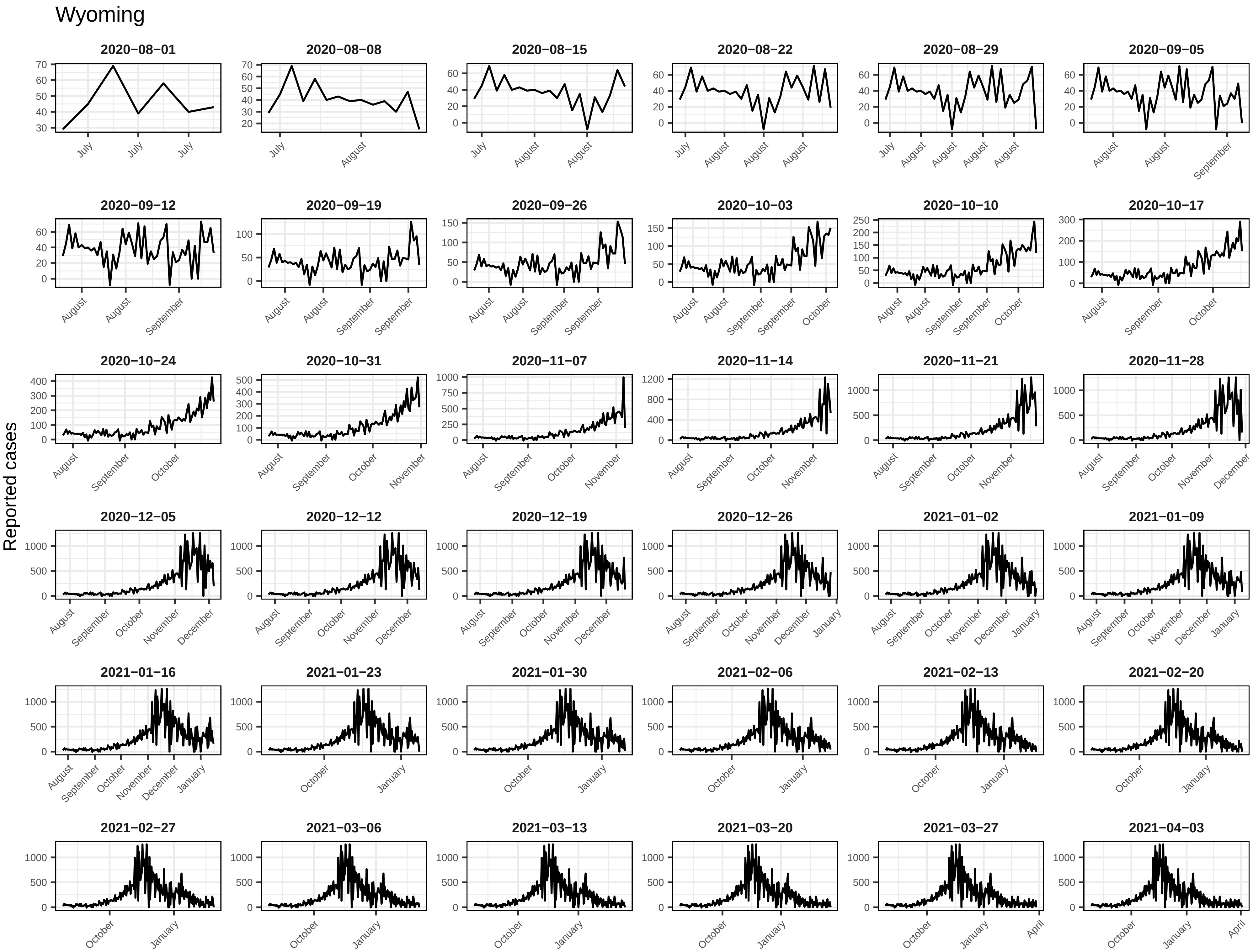
Wyoming
Reported cases
2020−08−01
2020−08−08
2020−08−15
2020−08−22
2020−08−29
2020−09−05
2020−09−12
2020−09−19
2020−09−26
2020−10−03
2020−10−10
2020−10−17
2020−10−24
2020−10−31
2020−11−07
2020−11−14
2020−11−21
2020−11−28
2020−12−05
2020−12−12
2020−12−19
2020−12−26
2021−01−02
2021−01−09
2021−01−16
2021−01−23
2021−01−30
2021−02−06
2021−02−13
2021−02−20
2021−02−27
2021−03−06
2021−03−13
2021−03−20
2021−03−27
2021−04−03

### Wyoming

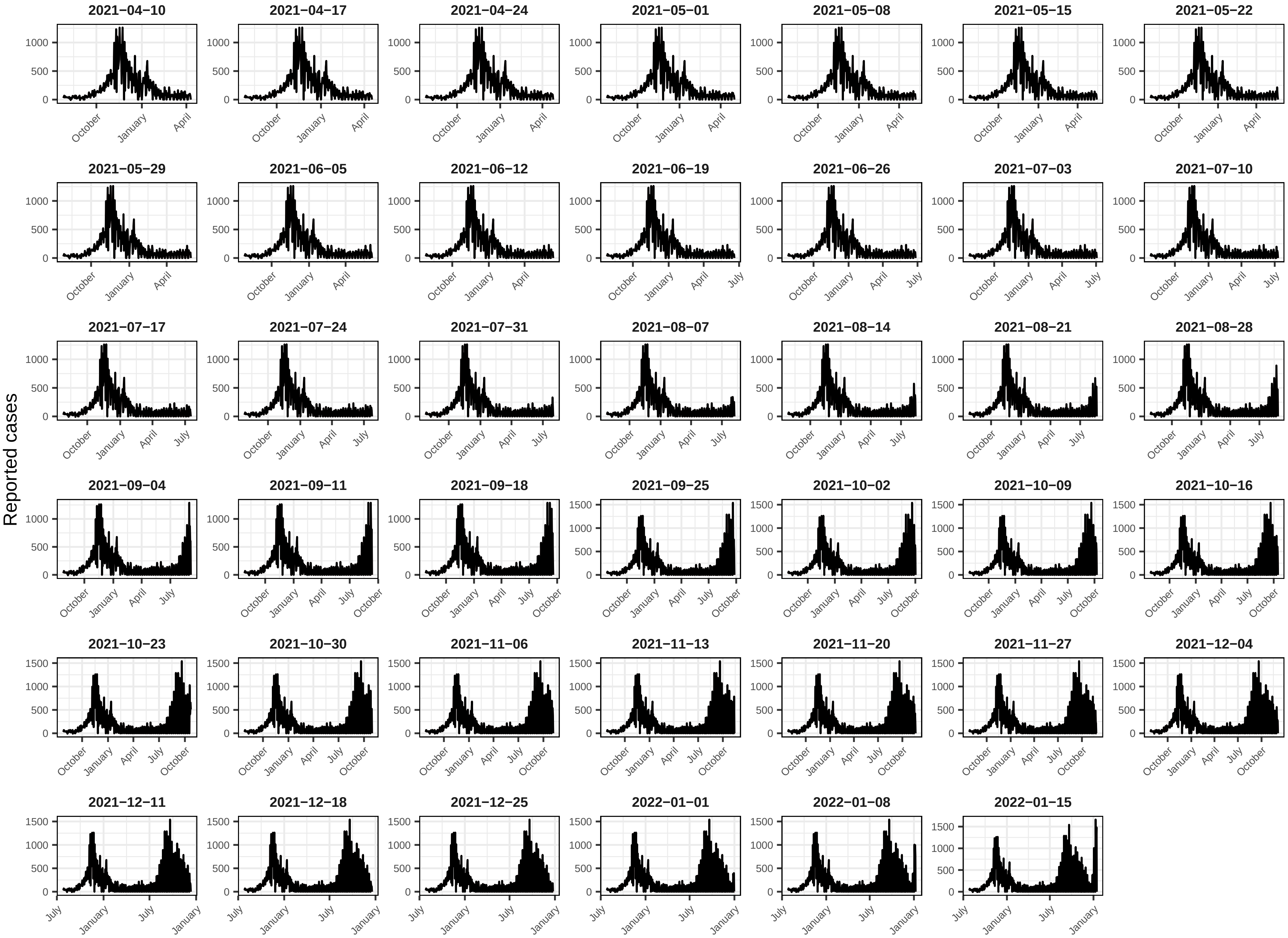
Reported cases
2021−04−10
2021−04−17
2021−04−24
2021−05−01
2021−05−08
2021−05−15
2021−05−22
2021−05−29
2021−06−05
2021−06−12
2021−06−19
2021−06−26
2021−07−03
2021−07−10
2021−07−17
2021−07−24
2021−07−31
2021−08−07
2021−08−14
2021−08−21
2021−08−28
2021−09−04
2021−09−11
2021−09−18
2021−09−25
2021−10−02
2021−10−09
2021−10−16
2021−10−23
2021−10−30
2021−11−06
2021−11−13
2021−11−20
2021−11−27
2021−12−04
2021−12−11
2021−12−18
2021−12−25
2022−01−01
2022−01−08
2022−01−15
