## Supplement 3 for "Challenges of COVID-19 Case Forecasting in the US, 2020-2021"

**Supporting Information 3**: Incident COVID-19 case forecasts were submitted for all US counties. The plots shown here depicted average, scaled pairwise Weighted Interval Score (WIS; see *Methods* for description), 95% coverage, and submissions (S3 Figure 3.1), average 50%, 80% and 95% coverage for eligible submitted forecasts (S3 Figure 3.2), and average WIS and 95% coverage over time (S3 Figure 3.2). Each figure shows spatial disaggregated results, with increasing population size and quantile numbers. For example, counties with the smallest population are grouped in Quantile 1 and the largest population sizes are grouped in Quantile 5. The following teams are included in these figures: CEID-Walk, LNQ-ens1, Microsoft_DeepSTIA, COVIDhub-4_week_ensemble, COVIDhub-trained_ensemble, COVIDhub-baseline, CU-select, FAIR-NRAR, FRBSF_Wilson-Econometric, IowasStateLW-STEM, JHU_IDD-CovidSP, JHU_CSSE-DECOM, JHUAPL-Bucky, LANL-GrowthRate, LNQ-esn1, UVA-Ensemble.

**S3 Figure 3.1.** Percent of weeks with complete submissions for all sets of team forecasts, scaled, pairwise relative Weighted Interval Score (rWIS), 95% coverage, and by geographical scale of submitted forecasts. Teams are sorted by increasing rWIS values.

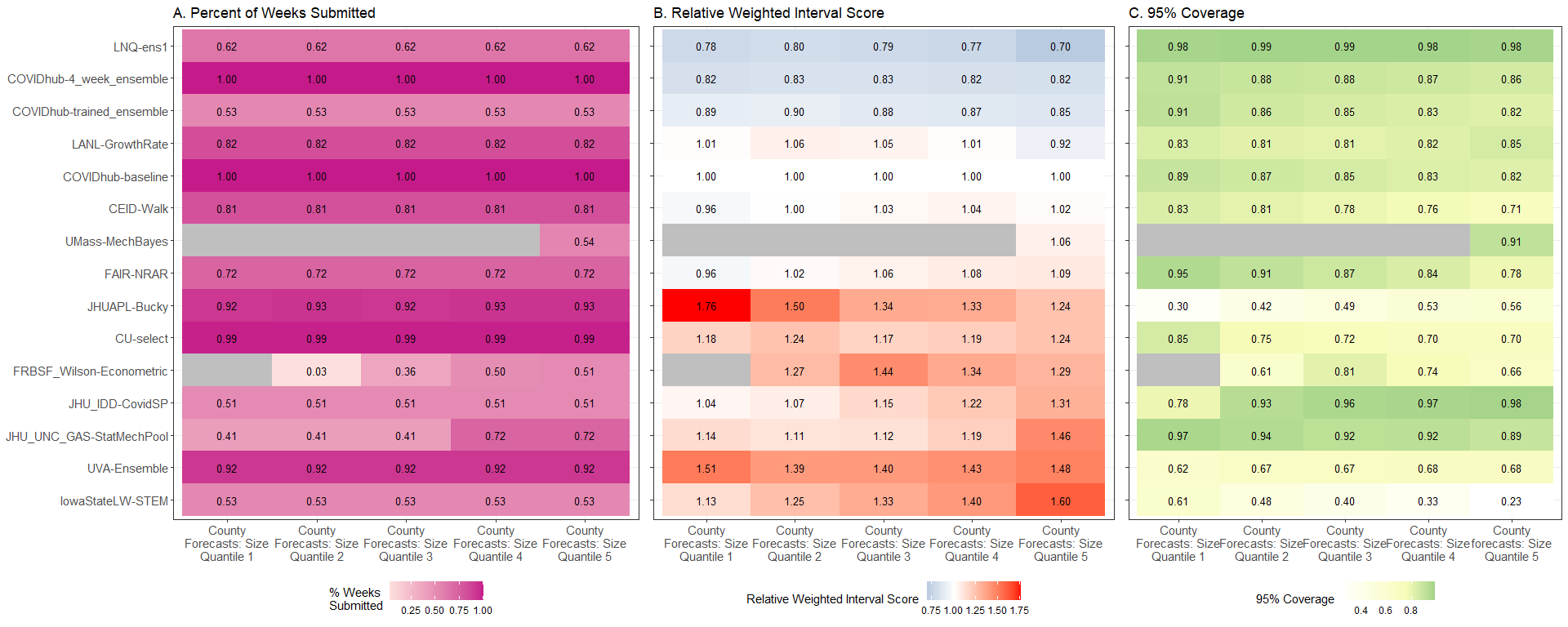

**S3 Figure 3.2**. Expected and observed coverage rates aggregated over time and horizon for county forecasts. The dashed line represents optimal expected coverage. Team forecasts that outperformed the COVIDhub-4_week_ensemble model at all coverage levels are labeled on the right hand side of the plots.

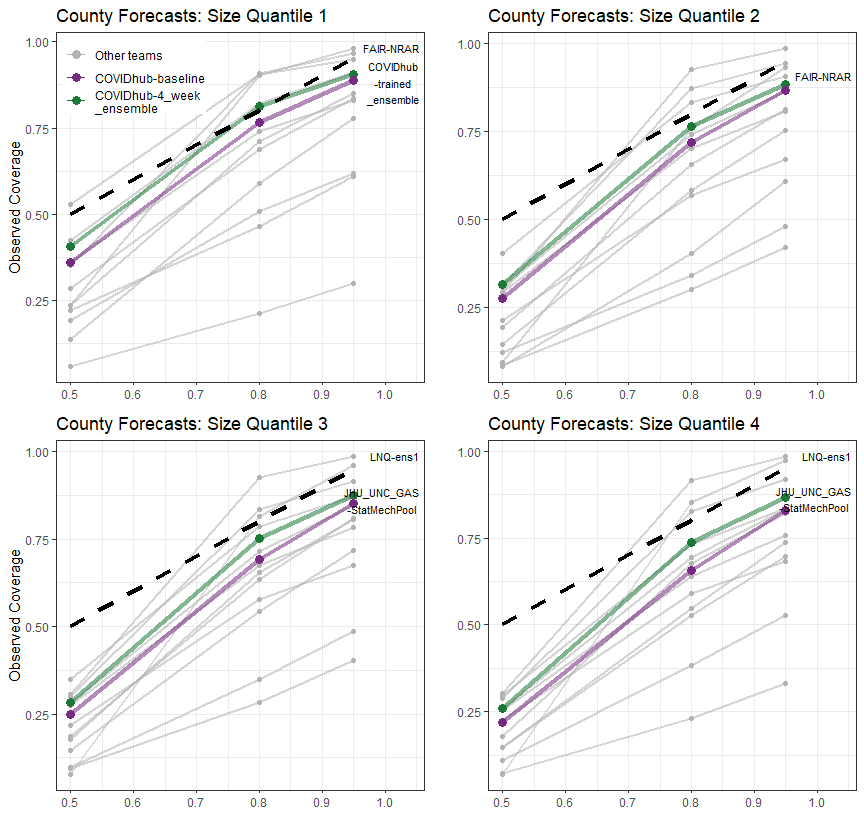

**S3 Figure 3.3.** Mean Weighted Interval Score (WIS) over time, aggregated by geographic units and forecast horizon in A and 95% coverage over time, aggregated by geographic units and forecast horizon in B. The black, dashed vertical line in all panels shows the date that public communication of the case forecasts was paused. The black, dashed horizontal line in panels B show nominal 95% interval coverage

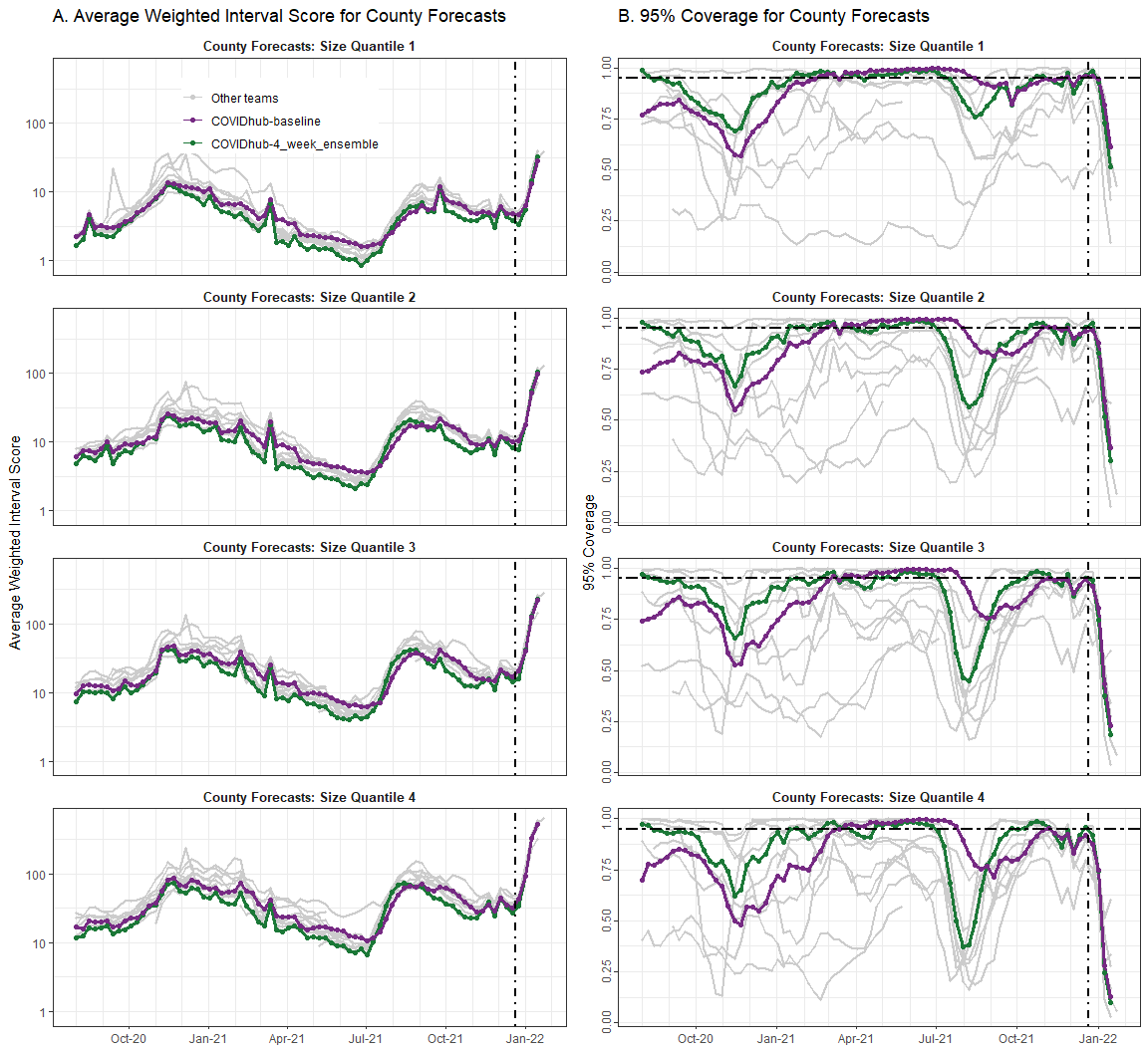
