## Supplement 4 for "Challenges of COVID-19 Case Forecasting in the US, 2020-2021"

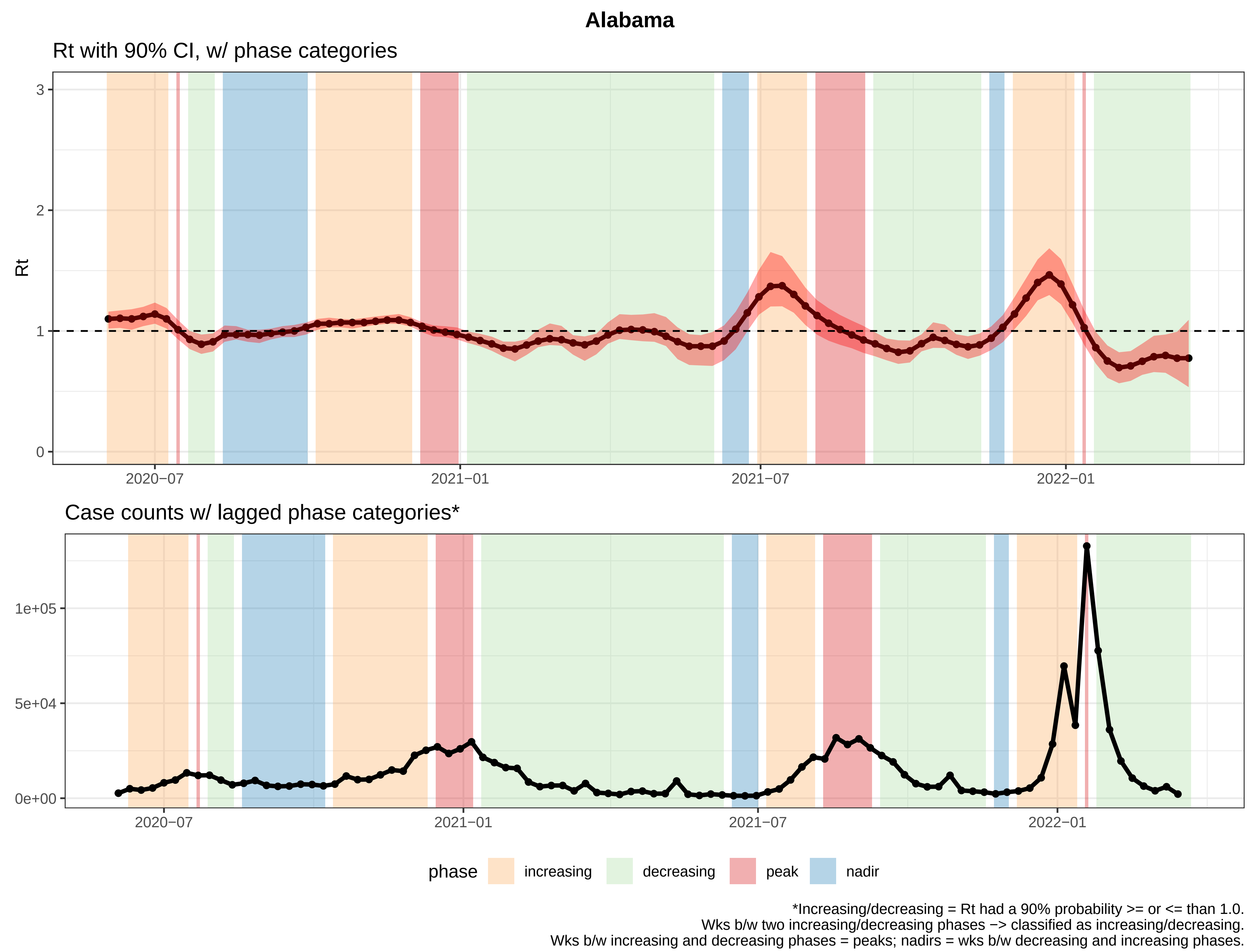
Alabama
Rt with 90% CI, w/ phase categories
Rt
0
1
2
3
2020−07
2021−01
2021−07
2022−01
Case counts w/ lagged phase categories*
0e+00
5e+04
1e+05
2020−07
2021−01
2021−07
2022−01
phase
increasing
decreasing
peak
nadir
*Increasing/decreasing = Rt had a 90% probability >= or <= than 1.0.
Wks b/w two increasing/decreasing phases −> classified as increasing/decreasing.
Wks b/w increasing and decreasing phases = peaks; nadirs = wks b/w decreasing and increasing phases.

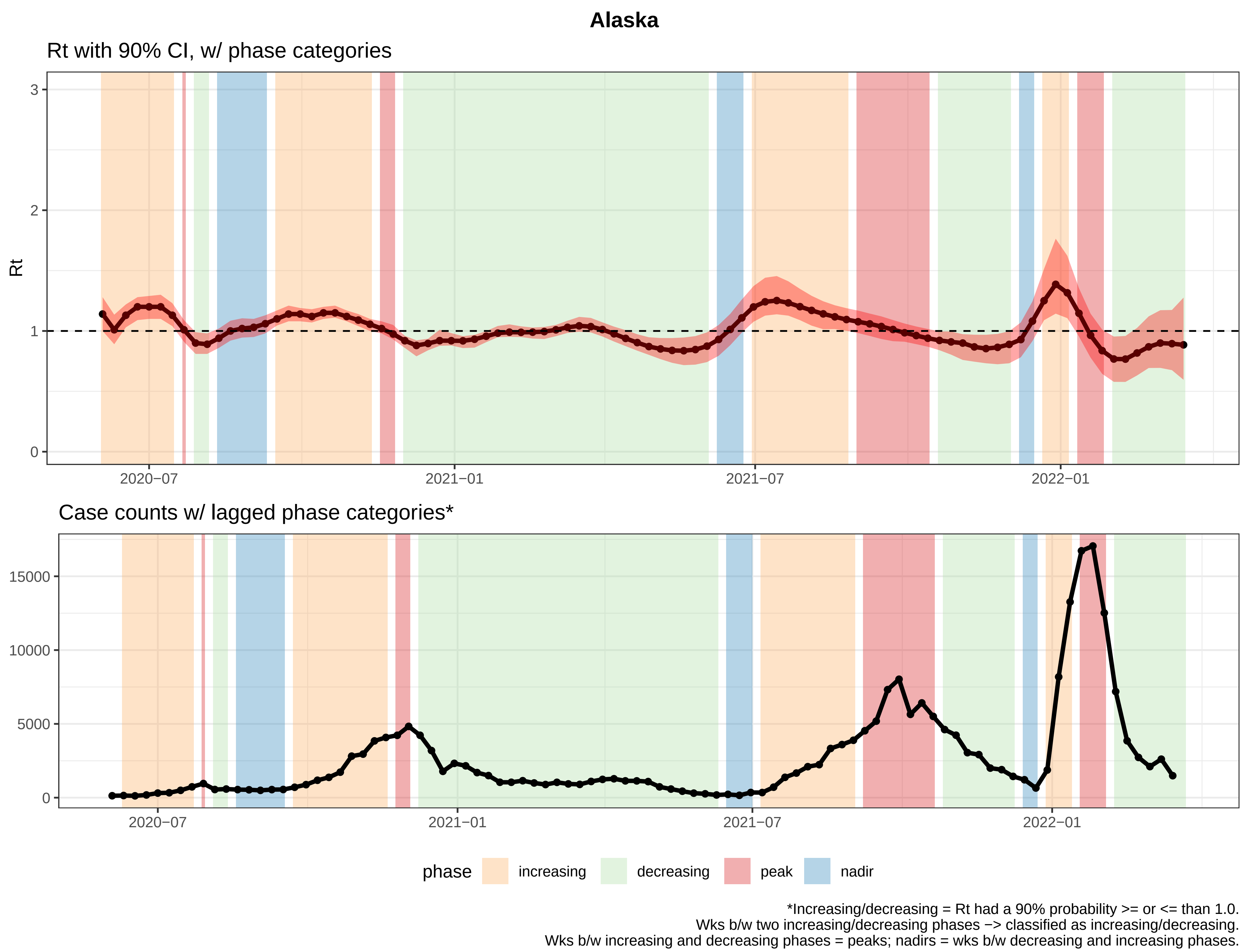
Alaska
Rt with 90% CI, w/ phase categories
Rt
3
2
1
0
2020−07
2021−01
2021−07
2022−01
Case counts w/ lagged phase categories*
15000
10000
5000
0
2020−07
2021−01
2021−07
2022−01
phase
increasing
decreasing
peak
nadir
*Increasing/decreasing = Rt had a 90% probability >= or <= than 1.0.
Wks b/w two increasing/decreasing phases −> classified as increasing/decreasing.
Wks b/w increasing and decreasing phases = peaks; nadirs = wks b/w decreasing and increasing phases.

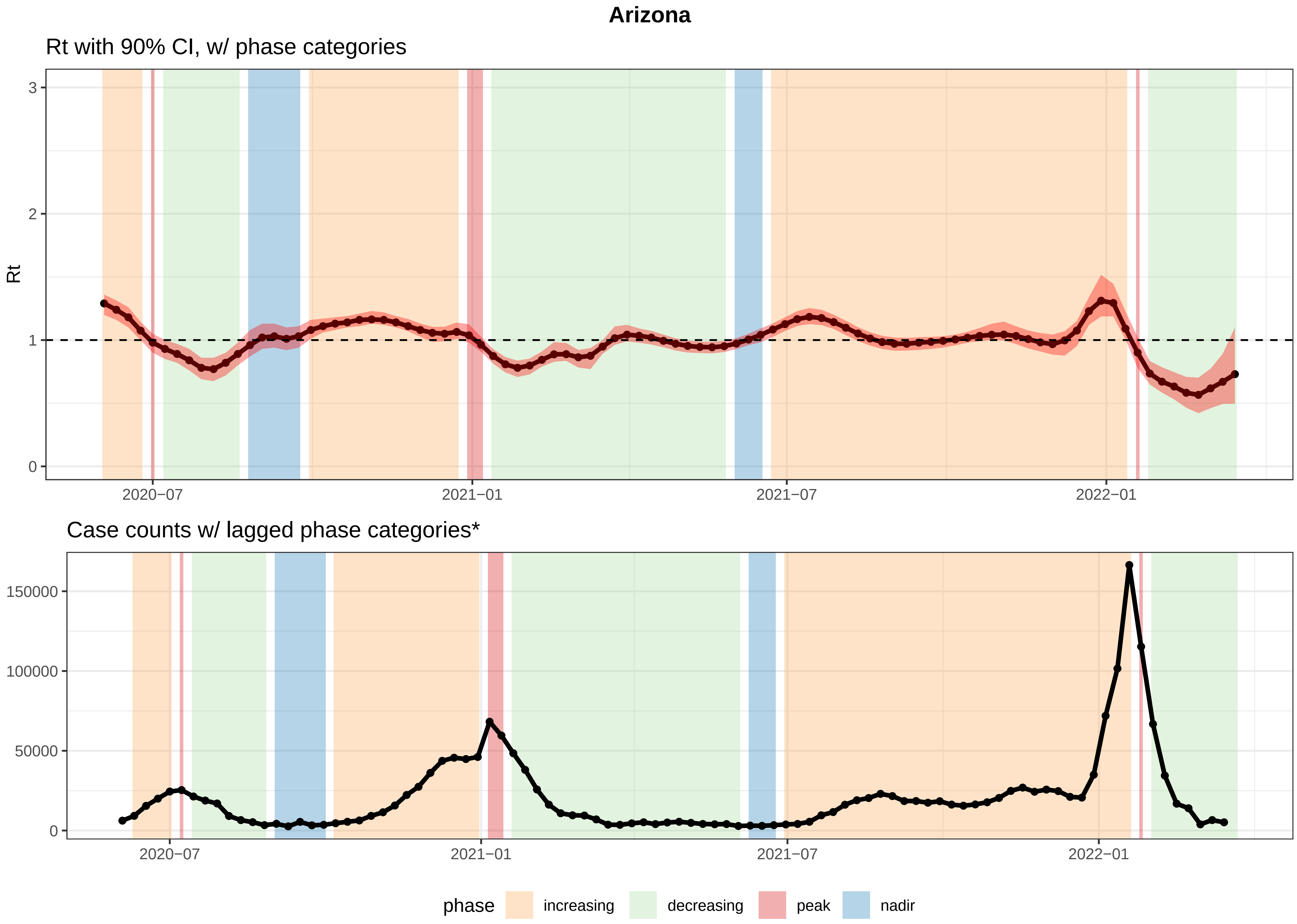


*Increasing/decreasing = Rt had a 90% probability >= or <= than 1.0.
Wks b/w two increasing/decreasing phases −> classified as increasing/decreasing.
Wks b/w increasing and decreasing phases = peaks; nadirs = wks b/w decreasing and increasing phases.

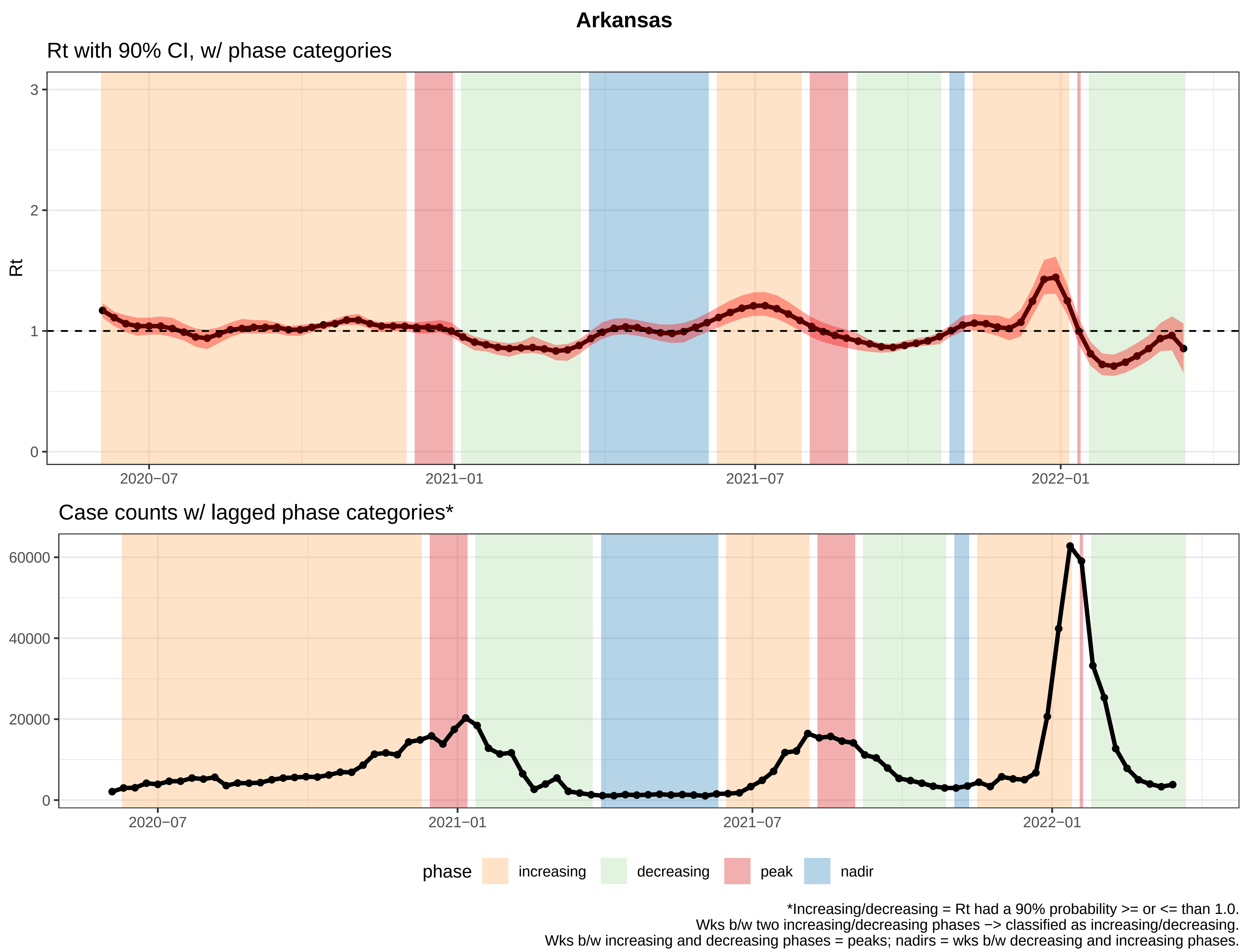
Arkansas
Rt with 90% CI, w/ phase categories
Rt
3
2
1
0
2020−07
2021−01
2021−07
2022−01
Case counts w/ lagged phase categories*
60000
40000
20000
0
2020−07
2021−01
2021−07
2022−01
phase
increasing
decreasing
peak
nadir
*Increasing/decreasing = Rt had a 90% probability >= or <= than 1.0.
Wks b/w two increasing/decreasing phases −> classified as increasing/decreasing.
Wks b/w increasing and decreasing phases = peaks; nadirs = wks b/w decreasing and increasing phases.

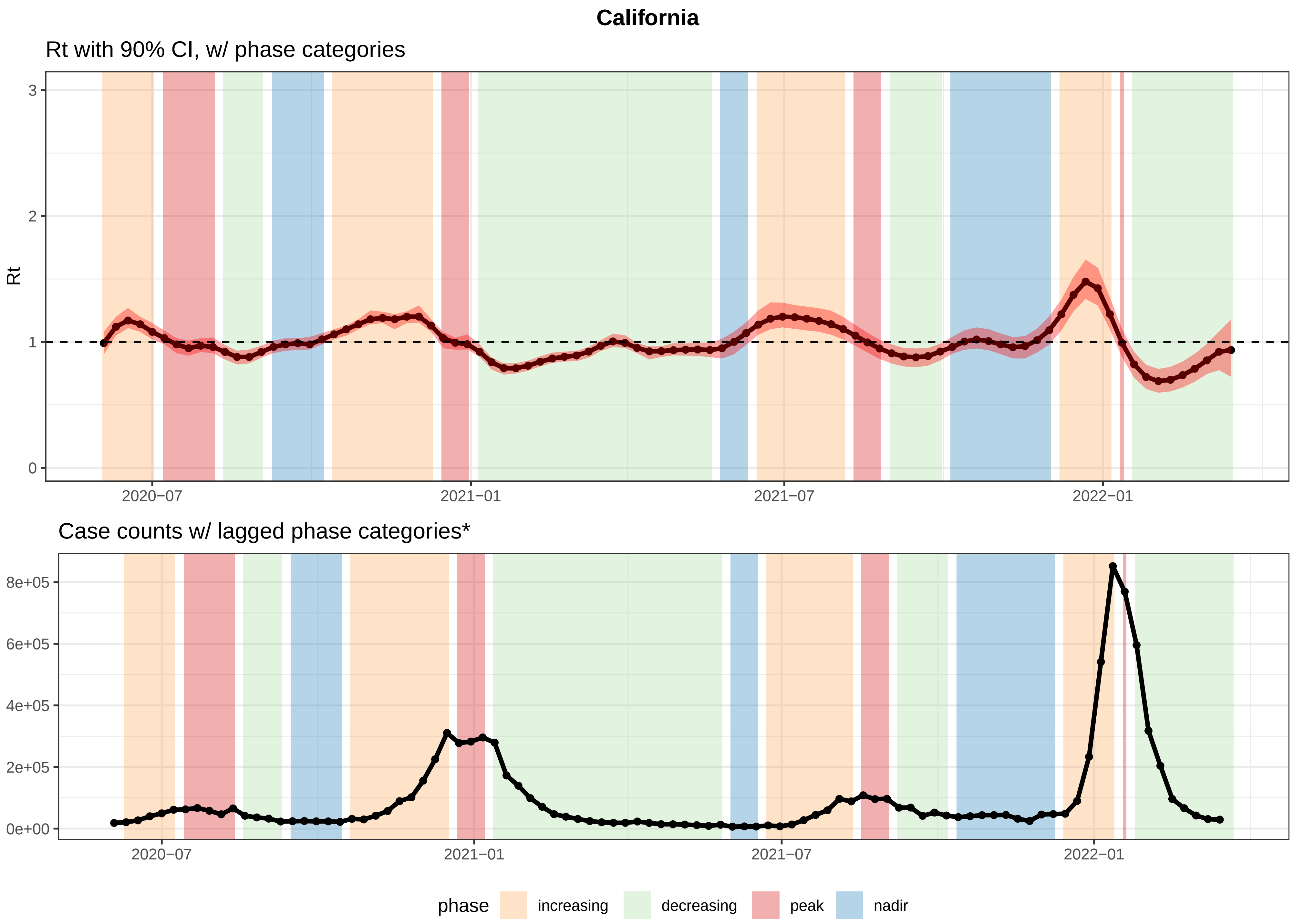


*Increasing/decreasing = Rt had a 90% probability >= or <= than 1.0.
Wks b/w two increasing/decreasing phases −> classified as increasing/decreasing.
Wks b/w increasing and decreasing phases = peaks; nadirs = wks b/w decreasing and increasing phases.

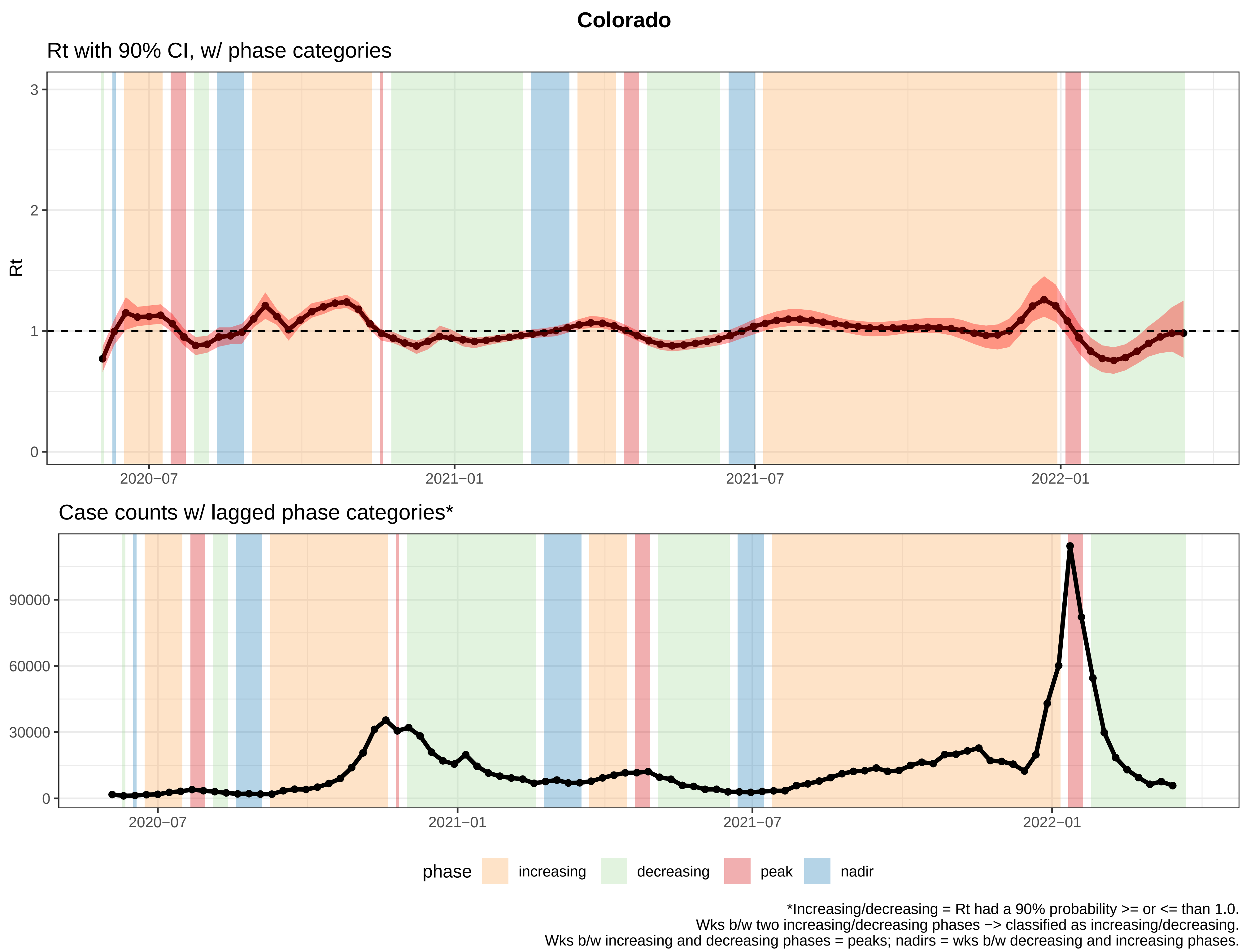
Colorado
Rt with 90% CI, w/ phase categories
Rt
3
2
1
0
2020−07
2021−01
2021−07
2022−01
Case counts w/ lagged phase categories*
90000
60000
30000
0
2020−07
2021−01
2021−07
2022−01
phase
increasing
decreasing
peak
nadir
*Increasing/decreasing = Rt had a 90% probability >= or <= than 1.0.
Wks b/w two increasing/decreasing phases −> classified as increasing/decreasing.
Wks b/w increasing and decreasing phases = peaks; nadirs = wks b/w decreasing and increasing phases.

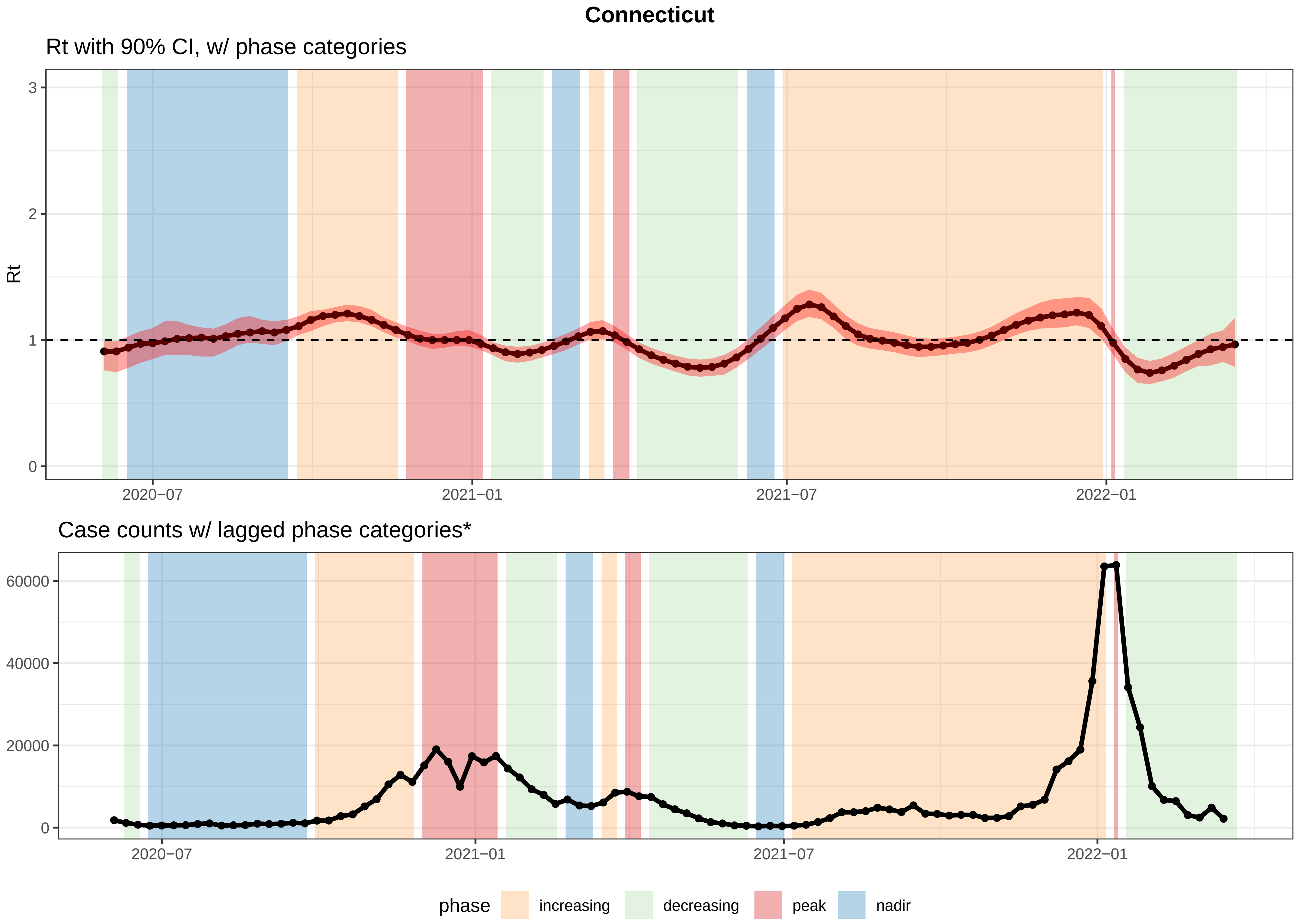


*Increasing/decreasing = Rt had a 90% probability >= or <= than 1.0.
Wks b/w two increasing/decreasing phases −> classified as increasing/decreasing.
Wks b/w increasing and decreasing phases = peaks; nadirs = wks b/w decreasing and increasing phases.

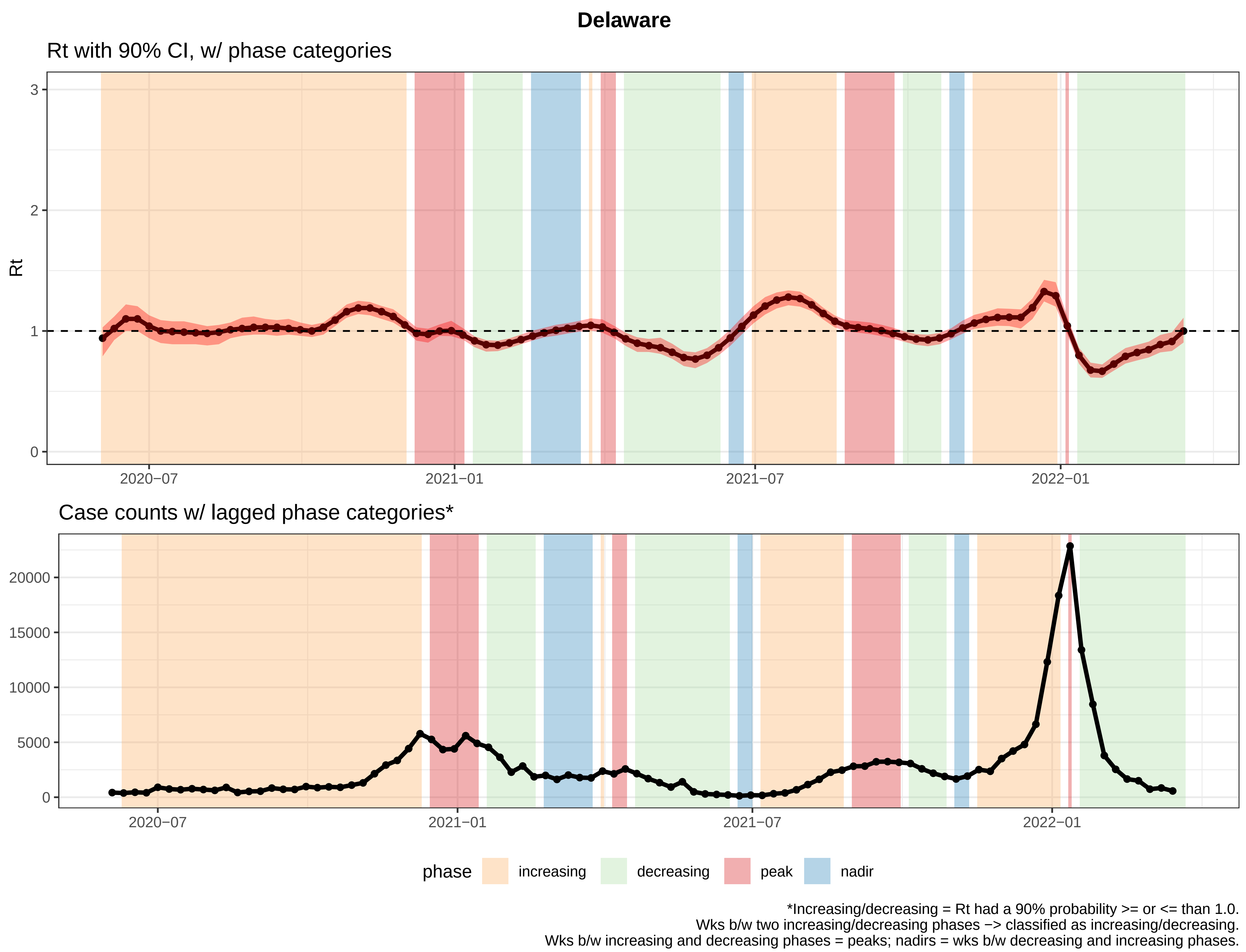

Delaware
Rt with 90% CI, w/ phase categories
Rt
0
1
2
3
2020−07
2021−01
2021−07
2022−01
Case counts w/ lagged phase categories*
0
5000
10000
15000
20000
2020−07
2021−01
2021−07
2022−01
phase
increasing
decreasing
peak
nadir
*Increasing/decreasing = Rt had a 90% probability >= or <= than 1.0.
Wks b/w two increasing/decreasing phases −> classified as increasing/decreasing.
Wks b/w increasing and decreasing phases = peaks; nadirs = wks b/w decreasing and increasing phases.

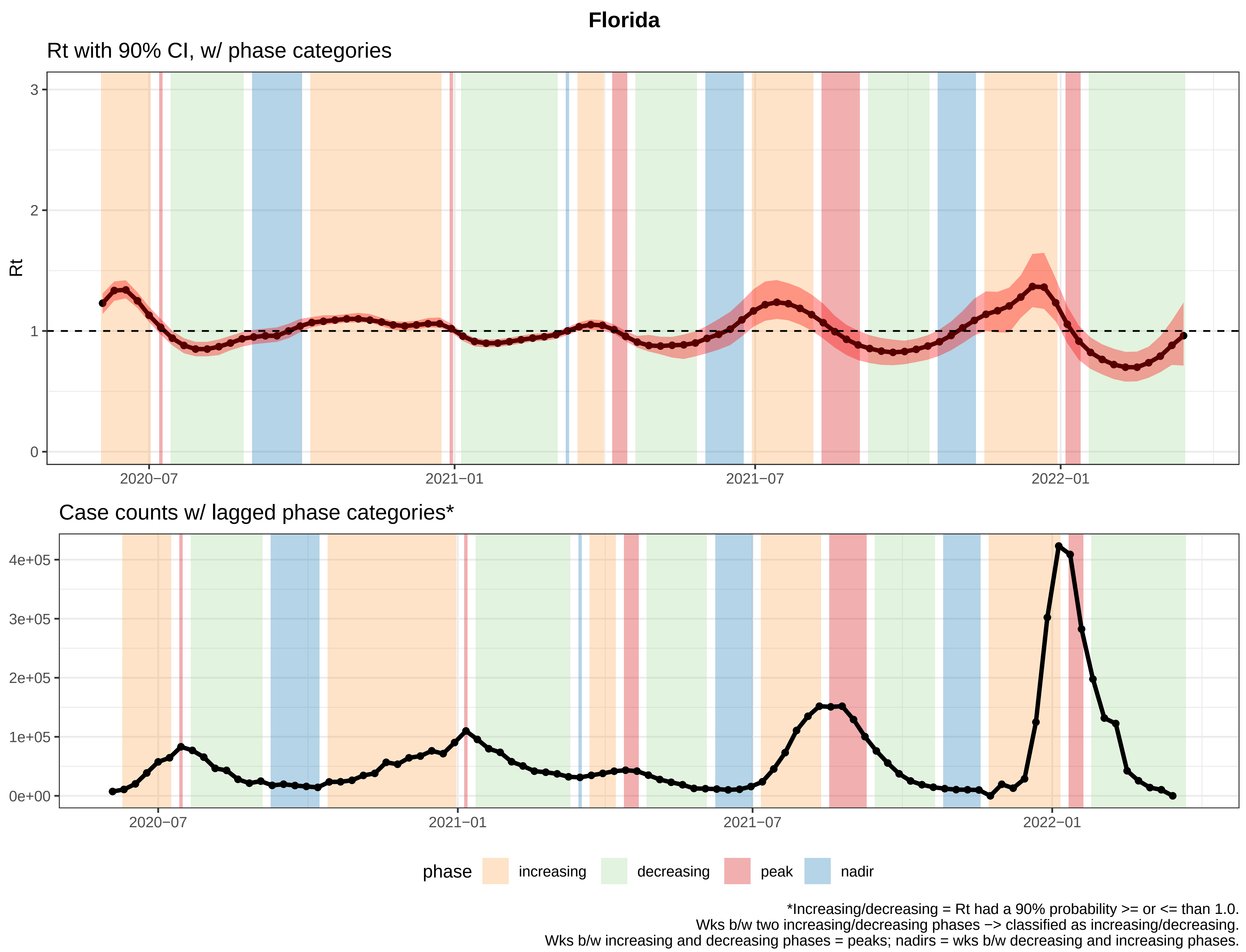
Florida
Rt with 90% CI, w/ phase categories
Rt
3
2
1
0
2020−07
2021−01
2021−07
2022−01
Case counts w/ lagged phase categories*
4e+05
3e+05
2e+05
1e+05
0e+00
2020−07
2021−01
2021−07
2022−01
phase
increasing
decreasing
peak
nadir
*Increasing/decreasing = Rt had a 90% probability >= or <= than 1.0.
Wks b/w two increasing/decreasing phases −> classified as increasing/decreasing.
Wks b/w increasing and decreasing phases = peaks; nadirs = wks b/w decreasing and increasing phases.

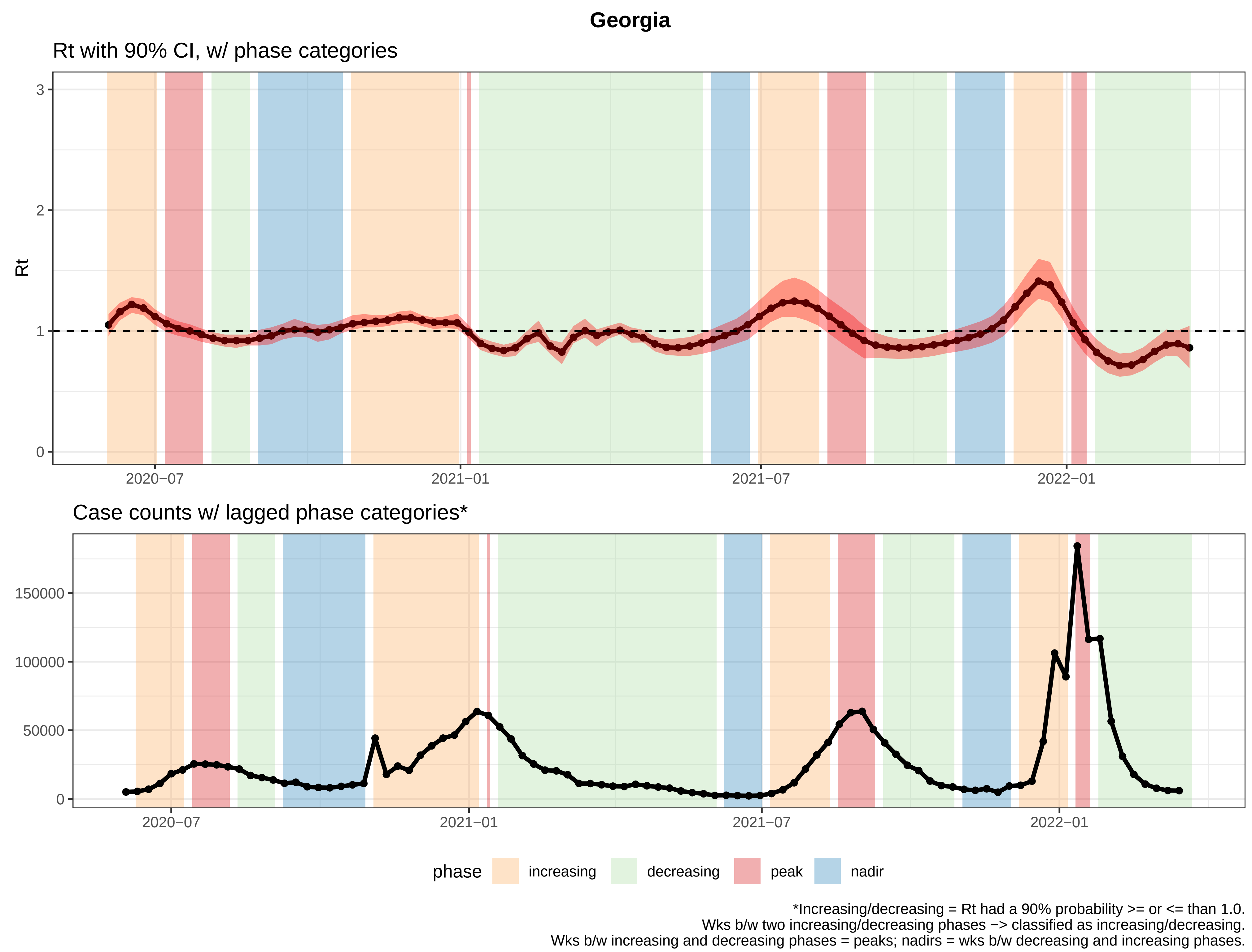

Georgia
Rt with 90% CI, w/ phase categories
Rt
3
2
1
0
2020−07
2021−01
2021−07
2022−01
Case counts w/ lagged phase categories*
150000
100000
50000
0
2020−07
2021−01
2021−07
2022−01
phase
increasing
decreasing
peak
nadir
*Increasing/decreasing = Rt had a 90% probability >= or <= than 1.0.
Wks b/w two increasing/decreasing phases −> classified as increasing/decreasing.
Wks b/w increasing and decreasing phases = peaks; nadirs = wks b/w decreasing and increasing phases.

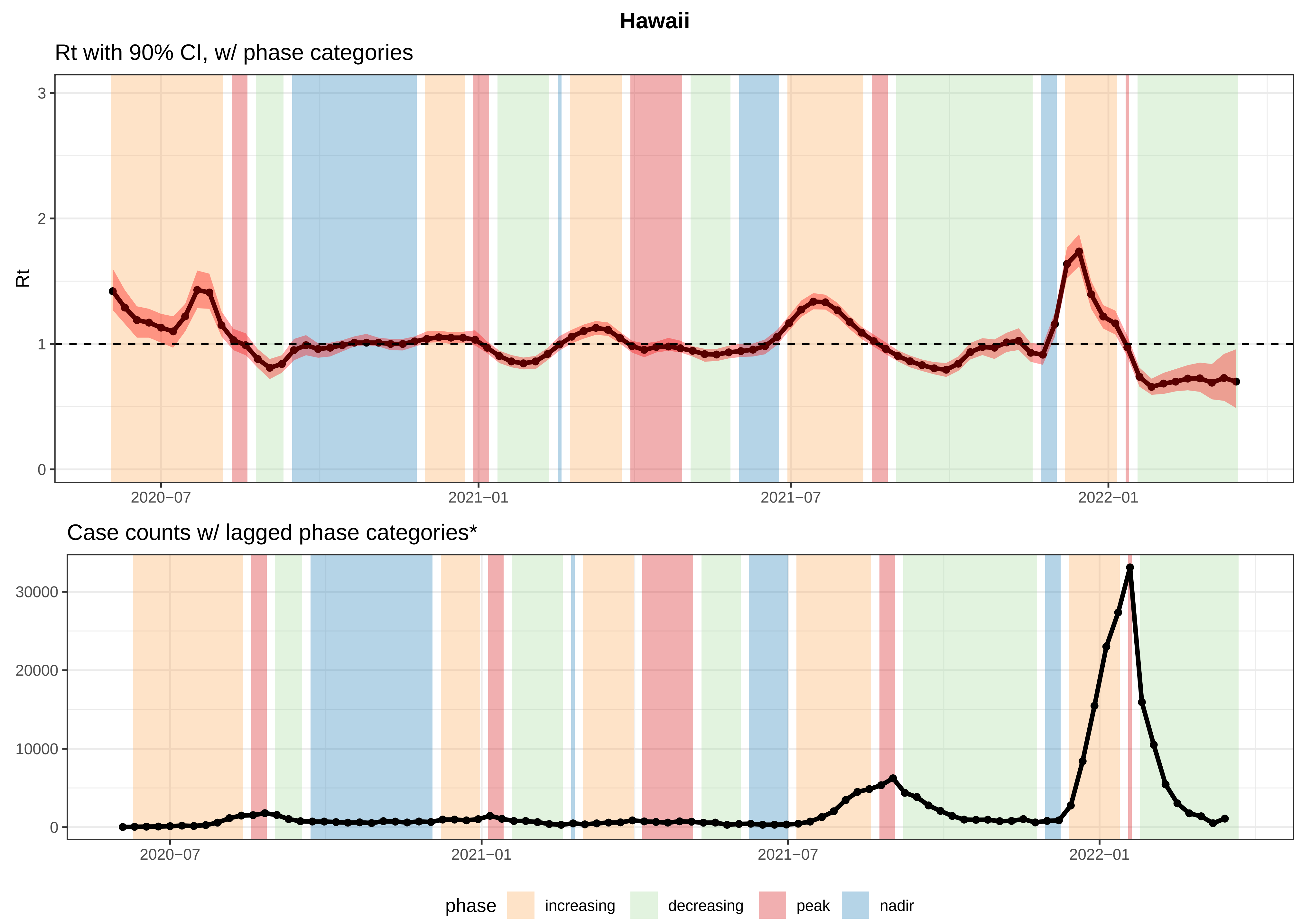


*Increasing/decreasing = Rt had a 90% probability >= or <= than 1.0.
Wks b/w two increasing/decreasing phases −> classified as increasing/decreasing.
Wks b/w increasing and decreasing phases = peaks; nadirs = wks b/w decreasing and increasing phases.

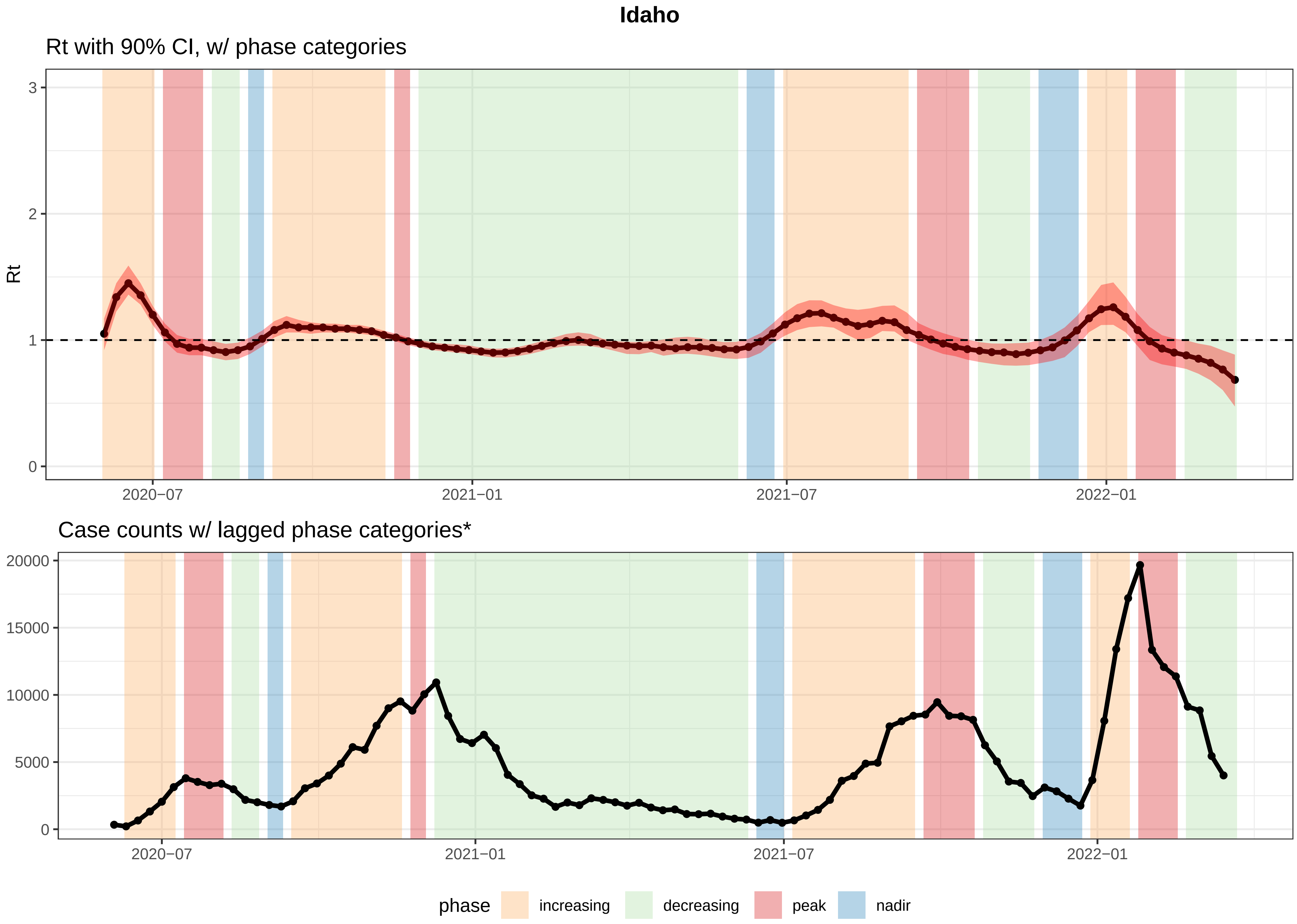


*Increasing/decreasing = Rt had a 90% probability >= or <= than 1.0.
Wks b/w two increasing/decreasing phases −> classified as increasing/decreasing.
Wks b/w increasing and decreasing phases = peaks; nadirs = wks b/w decreasing and increasing phases.

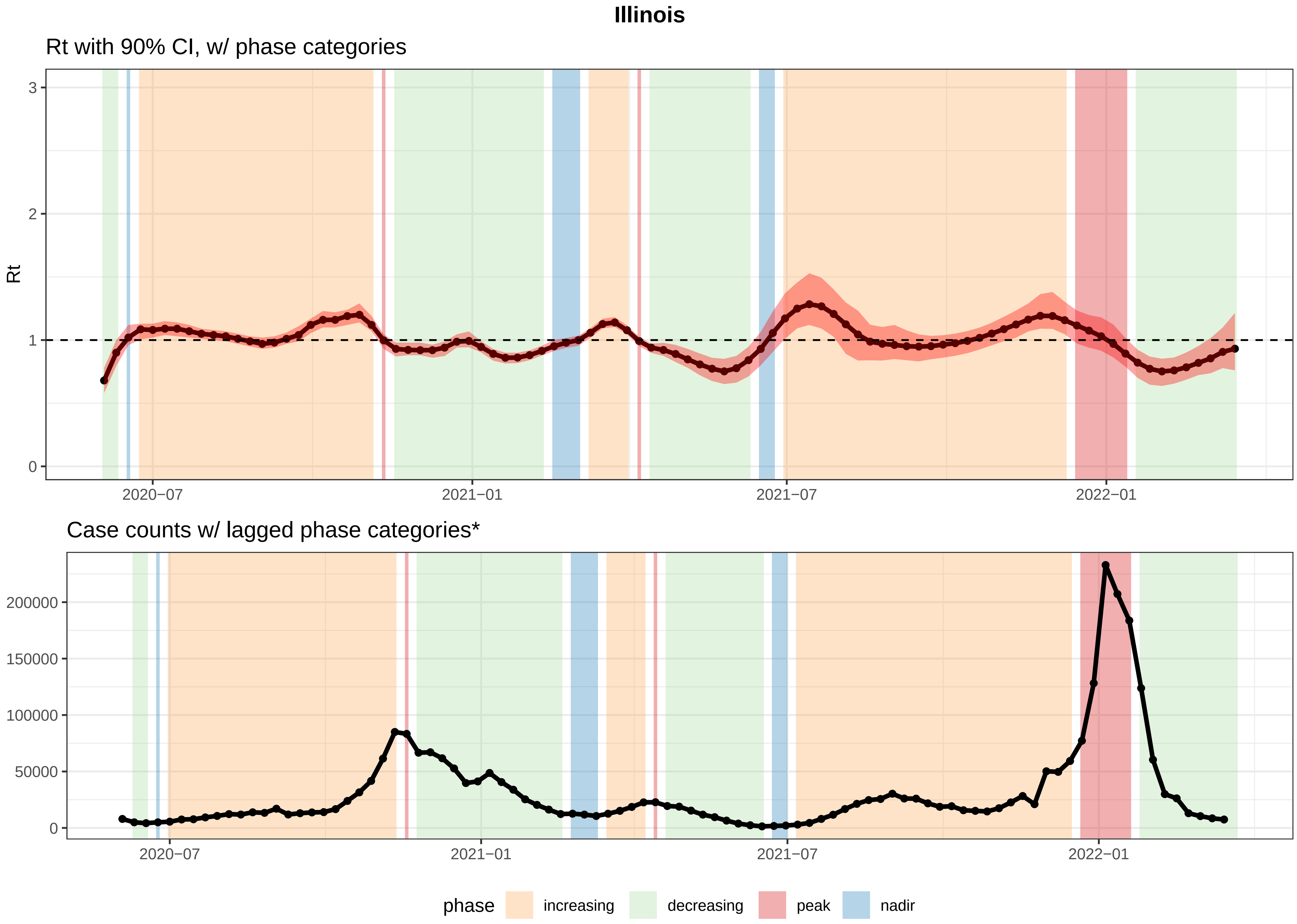


*Increasing/decreasing = Rt had a 90% probability >= or <= than 1.0.
Wks b/w two increasing/decreasing phases −> classified as increasing/decreasing.
Wks b/w increasing and decreasing phases = peaks; nadirs = wks b/w decreasing and increasing phases.

#### Indiana

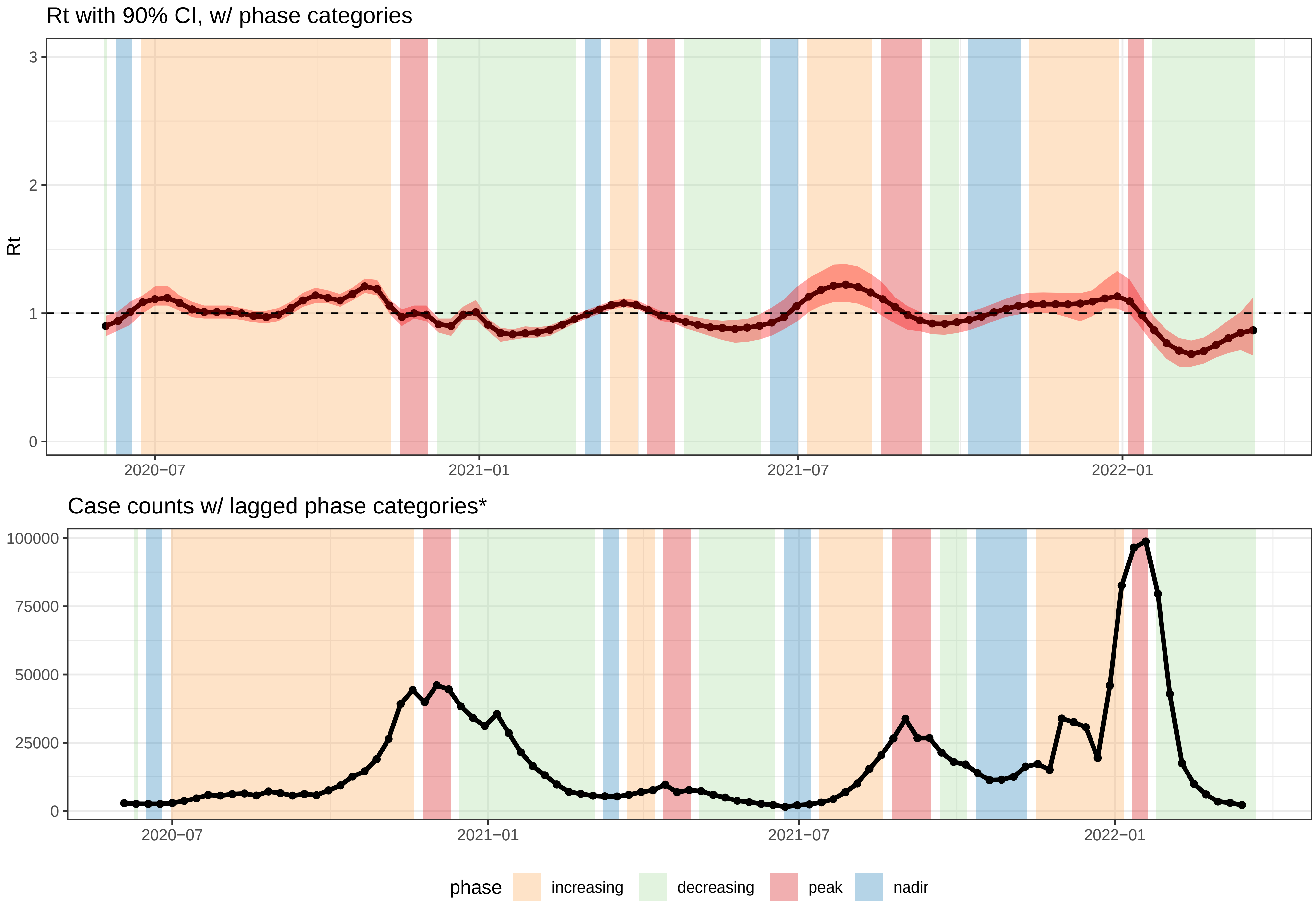


*Increasing/decreasing = Rt had a 90% probability >= or <= than 1.0.
Wks b/w two increasing/decreasing phases −> classified as increasing/decreasing.
Wks b/w increasing and decreasing phases = peaks; nadirs = wks b/w decreasing and increasing phases.

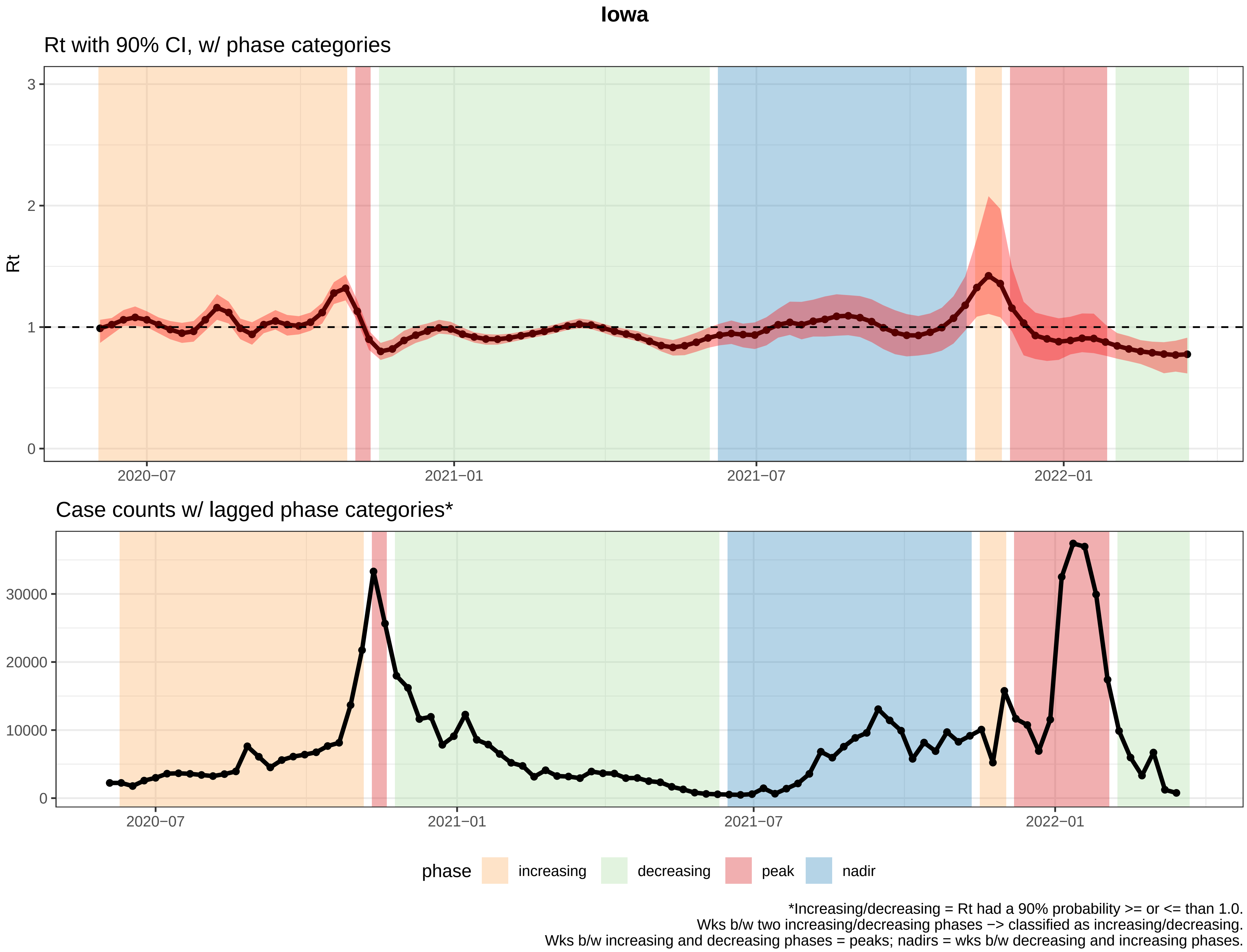
Iowa
Rt with 90% CI, w/ phase categories
Rt
3
2
1
0
2020−07
2021−01
2021−07
2022−01
Case counts w/ lagged phase categories*
30000
20000
10000
0
2020−07
2021−01
2021−07
2022−01
phase
increasing
decreasing
peak
nadir
*Increasing/decreasing = Rt had a 90% probability >= or <= than 1.0.
Wks b/w two increasing/decreasing phases −> classified as increasing/decreasing.
Wks b/w increasing and decreasing phases = peaks; nadirs = wks b/w decreasing and increasing phases.

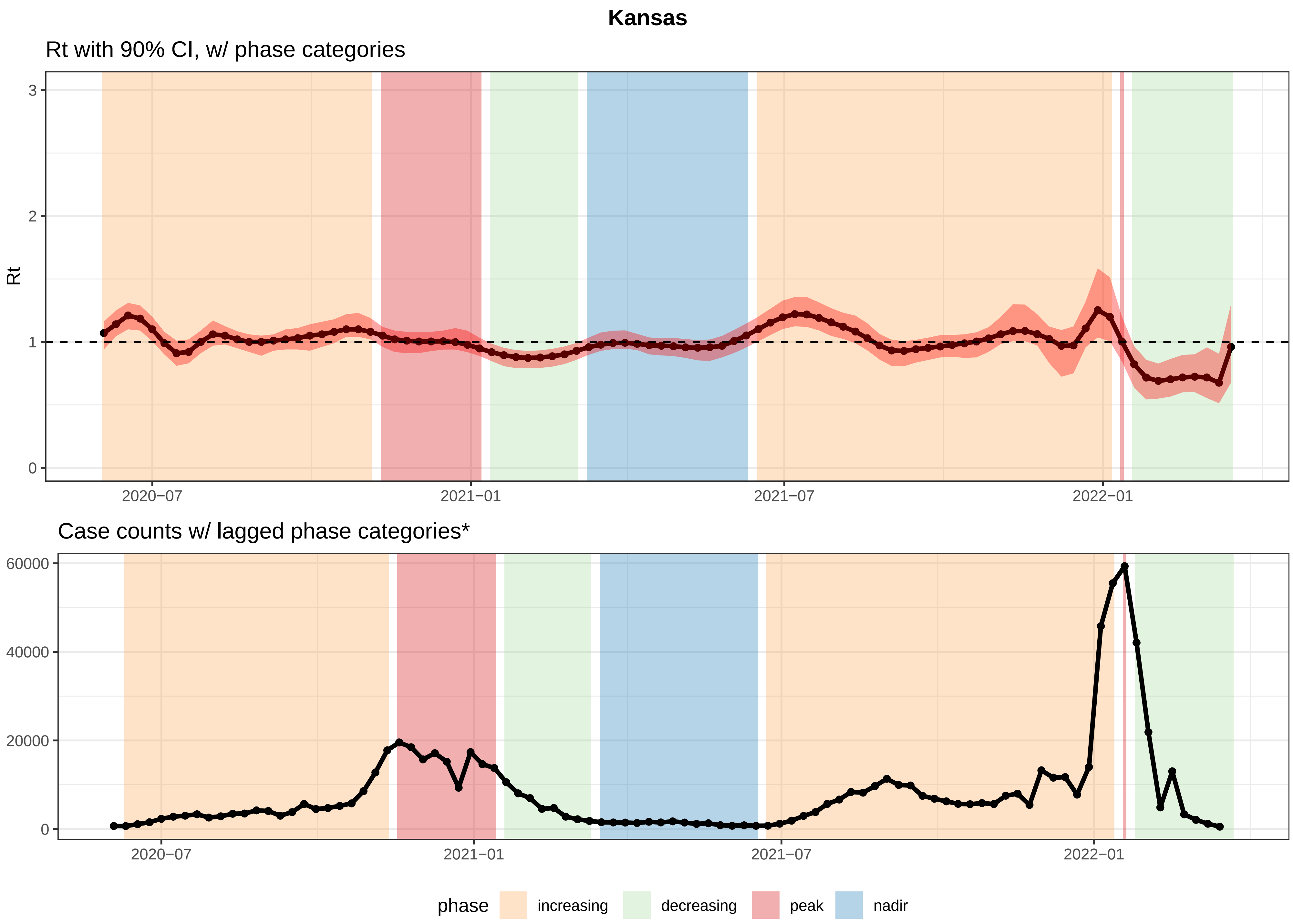


*Increasing/decreasing = Rt had a 90% probability >= or <= than 1.0.
Wks b/w two increasing/decreasing phases −> classified as increasing/decreasing.
Wks b/w increasing and decreasing phases = peaks; nadirs = wks b/w decreasing and increasing phases.

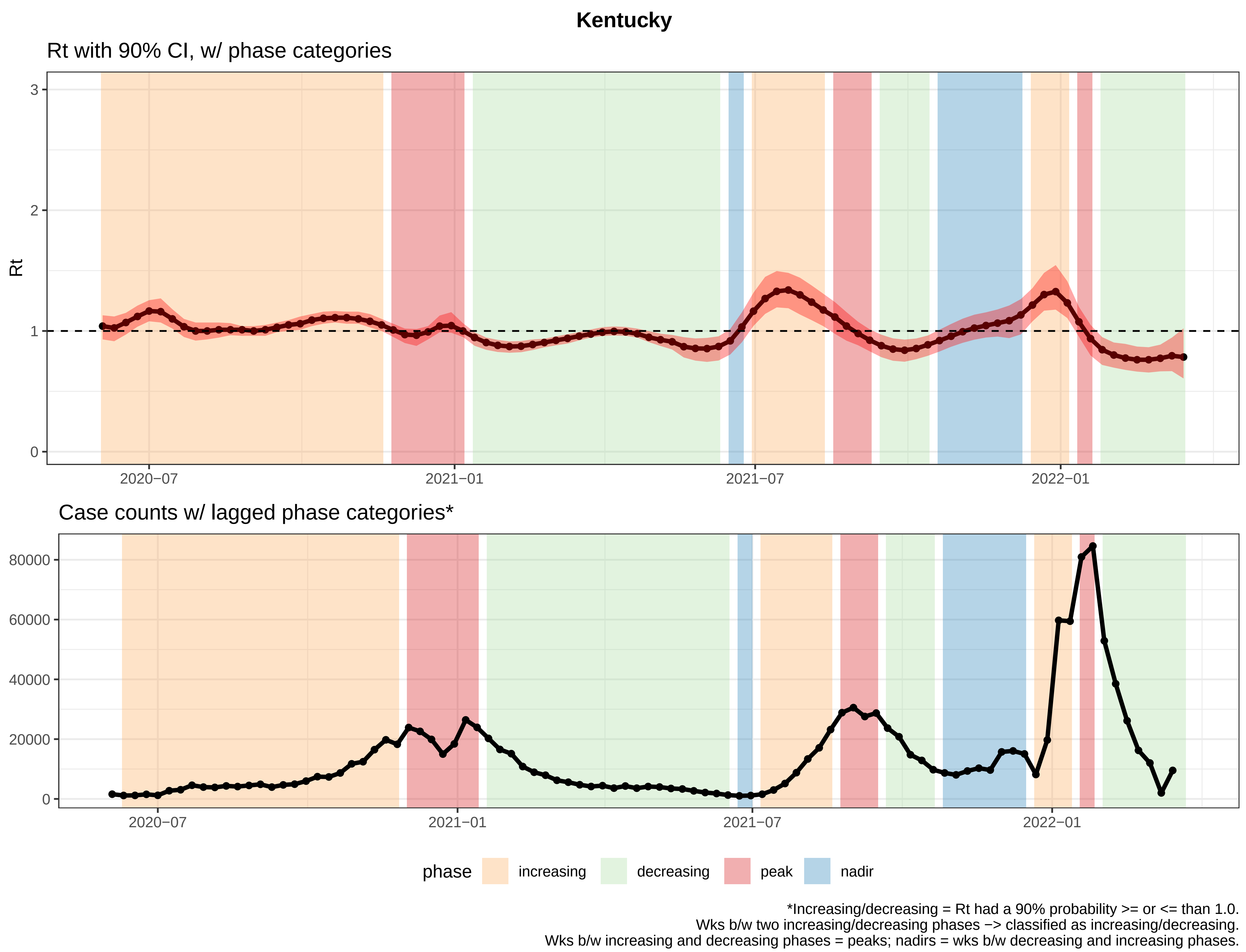
Kentucky
Rt with 90% CI, w/ phase categories
Rt
3
2
1
0
2020−07
2021−01
2021−07
2022−01
Case counts w/ lagged phase categories*
80000
60000
40000
20000
0
2020−07
2021−01
2021−07
2022−01
phase
increasing
decreasing
peak
nadir
*Increasing/decreasing = Rt had a 90% probability >= or <= than 1.0.
Wks b/w two increasing/decreasing phases −> classified as increasing/decreasing.
Wks b/w increasing and decreasing phases = peaks; nadirs = wks b/w decreasing and increasing phases.

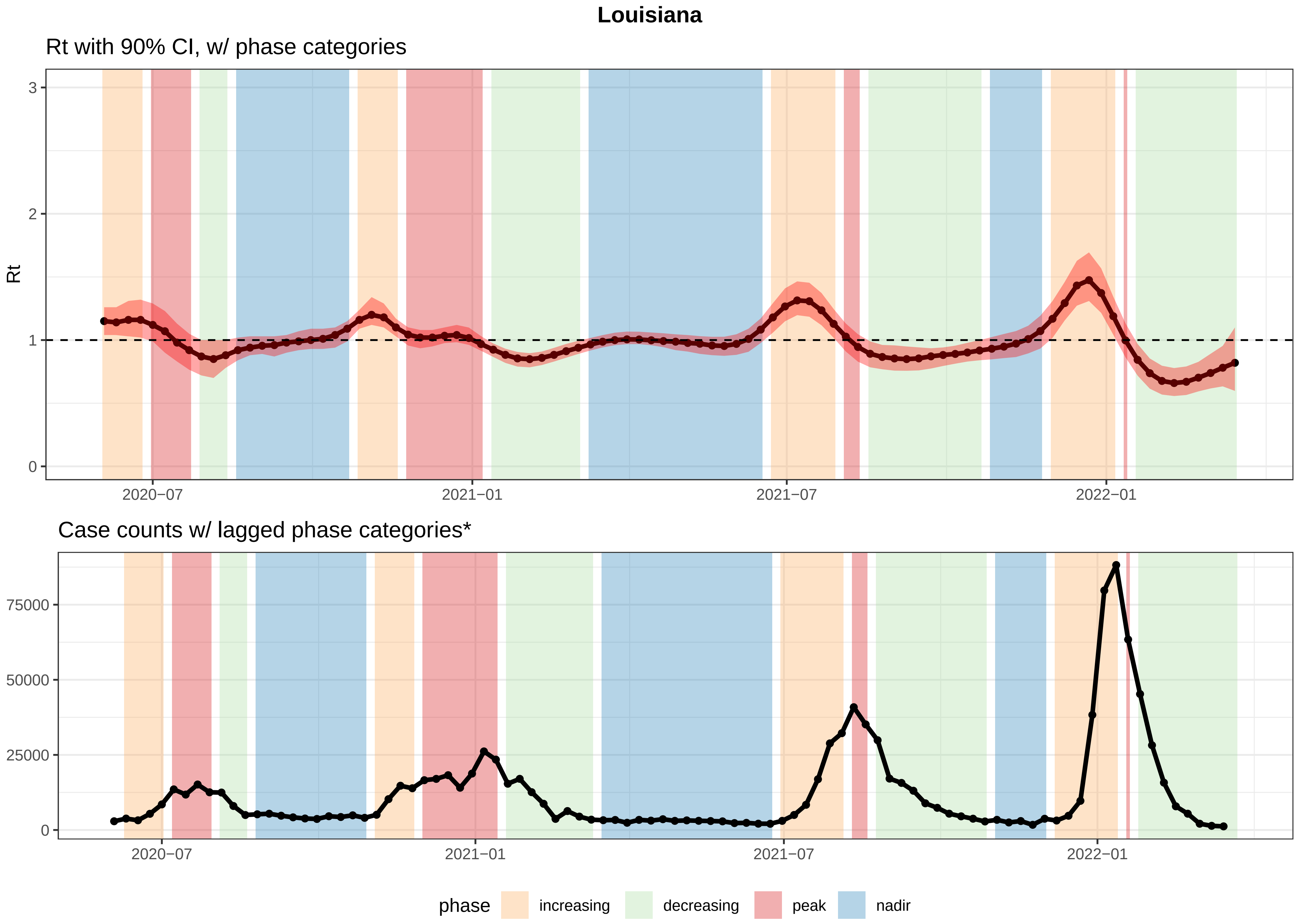


*Increasing/decreasing = Rt had a 90% probability >= or <= than 1.0.
Wks b/w two increasing/decreasing phases −> classified as increasing/decreasing.
Wks b/w increasing and decreasing phases = peaks; nadirs = wks b/w decreasing and increasing phases.

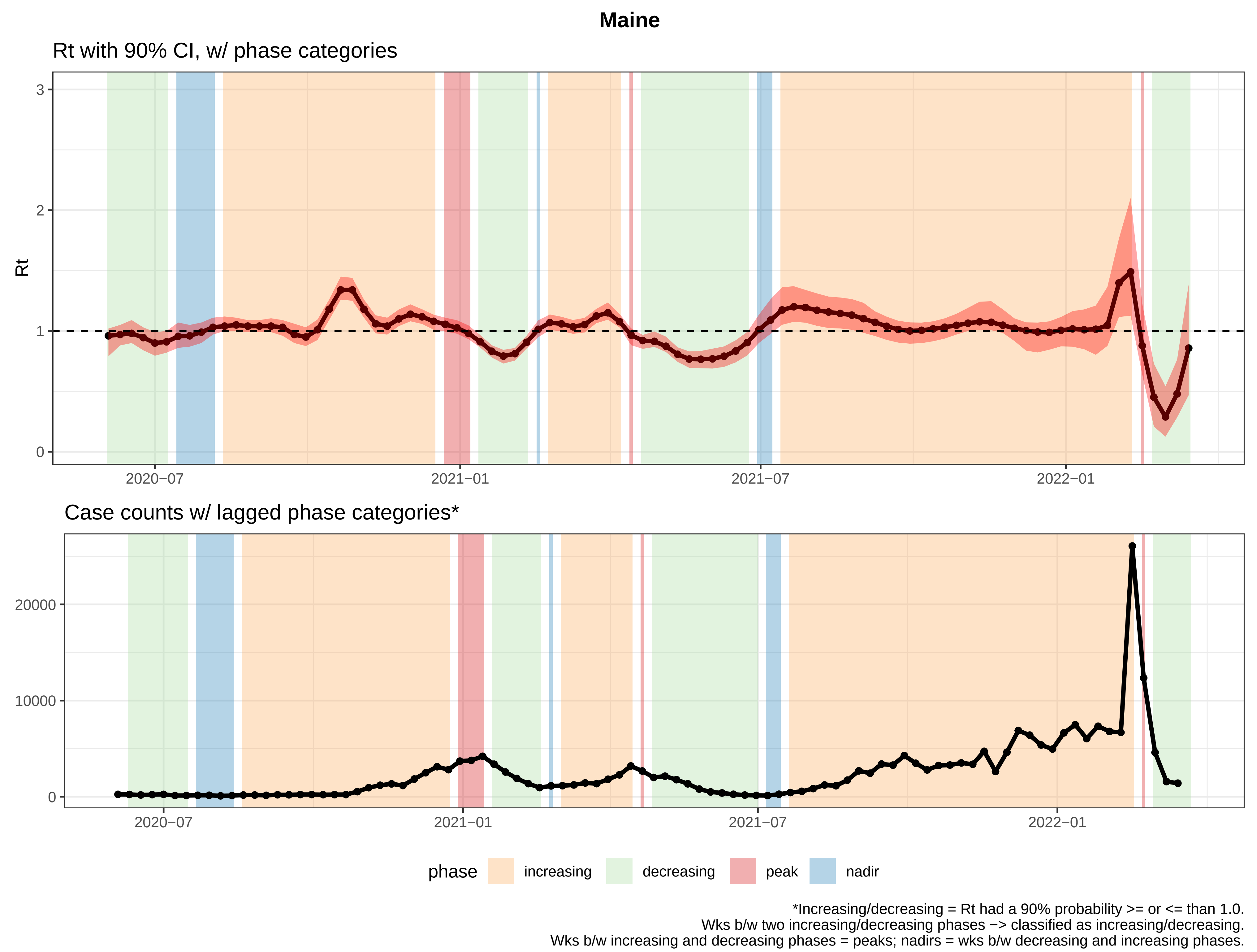
Maine
Rt with 90% CI, w/ phase categories
Rt
3
2
1
0
2020−07
2021−01
2021−07
2022−01
Case counts w/ lagged phase categories*
20000
10000
0
2020−07
2021−01
2021−07
2022−01
phase
increasing
decreasing
peak
nadir
*Increasing/decreasing = Rt had a 90% probability >= or <= than 1.0.
Wks b/w two increasing/decreasing phases −> classified as increasing/decreasing.
Wks b/w increasing and decreasing phases = peaks; nadirs = wks b/w decreasing and increasing phases.

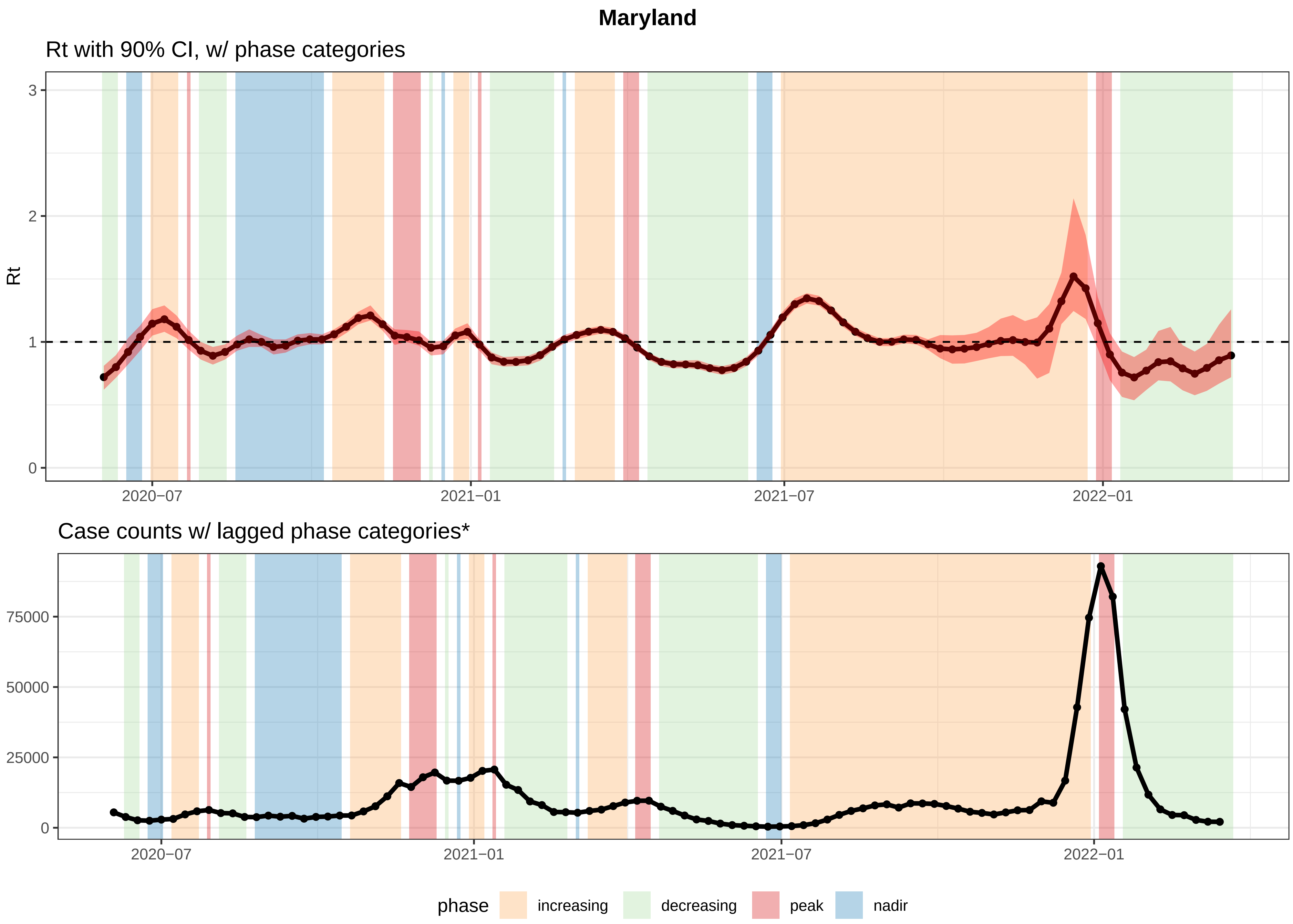


*Increasing/decreasing = Rt had a 90% probability >= or <= than 1.0.
Wks b/w two increasing/decreasing phases −> classified as increasing/decreasing.
Wks b/w increasing and decreasing phases = peaks; nadirs = wks b/w decreasing and increasing phases.

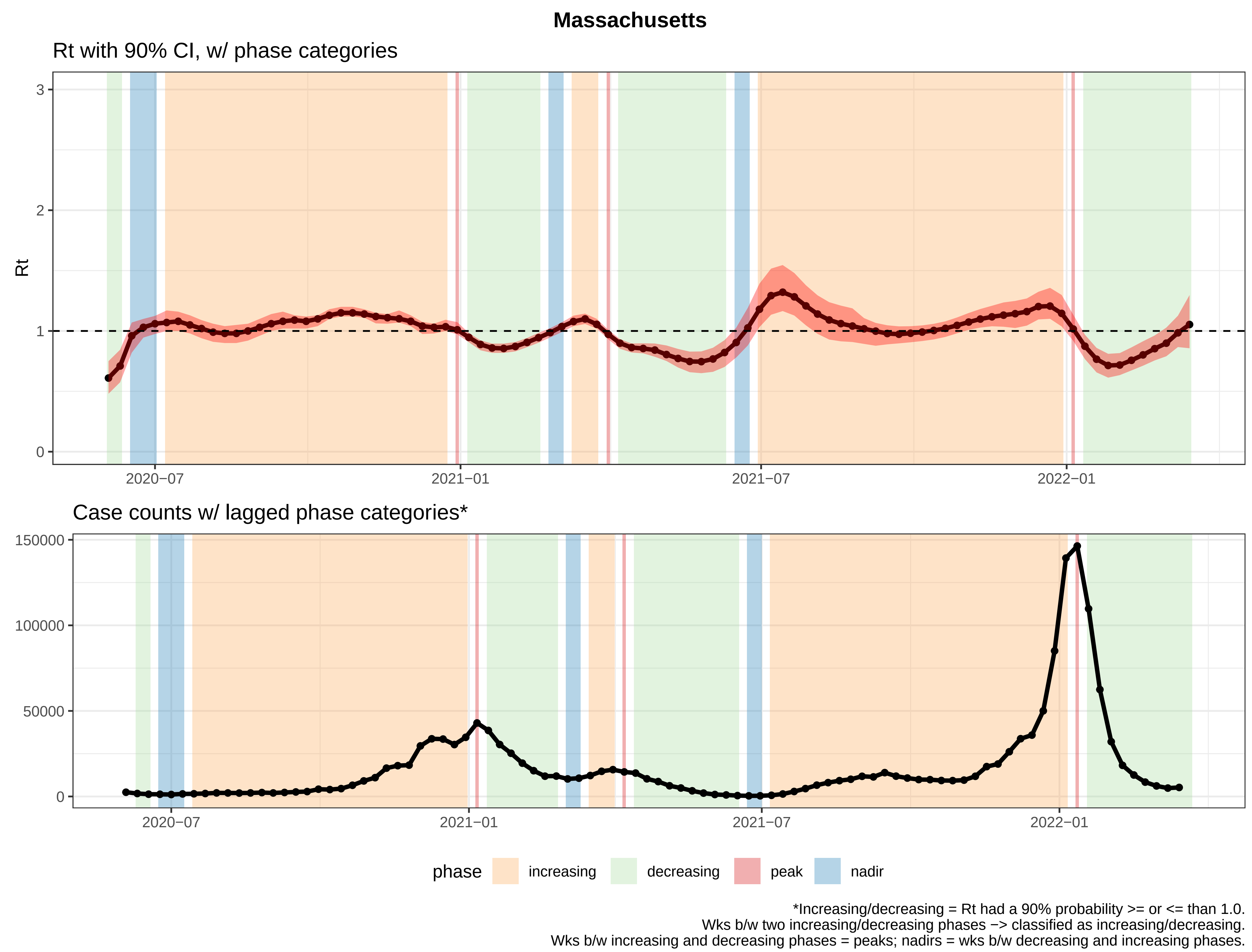
Massachusetts
Rt with 90% CI, w/ phase categories
Rt
0
1
2
3
2020−07
2021−01
2021−07
2022−01
Case counts w/ lagged phase categories*
0
50000
100000
150000
2020−07
2021−01
2021−07
2022−01
phase
increasing
decreasing
peak
nadir
*Increasing/decreasing = Rt had a 90% probability >= or <= than 1.0.
Wks b/w two increasing/decreasing phases −> classified as increasing/decreasing.
Wks b/w increasing and decreasing phases = peaks; nadirs = wks b/w decreasing and increasing phases.

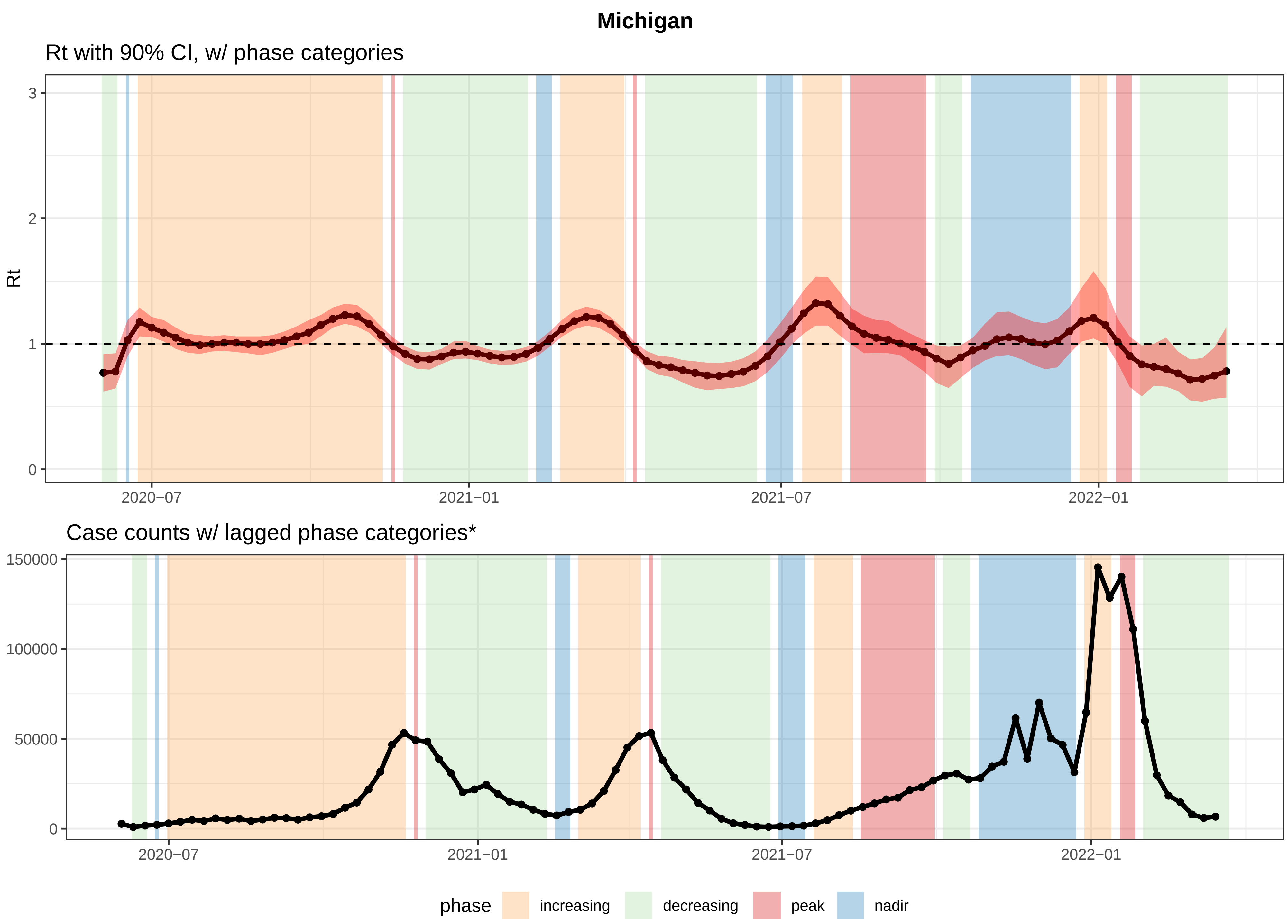


*Increasing/decreasing = Rt had a 90% probability >= or <= than 1.0.
Wks b/w two increasing/decreasing phases −> classified as increasing/decreasing.
Wks b/w increasing and decreasing phases = peaks; nadirs = wks b/w decreasing and increasing phases.

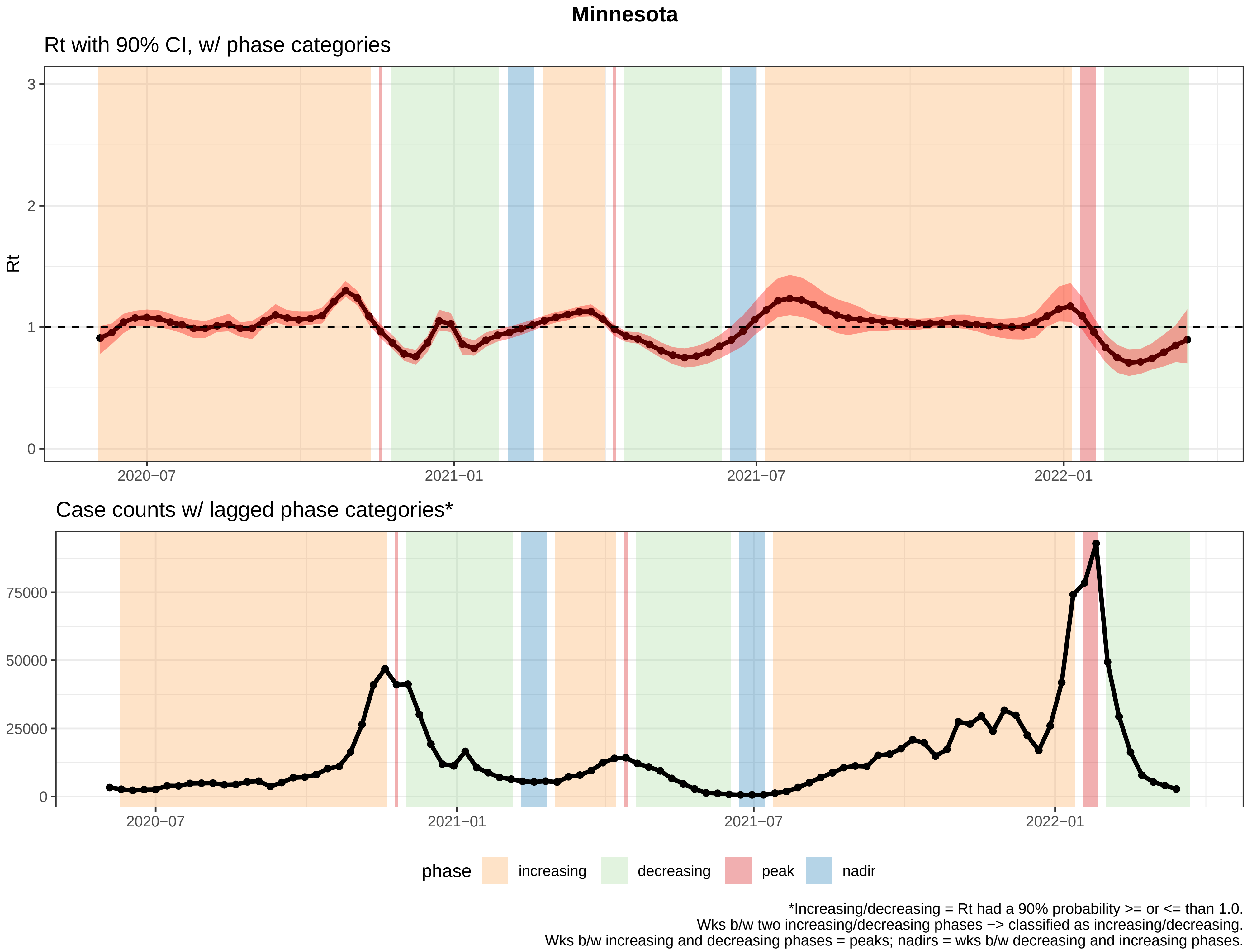

Minnesota
Rt with 90% CI, w/ phase categories
Rt
3
2
1
0
2020−07
2021−01
2021−07
2022−01
Case counts w/ lagged phase categories*
75000
50000
25000
0
2020−07
2021−01
2021−07
2022−01
phase
increasing
decreasing
peak
nadir
*Increasing/decreasing = Rt had a 90% probability >= or <= than 1.0.
Wks b/w two increasing/decreasing phases −> classified as increasing/decreasing.
Wks b/w increasing and decreasing phases = peaks; nadirs = wks b/w decreasing and increasing phases.

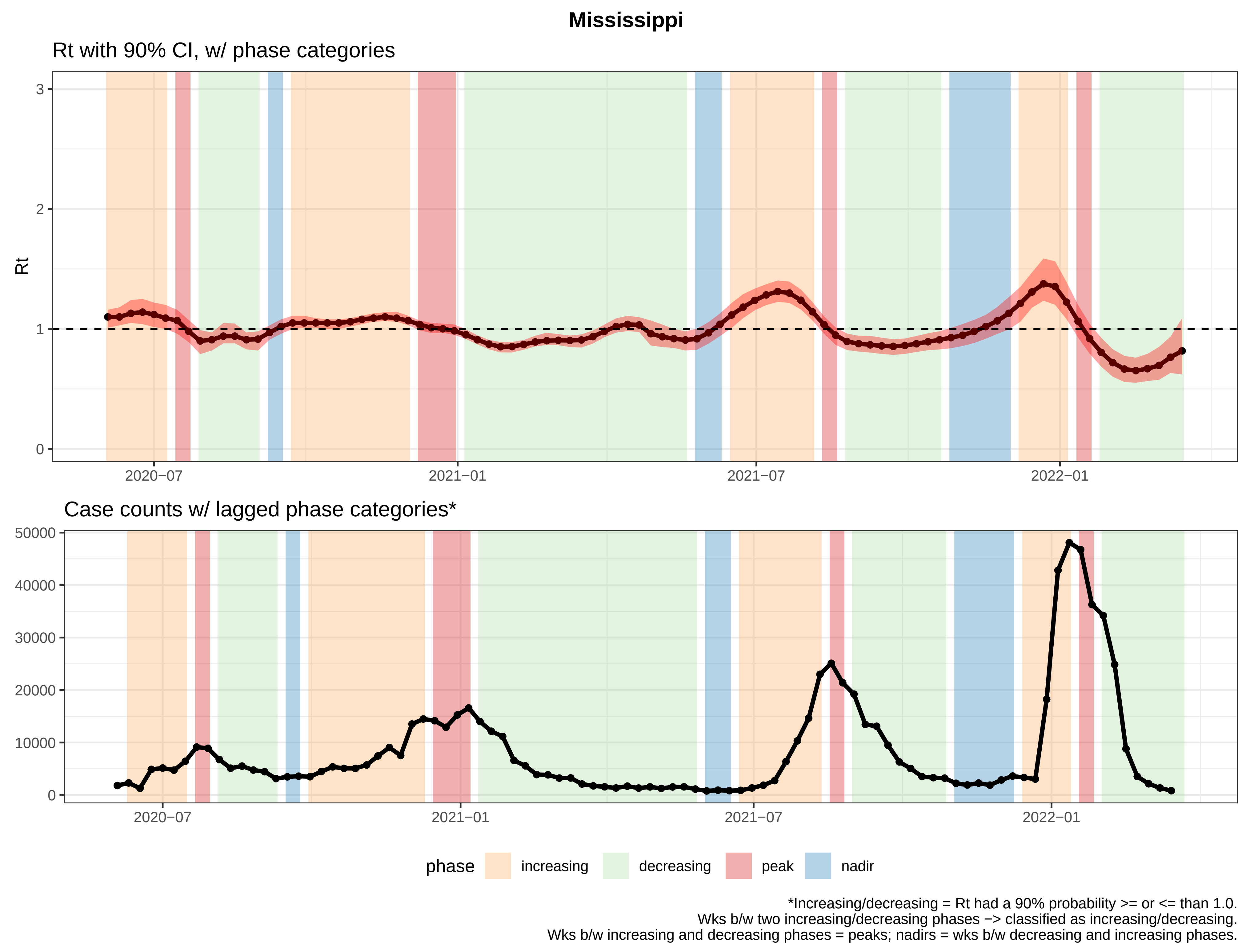

Mississippi
Rt with 90% CI, w/ phase categories
Rt
0
1
2
3
2020−07
2021−01
2021−07
2022−01
Case counts w/ lagged phase categories*
0
10000
20000
30000
40000
50000
2020−07
2021−01
2021−07
2022−01
phase
increasing
decreasing
peak
nadir
*Increasing/decreasing = Rt had a 90% probability >= or <= than 1.0.
Wks b/w two increasing/decreasing phases −> classified as increasing/decreasing.
Wks b/w increasing and decreasing phases = peaks; nadirs = wks b/w decreasing and increasing phases.

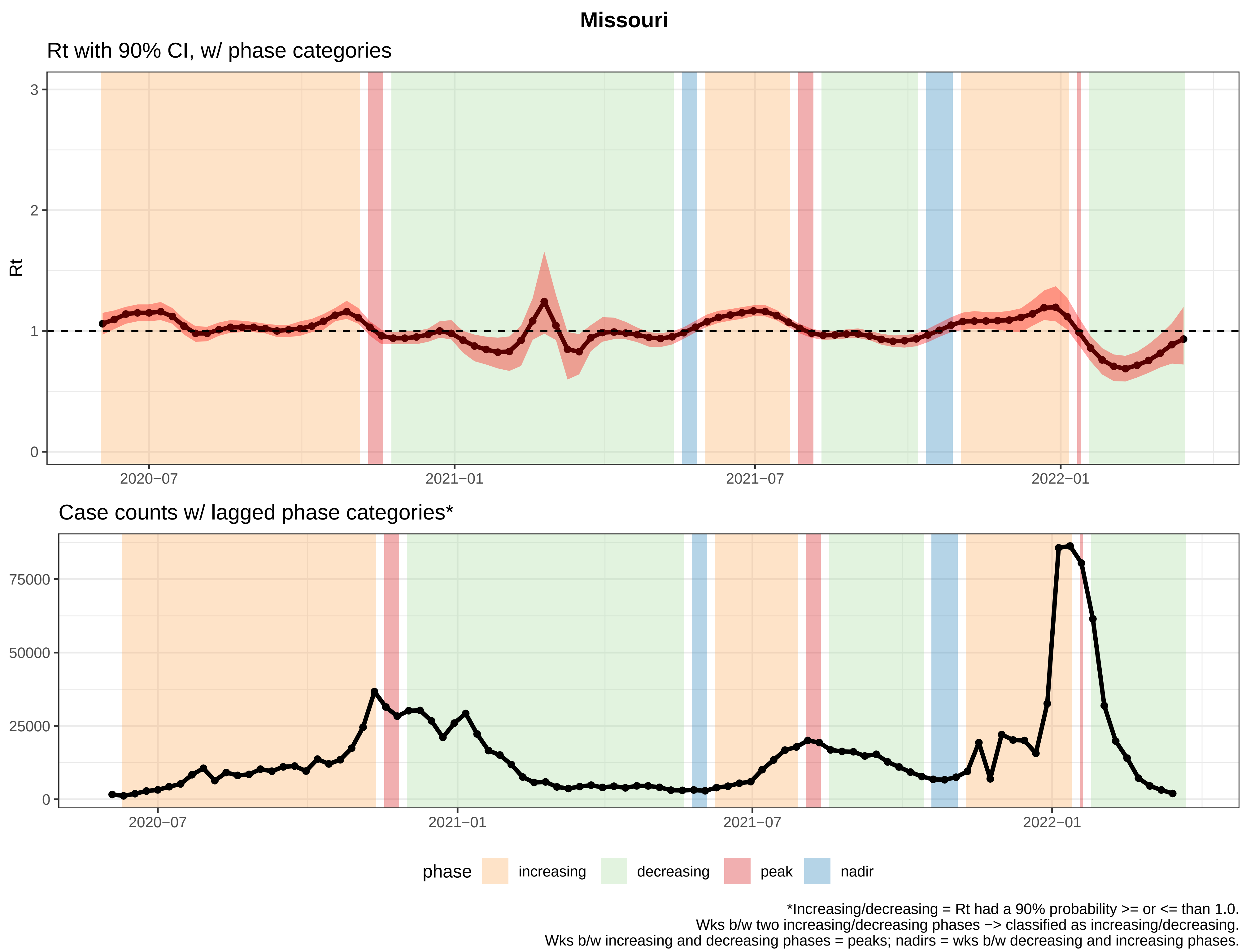

Missouri
Rt with 90% CI, w/ phase categories
Rt
3
2
1
0
2020−07
2021−01
2021−07
2022−01
Case counts w/ lagged phase categories*
75000
50000
25000
0
2020−07
2021−01
2021−07
2022−01
phase
increasing
decreasing
peak
nadir
*Increasing/decreasing = Rt had a 90% probability >= or <= than 1.0.
Wks b/w two increasing/decreasing phases −> classified as increasing/decreasing.
Wks b/w increasing and decreasing phases = peaks; nadirs = wks b/w decreasing and increasing phases.

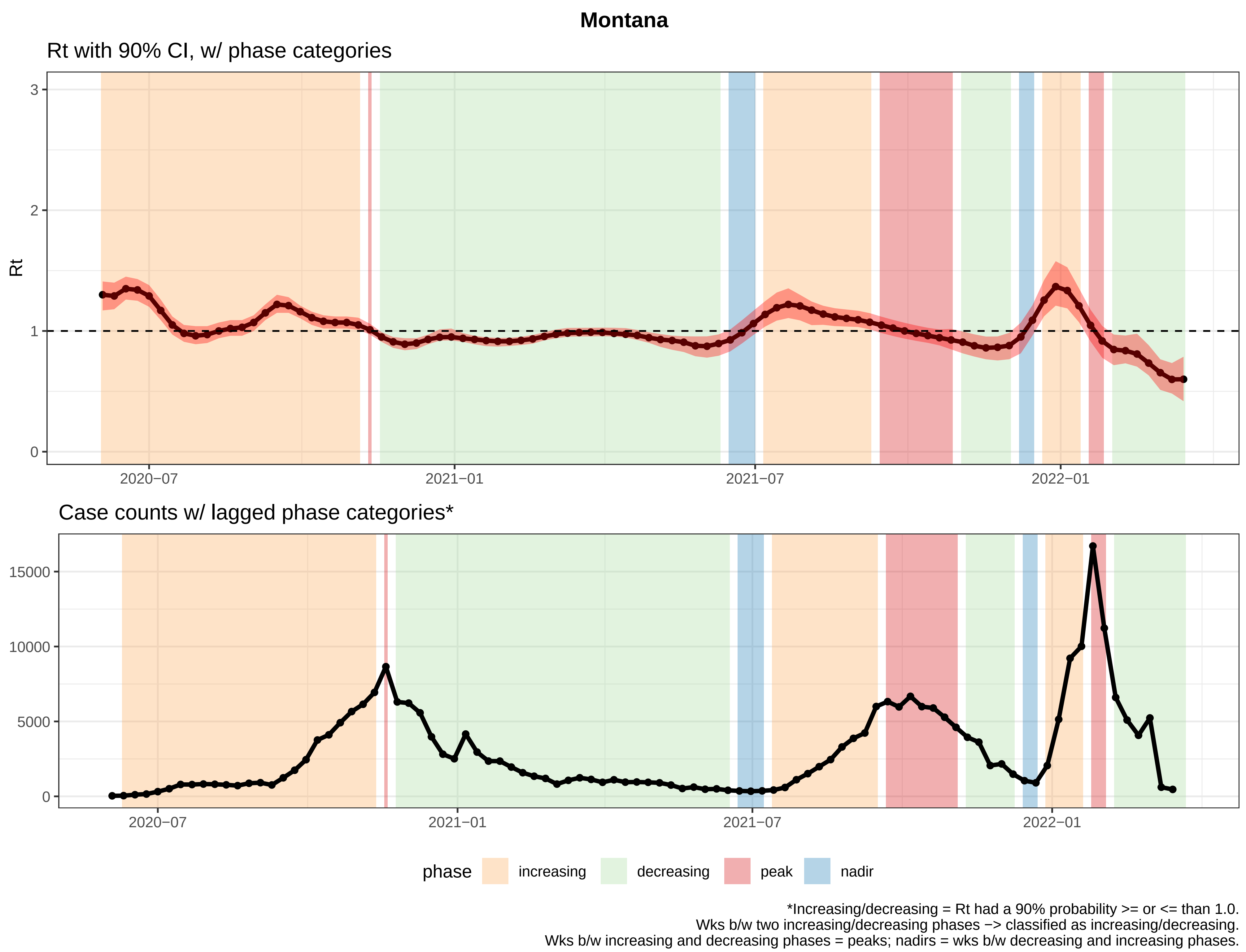

Montana
Rt with 90% CI, w/ phase categories
Rt
3
2
1
0
2020−07
2021−01
2021−07
2022−01
Case counts w/ lagged phase categories*
15000
10000
5000
0
2020−07
2021−01
2021−07
2022−01
phase
increasing
decreasing
peak
nadir
*Increasing/decreasing = Rt had a 90% probability >= or <= than 1.0.
Wks b/w two increasing/decreasing phases −> classified as increasing/decreasing.
Wks b/w increasing and decreasing phases = peaks; nadirs = wks b/w decreasing and increasing phases.

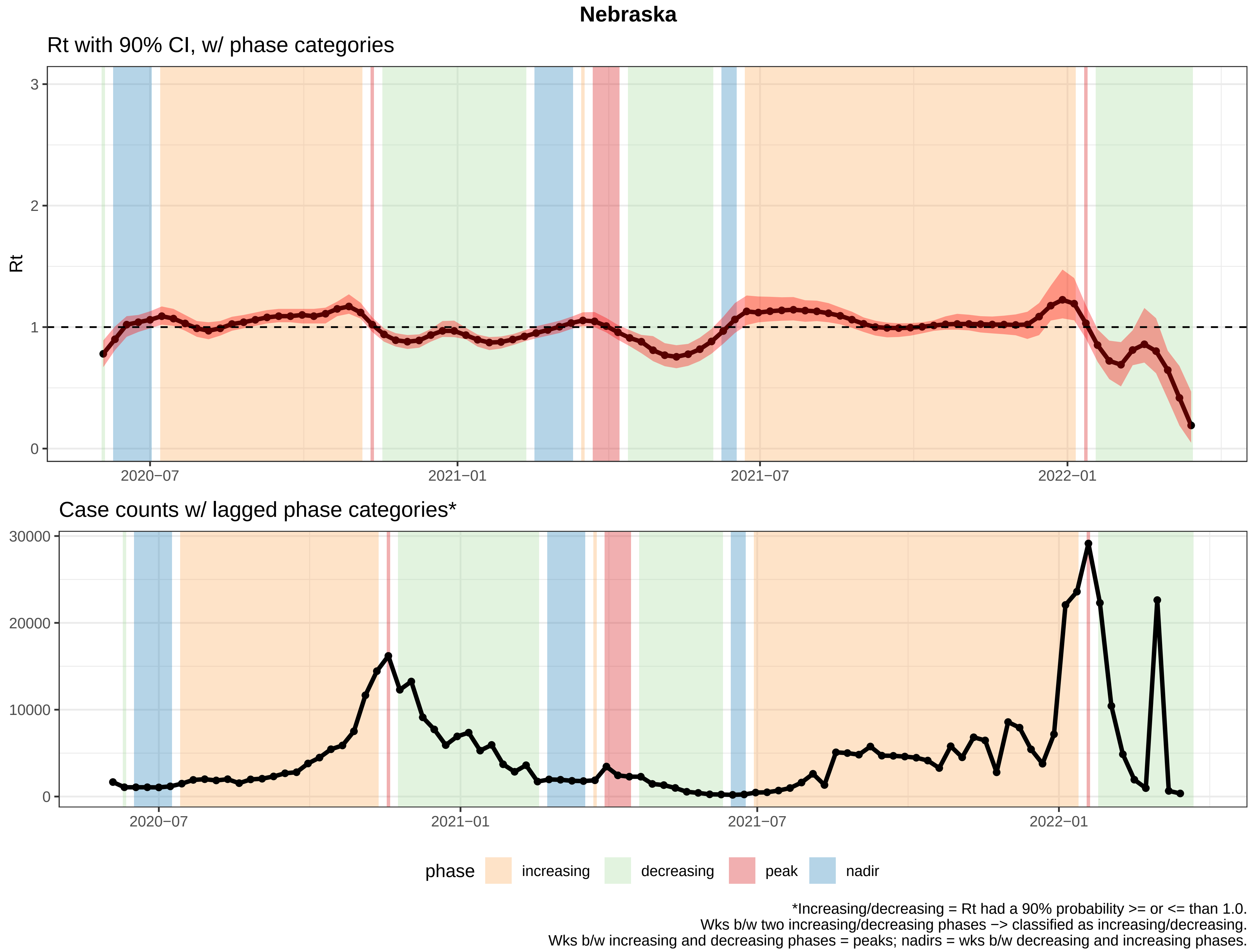

Nebraska
Rt with 90% CI, w/ phase categories
Rt
0
1
2
3
2020−07
2021−01
2021−07
2022−01
Case counts w/ lagged phase categories*
0
10000
20000
30000
2020−07
2021−01
2021−07
2022−01
phase
increasing
decreasing
peak
nadir
*Increasing/decreasing = Rt had a 90% probability >= or <= than 1.0.
Wks b/w two increasing/decreasing phases −> classified as increasing/decreasing.
Wks b/w increasing and decreasing phases = peaks; nadirs = wks b/w decreasing and increasing phases.

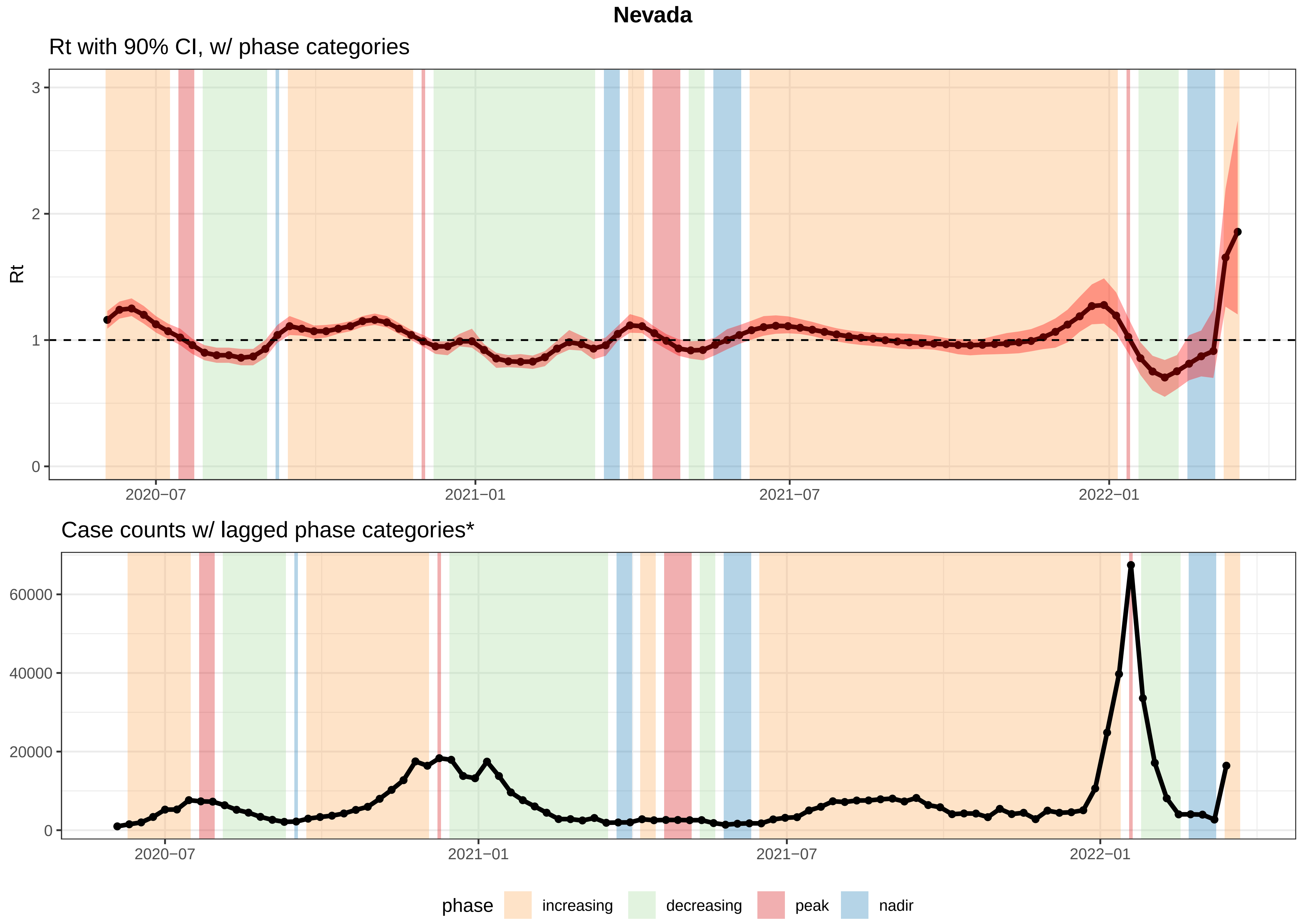


*Increasing/decreasing = Rt had a 90% probability >= or <= than 1.0.
Wks b/w two increasing/decreasing phases −> classified as increasing/decreasing.
Wks b/w increasing and decreasing phases = peaks; nadirs = wks b/w decreasing and increasing phases.

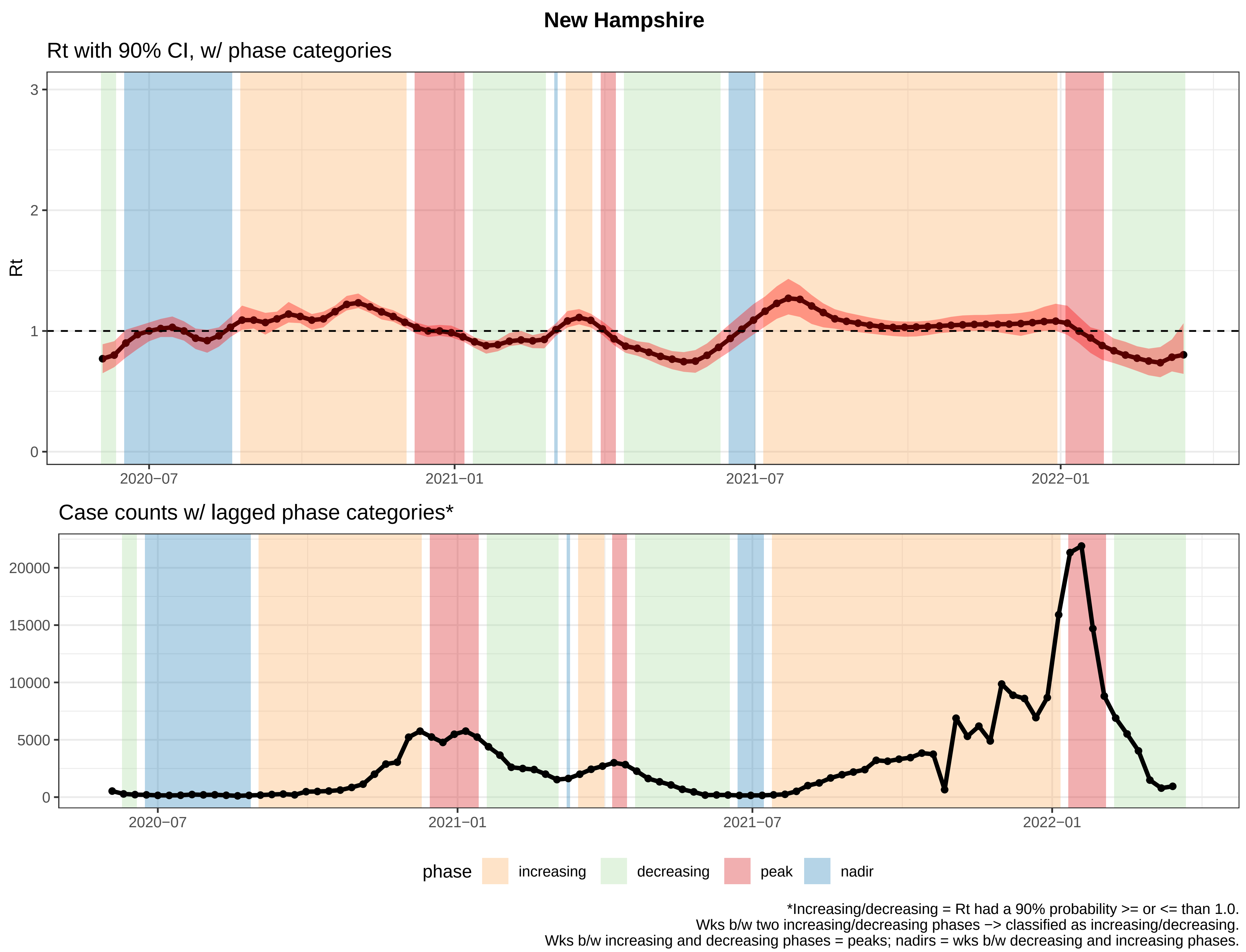
New Hampshire
Rt with 90% CI, w/ phase categories
Rt
3
2
1
0
2020−07
2021−01
2021−07
2022−01
Case counts w/ lagged phase categories*
20000
15000
10000
5000
0
2020−07
2021−01
2021−07
2022−01
phase
increasing
decreasing
peak
nadir
*Increasing/decreasing = Rt had a 90% probability >= or <= than 1.0.
Wks b/w two increasing/decreasing phases −> classified as increasing/decreasing.
Wks b/w increasing and decreasing phases = peaks; nadirs = wks b/w decreasing and increasing phases.

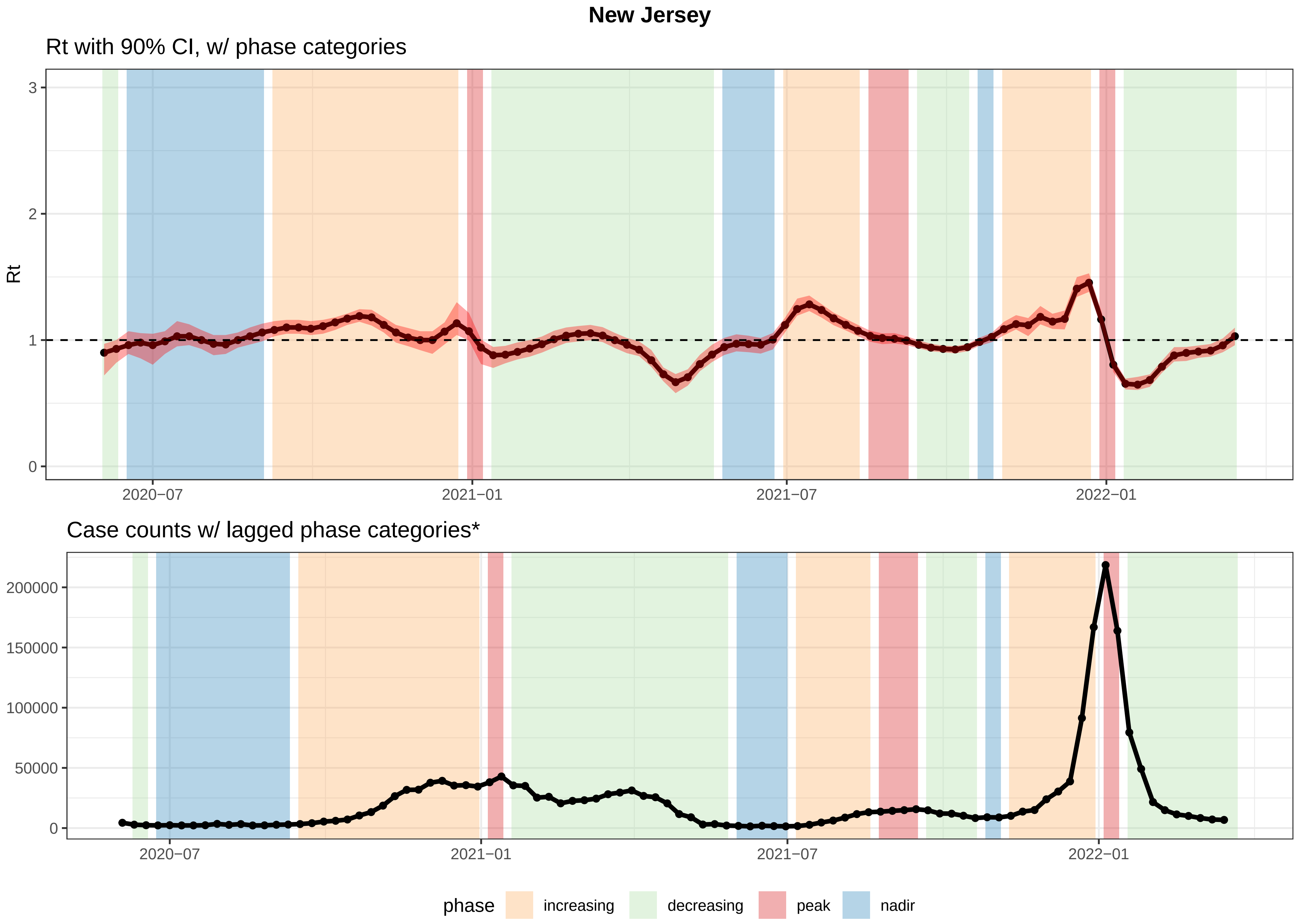


*Increasing/decreasing = Rt had a 90% probability >= or <= than 1.0.
Wks b/w two increasing/decreasing phases −> classified as increasing/decreasing.
Wks b/w increasing and decreasing phases = peaks; nadirs = wks b/w decreasing and increasing phases.

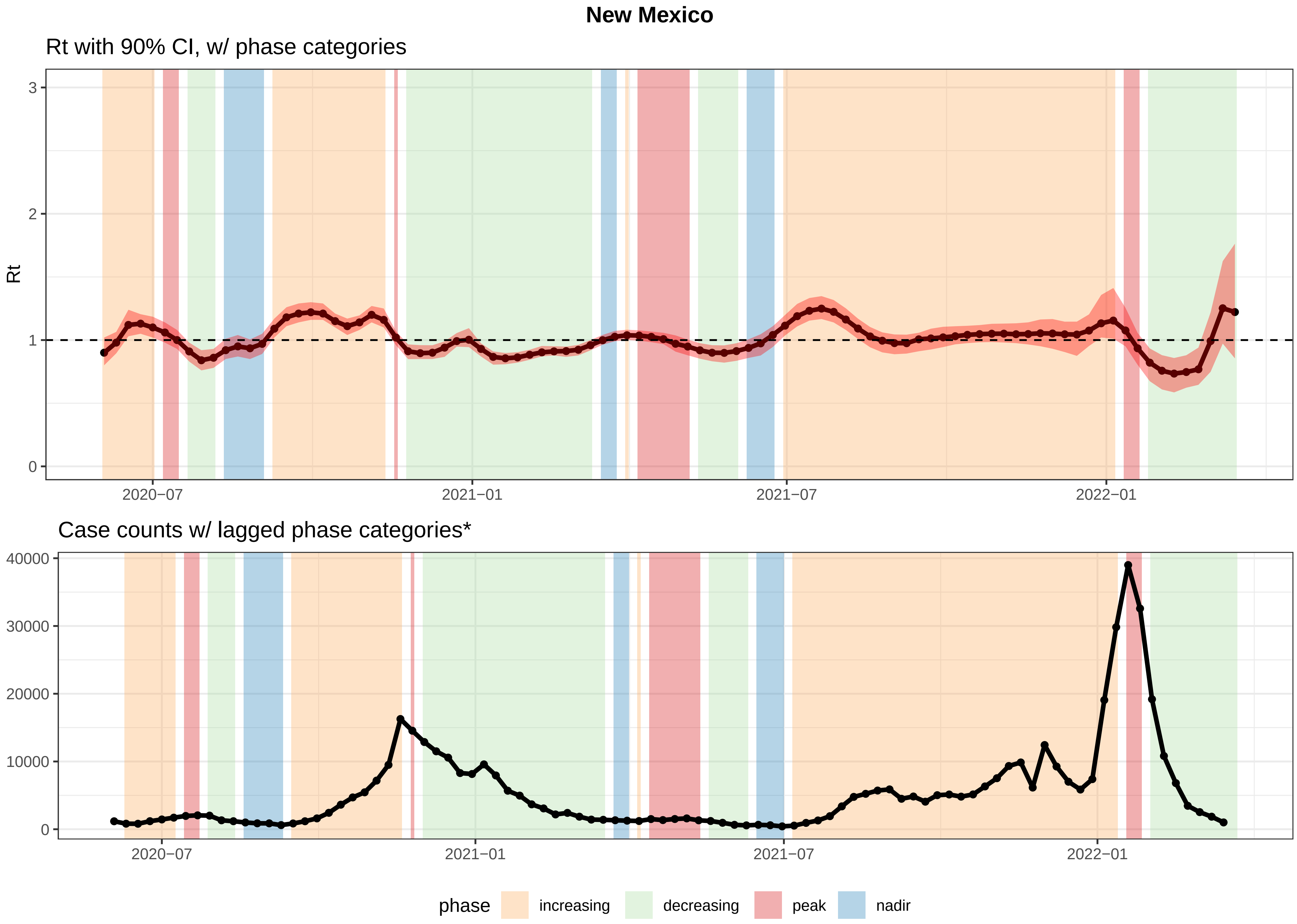


*Increasing/decreasing = Rt had a 90% probability >= or <= than 1.0.
Wks b/w two increasing/decreasing phases −> classified as increasing/decreasing.
Wks b/w increasing and decreasing phases = peaks; nadirs = wks b/w decreasing and increasing phases.

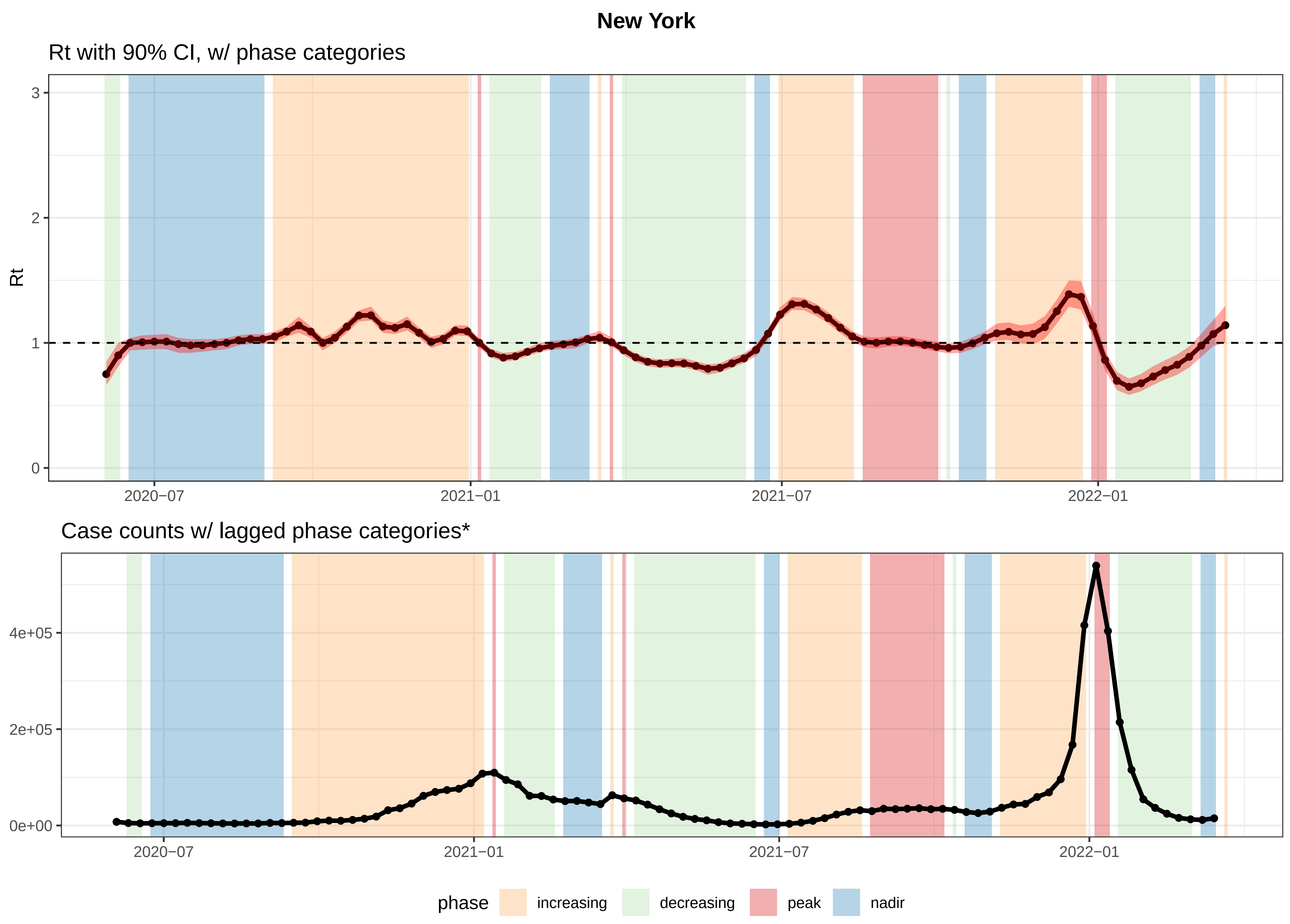


*Increasing/decreasing = Rt had a 90% probability >= or <= than 1.0.
Wks b/w two increasing/decreasing phases −> classified as increasing/decreasing.
Wks b/w increasing and decreasing phases = peaks; nadirs = wks b/w decreasing and increasing phases.

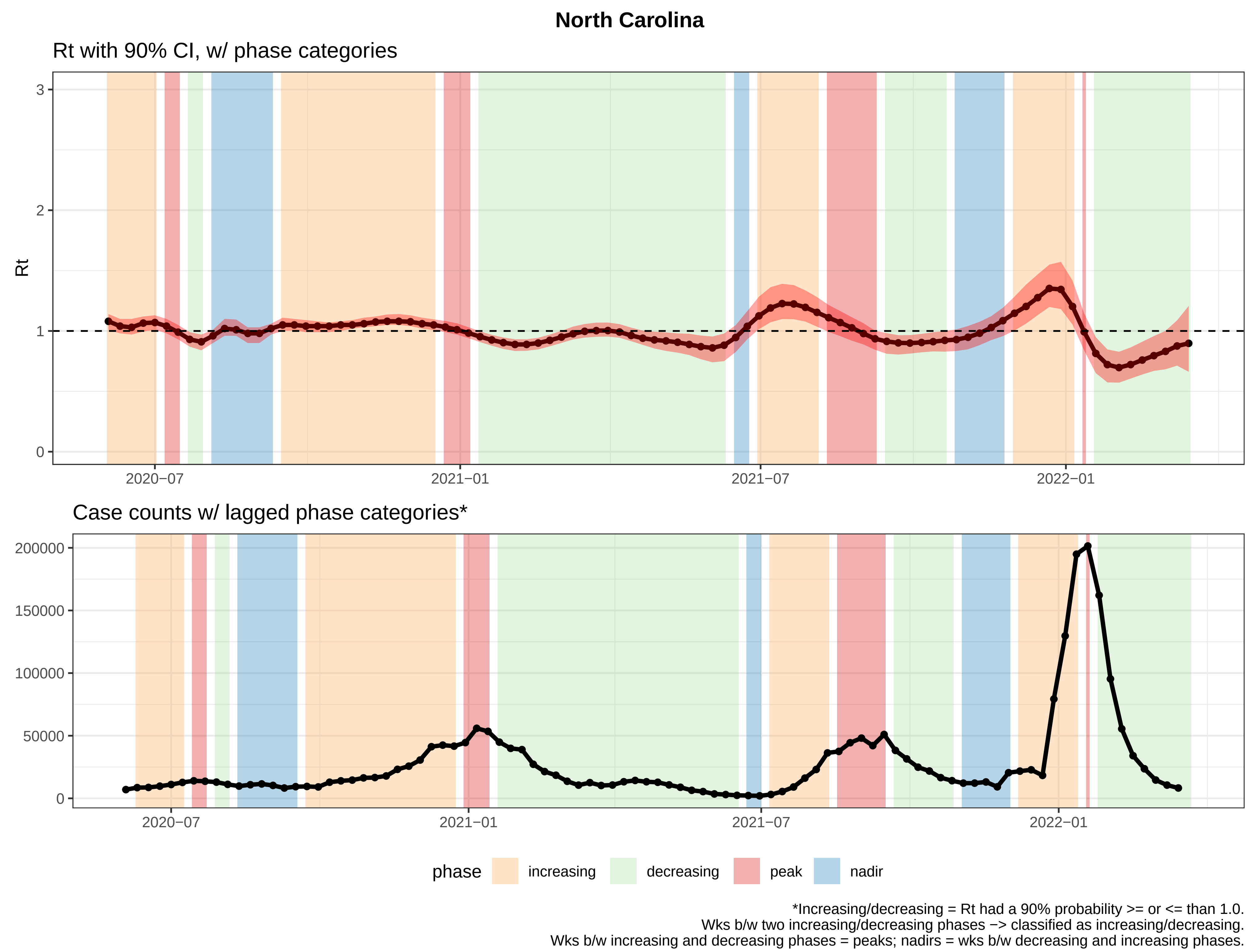

North Carolina
Rt with 90% CI, w/ phase categories
Rt
3
2
1
0
2020−07
2021−01
2021−07
2022−01
Case counts w/ lagged phase categories*
200000
150000
100000
50000
0
2020−07
2021−01
2021−07
2022−01
phase
increasing
decreasing
peak
nadir
*Increasing/decreasing = Rt had a 90% probability >= or <= than 1.0.
Wks b/w two increasing/decreasing phases −> classified as increasing/decreasing.
Wks b/w increasing and decreasing phases = peaks; nadirs = wks b/w decreasing and increasing phases.

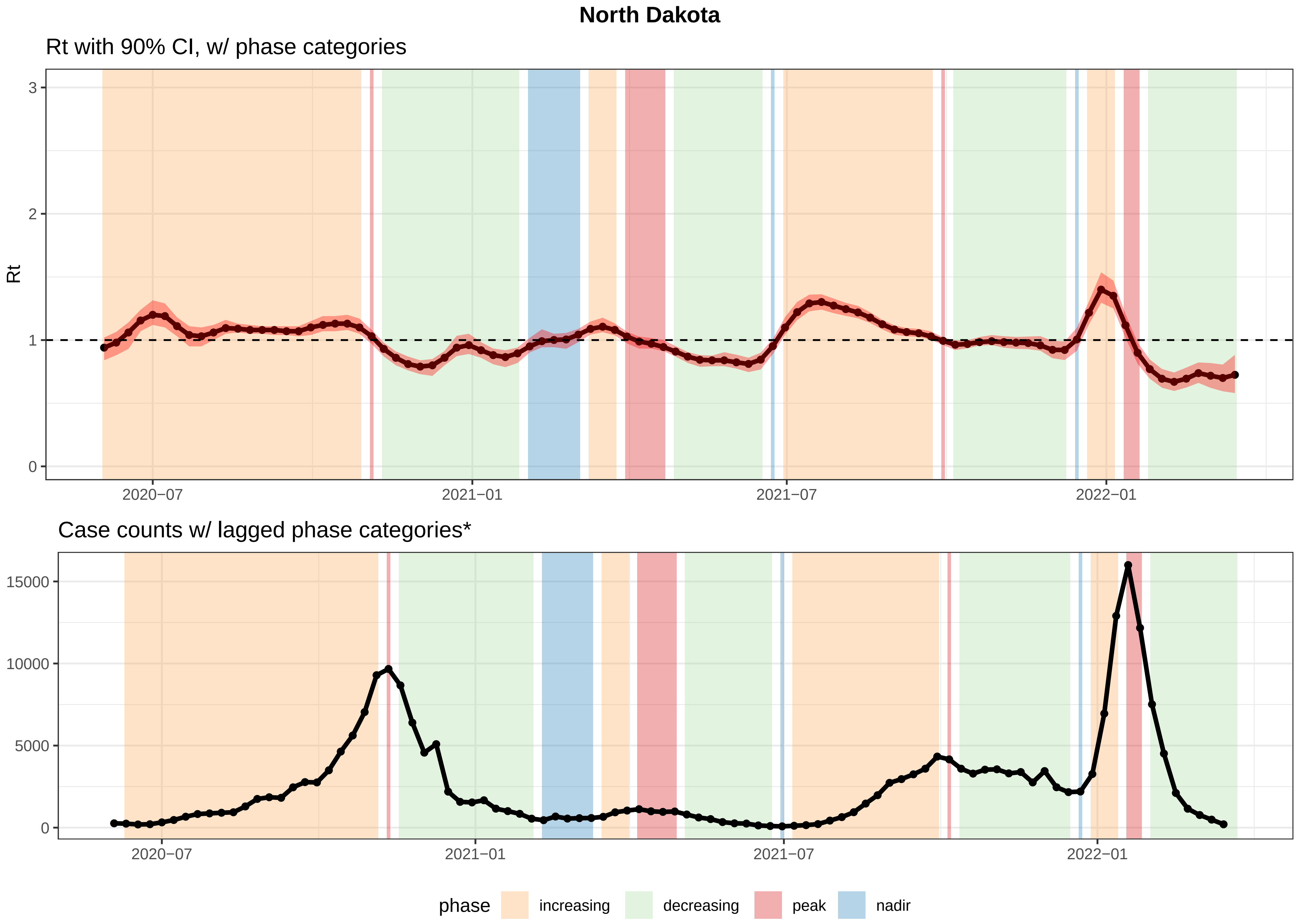


*Increasing/decreasing = Rt had a 90% probability >= or <= than 1.0.
Wks b/w two increasing/decreasing phases −> classified as increasing/decreasing.
Wks b/w increasing and decreasing phases = peaks; nadirs = wks b/w decreasing and increasing phases.

### Ohio

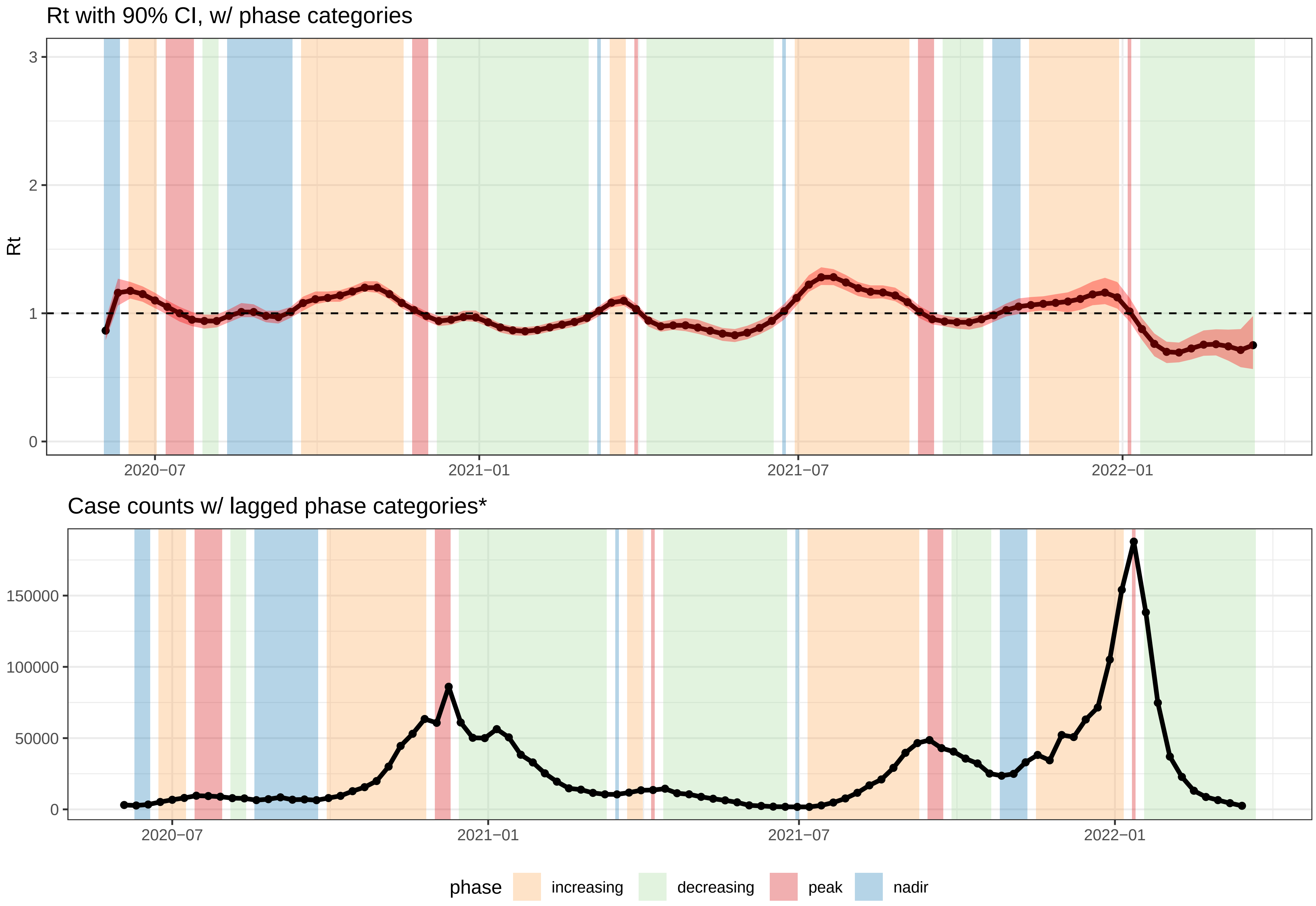


*Increasing/decreasing = Rt had a 90% probability >= or <= than 1.0.
Wks b/w two increasing/decreasing phases −> classified as increasing/decreasing.
Wks b/w increasing and decreasing phases = peaks; nadirs = wks b/w decreasing and increasing phases.

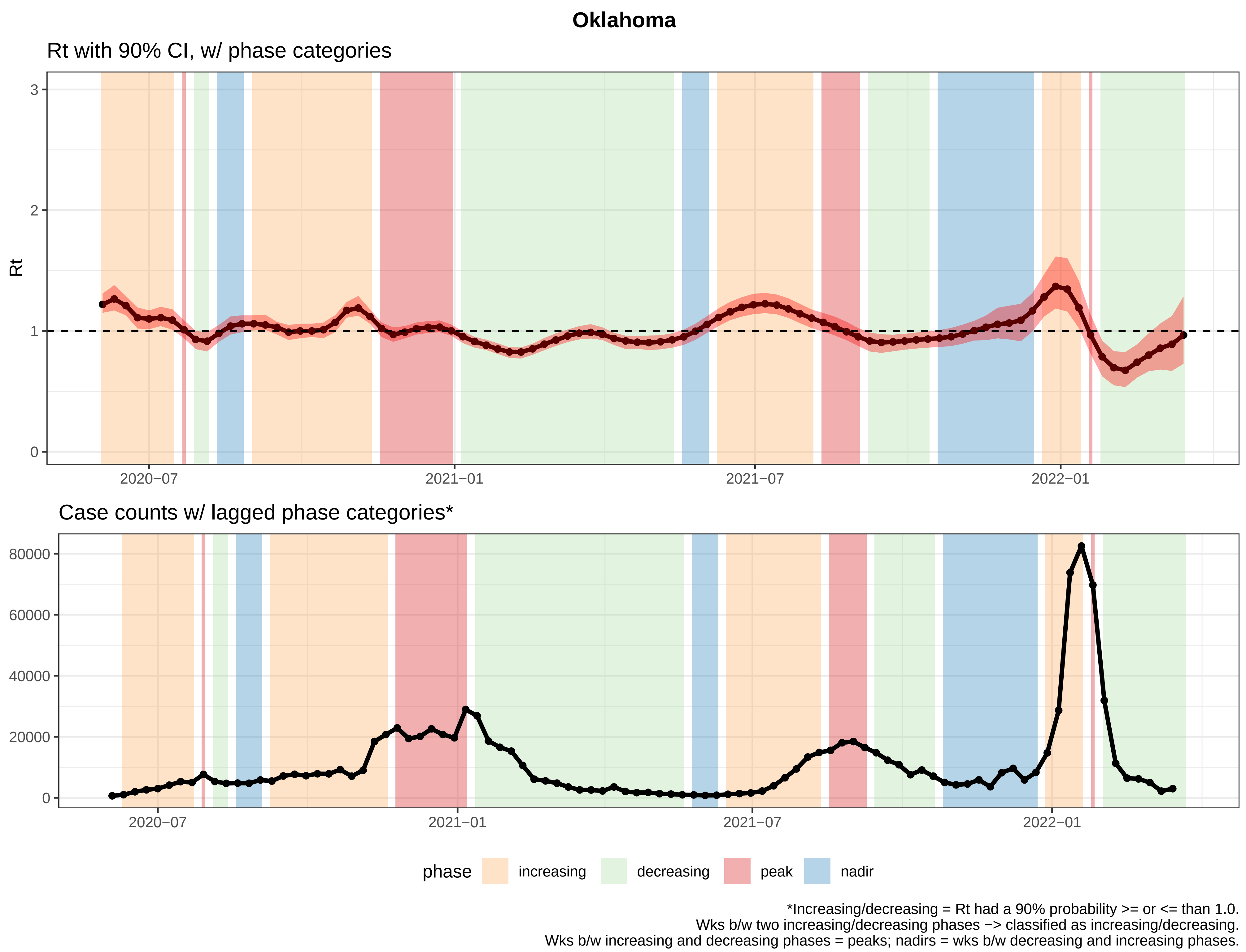

Oklahoma
Rt with 90% CI, w/ phase categories
Rt
3
2
1
0
2020−07
2021−01
2021−07
2022−01
Case counts w/ lagged phase categories*
80000
60000
40000
20000
0
2020−07
2021−01
2021−07
2022−01
phase
increasing
decreasing
peak
nadir
*Increasing/decreasing = Rt had a 90% probability >= or <= than 1.0.
Wks b/w two increasing/decreasing phases −> classified as increasing/decreasing.
Wks b/w increasing and decreasing phases = peaks; nadirs = wks b/w decreasing and increasing phases.

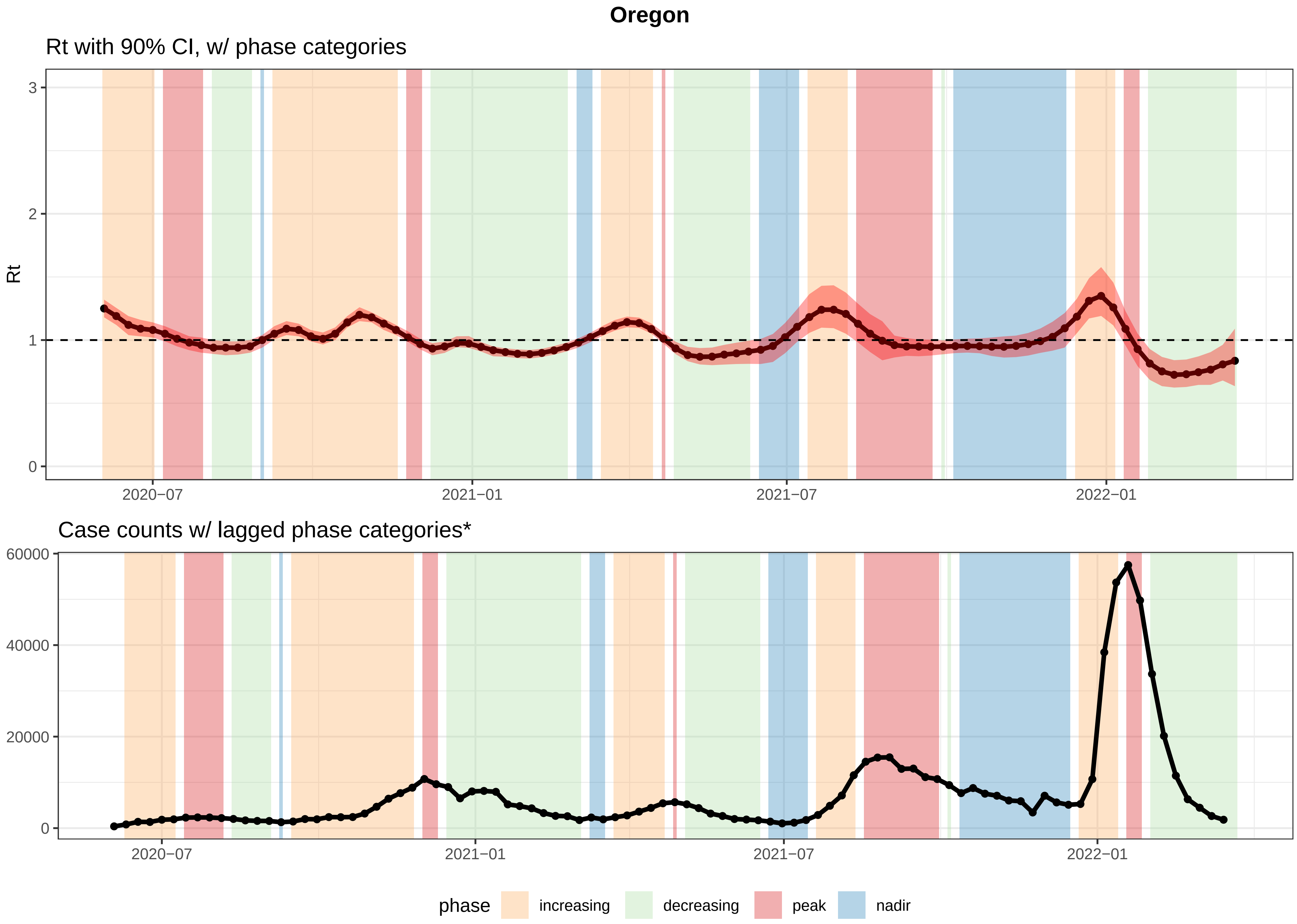


*Increasing/decreasing = Rt had a 90% probability >= or <= than 1.0.
Wks b/w two increasing/decreasing phases −> classified as increasing/decreasing.
Wks b/w increasing and decreasing phases = peaks; nadirs = wks b/w decreasing and increasing phases.

**Pennsylvania**

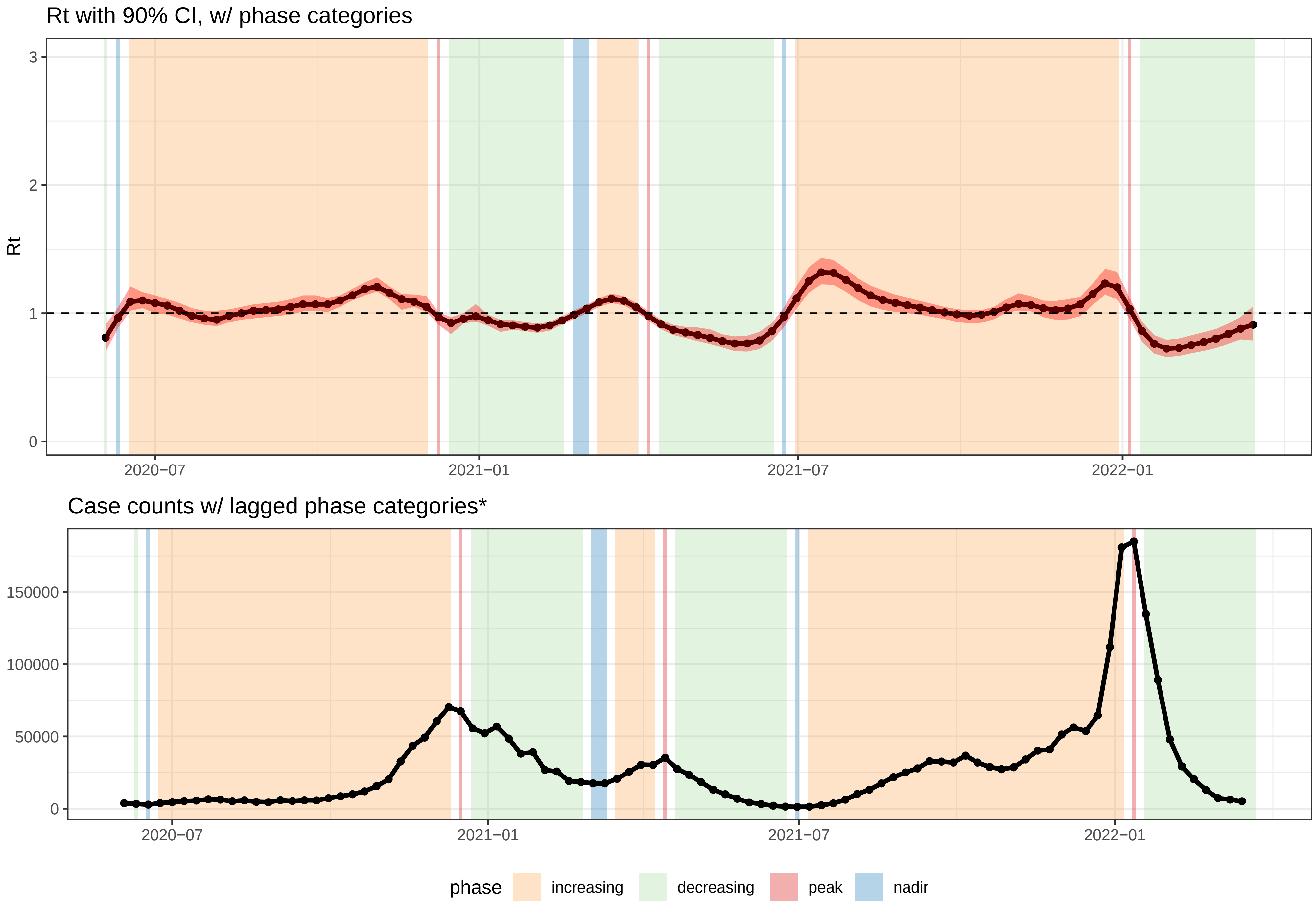


*Increasing/decreasing = Rt had a 90% probability >= or <= than 1.0.
Wks b/w two increasing/decreasing phases −> classified as increasing/decreasing.
Wks b/w increasing and decreasing phases = peaks; nadirs = wks b/w decreasing and increasing phases.

### Rhode Island

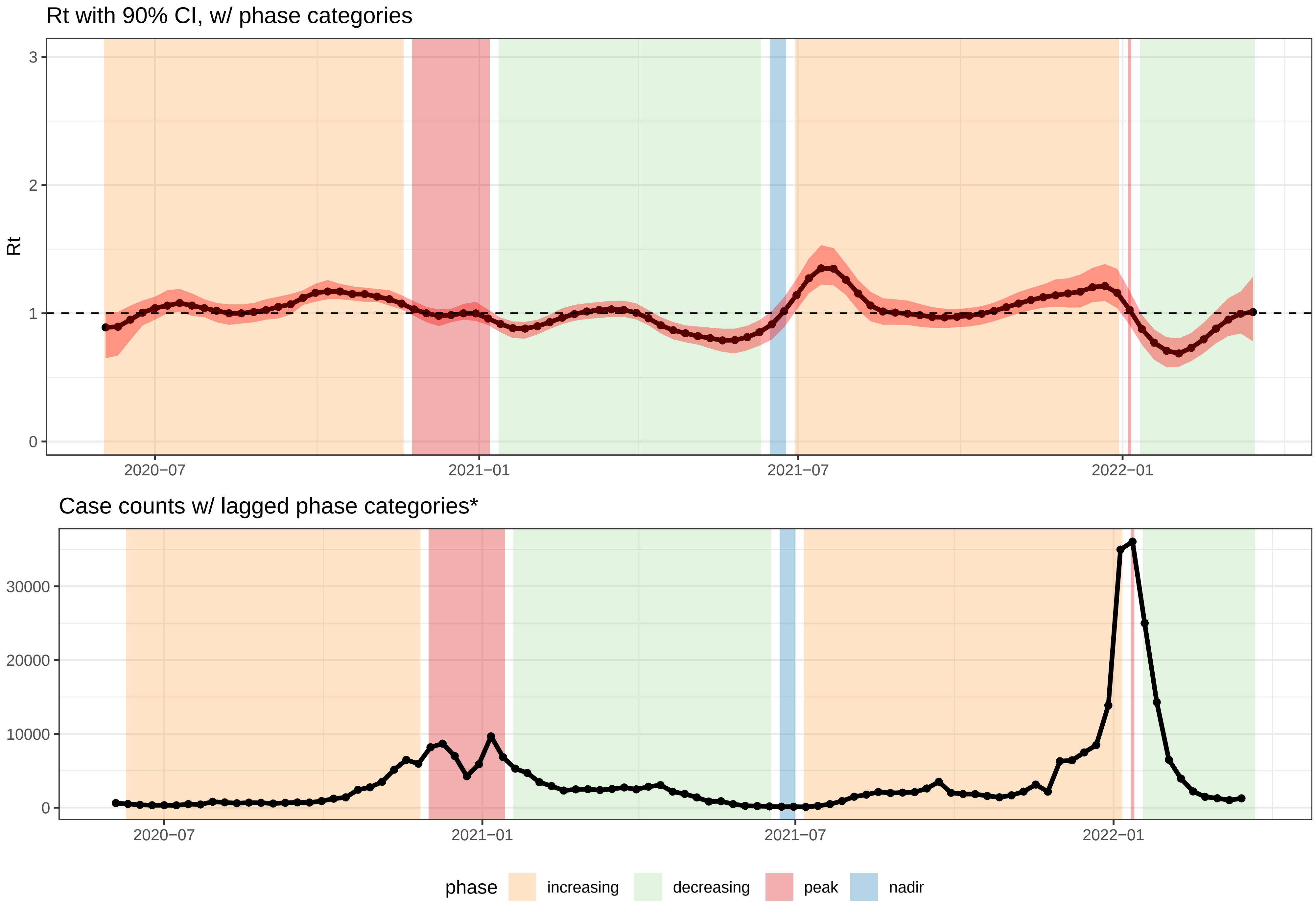


*Increasing/decreasing = Rt had a 90% probability >= or <= than 1.0.
Wks b/w two increasing/decreasing phases −> classified as increasing/decreasing.
Wks b/w increasing and decreasing phases = peaks; nadirs = wks b/w decreasing and increasing phases.

#### South Carolina

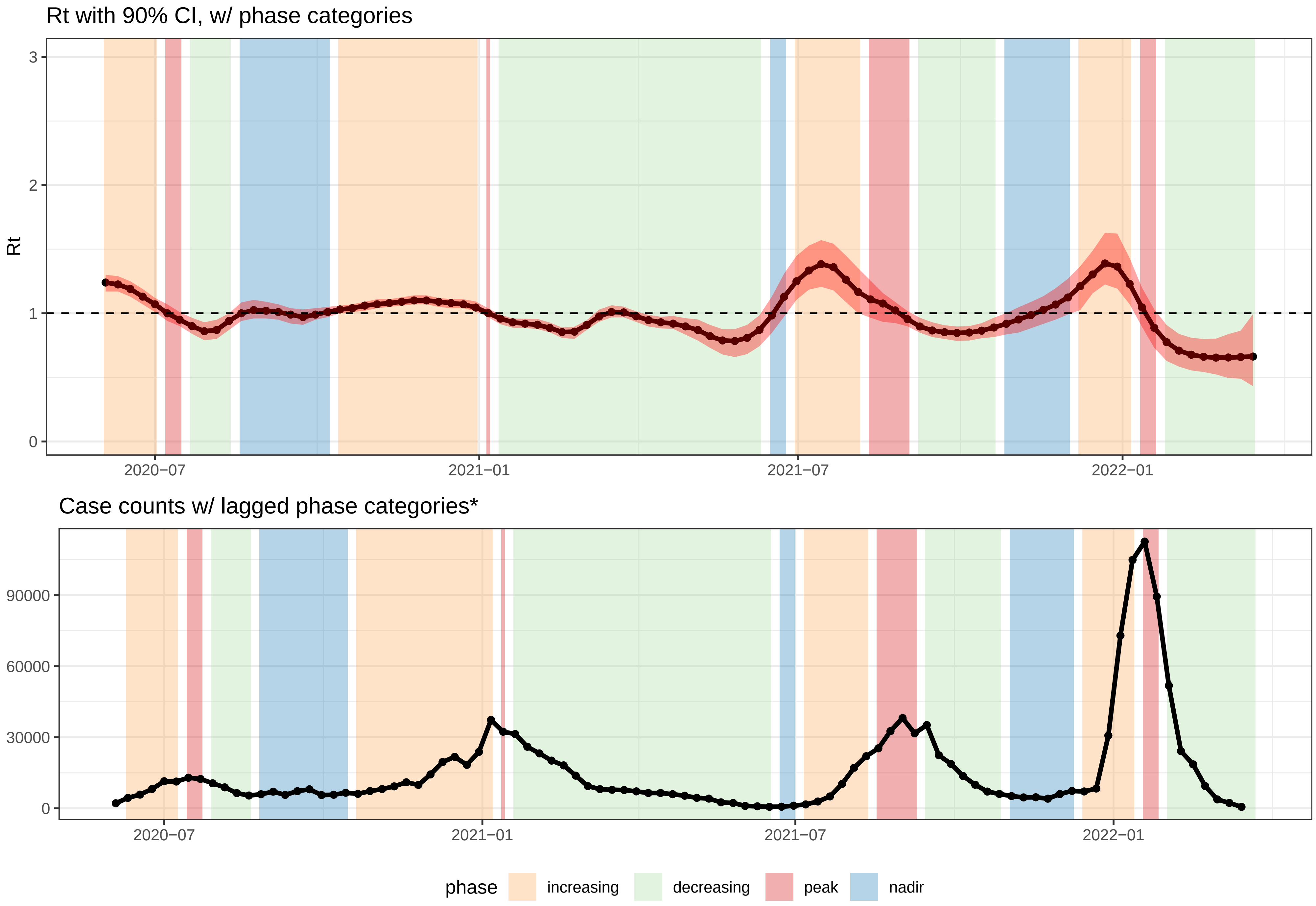


*Increasing/decreasing = Rt had a 90% probability >= or <= than 1.0.
Wks b/w two increasing/decreasing phases −> classified as increasing/decreasing.
Wks b/w increasing and decreasing phases = peaks; nadirs = wks b/w decreasing and increasing phases.

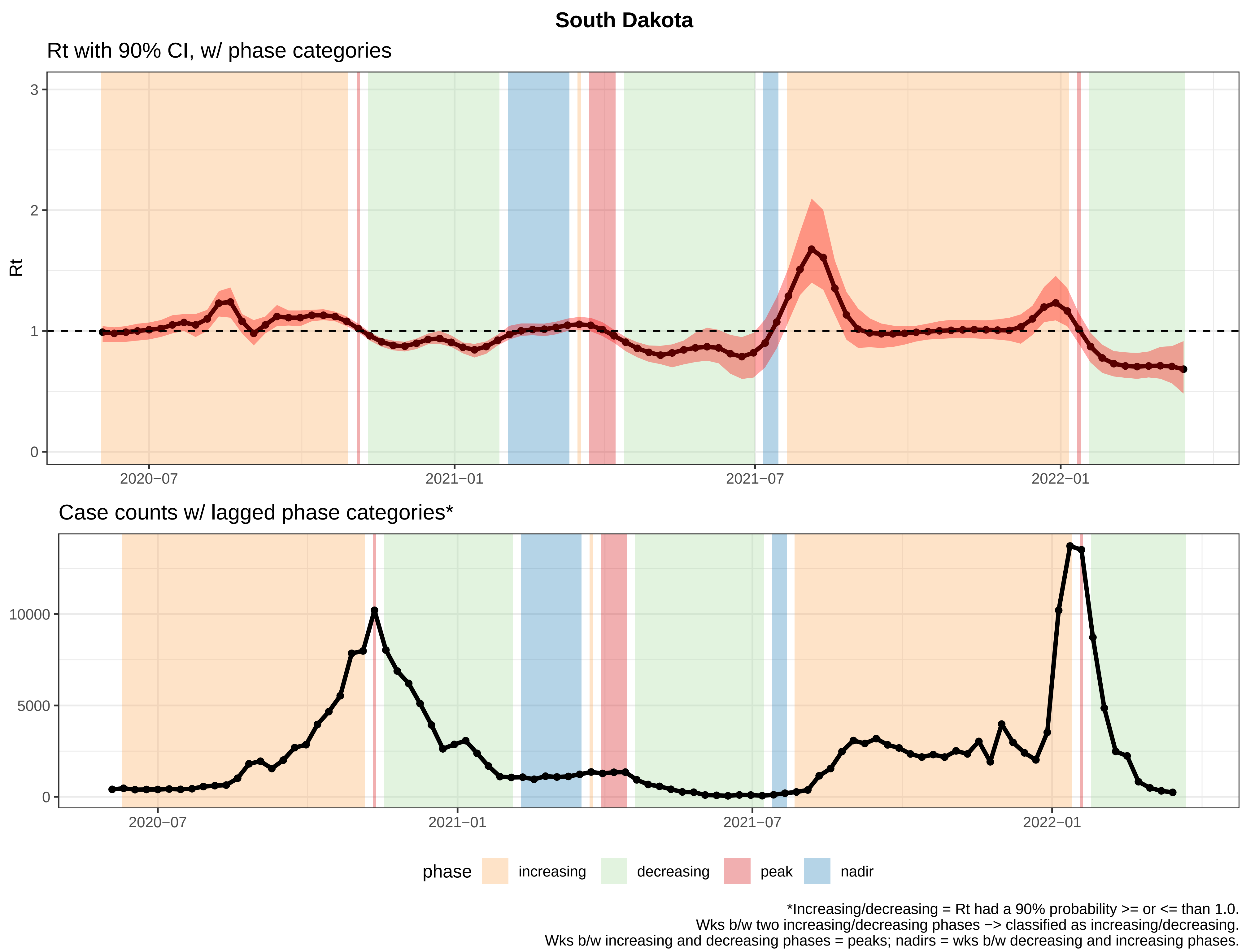
South Dakota
Rt with 90% CI, w/ phase categories
Rt
3
2
1
0
2020−07
2021−01
2021−07
2022−01
Case counts w/ lagged phase categories*
10000
5000
0
2020−07
2021−01
2021−07
2022−01
phase
increasing
decreasing
peak
nadir
*Increasing/decreasing = Rt had a 90% probability >= or <= than 1.0.
Wks b/w two increasing/decreasing phases −> classified as increasing/decreasing.
Wks b/w increasing and decreasing phases = peaks; nadirs = wks b/w decreasing and increasing phases.

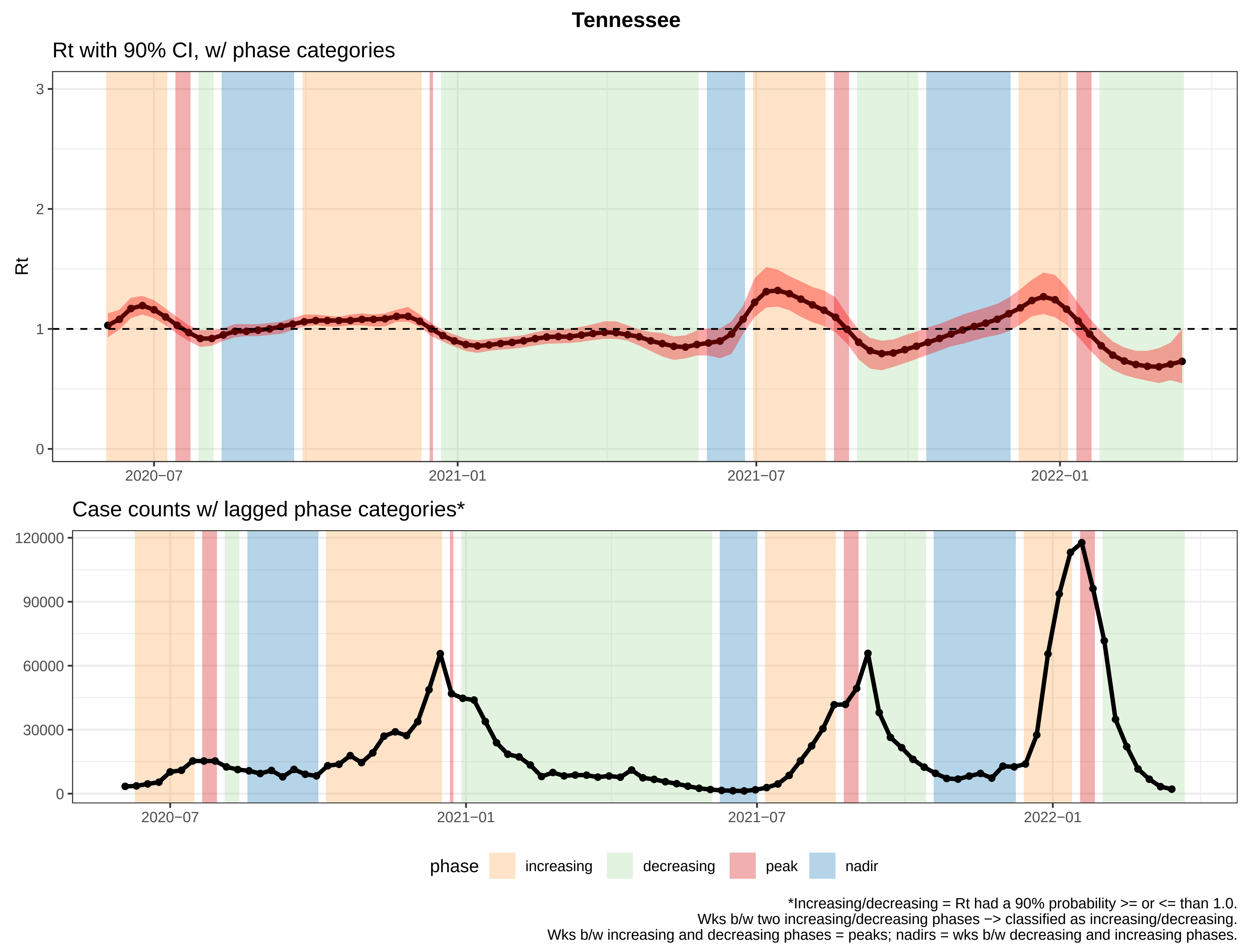
Tennessee
Rt with 90% CI, w/ phase categories
Rt
3
2
1
0
2020−07
2021−01
2021−07
2022−01
Case counts w/ lagged phase categories*
120000
90000
60000
30000
0
2020−07
2021−01
2021−07
2022−01
phase
increasing
decreasing
peak
nadir
*Increasing/decreasing = Rt had a 90% probability >= or <= than 1.0.
Wks b/w two increasing/decreasing phases −> classified as increasing/decreasing.
Wks b/w increasing and decreasing phases = peaks; nadirs = wks b/w decreasing and increasing phases.

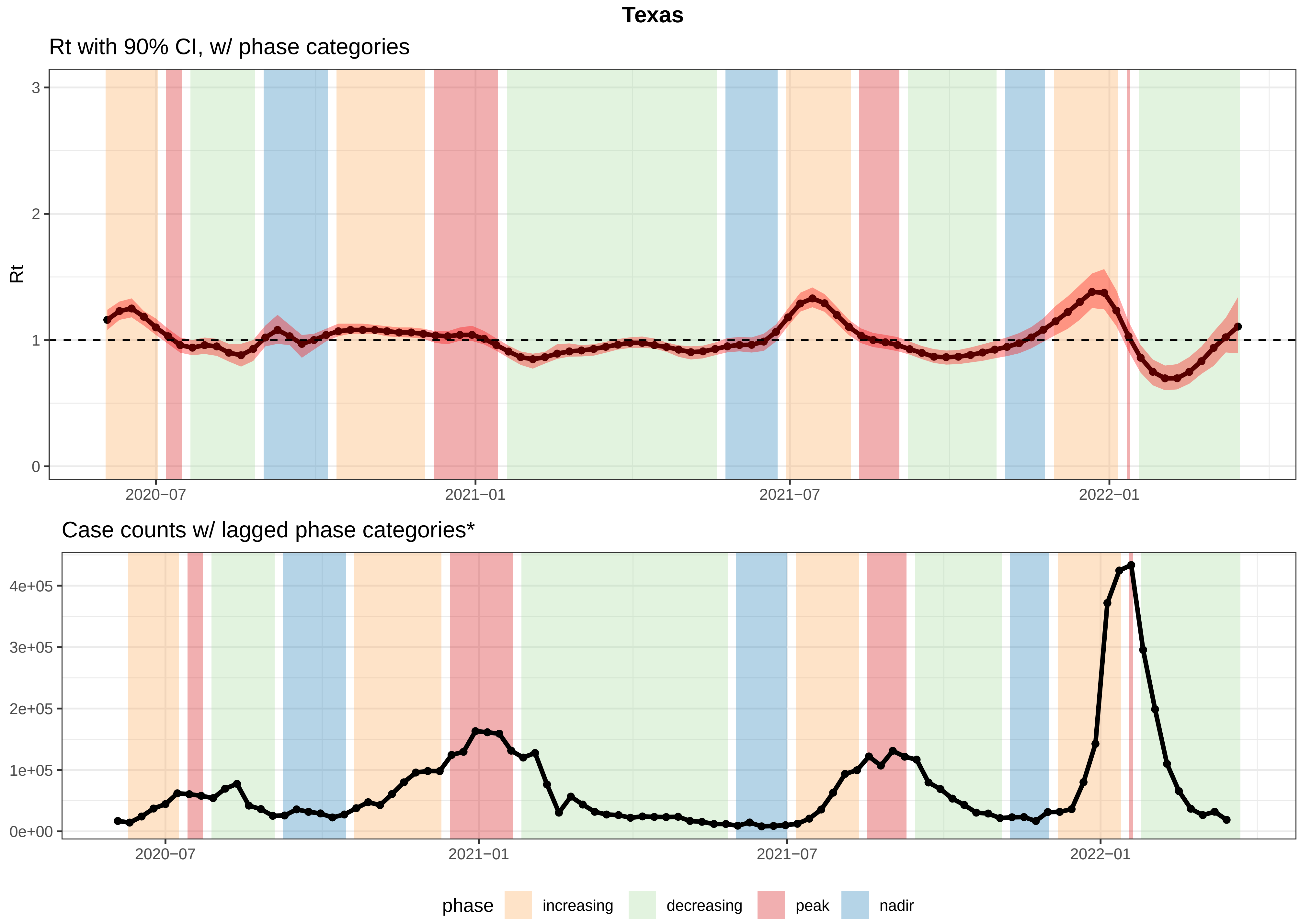


*Increasing/decreasing = Rt had a 90% probability >= or <= than 1.0.
Wks b/w two increasing/decreasing phases −> classified as increasing/decreasing.
Wks b/w increasing and decreasing phases = peaks; nadirs = wks b/w decreasing and increasing phases.

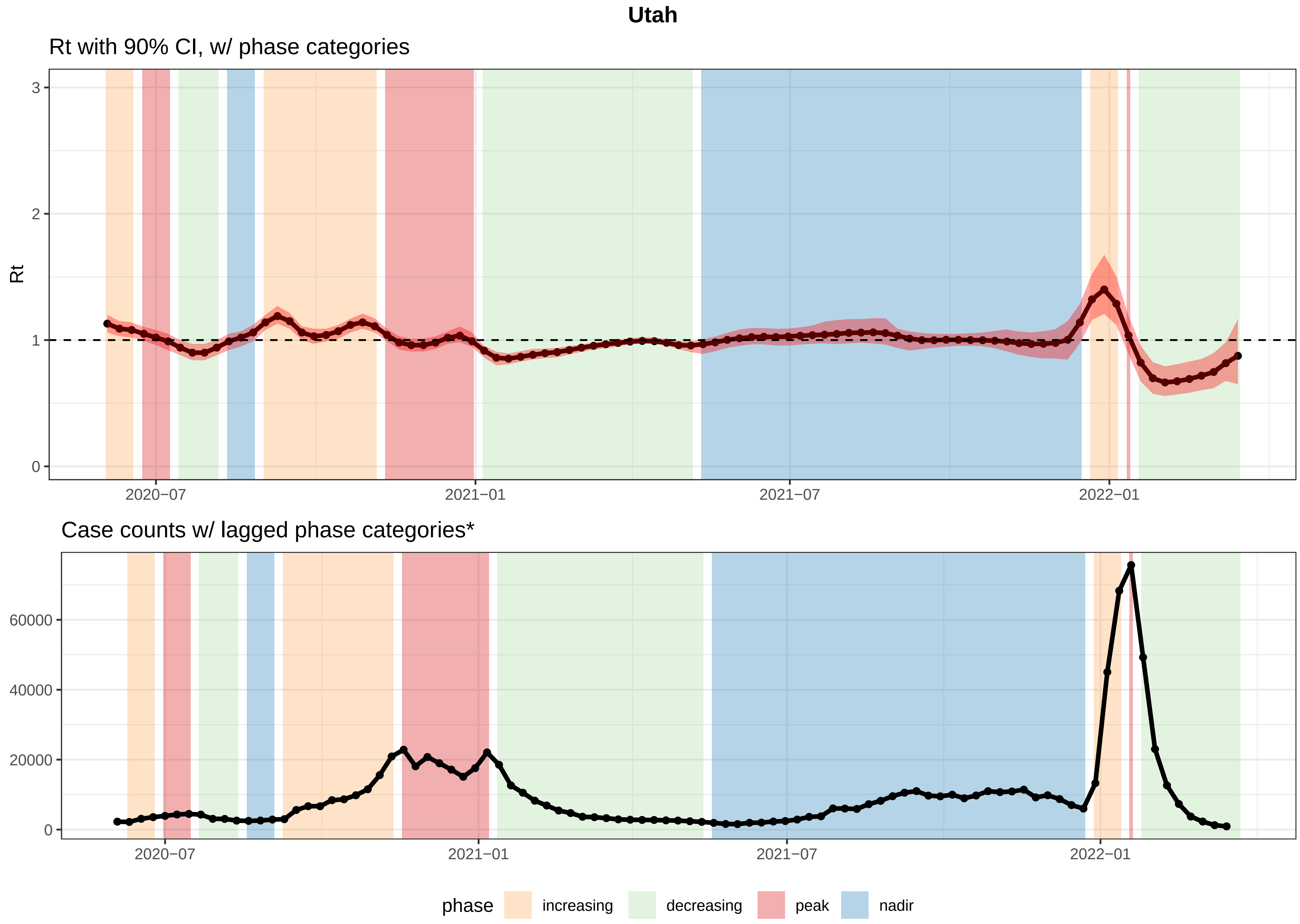


*Increasing/decreasing = Rt had a 90% probability >= or <= than 1.0.
Wks b/w two increasing/decreasing phases −> classified as increasing/decreasing.
Wks b/w increasing and decreasing phases = peaks; nadirs = wks b/w decreasing and increasing phases.

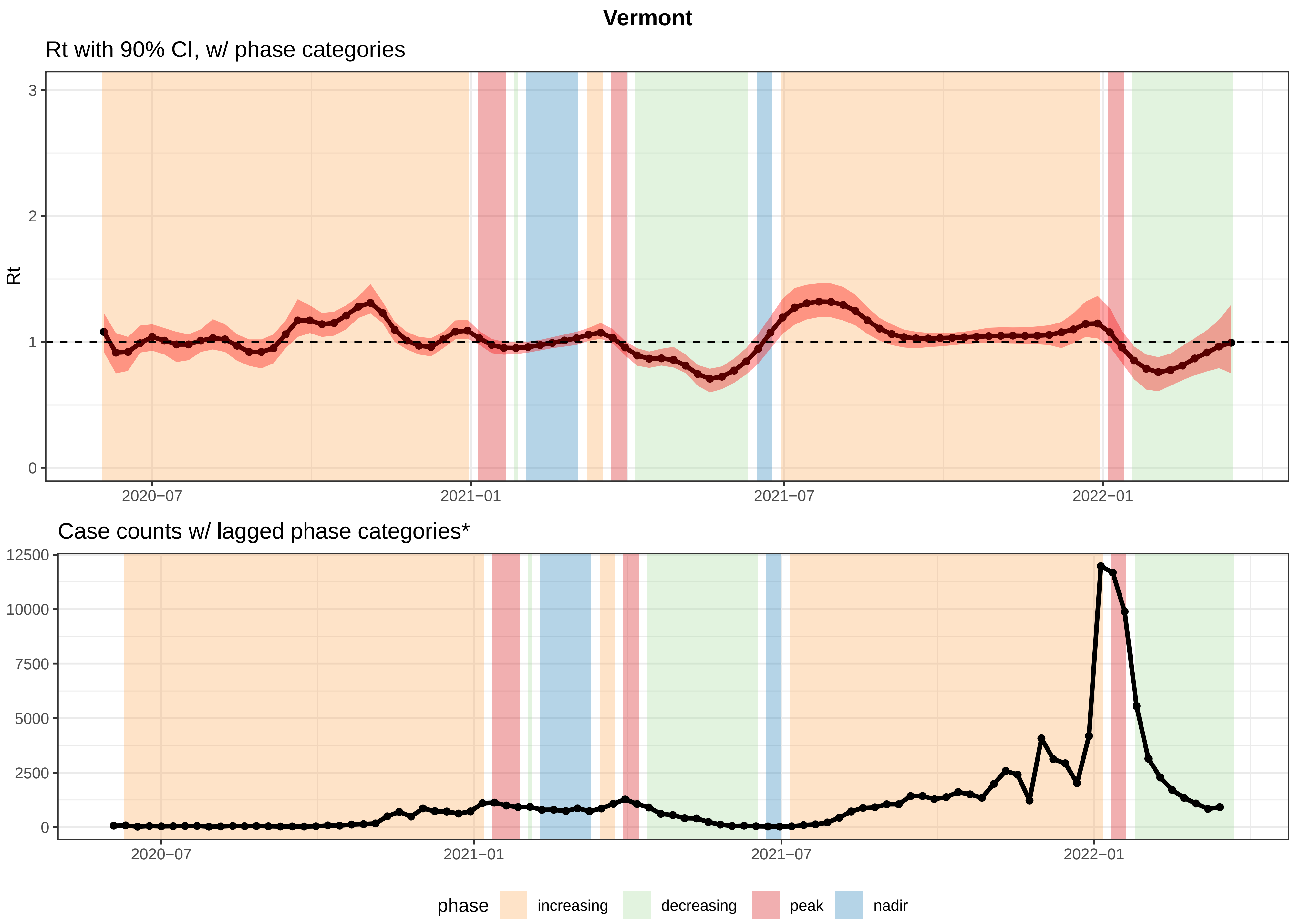


*Increasing/decreasing = Rt had a 90% probability >= or <= than 1.0.
Wks b/w two increasing/decreasing phases −> classified as increasing/decreasing.
Wks b/w increasing and decreasing phases = peaks; nadirs = wks b/w decreasing and increasing phases.

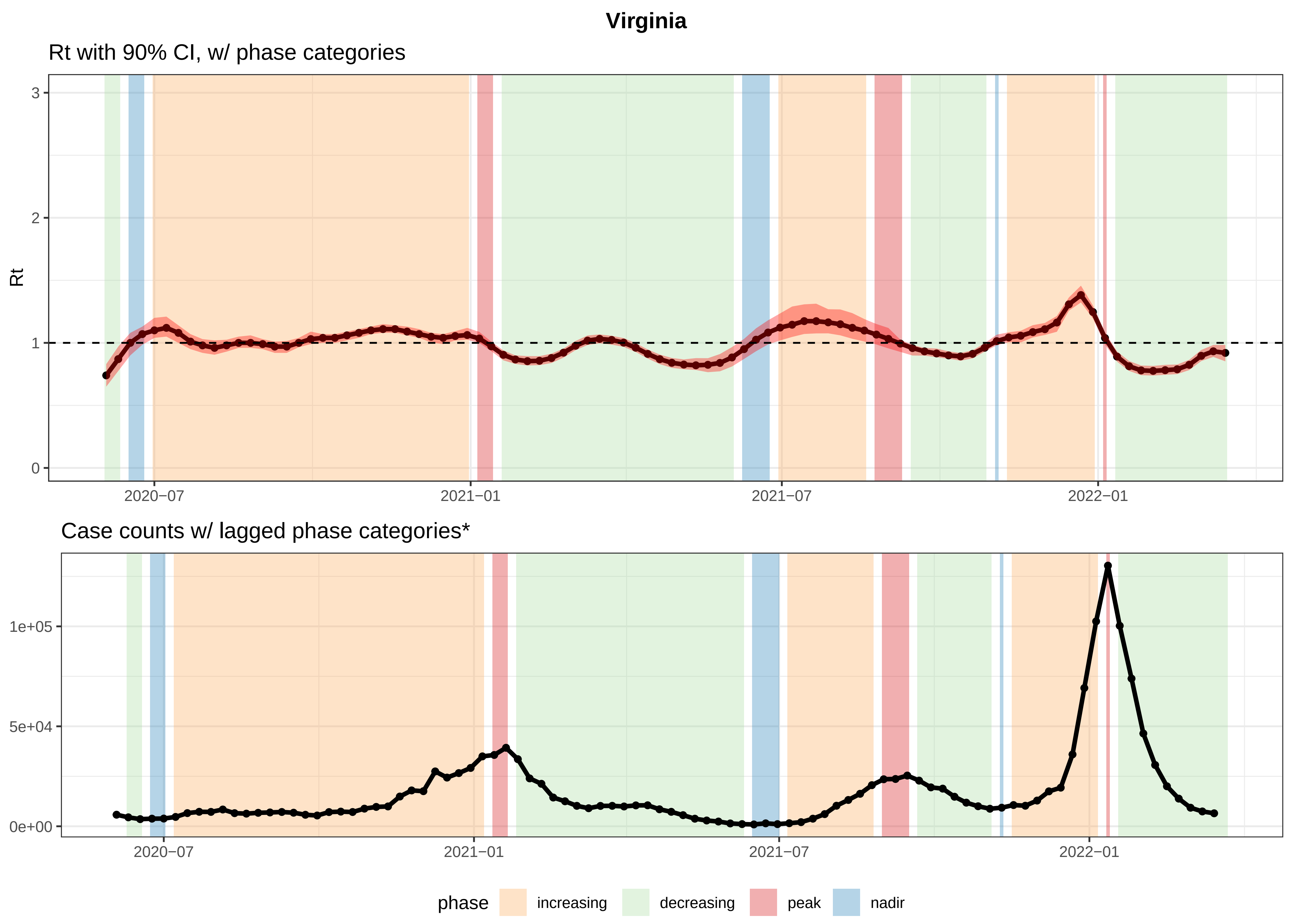


*Increasing/decreasing = Rt had a 90% probability >= or <= than 1.0.
Wks b/w two increasing/decreasing phases −> classified as increasing/decreasing.
Wks b/w increasing and decreasing phases = peaks; nadirs = wks b/w decreasing and increasing phases.

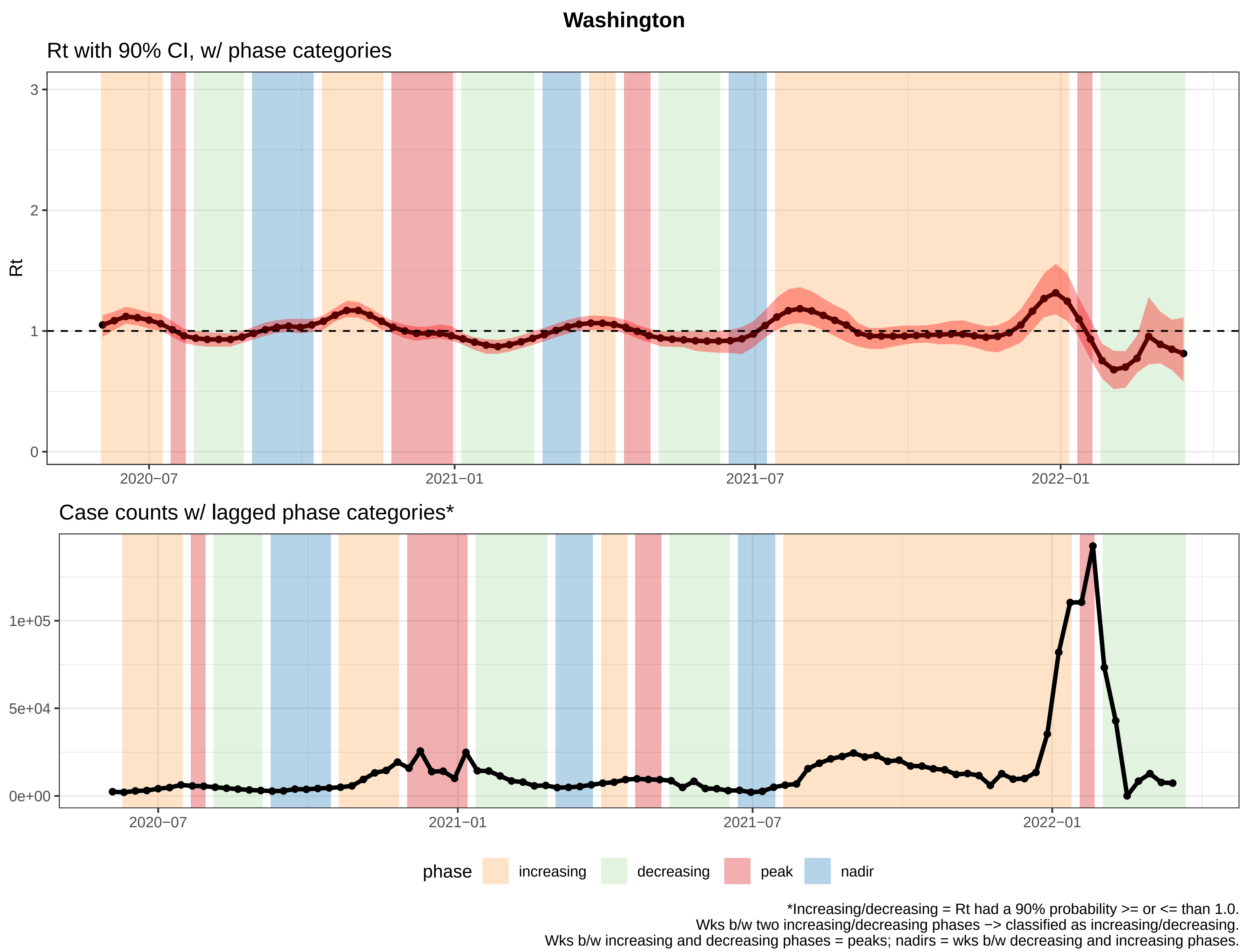

Washington
Rt with 90% CI, w/ phase categories
Rt
3
2
1
0
2020−07
2021−01
2021−07
2022−01
Case counts w/ lagged phase categories*
1e+05
5e+04
0e+00
2020−07
2021−01
2021−07
2022−01
phase
increasing
decreasing
peak
nadir
*Increasing/decreasing = Rt had a 90% probability >= or <= than 1.0.
Wks b/w two increasing/decreasing phases −> classified as increasing/decreasing.
Wks b/w increasing and decreasing phases = peaks; nadirs = wks b/w decreasing and increasing phases.

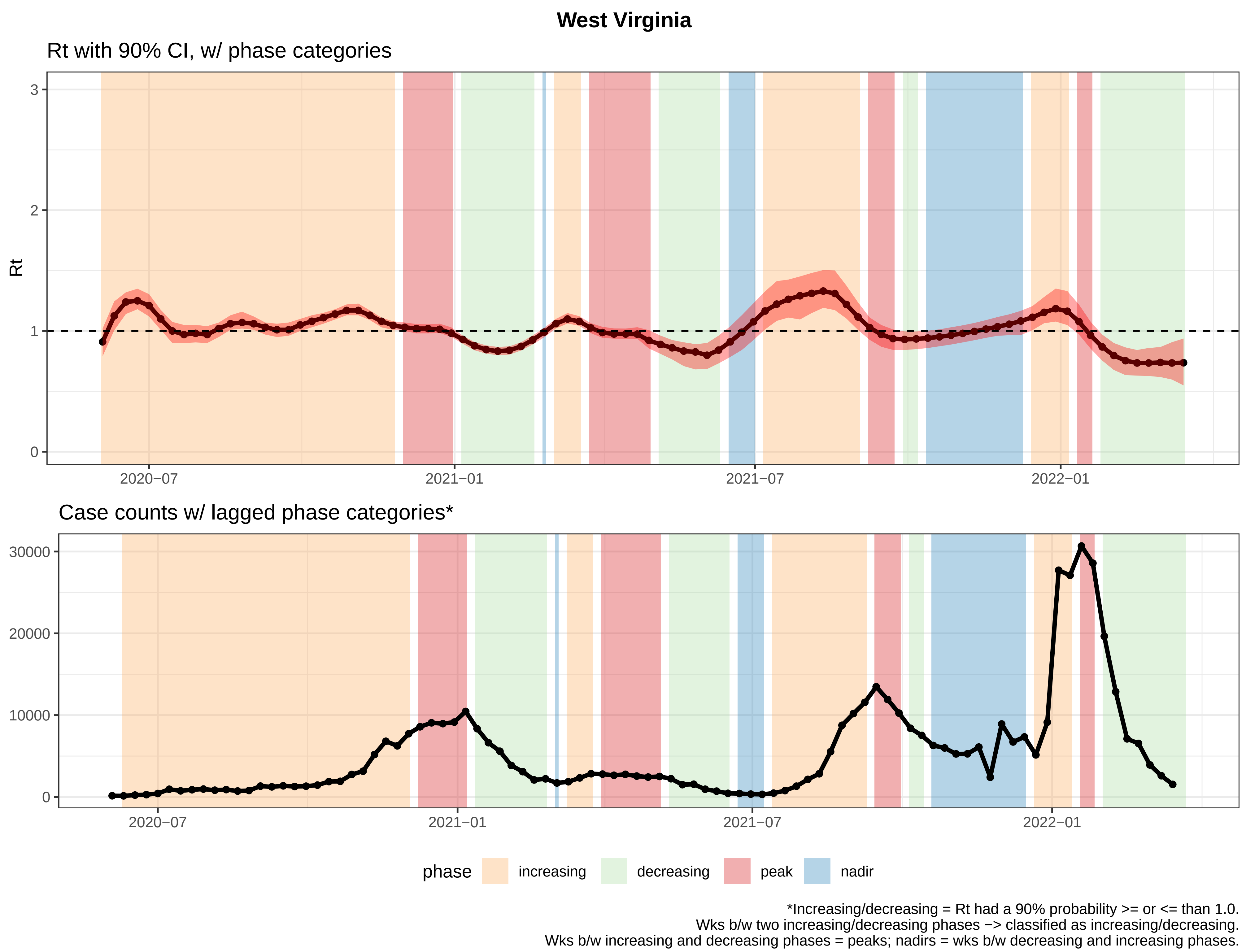
West Virginia
Rt with 90% CI, w/ phase categories
Rt
3
2
1
0
2020−07
2021−01
2021−07
2022−01
Case counts w/ lagged phase categories*
30000
20000
10000
0
2020−07
2021−01
2021−07
2022−01
phase
increasing
decreasing
peak
nadir
*Increasing/decreasing = Rt had a 90% probability >= or <= than 1.0.
Wks b/w two increasing/decreasing phases −> classified as increasing/decreasing.
Wks b/w increasing and decreasing phases = peaks; nadirs = wks b/w decreasing and increasing phases.

#### Wisconsin

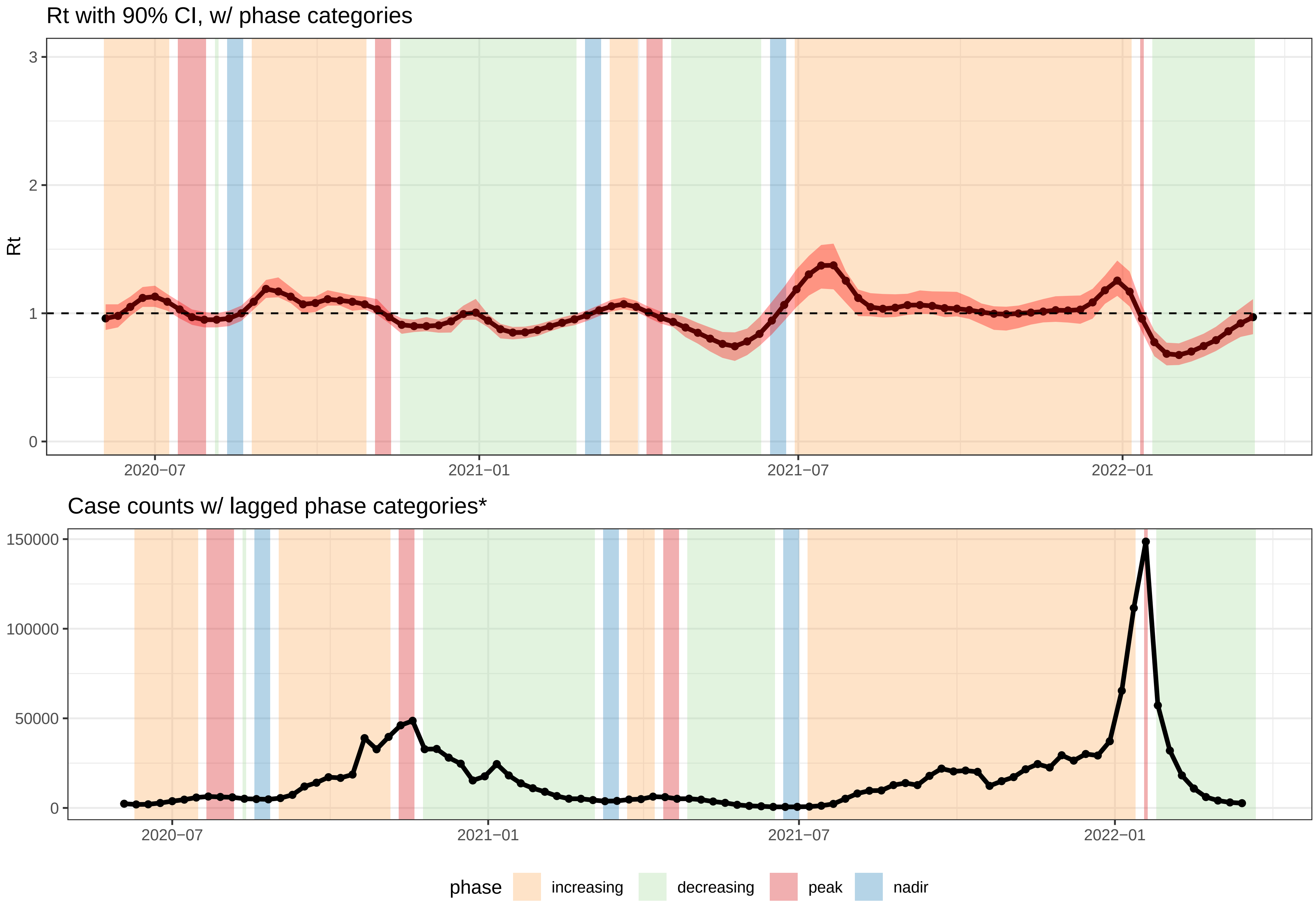


*Increasing/decreasing = Rt had a 90% probability >= or <= than 1.0.
Wks b/w two increasing/decreasing phases −> classified as increasing/decreasing.
Wks b/w increasing and decreasing phases = peaks; nadirs = wks b/w decreasing and increasing phases.

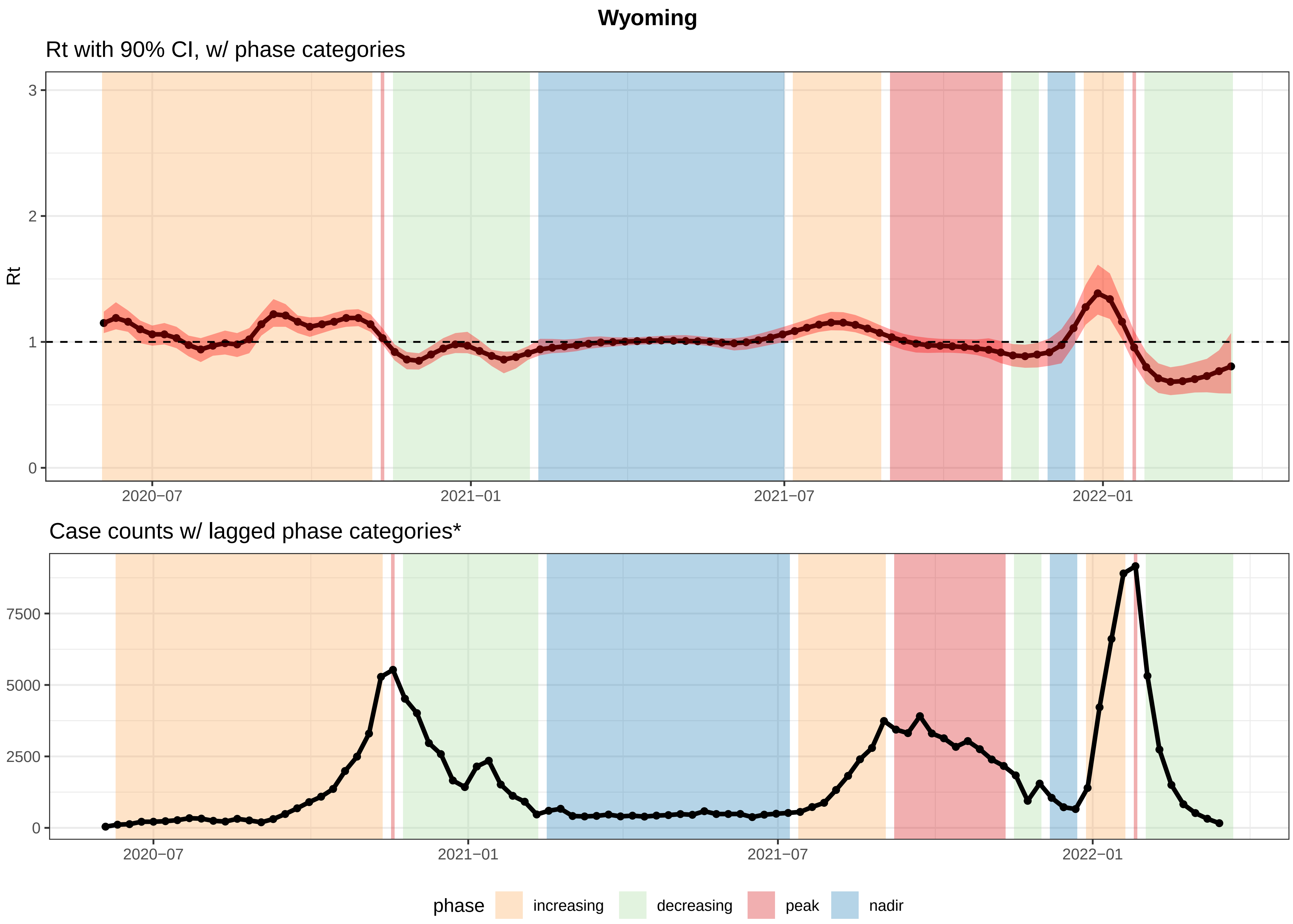


*Increasing/decreasing = Rt had a 90% probability >= or <= than 1.0.
Wks b/w two increasing/decreasing phases −> classified as increasing/decreasing.
Wks b/w increasing and decreasing phases = peaks; nadirs = wks b/w decreasing and increasing phases.
