## Supplement 5 for "Challenges of COVID-19 Case Forecasting in the US, 2020-2021"

**Supporting Information 5**: Each location specific forecast submitted to the COVID19 Forecast Hub included at least 4 weeks of future predictions. Here, we present disaggregated 1 and 4 week ahead predictions of model performance for each team model that submitted national and state/territory/DC forecasts and were included in the main analyses. Specific plots include the average 50%, 80% and 95% coverage for eligible submitted forecasts (S5 Figure 5.1), average absolute Weighted Interval Score (WIS) and 95% coverage over time (S5 Figure 5.2), and scaled, pairwise rWIS by location (S5 Figure 5.3)

**S5 Figure 5.1.** Expected and observed coverage rates aggregated for 1 and 4 week ahead forecasts over time for national forecasts in A, state/territory/DC forecasts in B, the largest country forecasts in C. The dashed line represents optimal expected coverage. Teams that outperformed the COVIDhub-4_week_ensemble model at all coverage levels are labeled on the right-hand side of the plots.

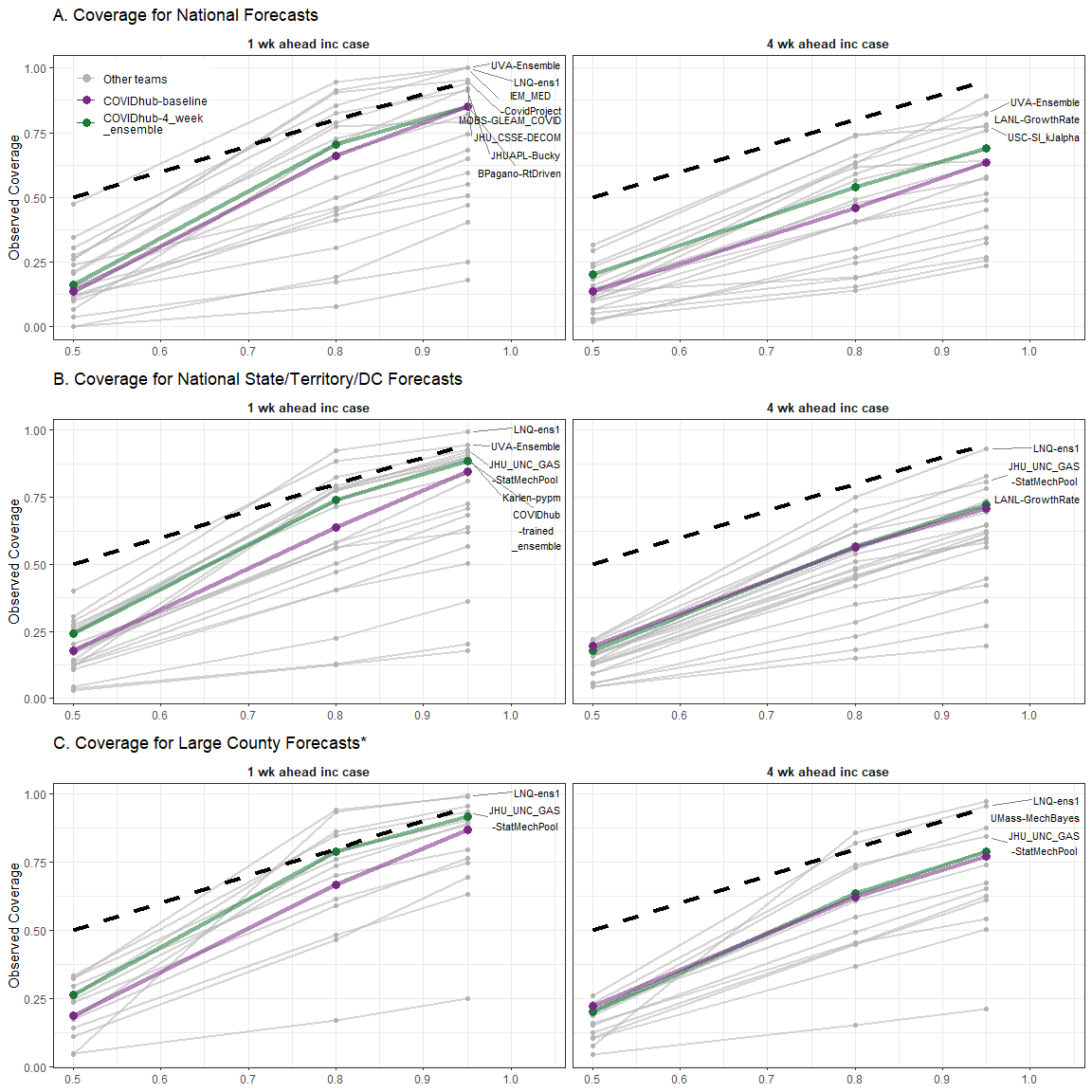

**S5 Figure 5.2.** Mean Weighted Interval Score (WIS) over time for 1 and 4 week ahead forecasts, aggregated by geographic units, and 95% coverage over time for 1 and 4 week ahead forecasts, aggregated by geographic units. The black, dashed vertical line in all panels shows the date that public communication of the case forecasts was paused. The black, dashed horizontal line in panels D, E, and F show nominal 95% interval coverage. Teams that submitted national forecasts are presented in A. and D., state/territory/DC forecasts presented in B. and E., and teams that submitted large county level forecasts are presented in C. and F.

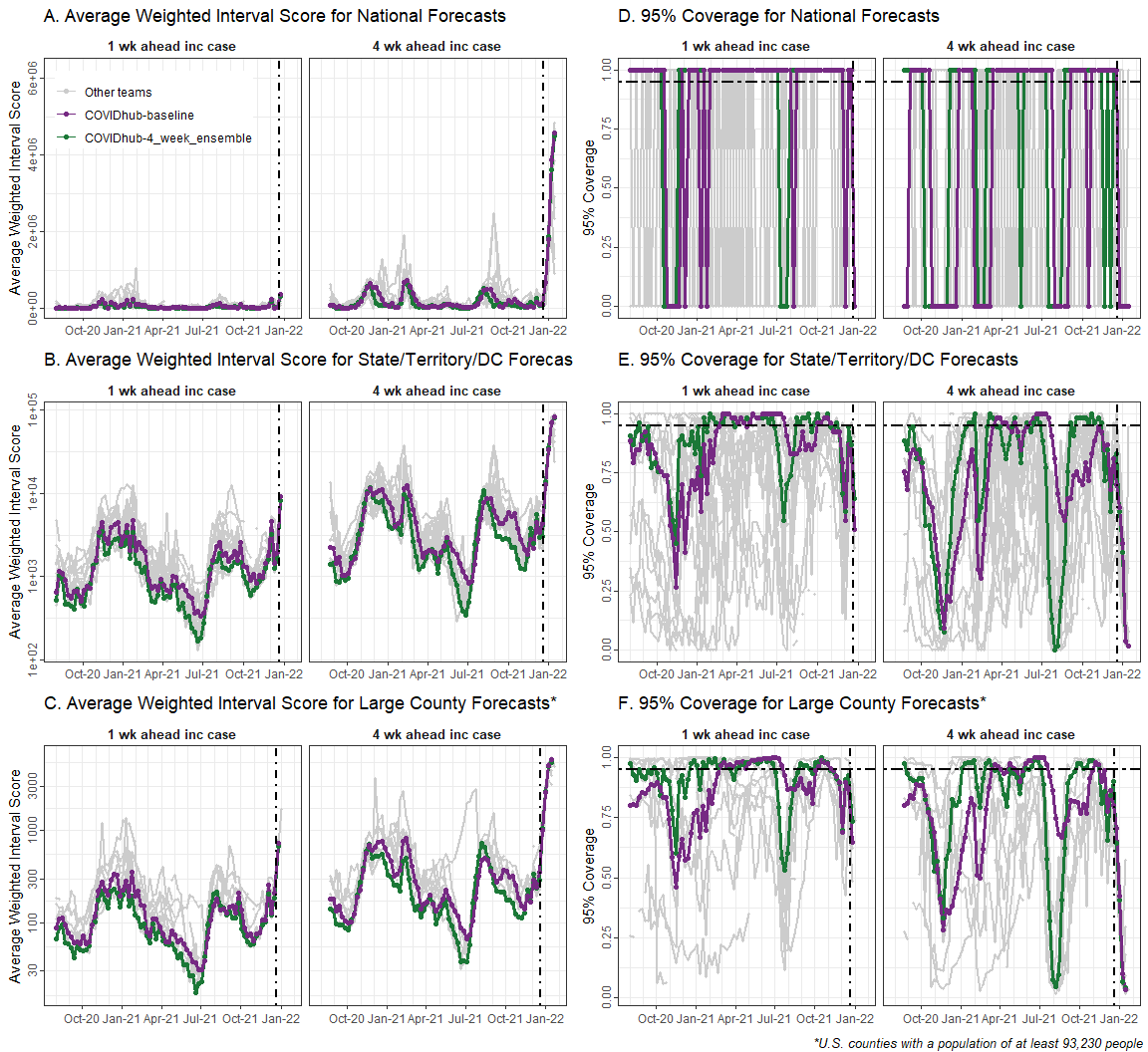

**S5 Figure 5.3.** Scaled, pairwise relative Weighted Interval Score (rWIS; see *Methods* for description) for all teams that submitted national and state/territory/DC forecasts by location for 1 and 4 week ahead horizon. National estimates are displayed first, followed by jurisdictions in alphabetical order. Teams are displayed by decreasing average rWIS across all forecast horizons and locations.

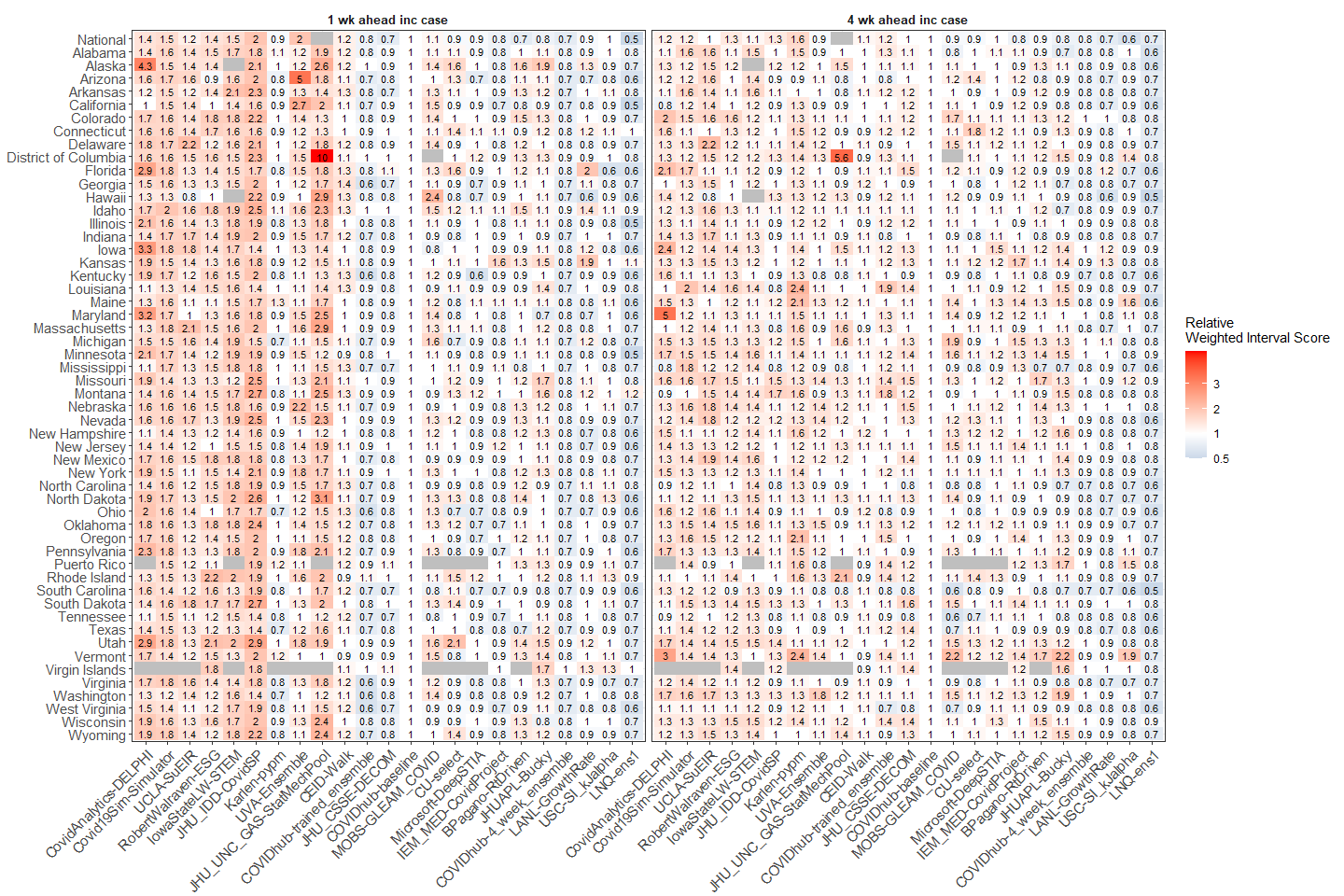
