## Supplement 6 for "Challenges of COVID-19 Case Forecasting in the US, 2020-2021"

**Supporting Information 6***:* Each team model’s estimated marginal mean Weighted Interval Score (WIS) over range of reported case counts per epidemic phase. Marginal mean WIS was estimated from GEE model results and reflect values across the 95% confidence interval of mean reported cases. Case counts differ per team model as each team forecasted a different set of locations over a different range of possible dates.

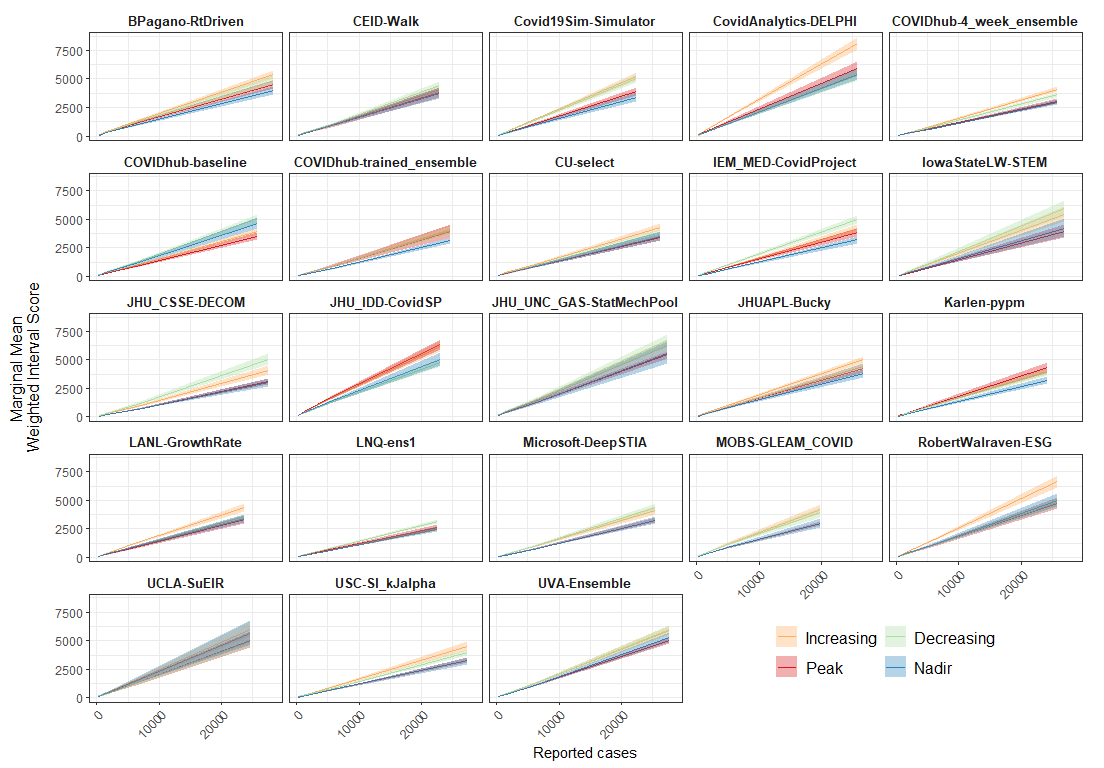
