## Supplement 7 for "Challenges of COVID-19 Case Forecasting in the US, 2020-2021"

**Supporting Information 7**: EPIFORGE 2020 guidelines outline 19 recommended reporting items for epidemic forecasting and prediction research (15). These items are included in the checklist below, which also include the page number where each item is described or presented within this evaluation.

| **Section of Manuscript** | **#** | **Checklist Item** | **Reported on Page** |
| --- | --- | --- | --- |
| Title/Abstract | 1 | Describe the study as forecast or prediction research in at least the title or abstract. | 1 |
| Introduction | 2 | Define the purpose of study and forecasting targets. | 1-2 |
| Methods | 3 | Fully document the methods. | 7-11 |
| Methods | 4 | Identify whether the forecast was performed prospectively, in real time, and/or retrospectively. | 8 |
| Methods | 5 | Explicitly describe the origin of input source data, with references. | 7-9 |
| Methods | 6 | Provide source data with publication, or document reasons as to why this was not possible. | 8-9 |
| Methods | 7 | Describe input data processing procedures in detail. | 7-9 |
| Methods | 8 | State and describe the model type, and document model assumptions, including references. | 27 |
| Methods | 9 | Make the model code available or document the reasons why this is not possible. | 11 |
| Methods | 10 | Describe the model validation and justify the approach. | 10 |
| Methods | 11 | Describe the forecast accuracy evaluation method used, with justification. | 10 |
| Methods | 12 | Where possible, compare results to a benchmark or other comparator model, with justification of comparator choice. | 10 |
| Methods | 13 | Describe the forecast horizon, with justification of its length. | 7-8 |
| Results | 14 | Present and explain uncertainty of forecasting results. | 3-4 |
| Results | 15 | Briefly summarize the results in nontechnical terms, including a nontechnical interpretation of forecast uncertainty. | 5 |
| Results | 16 | If results are published as a data object, encourage a time-stamped version number. | n/a |
| Discussion | 17 | Describe the weaknesses of the forecast, including weaknesses specific to data quality and methods. | 7 |
| Discussion | 18 | If the forecast research is applicable to a specific epidemic, comment on its potential implications and impact for public health action and decision-making. | 5-6 |
| Discussion | 19 | If the forecast research is applicable to a specific epidemic, comment on how generalizable it may be across populations. | 6 |
| n/a= Not Applicable | | | |
